# Genome-wide analyses identify 30 loci associated with obsessive-compulsive disorder

**DOI:** 10.1101/2024.03.13.24304161

**Authors:** Nora I. Strom, Zachary F. Gerring, Marco Galimberti, Dongmei Yu, Matthew W. Halvorsen, Abdel Abdellaoui, Cristina Rodriguez-Fontenla, Julia M. Sealock, Tim Bigdeli, Jonathan R. Coleman, Behrang Mahjani, Jackson G. Thorp, Katharina Bey, Christie L. Burton, Jurjen J. Luykx, Gwyneth Zai, Silvia Alemany, Christine Andre, Kathleen D. Askland, Julia Bäckmann, Nerisa Banaj, Cristina Barlassina, Judith Becker Nissen, O. Joseph Bienvenu, Donald Black, Michael H. Bloch, Sigrid Børte, Rosa Bosch, Michael Breen, Brian P. Brennan, Helena Brentani, Joseph D. Buxbaum, Jonas Bybjerg-Grauholm, Enda M. Byrne, Judit Cabana-Dominguez, Beatriz Camarena, Adrian Camarena, Carolina Cappi, Angel Carracedo, Miguel Casas, Maria Cristina Cavallini, Valentina Ciullo, Edwin H. Cook, Jesse Crosby, Bernadette A. Cullen, Elles J. De Schipper, Richard Delorme, Srdjan Djurovic, Jason A. Elias, Xavier Estivill, Martha J. Falkenstein, Bengt T. Fundin, Lauryn Garner, Christina Gironda, Fernando S. Goes, Marco A. Grados, Jakob Grove, Wei Guo, Jan Haavik, Kristen Hagen, Kelly Harrington, Alexandra Havdahl, Kira D. Höffler, Ana G. Hounie, Donald Hucks, Christina Hultman, Magdalena Janecka, Eric Jenike, Elinor K. Karlsson, Kara Kelley, Julia Klawohn, Janice E. Krasnow, Kristi Krebs, Christoph Lange, Nuria Lanzagorta, Daniel Levey, Kerstin Lindblad-Toh, Fabio Macciardi, Brion Maher, Brittany Mathes, Evonne McArthur, Nathaniel McGregor, Nicole C. McLaughlin, Sandra Meier, Euripedes C. Miguel, Maureen Mulhern, Paul S. Nestadt, Erika L. Nurmi, Kevin S. O’Connell, Lisa Osiecki, Olga Therese Ousdal, Teemu Palviainen, Nancy L. Pedersen, Fabrizio Piras, Federica Piras, Sriramya Potluri, Raquel Rabionet, Alfredo Ramirez, Scott Rauch, Abraham Reichenberg, Mark A. Riddle, Stephan Ripke, Maria C. Rosário, Aline S. Sampaio, Miriam A. Schiele, Anne Heidi Skogholt, Laura G. Sloofman, Jan Smit, María Soler Artigas, Laurent F. Thomas, Eric Tifft, Homero Vallada, Nathanial van Kirk, Jeremy Veenstra-VanderWeele, Nienke N. Vulink, Christopher P. Walker, Ying Wang, Jens R. Wendland, Bendik S. Winsvold, Yin Yao, Hang Zhou, Estonian Biobank, 23andMe Inc., Arpana Agrawal, Pino Alonso, Götz Berberich, Kathleen K. Bucholz, Cynthia M. Bulik, Danielle Cath, Damiaan Denys, Valsamma Eapen, Howard Edenberg, Peter Falkai, Thomas V. Fernandez, Abby J. Fyer, J. M. Gaziano, Dan A. Geller, Hans J. Grabe, Benjamin D. Greenberg, Gregory L. Hanna, Ian B. Hickie, David M. Hougaard, Norbert Kathmann, James Kennedy, Dongbing Lai, Mikael Landén, Stéphanie Le Hellard, Marion Leboyer, Christine Lochner, James T. McCracken, Sarah E. Medland, Preben B. Mortensen, Benjamin M. Neale, Humberto Nicolini, Merete Nordentoft, Michele Pato, Carlos Pato, David L. Pauls, John Piacentini, Christopher Pittenger, Danielle Posthuma, Josep Antoni Ramos-Quiroga, Steven A. Rasmussen, Margaret A. Richter, David R. Rosenberg, Stephan Ruhrmann, Jack F. Samuels, Sven Sandin, Paul Sandor, Gianfranco Spalletta, Dan J. Stein, S. Evelyn Stewart, Eric A. Storch, Barbara E. Stranger, Maurizio Turiel, Thomas Werge, Ole A. Andreassen, Anders D. Børglum, Susanne Walitza, Kristian Hveem, Bjarne K. Hansen, Christian Rück, Nicholas G. Martin, Lili Milani, Ole Mors, Ted Reichborn-Kjennerud, Marta Ribasés, Gerd Kvale, David Mataix-Cols, Katharina Domschke, Edna Grünblatt, Michael Wagner, John-Anker Zwart, Gerome Breen, Gerald Nestadt, Jaakko Kaprio, Paul D. Arnold, Dorothy E. Grice, James A. Knowles, Helga Ask, Karin J. Verweij, Lea K. Davis, Dirk J. Smit, James J. Crowley, Jeremiah M. Scharf, Murray B. Stein, Joel Gelernter, Carol A. Mathews, Eske M. Derks, Manuel Mattheisen

## Abstract

Obsessive-compulsive disorder (OCD) affects ∼1% of children and adults and is partly caused by genetic factors. We conducted a genome-wide association study (GWAS) meta-analysis combining 53,660 OCD cases and 2,044,417 controls and identified 30 independent genome-wide significant loci. Gene-based approaches identified 249 potential effector genes for OCD, with 25 of these classified as the most likely causal candidates, including *WDR6, DALRD3, CTNND1* and multiple genes in the MHC region. We estimated that ∼11,500 genetic variants explained 90% of OCD genetic heritability. OCD genetic risk was associated with excitatory neurons in the hippocampus and cortex, along with D1- and D2-type dopamine receptor-containing medium spiny neurons. OCD genetic risk was shared with 65 of 112 additional phenotypes, including all of the psychiatric disorders we examined. In particular, OCD shared genetic risk with anxiety, depression, anorexia nervosa, and Tourette syndrome, and was negatively associated with inflammatory bowel diseases, educational attainment, and body mass index.

## Introduction

Obsessive-compulsive disorder (OCD) is a chronic psychiatric illness that affects 1-3% of the population^1^ and is characterized by obsessions and compulsions that vary in type and severity, and over time. OCD is responsible for profound personal and societal costs^2^, including increased risk of suicide^3^ and overall mortality^4^. OCD is moderately heritable; twin-based heritability estimates range between 27-47% in adults and between 45-65% in children^5–8^, with SNP-based heritability estimates between 28-37%^9–11^.

Two earlier OCD GWAS meta-analyses, both containing a subset of the data included in this analysis^12,13^, showed SNP-based heritabilities of 8.5% (assuming a 3% population prevalence) and 16% (assuming a 2% population prevalence). The first GWAS (*n*_cases_ = 14,140, *n*_controls_ = 562,117)^12^ found one genome-wide significant locus associated with OCD, while the second (*n*_cases_ = 37,015, *n*_controls_ = 948,616)^13^ identified 15 independent genome-wide significant loci. As with other complex traits, increased sample sizes are needed for a more comprehensive understanding of the underlying genetic etiology of OCD and its genetic relationships with related disorders.

The current study combines data from the two unpublished OCD GWASs described above and includes additional cohorts (∼9,000 cases). This results in the largest and most well-powered GWAS of OCD to date, with a ∼20-fold increase of OCD cases compared to the previously published OCD GWASs^10^. Based on the results from the meta-analysis, we conducted secondary analyses, including positional and functional fine-mapping of SNPs and genes, structural equation modeling to examine possible genetic differences in sample ascertainment across cohorts, protein and transcriptome-wide association analyses, single-cell enrichment, and genetic correlations with other traits (**Supplementary Fig. 1**). Our results provide more detailed insight into the genetic underpinnings and biology of OCD.

## Results

### GWAS meta-analysis identifies 30 genome-wide significant loci

We conducted a GWAS meta-analysis of 28 European-ancestry OCD case-control cohorts, comprising 53,660 cases and 2,044,417 controls (effective sample size ∼210,000 individuals). Ascertainment of cases varied across cohorts: OCD diagnosis was determined (i) by a healthcare professional in a clinical setting (18 cohorts, *n* = 9,089 cases), (ii) from health records or biobanks (7 cohorts, *n* = 9,138 cases), (iii) in a clinical setting or from health records with the additional characteristic that all OCD cases were primarily collected for another psychiatric disorder (3 cohorts, *n* = 5,266 cases), or (iv) by self-reported diagnosis in a consumer-based setting (23andMe, Inc., *n* = 30,167 cases). Cohort details, including phenotypic assessment, quality control, and individual cohort GWAS analyses are described in **Supplementary Note 2** and **Supplementary Table 1**. We identified 30 independent (defined in **Supplementary Note 3**) loci among the 1,672 SNPs that exceeded the genome-wide threshold for significance (*P* < 5 × 10^−8^; Manhattan plot in **Fig. 1**, regional association plots and forest plots in **Supplementary Figs. 2-31**, and a list of all independent genome-wide significant SNPs in **Table 1** with additional details in **Supplementary Tables 2** and **3**). The independence of the 30 lead SNPs was subsequently validated using conditional and joint analysis (GCTA-COJO)^14^ (**Supplementary Table 4**). An analysis of the X-chromosome, conducted in a subset of the data for which this information was available (23andMe), yielded no significant associations (**Supplementary Note 4** and **Supplementary Figure 37e**). Of the 15 genome-wide significant loci previously reported in preprints^12,13^, 13 were genome-wide significant in the current GWAS, with the remaining two showing suggestive significance (*P* = 5.23 × 10^−8^ and *P* = 2.2 × 10^−7^; **Supplementary Table 5**). Using MiXeR^15^, we estimated that approximately 11,500 (s.e. = 607) causal variants account for 90% of the OCD SNP-based heritability.

**Figure 1.**
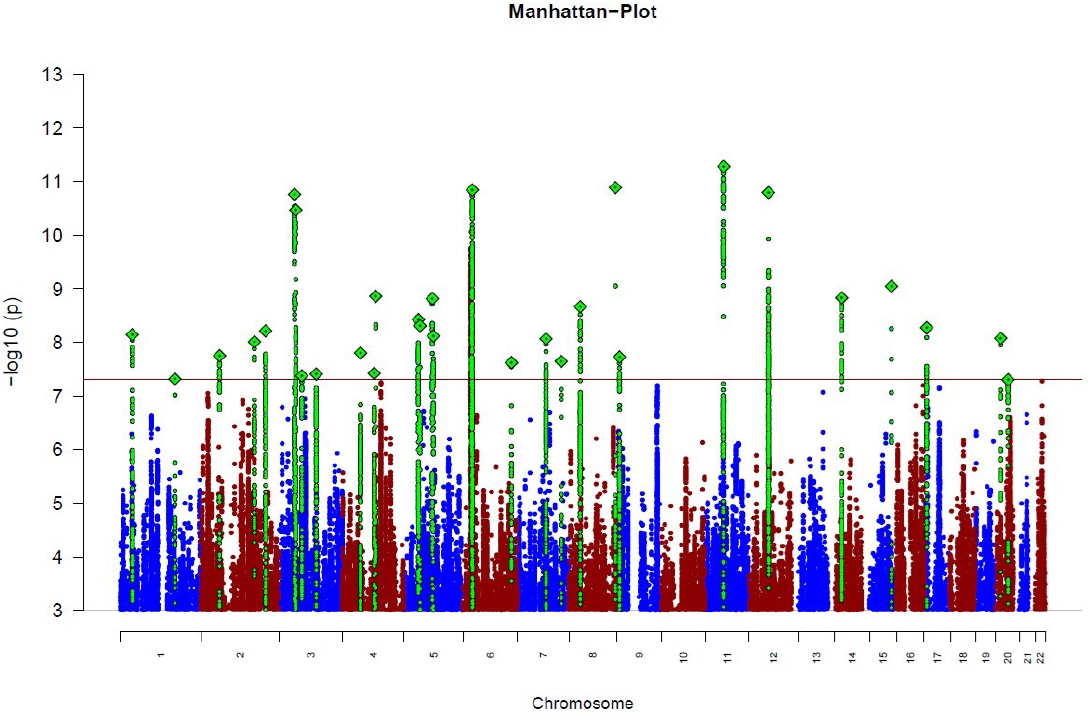
Manhattan plot of OCD GWAS meta-analysis. The *y*-axis represents –log_10_ *P*-values (two-sided, not adjusted for multiple testing) for the association of variants with OCD using an inverse-variance weighted fixed effects model (*n*_cases_ = 53,660 and *n*_controls_ = 2,044,417). The *x*-axis shows chromosomes 1 to 22. The horizontal red line represents the threshold for genome-wide significance (*P* = 5 × 10^−8^). Index variants of genome-wide significant loci are highlighted as a green diamond.

**Table 1.**
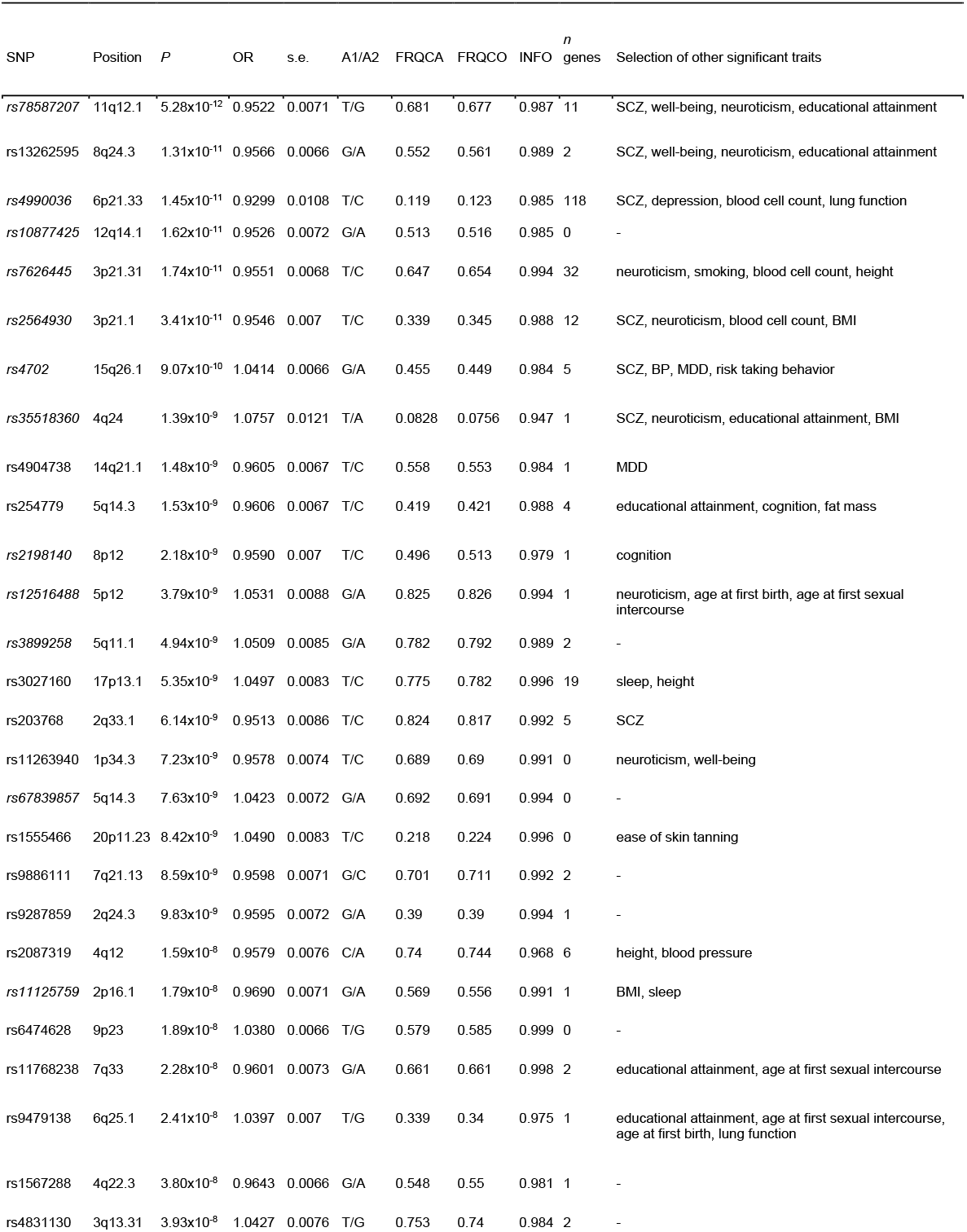
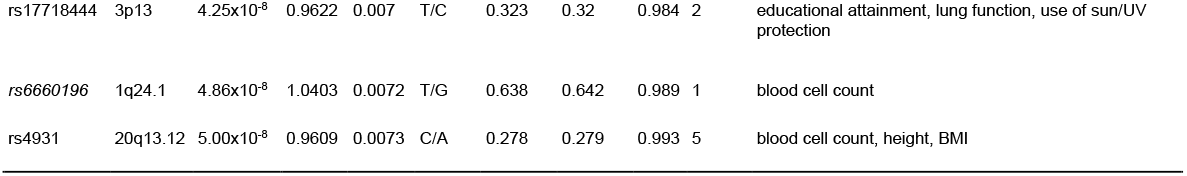
Genome-wide-significant loci associated with OCD. Shown are the lead SNP, the chromosome (CHR), base pair position on the genome (BP), *P*-value (*P*), effect estimate as an odds ratio (OR), standard error of the effect estimate (s.e.), effect allele and non-effect allele (A1/A2), frequency of A1 in cases (FRQCA) and in controls (FRQCO), imputation quality score (INFO), number of genes in a region of 6.5 kb around the SNP (*n* genes), and a curated list of phenotypes that also showed a genome-wide significant association with this SNP (in one or more of the following four databases: causalDB^90^, GenomeAtlas^52^, NHGRI-EBI GWAS catalog^91^, IEU Open GWAS project^92^. If fewer than four traits are significant across all four databases, all four traits are shown. If more than five traits are significant across the databases, neuropsychiatric traits are prioritized (closely related traits are summarized into one trait category). Previously identified GWAS hits for OCD (or SNPs in high LD with a previously identified SNP) are in italics. For a full list of associations in the four databases, see **Supplementary Table 18a-**d. A more detailed list of the significant loci can be found in **Supplementary Table 2**. Abbreviations in the last column are: schizophrenia (SCZ), bipolar disorder (BP), major depressive disorder (MDD), and body-mass index (BMI).

| SNP | Position | P | OR | s.e. | A1/A2 | FRQCA | FRQCO | INFO | <sup>n</sup><br>genes | Selection of other significant traits |
| --- | --- | --- | --- | --- | --- | --- | --- | --- | --- | --- |
| rs78587207 | 11q12.1 | 5.28x10<sup>-12</sup> | 0.9522 | 0.0071 | T/G | 0.681 | 0.677 | 0.987 | 11 | SCZ, well-being, neuroticism, educational attainment |
| rs13262595 | 8q24.3 | 1.31x10<sup>-11</sup> | 0.9566 | 0.0066 | G/A | 0.552 | 0.561 | 0.989 | 2 | SCZ, well-being, neuroticism, educational attainment |
| rs4990036 | 6p21.33 | 1.45x10<sup>-11</sup> | 0.9299 | 0.0108 | T/C | 0.119 | 0.123 | 0.985 | 118 | SCZ, depression, blood cell count, lung function |
| rs10877425 | 12q14.1 | 1.62x10<sup>-11</sup> | 0.9526 | 0.0072 | G/A | 0.513 | 0.516 | 0.985 | 0 | - |
| rs7626445 | 3p21.31 | 1.74x10<sup>-11</sup> | 0.9551 | 0.0068 | T/C | 0.647 | 0.654 | 0.994 | 32 | neuroticism, smoking, blood cell count, height |
| rs2564930 | 3p21.1 | 3.41x10<sup>-11</sup> | 0.9546 | 0.007 | T/C | 0.339 | 0.345 | 0.988 | 12 | SCZ, neuroticism, blood cell count, BMI |
| rs4702 | 15q26.1 | 9.07x10<sup>-10</sup> | 1.0414 | 0.0066 | G/A | 0.455 | 0.449 | 0.984 | 5 | SCZ, BP, MDD, risk taking behavior |
| rs35518360 | 4q24 | 1.39x10<sup>-9</sup> | 1.0757 | 0.0121 | T/A | 0.0828 | 0.0756 | 0.947 | 1 | SCZ, neuroticism, educational attainment, BMI |
| rs4904738 | 14q21.1 | 1.48x10<sup>-9</sup> | 0.9605 | 0.0067 | T/C | 0.558 | 0.553 | 0.984 | 1 | MDD |
| rs254779 | 5q14.3 | 1.53x10<sup>-9</sup> | 0.9606 | 0.0067 | T/C | 0.419 | 0.421 | 0.988 | 4 | educational attainment, cognition, fat mass |
| rs2198140 | 8p12 | 2.18x10<sup>-9</sup> | 0.9590 | 0.007 | T/C | 0.496 | 0.513 | 0.979 | 1 | cognition |
| rs12516488 | 5p12 | 3.79x10<sup>-9</sup> | 1.0531 | 0.0088 | G/A | 0.825 | 0.826 | 0.994 | 1 | neuroticism, age at first birth, age at first sexual intercourse |
| rs3899258 | 5q11.1 | 4.94x10<sup>-9</sup> | 1.0509 | 0.0085 | G/A | 0.782 | 0.792 | 0.989 | 2 | - |
| rs3027160 | 17p13.1 | 5.35x10<sup>-9</sup> | 1.0497 | 0.0083 | T/C | 0.775 | 0.782 | 0.996 | 19 | sleep, height |
| rs203768 | 2q33.1 | 6.14x10<sup>-9</sup> | 0.9513 | 0.0086 | T/C | 0.824 | 0.817 | 0.992 | 5 | SCZ |
| rs11263940 | 1p34.3 | 7.23x10<sup>-9</sup> | 0.9578 | 0.0074 | T/C | 0.689 | 0.69 | 0.991 | 0 | neuroticism, well-being |
| rs67839857 | 5q14.3 | 7.63x10<sup>-9</sup> | 1.0423 | 0.0072 | G/A | 0.692 | 0.691 | 0.994 | 0 | - |
| rs1555466 | 20p11.23 | 8.42x10<sup>-9</sup> | 1.0490 | 0.0083 | T/C | 0.218 | 0.224 | 0.996 | 0 | ease of skin tanning |
| rs9886111 | 7q21.13 | 8.59x10<sup>-9</sup> | 0.9598 | 0.0071 | G/C | 0.701 | 0.711 | 0.992 | 2 | - |
| rs9287859 | 2q24.3 | 9.83x10<sup>-9</sup> | 0.9595 | 0.0072 | G/A | 0.39 | 0.39 | 0.994 | 1 | - |
| rs2087319 | 4q12 | 1.59x10<sup>-8</sup> | 0.9579 | 0.0076 | C/A | 0.74 | 0.744 | 0.968 | 6 | height, blood pressure |
| rs11125759 | 2p16.1 | 1.79x10<sup>-8</sup> | 0.9690 | 0.0071 | G/A | 0.569 | 0.556 | 0.991 | 1 | BMI, sleep |
| rs6474628 | 9p23 | 1.89x10<sup>-8</sup> | 1.0380 | 0.0066 | T/G | 0.579 | 0.585 | 0.999 | 0 | - |
| rs11768238 | 7q33 | 2.28x10<sup>-8</sup> | 0.9601 | 0.0073 | G/A | 0.661 | 0.661 | 0.998 | 2 | educational attainment, age at first sexual intercourse |
| rs9479138 | 6q25.1 | 2.41x10<sup>-8</sup> | 1.0397 | 0.007 | T/G | 0.339 | 0.34 | 0.975 | 1 | educational attainment, age at first sexual intercourse, age at first birth, lung function |
| rs1567288 | 4q22.3 | 3.80x10<sup>-8</sup> | 0.9643 | 0.0066 | G/A | 0.548 | 0.55 | 0.981 | 1 | - |
| rs4831130 | 3q13.31 | 3.93x10<sup>-8</sup> | 1.0427 | 0.0076 | T/G | 0.753 | 0.74 | 0.984 | 2 | - |
| rs17718444 | 3p13 | 4.25x10<sup>-8</sup> | 0.9622 | 0.007 | T/C | 0.323 | 0.32 | 0.984 | 2 | educational attainment, lung function, use of sun/UV protection |
| rs6660196 | 1q24.1 | 4.86x10<sup>-8</sup> | 1.0403 | 0.0072 | T/G | 0.638 | 0.642 | 0.989 | 1 | blood cell count |
| rs4931 | 20q13.12 | 5.00x10<sup>-8</sup> | 0.9609 | 0.0073 | C/A | 0.278 | 0.279 | 0.993 | 5 | blood cell count, height, BMI |

No statistically significant heterogeneity was observed across individual cohorts for the 30 genome-wide significant loci, as assessed with Cochran’s Q-test (**Supplementary Fig. 32**), the I^2^ statistic, and GenomicSEM’s *Q*_*SNP*_-statistic^16^ (**Supplementary Table 2**). Genome-wide analyses of samples grouped by clinical, comorbid, biobank, and 23andMe (**Supplementary Table 3** and **Supplementary Figs. 33-37**) showed evidence that sample ascertainment impacted results at a genome-wide scale, although not beyond what is observed with closely related psychiatric disorders^17,18^. We observed moderate to high genetic correlations across the subgroups (between 0.63, s.e. = 0.11 for biobanks and comorbid, and 0.92, s.e. = 0.07 for 23andMe and comorbid; **Supplementary Table 7**), and a satisfactory fit for a one-factor GenomicSEM model (**Supplementary Table 8** and **Supplementary Fig. 39**). A common-factor GWAS based on the one-factor GenomicSEM model resulted in 20 significant loci, all of which were also significant in the primary GWAS (**Supplementary Table 8** and **Supplementary Fig. 40**; analysis details in **Supplementary Note 5**). SNP-heritability (assuming a 1% population prevalence) was 6.7% (s.e. = 0.3%), with slightly higher estimates for the clinical 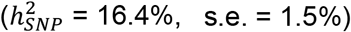 and comorbid 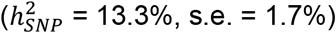 subgroups (**Supplementary Table 1**).

### Gene-based findings

We prioritized putative risk genes for OCD using six positional and functional QTL gene-based mapping approaches. Positional mapping was performed with mBAT-combo^19^. Functional eQTL mapping was performed with transcriptome-wide association study (TWAS)^20^ using PsychENCODE gene expression weights^21^, and summary-based Mendelian randomization (SMR)^22^ using whole blood eQTLGen^23^ and MetaBrain^24^ datasets. Functional protein QTL (pQTL) mapping was done using a protein-wide association study (PWAS) of human brain protein expression panels^25^. Finally, we used PsyOPS^26^, which combines positional mapping with biological annotations, to further prioritize risk genes within genome-wide significant loci. We identified 207 significant genes (Bonferroni correction, *P* < 2.67 × 10^−6^) with mBAT-combo, and 24 genes using TWAS (*P* < 4.76 × 10^−6^), 14 of which were conditionally independent. The SMR-eQTLGen analysis identified 39 significant risk genes (*P* < 4.28 × 10^−6^), and the SMR-MetaBrain analysis identified 14 risk genes (*P* < 9.23 × 10^−6^). The PWAS identified 3 significant genes (*P* < 3.39 × 10^−5^), while PsyOPS prioritized 29 genes. In total, 251 genes were significantly associated with OCD through at least one gene-based approach, and 48 were implicated by at least two methods (details in **Online Methods, Supplementary Note 7**, and **Supplementary Tables 9-14**).

From the 48 genes implicated by at least two approaches, we prioritized likely causal genes for OCD using colocalization (TWAS-COLOC)^27,28^ and SMR-heterogeneity in dependent instruments (SMR-HEIDI)^22^ tests. Colocalization was used to identify significant TWAS associations where the underlying GWAS and eQTL summary statistics are likely to share a single causal variant. Similarly, HEIDI was used to select SMR associations where the same causal variant affects gene expression and trait variation. 25 of the 48 genes implicated by at least two gene-based tests were also significant in either the TWAS-COLOC or SMR-HEIDI tests, implying causality (**Fig. 2a**). Only two of these 25 genes were prioritized by both TWAS-COLOC and SMR-HEIDI, WD repeat domain 6 (*WDR6*) and DALR Anticodon Binding Domain Containing 3 (*DALRD3*). Another gene of interest, Catenin Delta 1 (*CTNND1*), was implicated by three of our five approaches (mBAT-combo, TWAS, PWAS) and showed evidence for colocalization. Only three genes were implicated in the PWAS; of these, *CTNND1* was the only gene also implicated in the TWAS. In the PWAS, downregulation of *CTNND1* protein expression in human dorsolateral prefrontal cortex was significantly associated with OCD risk (*Z* = −4.49, *P* = 7.11 × 10^−6^; **Supplementary Table 13**), consistent with the downregulation of *CTNND1* gene expression in prefrontal cortex seen in the TWAS (*Z* = −6.86, *P* = 6.90 × 10^−12^; **Supplementary Table 10**). For a discussion of the overlap between the gene-findings with rare-coding variants in OCD, see **Supplementary Table 6** and **Supplementary Note 7**.

**Figure 2.**
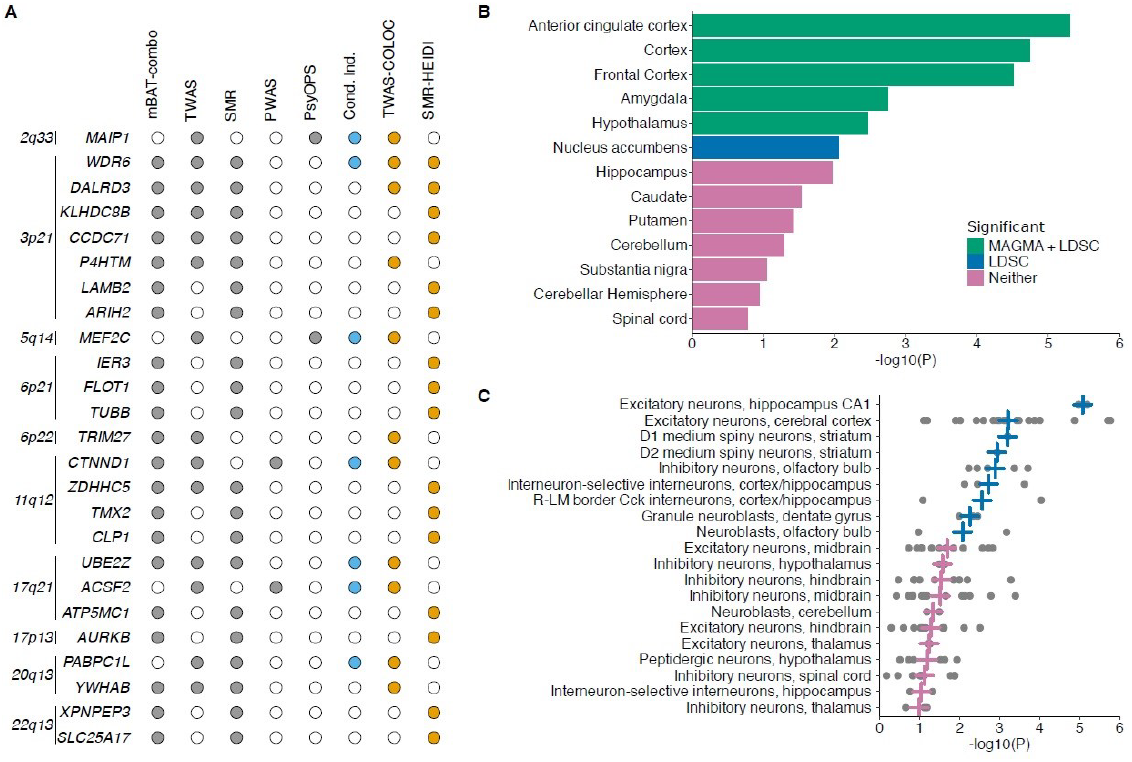
Gene-based, tissue, and cell type enrichment analyses. **a**, List of 25 genes that were implicated in at least two of the five different gene-based tests (significance indicated by grey dots) and passed the TWAS colocalization and/or SMR-HEIDI filters (significance indicated by orange dots). Conditionally independent genes within each locus are indicated by blue dots. **b**, Enrichment of OCD GWAS signal in human brain-related tissues from GTEx (v8). No significant enrichment was observed in the peripheral tissues (not included in the figure). The horizontal bar size represents the significance of the enrichment measured using the MAGMA gene set enrichment test or partitioned LDSC. **c**, Top 20 groups of brain cell types (*n* = 35 total tested) enriched with OCD GWAS signal using MAGMA. Dots represent –log_10_ *P*-values from MAGMA gene set enrichment tests of individual neuronal cell types from Zeisel et al.^30^. Vertical crosses represent the mean −log_10_ *P*-value observed for each brain cell type group. Blue crosses represent a significant enrichment of OCD GWAS signals (false discovery rate across 35 groups, FDR < 0.05), while pink crosses indicate non-significant enrichment. Grey points represent the association (−log_10_ *P*-value) for each single cell cluster (“level 5” analysis defined by Zeisel et al.^30^) in a given cell type (e.g., excitatory neurons, cerebral cortex).

### Tissue and cell type enrichment analysis

After mapping significantly associated SNPs from the GWAS meta-analysis to likely causal genes, we explored which tissues or cell types showed enriched gene expression of OCD associated genetic signals, using a previously described approach^29^ on published human gene expression datasets from bulk tissue RNA-seq data from GTEx and single-cell RNA-sequencing data from the adult mouse central and peripheral nervous systems (CNS and PNS)^30^. We found enrichment of OCD GWAS signals in 6 of 13 human brain tissue types in GTEx, but no enrichment in human peripheral tissues (**Fig. 2b** and **Supplementary Table 15**). In the adult mouse CNS and PNS, we found enrichment of OCD GWAS signals in 41 of 166 tested specific single cell types using the MAGMA gene-set enrichment test (**Supplementary Table 16**). When summarizing results of individual single cell types into groups of cell types defined by the same region/tissue and cell type, 9 of 35 are enriched for OCD GWAS signals (top 20 shown in **Fig. 2c**). Strong enrichment of OCD GWAS signal was especially observed in excitatory neurons of hippocampus and cerebral cortex, as well as D1 and D2 medium spiny neurons.

### Genetic relationship of OCD with other phenotypes

Using phenome-wide association analysis, we examined whether the 30 independent OCD-associated loci identified by our GWAS meta-analysis have previously been associated with other phenotypes (see **Supplementary Tables 17a-d** for look-ups in four, partially overlapping GWAS-databanks and **Table 1** for highlighted associations). We found that 22 of the 30 loci were associated with other phenotypes, including schizophrenia (seven loci), depression/major depressive disorder (two loci), bipolar disorder (one locus), neuroticism (seven loci), educational attainment (seven loci), and body fat mass or body mass index (eight loci).

We further used bivariate linkage-disequilibrium score regression (LDSC)^31^ to investigate the extent of genetic correlations between OCD and 112 previously published GWASs encompassing psychiatric, substance use, and neurological phenotypes, among others (**Fig. 3**). We found that 65 phenotypes were significantly correlated with OCD after correcting for multiple testing using the Benjamini-Hochberg^32^ procedure to control the false-discovery rate (FDR) at a threshold of 0.05. OCD was significantly positively correlated with all tested psychiatric phenotypes, the highest correlations with anxiety (ANX, *r*_*G*_ = 0.70), depression (DEP, *r*_*G*_ = 0.60), anorexia nervosa (AN, *r*_*G*_ = 0.52), Tourette syndrome (TS, *r*_*G*_ = 0.47), and post-traumatic stress disorder (PTSD, *r*_*G*_ = 0.48). Significant positive genetic correlations were also obtained for neuroticism (*r*_*G*_ = 0.53), in particular for the worry subcluster (*r*_*G*_ = 0.64), and all individual items in the worry subcluster, with slightly lower estimates for the depressive sub-cluster (*r*_*G*_ = 0.35). Suicide attempt (*r*_*G*_ = 0.40), history of childhood maltreatment (*r*_*G*_ = 0.37), and tiredness (*r*_*G*_ = 0.36) were also notable for strong positive associations with OCD. Of the assessed neurological disorders, OCD was only significantly correlated with migraine (*r*_*G*_ = 0.15). Some autoimmune disorders, such as Crohn’s disease (*r*_*G*_ = −0.13), ulcerative colitis (*r*_*G*_ = −0.14), and inflammatory bowel disease (*r*_*G*_ = −0.14), showed negative correlations with OCD (see **Fig. 3, Supplementary Table 18** for all genetic correlation estimates, 95% CI and *P*-values, **Supplementary Note 6** for a more in depth discussion of all significant genetic correlations, and **Supplementary Table 19** and **Supplementary Figs. 41** and **42** for sub-group specific genetic-correlation estimates).

**Figure 3.**
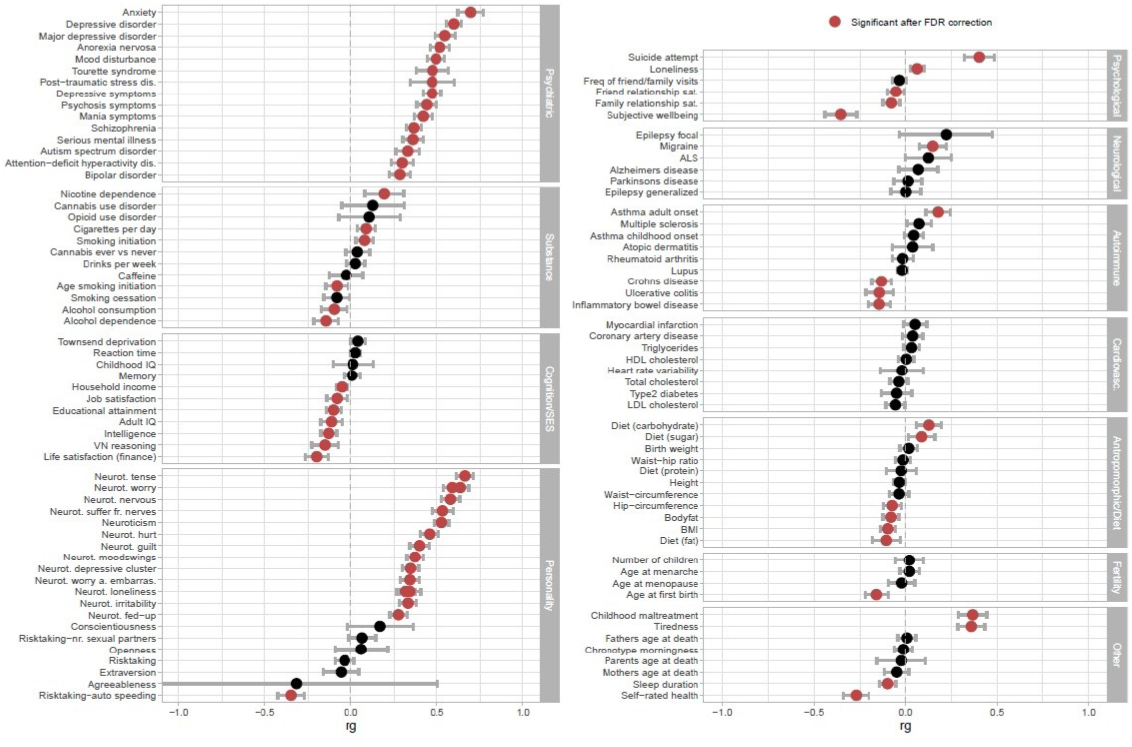
Genetic correlations (*r*_G_) between OCD and 112 phenotypes. This includes psychiatric, substance use, cognition/socioeconomic status (SES), personality, psychological, neurological, autoimmune, cardiovascular, anthropomorphic/diet, fertility, and other phenotypes. References and sample sizes of the corresponding summary statistics of the GWAS studies can be found in **Supplementary Table 18**. The OCD summary statistics are of the main meta-analysis (*n*_cases_ = 53,660 and *n*_controls_ = 2,044,417). Error bars represent the 95% confidence intervals for the genetic correlation estimates (*r*_G_). Red circles indicate significant associations with a *P*-value adjusted for multiple testing with the Benjamini-Hochberg procedure to control the FDR (< 0.05). Black circles indicate associations that are not significant.

## Discussion

The OCD GWAS reported here, comprising over 53,000 cases, identified 30 independent genome-wide significant loci. Common SNPs explained 6.7% of the variation in OCD risk in our meta-analysis (LDSC with an assumed population prevalence of 1%), a significant reduction from the 28% reported previously^10^. However, differences in the assumed population prevalence (where a lower assumed prevalence for LDSC heritability calculation results in a lower heritability estimate) and an increase in sample heterogeneity likely contributed to this discrepancy. The reduction in SNP-heritability is in line with previous observations for closely related psychiatric disorders such as ADHD^33,34^ or depression^17,35–37^, where expanding the phenotype definition increased genetic heterogeneity, potentially accounting for the observed decrease in SNP heritability. This aligns with the fact that heritability estimates for more homogeneous OCD subgroups were higher: 16.4% for the clinically-ascertained subgroup and 13.3% for the comorbid subgroup (**Supplementary Note 10**). The current estimates are comparable to those of other psychiatric and substance use disorders, with SNP-heritability estimates ranging between 9% and 28%^38^.

The most significant SNP rs78587207 (*P* = 5.28 × 10^−12^) identified in the GWAS is located on chr11q12.1 and has been previously associated with several traits, including neuropsychiatric phenotypes^39^ such as depressive symptoms^40^ and neuroticism^40^. Gene-based analyses identified four putative causal genes within this locus. The closest gene to rs78587207 is catenin delta 1 (*CTNND1*), which encodes the cell adhesion molecule p120-catenin. This gene was associated with OCD using three gene-based tests (mBAT-combo, TWAS, and PWAS) and we found strong evidence for colocalization of the TWAS signal for *CTNND1* in dorso-lateral prefrontal cortex (dlPFC). dlPFC has been consistently implicated in the neural circuitry of OCD as well as in compulsivity more broadly as part of the cortico-striatal-thalamo-cortical circuitry^41,42^. The protein product of *CTNND1* is a regulator of cell-cell adhesion^43^ and plays a crucial role in gene transcription, Rho GTPase activity, and cytoskeletal organization^44–46^. Other credible causal genes in the locus include cleavage factor polyribonucleotide kinase subunit 1 (*CLP1*), thioredoxin related transmembrane protein 2 (*TMX2*), and zinc finger DHHC-type palmitoyltransferase 5 (*ZDHHC5*). Rare genetic mutations in *CLP1* are associated with pontocerebellar hypoplasia type 10, a very rare autosomal recessive neurodegenerative disease characterized by brain atrophy and delayed myelination resulting in intellectual disability^47^. *TMX2* is associated with increased risk of neurodevelopmental disorders with microcephaly, cortical malformations, spasticity, and congenital nervous system abnormalities^48^. *ZDHHC5* is broadly expressed in the brain, including the frontal cortex. *ZDHHC5* has not been implicated in brain development but has been linked to lung acinar adenocarcinoma and lung papillary adenocarcinoma in prior studies^49^.

Our finding that approximately 11,500 (s.e. = 607) causal variants account for 90% of the SNP-based heritability of OCD suggests that OCD is more polygenic than other complex traits such as height (*n*_causal_ = 4,000), schizophrenia (*n*_causal_ = 9,600) and ADHD (*n*_causal_ = 5,600), but less polygenic than major depression (*n*_causal_ = 14,500) and educational attainment (*n*_causal_ = 13,200)^50^.

We identified a total of 25 credible causal genes based on robust evidence using multiple positional and functionally-informed gene-based approaches. Notably, *DLGAP1*, which has been previously implicated in OCD pathogenesis^10,51^, was not identified in either the GWAS or in the gene-based analyses. Of the 25 genes that were implicated, 15 were within 6.5 kb of a SNP that surpassed genome-wide significance in the meta-analysis. In addition to the four genes discussed above, several others are of particular interest, including *WDR6* and *DALRD3*, which had the strongest evidence from the gene-based analyses. These genes lie in a gene-rich region on chr3p21.31, which in addition to harboring multiple genome-wide significant SNPs, has been previously associated with a broad range of psychiatric disorders and related traits, including schizophrenia^39^, well-being^52^, and the worry-subcluster of neuroticism^53^.

*WDR6* (WD repeat domain 6) is broadly expressed in the brain, particularly the hypothalamus. Its protein product is involved in cell growth arrest^54^, and recent studies have implicated it in anorexia nervosa^55^ and Parkinson’s disease^56^. *DALRD3* (DALR anticodon binding domain containing 3) is located on chromosome 3 in the same region as *WDR6. DALRD3*, when fully disrupted, is implicated in a form of epileptic encephalopathy with associated developmental delay^57^. Finally, a third gene in the 3p21 locus, *CELSR3* (cadherin EGF LAG seven-pass G-type receptor 3), encodes a protocadherin that is highly expressed in the developing basal ganglia^58^. Multiple loss-of-function mutations in *CELSR3* have been associated with Tourette syndrome^59,60^, which co-occurs with OCD in 10-20% of patients.

Four other genes identified through these analyses are located in the MHC locus, a region on chromosome 6 that plays a major role in the adaptive immune system and has been repeatedly linked to major psychiatric disorders^61^. The newly identified MHC association for OCD is noteworthy given evidence linking OCD with autoimmune disorders^62–64^. Genetic pleiotropy may underlie this connection, with variants predisposing individuals to both autoimmune conditions and OCD^65^. Further, some OCD subtypes, such as pediatric acute-onset neuropsychiatric disorders associated with streptococcus (PANDAS) and pediatric acute-onset neuropsychiatric syndrome (PANS) may have autoimmune origins^66,67^. Nevertheless, we were surprised to discover several negative genetic correlations between OCD and autoimmune disorders such as Crohn’s disease, ulcerative colitis, and inflammatory bowel disease in our analyses, suggesting that there is heterogeneity (and perhaps pleiotropy) in the genetic relationships between autoimmune disorders and OCD.

Tissue and cell type enrichment analysis revealed significant enrichment of OCD SNP-heritability in several tissues and cell types, with the strongest enrichment in excitatory neurons of hippocampus and cerebral cortex, and in dopamine D1 receptor (D1R)-positive and dopamine D2 receptor (D2R)-positive medium spiny neurons (MSNs) in the striatum. These findings are in line with traditional neural circuitry models of OCD, which focus on frontal cortical-striatal pathways^68,69^. These findings are consistent with and build on previous work linking various neuronal cell types to psychiatric and cognitive phenotypes^70^.

Interestingly, frontal and anterior cingulate cortex, which were enriched in our tissue-based analyses, as well as hippocampus and striatum, which were implicated in our cell-type based analyses, are among the regions that are consistently implicated in neuroimaging studies of OCD^41,71–73^. Enrichment in MSNs in striatum is consistent with their role in the observed aberrant circuitry in OCD, where the medium spiny neurons D1 project to the globus pallidus interna and substantia nigra in the direct pathway, and the D2-type MSNs project to the globus pallidus externa in the indirect pathway^74^. However, medium spiny neurons are also enriched in major depressive disorder^75^, schizophrenia^76^, and intelligence^77^, suggesting that the observed enrichment is not specific for OCD.

Our analyses of the shared genetic risk between OCD and other psychiatric disorders provides further insights into the etiology of OCD. In line with previous observations^38,78^, OCD was significantly genetically correlated with multiple psychiatric disorders and traits. The strongest genetic correlations were observed for anxiety disorders, depression, and anorexia nervosa, all of which are highly comorbid with OCD^79^. This aligns with previous findings from cross-disorder analyses suggesting a shared genetic susceptibility among most psychiatric disorders^38,80,81^. A notable exception is our finding that risk variants for OCD are protective for alcohol dependence^82^, which is at odds with epidemiological evidence strongly linking OCD and alcohol related disorders^83^, but in line with a recent paper^79^ reporting a lower than expected lifetime comorbidity of substance use disorders in OCD. The observed pattern of correlations with other phenotypes can be thought of as falling into two categories: compulsivity/impulsivity and rumination/worry/neuroticism. In both categories, the patterns of genetic correlations appear to follow a gradient across disorders/traits. For example, in the compulsivity/impulsivity category, strong positive correlations are seen with anorexia nervosa and Tourette syndrome, which are disorders with strong compulsive features, with less positive associations seen with ADHD, and negative correlations with alcohol dependence and risk-taking behaviors, which are all phenotypes characterized by impulsivity. A similar gradient is observed for the rumination/worry/neuroticism related phenotypes, with strong positive correlations with anxiety, and other ruminative phenotypes such as worry, transitioning to less strong correlations with individual depression-related items.

This study marks the transition from the flat (sample building) phase of SNP discovery described for GWAS^84^ (**Supplementary Fig. 20**), where few to no genome-wide significant loci are identified^10,12,51,85^, to the linear phase of SNP discovery, where even relatively small increases in sample size identify additional genome-wide significant loci^18^. The strengths of the current study hence include the marked increase in the number of OCD cases and the rigorous analytic methods, including two multivariate approaches (MTAG and GenomicSEM) to control for potential overlapping study participants and to examine potential heterogeneity between the multiple ascertainment approaches. Potential weaknesses include the inability to document comorbid psychiatric disorders in the majority of cases that were not ascertained from clinical collections or electronic registries, the lack of inclusion of non-European ancestries, and the limited availability of sex-chromosome data. Due to the nature of our study, imputation references used in the different cohorts were heterogeneous and did not allow for confident analysis of rare variant associations. Future larger scale sequencing studies that are currently underway will be needed to identify associations in this allele frequency spectrum. We also note that the genetic correlation analyses are impacted by residual heterogeneity in genetic signals due to the employment of heterogeneous ascertainment strategies.

In summary, this work substantially advances the field of OCD genetics by identifying new OCD genetic risk loci and multiple credible candidate causal genes, including those expressed in brain regions and cell types previously implicated in OCD^86^. We have also shown that OCD is highly polygenic in nature, with many variants implicated not only in OCD but also in commonly comorbid disorders or traits, in particular anxiety, neuroticism, anorexia nervosa, and depression. The observation that common variants explain only a modest amount of the phenotypic variation in OCD suggests that other types of genetic variation may also contribute to the etiology of OCD. Notably, whole-exome sequencing studies have suggested that a substantial proportion of OCD cases (22%) may be influenced by rare de novo coding variants^87^, especially in genes that are intolerant to loss of function^88^. Similarly, rare potentially damaging copy number variations represent part of the risk architecture for OCD^9^. These findings emphasize the need for a comprehensive exploration of the contribution of both common and rare genetic factors, as well as their interplay, to OCD risk. Finally, with the implication of the MHC complex, we provide additional evidence for potential shared genetic influences underlying both OCD and increased liability to autoimmune processes, although the directionality of those relationships remains to be definitively elucidated. In addition to continuing to increase sample sizes, future studies will require ancestrally diverse samples to further facilitate the discovery of additional OCD risk variants. Similarly, sex-specific analyses and additional clinical phenotyping will allow for the further elucidation of genetic and clinical relationships between OCD and co-occurring disorders. Finally, with the emergence of drug databases describing the relations between drugs and molecular phenotypes^89^, our results may be useful for drug repurposing (i.e., identifying existing drugs targeting OCD risk genes), leading to new opportunities to find more effective treatments.

## Consortium author lists and affiliations

**23andMe Inc**.

Chris German^268^

^268^23andMe, Inc., Sunnyvale, CA, USA.

**Estonian Biobank**

Andres Metspalu^98^, Tõnu Esko^98^, Reedik Mägi^98^, Mari Nelis^98^, Georgi Hudjashov^98^

## Acknowledgements

We thank the research participants and employees of **all cohorts included in this study** for making this work possible. A list of members of the **23andMe** Research Team, **HUNT, CoGa**, and **MVP** that contributed to this study can be found in **Supplementary Note 1**. We thank the research participants and employees of **23andMe, Inc**. for making this work possible. The **Trøndelag Health Study (HUNT)** is a collaboration between HUNT Research Centre (Faculty of Medicine and Health Sciences, Norwegian University of Science and Technology NTNU), Trøndelag County Council, Central Norway Regional Health Authority, and the Norwegian Institute of Public Health. The genotype quality control and imputation has been conducted by the K.G. Jebsen Center for Genetic Epidemiology, Department of Public Health and Nursing, Faculty of Medicine and Health Sciences, Norwegian University of Science and Technology (NTNU). HUNT analyses were performed in digital labs at HUNT Cloud, HUNT Research Centre Department of Public Health and Nursing, Faculty of Medicine and Health Sciences, Norwegian University of Science and Technology (NTNU), Trondheim, Norway. **NORDiC** is funded by NIMH R01 MH110427 (PI Crowley), NIMH R01 MH105500 (PI Crowley) and the Swedish Research Council grant # 2015– 02271 (PI Mataix-Cols). NORDiC was further supported by the Swedish Research Council (grants 2012-07111 and 2018-02487), Swedish Research Council for Health, Working Life and Welfare 2018-00221 and Center for Innovative Medicine – CIMED. We are deeply grateful for the study participants contributing to the **NORDiC** research. We thank the collection team that worked to recruit them: Anders Juréus, Jessica Pege, Malin Rådström, Radja Satgunanthan-Dawoud, Milka Krestelica, and Birgitta Ohlander, as well as data manager Bozenna Iliadou. We also thank the National Quality Registry for Eating Disorders (RIKSÄT) for help with recruiting patients. We finally thank the BBMRI.se and KI Biobank at Karolinska Institutet for professional biobank service. Grant support for the **MoBa** team was also provided from RCN (273291, 262656, 248778, 223273) and the KG Jebsen Stiftelsen.). MoBa is supported by the Norwegian Ministry of Health and Care Services and the Ministry of Education and Research. We are grateful to all the participating families in Norway who take part in this on-going cohort study. The **AGDS** was primarily funded by National Health and Medical Research Council (NHMRC) of Australia grant 1086683. This work was further supported by NHMRC grants 1145645, 1078901 and 1087889. LCC is supported by a QIMR Berghofer Institute fellowship. We thank all the people who helped in the conception, implementation, beta testing, media campaign and data cleaning of the AGDS data. We specifically acknowledge Dale Nyholt for advice on using the PBS for research; Ken Kendler, Patrick Sullivan, Andrew McIntosh and Cathryn Lewis for input on the questionnaire; Lorelle Nunn, Mary Ferguson, Lucy Winkler and Natalie Garden for data and sample collection; Natalia Zmicerevska, Alissa Nichles and Candace Brennan for participant recruitment support; Jonathan Davies, Luke Lowrey and Valeriano Antonini for support with IT aspects; Vera Morgan and Ken Kirkby for help with the media campaign. We thank VIVA! Communications for their effort in promoting the study. We also acknowledge David Whiteman and Catherine Olsen from QSkin. The work done by the **EstBB** team has received funding from the European Union’s Horizon 2020 Research and Innovation Programme under Grant agreement 847776 (CoMorMent). Data analysis for EstBB was carried out in part in the High-Performance Computing Center of University of Tartu. We thank participants, families and staff of primary and secondary schools who kindly contribute to this research (**Marta Ribasés, Metal-Cat and INSchool**). **EGOS** was supported by a grant from the Beatrice and Samuel A. Seaver Foundation to DEG. The genotyping of **HUNT** was financed by the National Institute of health (NIH), University of Michigan, The Norwegian Research council, and Central Norway Regional Health Authority and the Faculty of Medicine and Health Sciences, Norwegian University of Science and Technology (NTNU). This research is based in part on data from the **Million Veteran Program**, Office of Research and Development, Veterans Health Administration, and was supported by awards CSP575b, I01CX001849-01, 1P1HX002375, and the National Center for PTSD Research. MVP was supported by funding from the Department of Veterans Affairs Office of Research and Development, USVA, grants CSP575B and I01CX001849, MVP-025, and the VA Cooperative Studies Program study, no. 575B; the VA National Center for PTSD Research, and the West Haven VA Mental Illness Research, Education and Clinical Center; and by NIH grant R01 AA026364 (JG). D.F.L. is supported by a Career Development Award CDA-2 from the Veterans Affairs Office of Research and Development (1IK2BX005058-01A2) and is Aimee Mann Fellow of Psychiatric Genetics. This publication does not represent the views of the Department of Veteran Affairs or the United States Government. The **EPOC** study was funded by the Deutsche Forschungsgemeinschaft (DFG; KA815/6-1 and WA731/10-1). **LifeGene** was supported by the Swedish Research Council, the Karolinska Institutet/Stockholm County Council research grants, AFA Insurance and the Torsten and Ragnar Söderbergs Foundation. **GENOS** was supported by the German Research Fundation (GR 1912/1-1). The OCD Collaborative Genetics Association Study (**OCGAS**) is a collaborative research study and was funded by the following NIMH Grant Numbers: MH071507, MH079489, MH079487, MH079488 and MH079494. This work (**OCGAS and IOCDF**) is supported by the Netherlands Organization for Scientific Research-Gravitation project ‘BRAINSCAPES: a Roadmap from Neurogenetics to Neurobiology’ (024.004.012) and the European Research Council advanced grant ‘From GWAS to Function’ (ERC-2018-ADG 834057). The **OCGAS and IOCDF** samples are supported through NIMH Grant Numbers: MH071507 (G.N.), MH079489 (D.A.G.), MH079487 (J.M.), MH079488 (A.F.), and MH079494 (J.K.). The **iPSYCH** team was supported by grants from the Lundbeck Foundation (R102-A9118, R155-2014-1724, and R248-2017-2003), NIH/NIMH (1R01MH124851-01 to A.D.B.) and the Universities and University Hospitals of Aarhus and Copenhagen. The Danish National Biobank resource was supported by the Novo Nordisk Foundation. High-performance computer capacity for handling and statistical analysis of iPSYCH data on the GenomeDK HPC facility was provided by the Center for Genomics and Personalized Medicine and the Centre for Integrative Sequencing, iSEQ, Aarhus University, Denmark (grant to A.D.B.). **Anders D. Borglum** was also supported by the EU’s HORIZON-HLTH-2021-STAYHLTH-01programme, project number 101057385: Risk and Resilience in Developmental Diversity and Mental Health (R2D2-MH). **Mental-Cat and INSchool** was supported by the Agència de Gestió d’Ajuts Universitaris i de Recerca (AGAUR, 2017SGR-1461, 2021SGR-00840), the Instituto de Salud Carlos III (PI20/00041, PI23/00404 and PI23/00026), the European Regional Development Fund (ERDF); the ECNP Network ‘ADHD across the Lifespan’; “la Marató de TV3” (202228-30 and 202228-31). **BioVU**: CTSA (SD, Vanderbilt Resources) was supported by the National Center for Research Resources, Grant UL1 RR024975-01, and is now at the National Center for Advancing Translational Sciences, Grant 2 UL1 TR000445-06. The content is solely the responsibility of the authors and does not necessarily represent the official views of the NIH.The dataset(s) used for the analyses (BioVU) described were obtained from Vanderbilt University Medical Center’s BioVU which is supported by numerous sources: institutional funding, private agencies, and federal grants. These include the NIH funded Shared Instrumentation Grant S10RR025141; and CTSA grants UL1TR002243, UL1TR000445, and UL1RR024975. Genomic data are also supported by investigator-led projects that include U01HG004798, R01NS032830, RC2GM092618, P50GM115305, U01HG006378, U19HL065962, R01HD074711; and additional funding sources listed at https://victr.vumc.org/biovu-funding/. **Silvia Alemany** acknowledges Miguel Servet contract (CP22/00026) awarded by the Instituto de Salud Carlos III and co-funded by the European Union Found: Fondo Social Europeo Plus, FSE +.HYPERGENES and InterOmics cohorts provided controls of Italian origin for the present study. **Judit Cabana-Dominguez** acknowledges her contract from the Network Center for Biomedical Research (CIBER). **Richard Delorme** acknowledges the Clinical Investigation Centre, Robert Debré Hospital. INSERM @ APHP granted the study. **Bengt Fundin** acknowledges The Anorexia Nervosa Genetics Initiative (ANGI), an initiative of the Klarman Family Foundation. **Jan Haavik** acknowledges the Trond Mohn Foundation, Bergen, Norway. **Christine Lochner** acknowledges the South African Medical Research Council and the National Research Foundation for their support. **Thomas V Fernandez:** Research reported in this publication was supported by the National Institute of Mental Health of the National Institutes of Health under Award Number R01MH114927. The content is solely the responsibility of the authors and does not necessarily represent the official views of the National Institutes of Health. **Nicholas G. Martin** has received funding from a project grant from Australian NHMRC. The Research Council of Norway supported **H. Ask, A. Havdahl** and **T. Reichborn-Kjennerud** (274611). A. Havdahl was also supported by South East Norway Health Authority (2020022). **Zachary F. Gerring** is supported by an Australian NHMRC EL1 Investigator Grant (2034743) and NIH/NIA Grant AG068026. **Marco Galimberti** received support from the following grants (Joel Gelernter): CSP575b, I01CX001849-01, 1P1HX002375, National Center for PTSD Research, 5R01DA054869-01. **Abdel Abdellaoui** was supported by the Foundation Volksbond Rotterdam. **Tim Bigdeli** is supported by NIMH grant 7R01MH103657 (GPC-OCD). **Jonathan Coleman:** This study represents independent research partly funded by the National Institute for Health Research (NIHR) Maudsley Biomedical Research Centre at South London and Maudsley NHS Foundation Trust and King’s College London. The views expressed are those of the authors and not necessarily those of the NHS, the NIHR or the Department of Health and Social Care. **Christina Barlassina** was supported by grant EU FP7-HEALTH-2007-A-201550, and grant MIUR-CNR PB05. **Enda Byrne** was supported by the NHMRC Project Grant 1145645; University of Queensland Health Research Accelerator Program (HeRA). **Carolina Cappi** was supported by grant K99MH128540-01A1. **Valentina Ciullo** was supported by the Italian Ministry of Health grant RC-18-19-20-21/A. **Marco A. Grados** was supported by NIMH K23 MH066284. **Jan Haavik** was supported by Stiftelsen KG Jebsen (SKGJ MED-02). **Kristen Hagen** was supported by the Trond Mohn Foundation. **Elinor K. Karlsson** was supported by NIH R21 MH109938. **Paul S. Nestadt** was supported by R01MH071507. **Fabrizio and Federica Piras** are supported by the Italian Ministry of Health RC18-19-20-21/A grant. This work was in part supported by the German Research Foundation (DFG) grants: [RA1971/8-1], [RA1971/7-1]; and by the Bundesministerium für Bildung und Forschung (BMBF) grant: 01ED2007A to **Alfredo Ramirez. Stephan Ripke** was supported by research grant 1R01MH124873-01. **Maria Soler Artigas** was supported by The Instituto de Salud Carlos III (P19/01224, PI22/00464 and CP22/00128) and the European Regional Development Fund (ERDF). **Arpana Agrawal** was supported by grant U10AA008401. **Pino Alonso** was supported by the Spanish Ministry of Science, Innovation and Universities (ISCIII PI22/00752) and Fundació La Marató 202201-30. **Cynthia M. Bulik** was supported by R01 MH124871 (Sullivan/Bulik) PGC4. **Howard Edenberg** was supported by grant U10AA008401. **Dan A. Geller** was supported by NIMH (OCGAS and OCGS). **Gregory L. Hanna** was supported by the National Institute of Mental Health (R01 MH58376), National Institute of Mental Health (K20 MH01065), National Institute of Mental Health (R01 MH101493), National Institute of Mental Health (R01 MH085321) **Norbert Kathmann** has received funding from Deutsche Forschungsgemeinschaft (DFG) KA815/6-1. **Sarah E. Medland** is supported by an Australian NHMRC Investigator Grant (APP1172917). **Benjamin M. Neale** is funded by grant R01MH124851. **Michele Pato and Carlos Pato** have received support from R01MH103657 and R01MH079494 from the National Institutes of Mental Health (NIMH) and the Della Martin Foundation, Los Angeles CA. **John Piacentini** has received support through the National Institute of Mental Health: R01MH50214: Collaborative OCD Genetics Study (G. Nestadt, PI; J. McCracken, UCLA PI). **Margaret A. Richter** was supported by funding from the Canadian Institutes for Health Research and the Ontario Mental Health Foundation. **David R. Rosenberg** was supported by NIMH R01MH059299. **Jack F. Samuels** was supported by NIMH Grant Number: MH071507. **Gianfranco Spalletta** is supported by the Italian Ministry of Health RC18-19-20-21/A grant. **Eric A. Storch** collected data as part of the following NIH grant: 1R01MH093381. **Ole A. Andreassen** (MoBa) has received grant support from RCN (324499,273291,262656,248778,223273), KG Jebsen Stiftelsen, NordForsk #164218. **Jaakko Kaprio** has been supported by the Academy of Finland (grant 336823). **Paul D. Arnold** is supported by the Alberta Innovates Translational Health Chair in Child and Youth Mental Health. **Dorothy E. Grice** is supported by the grant MH124679-01. **James A. Knowles** is supported through the following grants: R01MH103657 and R01MH079494 from the National Institutes of Mental Health (NIMH) and the Della Martin Foundation, Los Angeles CA. **Karin J. H. Verweij** is supported by the Foundation Volksbond Rotterdam. **Lea K. Davis** was supported by grants from the National Institutes of Health including R01NS102371, R01MH113362, R01MH118223, R01NS105746, and R56MH120736. J.S. was supported by a NIH training grant in Human Genetics, 2T32GM080178. **James Crowley** was supported by NIH grants R01MH105500 and R01MH110427. **Murray B. Stein** has been funded by the Veterans Affairs Administration (United States VA).

## Author Contributions

J.M. Scharf, M.B.S., J. Gelernter, C.A.M., E.M.D. and M. Mattheisen designed the study. N.I.S., Z.F.G., M.G., D.Y. and M.W.H. conducted data analysis. N.I.S., Z.F.G., M.G., D.Y., M.W.H., A. Abdellaoui, C.R.-F., J.M. Sealock, T.B., J.R.C., B. Mahjani, J.G.T., K.B., C.L.B., J.J.L., G.Z., S.A., C.A., K.D.A., J.B., N.B., C.B., J.B.N., O.J.B., D.B., M.H.B., S.B., R.B., M.B., B.P.B., H.B., J.D.B., J.B.-G., E.M.B., J.C.-D., B.C., A. Camarena, C.C., A. Carracedo, M.C., M.C.C., V.C., E.H.C., J.C., B.A.C., E.J.D.S., R.D., S.D., J.A.E., X.E., M.J.F., B.T.F., L.G., C.G., F.S.G., M.A.G., J. Grove, W.G., J.H., K. Hagen, K. Harrington, A.H., K.D.H., A.G.H., D.H., C.H., M.J., E.J., E.K.K., K. Kelley, J. Klawohn, J.E.K., K. Krebs, C. Lange, N.L., D. Levey, K.L.-T., F.M., B. Maher, B. Mathes, E.M., N.M., N.C.M., S.M., E.C.M., M. Mulhem, P.S.N., E.L.N., K.S.O., L.O., O.T.O., T.P., N.L.P., Fabrizio Piras, Federica Piras, S.P., R.R., A. Ramirez, S. Rauch, A. Reichenberg, M.A. Riddle, S. Ripke, M.C.R., A.S.S., M.A.S., A.H.S., L.G.S., J.S., M.S.A., L.F.T., E.T., H.V., N.v.K., J.V.-V., N.N.V., C.P.W., Y.W., J.R.W., B.S.W., Y.Y., H.Z., A. Agrawal, P.A., G. Berberich, K.K.B., C.M.B., D.C., D.D., V.E., H.E., P.F., T.V.F., A.J.F., J.M.G., D.A.G., H.J.G., B.D.G., G.L.H., I.B.H., D.M.H., N.K., J. Kennedy, D. Lai, M. Landén, S.L.H., M. Leboyer, C. Lochner, J.T.M., S.E.M., P.B.M., B.M.N., H.N., M.N., M.P., C. Pato, D.L.P., J.P., C. Pittenger, D.P., J.A.R.-Q., S.A.R., M.A. Richter, D.R.R., S. Ruhrmann, J.F.S., S.S., P.S., G.S., D.J. Stein, S.E.S., E.A.S., B.E.S., M.T., T.W., O.A.A., A.D.B., S.W., K. Hveem, B.K.H., C.R., N.G.M., L.M., O.M., T.R.-K., M.R., G.K., D.M.-C., K.D., E.G., M.W., J.-A.Z., G. Breen, G.N., J. Kaprio, P.D.A., D.E.G., J.A.K., H.A., K.J.V., L.K.D., D.J. Smit, J.J.C., J.M. Scharf, M.B.S., J. Gelernter, C.A.M., E.M.D. and M. Mattheisen provided samples and/or processed individual cohort data. N.I.S., Z.F.G., M.G., D.Y., M.W.H., J.M. Scharf, M.B.S., J. Gelernter, C.A.M., E.M.D. and M. Mattheisen wrote the paper and formed the core revision group. J.M. Scharf, M.B.S., J. Gelernter, C.A.M., E.M.D. and M. Mattheisen supervised and directed the study. All authors discussed the results and approved the final version of the manuscript.

## Methods

### Ethics

All relevant ethics approvals have been obtained by the respective cohort’s institutions and a list of all respective approvals can be found in **Supplementary Note 2**.

### Study participants

We analyzed genomic data from 28 OCD case-control cohorts including 53,660 OCD cases and 2,044,417 controls of European ancestry. **Supplementary Table 1** provides an overview of the individual cohorts. A subset of the cases and controls have been included in previous studies^10,51,85^ and preprints^12,13^, as described in **Supplementary Note 2**. Among all included individuals, 323 cases were part of a parent-proband trio; in these cases, parents were used as pseudo-controls. A total of 20,427 cases met DSM-5^93^ or ICD-10^94^ criteria for OCD as assessed by a healthcare professional or derived from (electronic) health records, while the remaining 32,233 cases were based on self-reported OCD diagnosis (23andMe, AGDS, and parts of UKBB). Cohort-specific sample and analytic details can be found in **Supplementary Note 2**. Data collections were approved by the relevant institutional review boards at all participating sites, and all participants provided written informed consent.

### Individual GWAS analyses and harmonizing of results

First, the data of each participating cohort were analyzed individually (see **Supplementary Note 2** for details). Genetic data were imputed using either the Haplotype Reference Consortium (HRC)^95^ or 1000 Genomes Project Phase 3 reference panels^96^. The resulting GWAS summary statistics were then harmonized before a conjoint meta-analysis of all autosomes was conducted. Each summary statistic data set was transformed to ‘daner’ file format following RICOPILI^97^ specifications. All variants had to meet the following criteria for inclusion: minor allele frequency (MAF) > 1% in cases and controls, imputation quality (INFO) score > 0.8 and < 1.2. If the effect measure, *P*-value or standard error (s.e.) was missing or was out of bounds (infinite), the SNP was removed. Once cleaned summary statistics were produced, all datasets were aligned to the HRC reference panel. If variants were reported on different strands, they were flipped to the orientation in the HRC-reference. Furthermore, strand-ambiguous A/T and C/G SNPs were removed if their MAF was > 0.4. In the case that A/T and C/G SNPs showed a MAF < 0.4, allele frequencies were compared to frequencies in the HRC-reference. If an allele frequency match was found, i.e., minor alleles were the same in the summary statistics and the HRC reference, the same strand orientation was assumed. If an allele mismatch was found, i.e., the allele had a frequency > 0.5 in HRC, it was assumed that alleles were reported on different strands and alleles were flipped subsequently. Marker-names were uniformly switched to those present in the HRC reference. If a variant did not overlap with the variants in the HRC reference, it was removed.

### GWAS meta-analysis

Inverse variance weighted meta-analysis was conducted on 28 European cohorts using METAL^98^. Weighting was based on standard error primarily to account for the large case/control imbalances in cohorts that used linear mixed model approaches in their primary GWAS. Heterogeneity was assessed with Cochran’s *Q* statistic and *I*^2^ statistic^99,100^ (see **Supplementary Note 5** for details). The genomic control factor lambda (λ) was calculated for each individual GWAS and for the overall meta-analysis to identify residual population stratification or systematic technical artifact. GWAS summary statistics were subjected to linkage disequilibrium (LD) score regression (LDSC) analyses on high-quality common SNPs (INFO score > 0.9) to examine the LDSC intercept to distinguish polygenicity from other types of inflation, and to estimate the genetic heritability from the meta-analysis and genetic correlations between cohorts. The genomic inflation factor lambda (λ) was estimated at 1.330 with an λ_1000_ of 1.033, while the LDSC intercept was 1.0155 (*s. e*. = 0.0085), indicating that the inflation was mostly due to polygenic signal and unlikely to be significantly confounded by population structure. The genome-wide significance threshold for the GWAS was set at a *P*-value of 5.0 × 10^−8^. The 23andMe data included information on the X-chromosome; as this information was not present for all other cohorts, analysis of the X-chromosome was only conducted in this sub-cohort (see **Supplementary Note** 4 for details).

We further conducted GWAS meta-analyses on four subgroups, defined by differences in their sample ascertainment, either (i) clinical-OCD cases diagnosed by a health care professional in a clinical setting (*n*_*cases*_ = 9,089, *n*_*controls*_ = 21,077; including IOCDF, IOCDF_trio, EPOC, NORDiC-nor, NORDiC-swe, EGOS, OCGAS, OCGAS-ab, OCGAS-gh, OCGAS-nes, Psych_Broad, WWF, MVP, Michigan/Toronto IGS, YalePenn, Chop, CoGa), (ii) comorbid-individuals that were primarily ascertained for another comorbid psychiatric disorder (*n*_*cases*_ = 5,266, *n*_*controls*_ = 43,760; AGDS, IPSYCH), (iii) biobank data from large-scale biobanks or registries with ICD or DSM codes (*n*_*cases*_ = 9,138, *n*_*controls*_ = 1,049,776; BioVU, EstBB, FinnGen, HUNT, MoBa, UKBB) or (iv) 23andMe data (*n*_*cases*_ = 30,167, *n*_*controls*_ = 929,804). While these groups are not exclusive (e.g., diagnoses in health records were originally given in a clinical setting, or comorbid cases were also assessed in a clinical setting or derived from health records), we defined these groups by the cohort’s primary characteristic. We also conducted one meta-analysis including all clinical, comorbid, and biobank subgroups, while excluding the 23andMe data, resulting in 23,493 cases and 1,114,613 controls. As 23andMe is the only consumer-based dataset, we intended to compare this dataset to all others.

### Number of Trait specific causal Variants (MiXeR analysis)

We applied MiXeR v1.3^15^ to quantify the polygenicity of OCD (i.e., estimate the total number of trait-influencing genetic variants). MiXeR fits a Gaussian mixture model assuming that common genetic effects on a trait are a mixture of causal variants and noncausal variants. Polygenicity is reported as the number of the causal variants that explain 90% of SNP heritability of OCD (to avoid extrapolating model parameters into the area of infinitesimally small effects).

### SNP-based fine-mapping - GCTA-COJO

We performed a conditional-and-joint analysis (GCTA-COJO)^14^ to identify independent signals within significant OCD loci. This approach performs a conditional and joint analysis on the basis of conditional *P*-values before calculating the joint effects of all selected SNPs. We used the stepwise model selection procedure to select independently associated SNPs. The linkage disequilibrium reference sample was created from 73,005 individuals from the QIMR Berghofer Medical Research Institute genetic epidemiology cohort. The distance assumed for complete linkage disequilibrium was 10 Mb, and we used the default *P*-value threshold of 5 × 10^−8^ to define a genome-wide significant hit.

### Multi-trait analysis of ascertainment subgroups

We used multi-trait analysis of GWAS (MTAG)^101^ to conduct multivariable GWAS analyses, reporting GWAS results for each of the ascertainment-specific sub-groups. Through this approach, we aimed to address potential concerns about heterogeneity in genetic liability for individual sub-groups following different ascertainment strategies. MTAG is a multi-trait analysis that is usually used to combine different but related traits into one meta-analysis by leveraging the shared heritability among the different traits and thereby gaining power. In this case, our aim was to generate ascertainment-specific estimates, while boosting power by leveraging the high shared heritability between the subgroups. The MTAG analysis resulted in four different GWAS summary statistics, one for each subgroup (clinical, comorbid, biobanks, 23andMe). We performed maxFDR analyses to approximate the upper bound on the FDR of MTAG results.

### GenomicSEM

Similarly, we used genomic structural equation modeling (GenomicSEM^16^) to model the joint genetic architecture of the four subgroups. First, we ran a common-factor model without individual SNP effects, following the tutorial ‘Models without individual SNP effects’ on the GenomicSEM github website (see web resources). Second, we ran a multivariate GWAS of the common factor (see **Supplementary Note 5** for details). We specified the model using unit-variance identification, for which the latent factor variance was fixed to 1 and the loadings of the traits are estimated freely. This ensures that we capture how much of each subgroup contributes to the latent factor. GenomicSEM also generates *Q*_*SNP*_-values, which indicate possible heterogeneous effects across the subgroups. The *Q*_*SNP*_ statistic is mathematically similar to the Q-statistic from standard meta-analysis and is a *X*^2^-distributed test statistic with larger values indexing a violation of the null hypothesis that the SNP acts entirely through the common factor.

### SNP heritability estimation

The proportion of the phenotypic variance that could be explained by the aggregated effect of all included SNPs (SNP-based heritability, 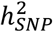 was estimated using LDSC^31^. The analysis was performed using pre-computed LDscores from samples restricted to European-ancestry in the 1000 Genomes Project^96^, filtered for SNPs included in the HapMap3 reference panel^102^. SNP heritability was estimated based on the slope of the LDSC, with heritability on the liability scale calculated assuming a 1% population prevalence of OCD^1^. To omit a downward bias in our estimates of liability scale heritability, following Grotzinger *et al*.^103^, we accounted for varying levels of ascertainment across cohorts in our meta-analysis by summing the effective sample sizes across the contributing cohorts and using that as the input sample size for LDSC. For the conversion to the liability scale (1%), the sample prevalence was then specified as 0.5. The SNP-heritability was calculated for the whole OCD sample as well as for ascertainment-specific sub-groups.

### Genetic correlations

We used cross-trait LDSC^31^, a method that computes genetic correlations between GWASs without bias from ancestry differences or sample overlap to calculate genetic correlations between the primary OCD meta-analysis and other phenotypes of interest. The selection of traits was based on phenotypic relevance and/or prior report of a genetic relationship with OCD. The genetic correlation between traits is based on the estimated slope from the regression of the product of *Z*-scores from two GWASs on the LD score and represents the genetic covariation between two traits based on all polygenic effects captured by the included SNPs. The genome-wide LD information used by these methods was based on European populations from the HapMap 3 reference panel^102^, and GWAS summary statistics were filtered to only include SNPs that were part of the 1,290,028 HapMap 3 SNPs.

To ensure the internal consistency of the datasets included in our meta-analysis, we calculated genetic correlations between all cohorts we considered to have a sample size large enough for LDSC (effective sample size of ≥ 1,000) and between the four ascertainment-specific subgroups.

We further calculated genetic correlations between OCD and 112 other disorders and traits. The source studies of the GWAS summary statistics can be found in **Supplementary Table 18**. As a follow-up, we also calculated genetic correlations between the 112 phenotypes and each ascertainment-specific sub-cohort and compared the genetic correlation patterns between the four groups. For all cross-phenotype genetic correlation analyses, we adjusted *P*-values for multiple testing using the Benjamini-Hochberg procedure to control for FDR (< 0.05).

### Gene-based analyses

To match the significant SNPs to the genes whose function they likely influence, we conducted a series of positional and functional gene mapping analyses. The positional mapping employed MBAT-combo^19^, while the functional mapping tested whether genetic variants associated with OCD were also associated with differential expression of nearby genes (within 1-Mb window) using (i) transcriptome-wide association study (TWAS)^20^ utilizing PsychENCODE data and included colocalization with COLOC^27,28^ and (ii) summary-based Mendelian randomization (SMR^22^) using whole-blood eQTL information and brain tissues from MetaBrain, alongside the HEIDI test which tests for heterogeneity in GWAS signal and eQTL association. Further, a protein-wide association study (PWAS) was conducted. As a final step, genes within each locus were prioritized using PsyOPS^26^, which integrates both positional and functional information. The details of each method are described below.

### Positional gene mapping (MBAT-combo)

A gene-based analysis was conducted using multivariate Set-Based Association Test (mBAT-combo)^19^ within GCTA version 1.94.1^14^. The European subsample (*n* = 503 individuals) from Phase 3 of the 1000 Genomes Project^96^ was used as the LD reference panel with the fastBAT default LD cut-off of 0.9 applied. After filtering SNPs with MAF > 0.01, there were 6,629,124 SNPs for analysis in our sample. A gene list consisting of 19,899 protein coding genes was used to map the base pair position of genes using genome build hg19 (see **Supplementary Note 7** for details).

### Functional gene mapping

#### Transcriptome-wide association study (TWAS)

We used TWAS FUSION^20^ to perform a transcriptome-wide association study of OCD. We used brain gene expression weights from the PsychENCODE (104) and LD information from the 1000 Genomes Project Phase 3^96^. TWAS FUSION uses reference LD and reference gene expression panels with GWAS summary statistics to estimate the association between gene expression and OCD risk. These data were processed with the test statistics from the OCD GWAS to estimate the expression-GWAS association statistic. We corrected for multiple testing using Bonferroni correction.

We performed colocalization analyses using the COLOC R function^27,28^ implemented in TWAS FUSION. Colocalization is a Bayesian method used to calculate the posterior probabilities (PP) that individual lead SNPs within a significant TWAS locus are (i) independent (e.g., two causal SNPs in LD, one affecting transcription, and one affecting OCD; posterior probability (PP3)) or (ii) share the same associated variant (e.g., a single causal SNP affects both transcription and OCD (PP4)). We also performed a conditional analysis to determine whether identified associations represented independent associations. This was performed using the FUSION software, which jointly estimates the effect of all significant features within each locus by using residual SNP associations with OCD after accounting for the predicted expression of other features.

#### Summary-based Mendelian randomization (SMR)

SMR^22^ was performed using default settings and eQTL meta-analysis summary statistics from European populations for whole blood from eQTLGen^23^, and all five nervous system tissues from MetaBrain (basal ganglia, cerebellum, cortex, hippocampus and spinal cord)^105^. The HEIDI (heterogeneity in dependent instruments) test is performed alongside SMR to test for effect size heterogeneity between the GWAS and eQTL summary statistics. Both SMR and TWAS have a number of important assumptions and limitations, which we discuss in **Supplementary Note 9**.

### Psychiatric omnilocus prioritization score

We used the gene prioritization method PsyOPS (Psychiatric Omnilocus Prioritization Score)^26^ to rank genes within genome-wide significant loci. This supervised approach integrates biological annotations on mutational intolerance, brain-specific expression, and involvement in neurodevelopmental disorder for genes within significant loci. Genes with the top PsyOPS score within each locus were used for further gene prioritization (see “Gene prioritization” below). In the instance where two genes in the same locus had the same PsyOPS score, the gene nearest the index SNP was prioritized.

### Protein-wide association study (PWAS)

We performed a protein-wide association study (PWAS) using protein expression data from human brain samples. Human brain proteome reference weight data were obtained using the Religious Orders Study and Rush Memory and Aging Project (ROS/MAP) and Banner Sun Health Research Institute (Banner) study. The ROS/MAP proteomes were generated from the dorsolateral prefrontal cortex (DLPFC) of 376 participants of European ancestry and included 1,476 proteins with significant SNP-based heritability (*P* < 0.01). The Banner PWAS weights were generated from 152 individuals of European ancestry and included 1,147 proteins with significant SNP-based heritability. The PWAS was performed using the TWAS FUSION software^20^ with LD reference information from the 1000 Genomes Project Phase 3^96^. We corrected for multiple testing using Bonferroni correction.

### Gene prioritization

We created a list of prioritized genes using both gene-based tests and colocalization/HEIDI filters. Results from each gene-based test were first restricted to protein coding genes with unique gene identifiers based on the release from GENCODE (v40) for hg19. The following criteria were then used to prioritize genes: (i) a significant (Bonferroni corrected) association from at least two gene-based tests (mBAT-combo, TWAS FUSION, SMR, or PsyOPS), and (ii) evidence of colocalization (COLOC PP4 > 0.8) and/or significant SMR association with HEIDI *P* > 0.05. Joint/conditional tests of association and significant PWAS associations were used as ancillary approaches to further annotate the prioritized gene list.

### Tissue and cell-type enrichment analysis

An analysis of tissue and cell-type enrichment of OCD GWAS association signals was conducted using MAGMA (v1.08)^106^ and partitioned LD score regression^107^. We employed Bryois et al.’s approach^29^ to determine gene expression specificity in bulk tissue RNA-seq data from 37 tissues in GTEx (v8) and single-cell RNA sequencing data from 19 regions in the mouse central and peripheral nervous systems^30^. The analysis was limited to protein-coding genes with 1:1 orthologs between mice and humans. Gene expression in each tissue or cell type was calculated relative to total expression across all tissues or cell types. Enrichment analysis was performed on genes with the top 10% specificity values in each tissue or cell type, as previously defined^29^.

To evaluate the enrichment of tissue and cell type specific genes in OCD genetic association signals, we applied MAGMA and partitioned LDSC. We restricted the analysis to summary statistics for SNPs with a high INFO score (> 0.6) and frequency in the entire cohort (MAF > 0.01). Using MAGMA (v1.08), we tested if genes with the top 10% specificity in a tissue or cell type showed enrichment in gene-level genetic associations for OCD, with the 1000 Genomes Phase 3 European sample genotypes serving as the LD reference panel. We used standard gene boundaries (35 kb upstream of the transcription start site to 10 kb downstream of the transcription stop site). Partitioned LDSC was used to examine whether SNPs within 100-kb regions of the top 10% specifically expressed genes were enriched for SNP-based heritability for OCD. All results were corrected for multiple testing with an FDR threshold of 0.05.

### SNP and gene findings in the context of previous analyses

#### Previously reported associations for significant SNPs (PheWAS)

Multiple resources were used to identify previously reported associations of our 30 significant SNPs with other phenotypes. We used the IEU open gwas project^92^, PheWAS analysis of gwasATLAS^52^, the NHGRI-EBI GWAS Catalog^91^, and identified credible SNPs through causaldb^90^. Causaldb estimates causal probabilities of all genetic variants in GWAS significant loci using three state-of-the-art fine-mapping tools including PAINTOR, CAVIARBF and FINEMAP^108–111^. We used default settings for our causaldb queries.

#### Lookup of previous OCD GWAS findings

We performed a look-up of SNPs, identified to be significantly associated with OCD-related phenotypes in previous GWASs. Note that this is not an independent replication as previous studies partially overlap with the cohorts included in this GWAS.

#### Overlap of previous rare coding variants in OCD and GWAS gene-findings

We performed a bi-directional look-up, assessing (i) whether gene-findings from our GWAS showed evidence for rare variant involvement and (ii) vice versa, whether findings from rare variant testing showed evidence of common variant association in our GWAS.

First, we comprehensively assessed the overlap between 251 genes that we highlighted in our manuscript as carrying common risk variation for OCD (**Supplementary Table 14**), and current gene-based summary statistics from OCD exome sequencing data. We utilized data from Halvorsen *et al*.^88^ since it is the largest published exome sequencing study of OCD presently. The supplemental materials from that paper include *de novo* variant calls from 771 case trios and 1,911 controls (Table S14 in Halvorsen *et al*.^88^). We compared the burden of *de novo* variants, partitioned by variant annotation (synonymous, missense, loss of function) in trio cases versus trio controls within these 251 GWAS-prioritized genes. As was done in the Halvorsen et al.^88^, we only included de novo variants that were in loci well-covered in both case and control data (In_Jointly_Covered_Loci==TRUE). We also excluded all calls from quartet samples in Halvorsen et al. (Cohort!=“OCD_JHU_quartets”). For each of the four variant annotation classes, we compared the proportion of cases that have at least one qualifying *de novo* variant to the proportion of controls using a two-sided Fisher’s exact test.

Second, since Halvorsen *et al*.^88^ describe an overall excess of loss of function variants in OCD cases relative to controls specifically within loss of function intolerant genes (Table S4 in Halvorsen *et al*.^88^), we analyzed the overlap between those genes and our GWAS-derived genes. We looked up 200 genes with a probability of loss of function intolerance > 0.995 (derived from Lek *et al*.^112^) and effect size estimate > 1. We further tested for a difference in the proportion of these pLI > 0.995 genes with effect size estimate >1 vs ≤1 within the set of genes highlighted in the OCD GWAS (*n* = 251) vs. outside this set using a two-sided Fisher’s exact test.

## Data availability

The meta-analyzed summary statistics (not including 23andMe data) are available from the Psychiatric Genomics Consortium Download page (https://www.med.unc.edu/pgc/download-results/). In line with 23andMe regulations, 10,000 SNPs from the full GWAS including 23andMe are also being made available at the https://www.med.unc.edu/pgc/download-results/.

The full GWAS summary statistics for the 23andMe discovery data set will be made available through 23andMe to qualified researchers under an agreement with 23andMe that protects the privacy of the 23andMe participants. Datasets will be made available at no cost for academic use. Please visit https://research.23andme.com/collaborate/#dataset-access/ for more information and to apply to access the data.

MVP summary statistics are made available through dbGAP request under accession phs001672.v12.p1.

## Code availability

Core analysis code for RICOPILI can be found at https://sites.google.com/a/broadinstitute.org/ricopili/. This includes PLINK (https://www.cog-genomics.org/plink2/), EIGENSOFT (https://www.hsph.harvard.edu/alkes-price/software/), Eagle2 (https://alkesgroup.broadinstitute.org/Eagle/), Minimac3 (https://genome.sph.umich.edu/wiki/Minimac3), SHAPEIT3 (https://mathgen.stats.ox.ac.uk/genetics_software/shapeit/shapeit.html), METAL (https://genome.sph.umich.edu/wiki/METAL_Documentation) and LDSR (https://github.com/bulik/ldsc). MAGMA can be found at https://ctg.cncr.nl/software/magma. Genomic SEM, specifically the tutorial ‘Models without Individual SNP effects’ can be found here: https://github.com/GenomicSEM/GenomicSEM/wiki/3.-Models-without-Individual-SNP-effects

TWAS FUSION: http://gusevlab.org/projects/fusion/

PWAS: For access to the protein weights, see: https://www.synapse.org/#!Synapse:syn24872746

GCTA (mBAT-combo and COJO): https://yanglab.westlake.edu.cn/software/gcta/#Overview

LDSC and partitioned heritability: https://github.com/bulik/ldsc

Additional code for data processing (e.g., harmonization of summary statistics) can be found at https://www.doi.org/10.6084/m9.figshare.28451894

## References

1. Fawcett, E. J., Power, H. & Fawcett, J. M. Women are at greater risk of OCD than men: a meta-analytic review of OCD prevalence worldwide. J. Clin. Psychiatry 81, 19r13085 (2020).

2. World Health Organization. The Global Burden of Disease: 2004 Update. Geneva: WHO Press (2008).

3. Fernández de la Cruz, L. et al. Suicide in obsessive–compulsive disorder: a population-based study of 36 788 Swedish patients. Mol. Psychiatry 22, 1626–1632 (2017).

4. Meier, S. M., Mattheisen, M., Mors, O., Schendel, D. E., Mortensen, P. B. & Plessen, K. J. Mortality among persons with obsessive-compulsive disorder in Denmark. JAMA Psychiatry 73, 268–274 (2016).

5. Blanco-Vieira, T., Radua, J., Marcelino, L., Bloch, M., Mataix-Cols, D. & do Rosário, M. C. The genetic epidemiology of obsessive-compulsive disorder: a systematic review and meta-analysis. Trans.l Psychiatry 13, 230 (2023).

6. Burton, C. L. et al. Heritability of obsessive–compulsive trait dimensions in youth from the general population. Transl. Psychiatry 8, 191 (2018).

7. Pauls, D. L. The genetics of obsessive compulsive disorder: a review of the evidence. Am. J. Med. Genet. C Semin. Med. Genet. 148, 133–139 (2008).

8. van Grootheest, D. S., Cath, D. C., Beekman, A. T. & Boomsma, D. I. Twin studies on obsessive–compulsive disorder: a review. Twin Res. Hum. Genet. 8, 450–458 (2005).

9. Mahjani, B. et al. The genetic architecture of obsessive-compulsive disorder: contribution of liability to OCD from alleles across the frequency spectrum. Am. J. Psychiatry 179, 216–225 (2022).

10. International Obsessive Compulsive Disorder Foundation Genetics Collaborative (IOCDF-GC) and OCD Collaborative Genetics Association Studies (OCGAS). Revealing the complex genetic architecture of obsessive-compulsive disorder using meta-analysis. Mol. Psychiatry 23, 1181–1188 (2018).

11. Davis, L. K. et al. Partitioning the heritability of Tourette syndrome and obsessive compulsive disorder reveals differences in genetic architecture. PLoS Genet. 9, e1003864 (2013).

12. Strom, N. I. et al. Genome-wide association study identifies new locus associated with OCD. medRxiv (10.1101/2021.10.13.21261078).

13. Strom, N. I. et al. Genome-wide association study identifies new loci associated with OCD. medRxiv (10.1101/2024.03.06.24303776).

14. Yang, J., Lee, S. H., Goddard, M. E. & Visscher, P. M. GCTA: a tool for genome-wide complex trait analysis. Am. J. Hum. Genet. 88, 76–82 (2011).

15. Holland, D. et al. Beyond SNP heritability: polygenicity and discoverability of phenotypes estimated with a univariate Gaussian mixture model. PLoS Genet. 16, e1008612 (2020).

16. Grotzinger, A. D. et al. Genomic structural equation modelling provides insights into the multivariate genetic architecture of complex traits. Nat. Hum. Behav. 3, 513–525 (2019).

17. Wray, N. R. et al. Genome-wide association analyses identify 44 risk variants and refine the genetic architecture of major depression. Nat. Genet. 50, 668–681 (2018).

18. Strom, N. I. et al. Genome-wide association study of major anxiety disorders in 122,341 European-ancestry cases identifies 58 loci and highlights GABAergic signaling. medRxiv (10.1101/2024.07.03.24309466).

19. Li, A. et al. mBAT-combo: a more powerful test to detect gene-trait associations from GWAS data. Am. J. Hum. Genet. 110, 30–43 (2023).

20. Gusev, A. et al. Integrative approaches for large-scale transcriptome-wide association studies. Nat. Genet. 48, 245–252 (2016).

21. Gandal, M. J. et al. Shared molecular neuropathology across major psychiatric disorders parallels polygenic overlap. Science 359, 693–697 (2018).

22. Zhu, Z. et al. Integration of summary data from GWAS and eQTL studies predicts complex trait gene targets. Nat. Genet. 48, 481–487 (2016).

23. Võsa, U. et al. Large-scale cis- and trans-eQTL analyses identify thousands of genetic loci and polygenic scores that regulate blood gene expression. Nat. Genet. 53, 1300–1310 (2021).

24. Qi, T. et al. Identifying gene targets for brain-related traits using transcriptomic and methylomic data from blood. Nat. Commun. 9, 2282 (2018).

25. Wingo, T. S. et al. Brain proteome-wide association study implicates novel proteins in depression pathogenesis. Nat. Neurosci. 24, 810–817 (2021).

26. Wainberg, M., Merico, D., Keller, M. C., Fauman, E. B. & Tripathy, S. J. Predicting causal genes from psychiatric genome-wide association studies using high-level etiological knowledge. Mol. Psychiatry 27, 3095–3106 (2022).

27. Giambartolomei, C. et al. Bayesian test for colocalisation between pairs of genetic association studies using summary statistics. PLoS Genet. 10, e1004383 (2014).

28. Wallace, C. Eliciting priors and relaxing the single causal variant assumption in colocalisation analyses. PLoS Genet. 16, e1008720 (2020).

29. Bryois, J. et al. Genetic identification of cell types underlying brain complex traits yields insights into the etiology of Parkinson’s disease. Nat. Genet. 52, 482–493 (2020).

30. Zeisel, A. et al. Molecular architecture of the mouse nervous system. Cell 174, 999–1014.e22 (2018).

31. Bulik-Sullivan, B. K. et al. LD Score regression distinguishes confounding from polygenicity in genome-wide association studies. Nat. Genet. 47, 291–295 (2015).

32. Benjamini, Y. & Hochberg, Y. Controlling the false discovery rate: a practical and powerful approach to multiple testing. J. R. Statist. Soc. B 57, 289–300 (1995).

33. Demontis, D. et al. Discovery of the first genome-wide significant risk loci for attention deficit/hyperactivity disorder. Nat. Genet. 51, 63–75 (2019).

34. Demontis, D. et al. Genome-wide analyses of ADHD identify 27 risk loci, refine the genetic architecture and implicate several cognitive domains. Nat. Genet. 55, 198–208 (2023).

35. Hall, L. S. et al. Genome-wide meta-analyses of stratified depression in Generation Scotland and UK Biobank. Transl. Psychiatry 8, 9 (2018).

36. Als, T. D. et al. Depression pathophysiology, risk prediction of recurrence and comorbid psychiatric disorders using genome-wide analyses. Nat. Med. 29, 1832–1844 (2023).

37. Cai, N. et al. Minimal phenotyping yields genome-wide association signals of low specificity for major depression. Nat. Genet. 52, 437–447 (2020).

38. Derks, E. M., Thorp, J. G. & Gerring, Z. F. Ten challenges for clinical translation in psychiatric genetics. Nat. Genet. 54, 1457–1465 (2022).

39. Pardiñas, A. F. et al. Common schizophrenia alleles are enriched in mutation-intolerant genes and in regions under strong background selection. Nat. Genet. 50, 381–389 (2018).

40. Baselmans, B. M. L. et al. Multivariate genome-wide analyses of the well-being spectrum. Nat. Genet. 51, 445– 451 (2019).

41. van den Heuvel, O. A. et al. (Brain circuitry of compulsivity. Eur. Neuropsychopharmacol. 26, 810–827 (2016).

42. Shephard, E. et al. Toward a neurocircuit-based taxonomy to guide treatment of obsessive-compulsive disorder. Mol. Psychiatry 26, 4583–4604 (2021).

43. Davis, M. A., Ireton, R. C. & Reynolds, A. B. A core function for p120-catenin in cadherin turnover. J. Cell Biol. 163, 525–534 (2003).

44. Yanagisawa, M. et al. A p120 catenin isoform switch affects Rho activity, induces tumor cell invasion, and predicts metastatic disease. J. Biol. Chem. 283, 18344–18354 (2008).

45. Daniel, J. M. & Reynolds, A. B. The catenin p120(ctn) interacts with Kaiso, a novel BTB/POZ domain zinc finger transcription factor. Mol. Cell. Biol. 19, 3614–3623 (1999).

46. Ishiyama, N. et al. Dynamic and static interactions between p120 catenin and E-cadherin regulate the stability of cell-cell adhesion. Cell 141, 117–128 (2010).

47. Schaffer, A. E. et al. CLP1 founder mutation links tRNA splicing and maturation to cerebellar development and neurodegeneration. Cell 157, 651–663 (2014).

48. Vandervore, L. V. et al. TMX2 is a crucial regulator of cellular redox state, and its dysfunction causes severe brain developmental abnormalities. Am. J. Hum. Genet. 105, 1126–1147 (2019).

49. Zhang, Y., Li, F., Fu, K., Liu, X.Lien, I.-C. & Li, H. Potential role of S-palmitoylation in cancer stem cells of lung adenocarcinoma. Front. Cell Dev. Biol. 9, 734897 (2021).

50. Hindley, G. et al. Charting the landscape of genetic overlap between mental disorders and related traits beyond genetic correlation. Am. J. Psychiatry 179, 833–843 (2022).

51. Mattheisen, M. et al. Genome-wide association study in obsessive-compulsive disorder: results from the OCGAS. Mol. Psychiatry 20, 337–344 (2015).

52. Watanabe, K. et al. A global overview of pleiotropy and genetic architecture in complex traits. Nat. Genet. 51, 1339–1348 (2019).

53. Nagel, M., Watanabe, K., Stringer, S., Posthuma, D. & van der Sluis, S. Item-level analyses reveal genetic heterogeneity in neuroticism. Nat. Commun. 9, 905 (2018).

54. Xie, X., Wang, Z. & Chen, Y. Association of LKB1 with a WD-repeat protein WDR6 is implicated in cell growth arrest and p27Kip1 induction. Mol. Cell. Biochem. 301, 115–122 (2007).

55. Adams, D. M., Reay, W. R. & Cairns, M. J. Multiomic prioritisation of risk genes for anorexia nervosa. Psychol. Med. 53, 6754–6762 (2023).

56. Kia, D. A. et al. Identification of candidate Parkinson disease genes by integrating genome-wide association study, expression, and epigenetic data sets. JAMA Neurol. 78, 464–472 (2021).

57. Lentini, J. M., Alsaif, H. S., Faqeih, E., Alkuraya, F. S. & Fu, D. DALRD3 encodes a protein mutated in epileptic encephalopathy that targets arginine tRNAs for 3-methylcytosine modification. Nat. Commun. 11, 2510 (2020).

58. Wu, J., Poppi, L. A. & Tischfield, M. A. Planar cell polarity and the pathogenesis of Tourette disorder: new hypotheses and perspectives. Dev. Biol. 489, 14–20 (2022).

59. Willsey, A. J. et al. De novo coding variants are strongly associated with Tourette disorder. Neuron 94, 486– 499.e9 (2017).

60. Zhao, X., Wang, S., Hao, J., Zhu, P., Zhang, X. & Wu, M. A whole-exome sequencing study of Tourette disorder in a Chinese population. DNA Cell Biol. 39, 63–68 (2020).

61. Debnath, M., Berk, M., Leboyer, M. & Tamouza, R. The MHC/HLA gene complex in major psychiatric disorders: emerging roles and implications. Curr. Behav. Neurosci. Rep. 5, 179–188 (2018).

62. Mataix-Cols, D. et al. A total-population multigenerational family clustering study of autoimmune diseases in obsessive–compulsive disorder and Tourette’s/chronic tic disorders. Mol. Psychiatry 23, 1652–1658 (2018).

63. Tylee, D. S. et al. Genetic correlations among psychiatric and immune-related phenotypes based on genome-wide association data. Am J Med Genet B Neuropsychiatr Genet 177, 641–657 (2018).

64. Westwell-Roper, C. et al. Immune-related comorbidities in childhood-onset obsessive compulsive disorder: lifetime prevalence in the Obsessive Compulsive Disorder Collaborative Genetics Association Study. J. Child Adolesc. Psychopharmacol. 29, 615–624 (2019).

65. Zhang, T. et al. Prenatal and early childhood infections and subsequent risk of obsessive-compulsive disorder and tic disorders: a nationwide, sibling-controlled study. Biol. Psychiatry 93, 1023–1030 (2023).

66. Swedo, S. E. et al. Pediatric autoimmune neuropsychiatric disorders associated with streptococcal infections: clinical description of the first 50 cases. Am. J. Psychiatry 155, 264–271 (1998).

67. Wilbur, C. et al. PANDAS/PANS in childhood: controversies and evidence. Paediatr. Child Health 24, 85–91 (2019).

68. Ahmari, S. E. et al. Repeated cortico-striatal stimulation generates persistent OCD-like behavior. Science 340, 1234–1239 (2013).

69. Ahmari, S. E. & Rauch S. L. The prefrontal cortex and OCD. Neuropsychopharmacol. 47, 211–224 (2022).

70. Olislagers, M., Rademaker, K., Adan, R. A. H., Lin, B. D. & Luykx, J. J. Comprehensive analyses of RNA-seq and genome-wide data point to enrichment of neuronal cell type subsets in neuropsychiatric disorders. Mol. Psychiatry 27, 947–955 (2022).

71. Boedhoe, P. S. W. et al. Cortical abnormalities associated with pediatric and adult obsessive-compulsive disorder: findings from the ENIGMA Obsessive-Compulsive Disorder Working Group. Am. J. Psychiatry 175, 453–462 (2018).

72. Bruin, W. B. et al. Structural neuroimaging biomarkers for obsessive-compulsive disorder in the ENIGMA-OCD consortium: medication matters. Transl. Psychiatry 10, 342 (2020).

73. van den Heuvel, O. A. et al. An overview of the first 5 years of the ENIGMA obsessive–compulsive disorder working group: the power of worldwide collaboration. Human Brain Mapping 43, 23–36 (2022).

74. Haber, S. N. Corticostriatal circuitry. Dialogues Clin. Neurosci. 18, 7–21 (2016).

75. Major Depressive Disorder Working Group of the Psychiatric Genomics Consortium. Trans-ancestry genome-wide study of depression identifies 697 associations implicating cell types and pharmacotherapies. Cell 188, 640–652.e9 (2025).

76. Trubetskoy, V. et al. Mapping genomic loci implicates genes and synaptic biology in schizophrenia. Nature 604, 502–508 (2022).

77. Savage, J. E. et al. Genome-wide association meta-analysis in 269,867 individuals identifies new genetic and functional links to intelligence. Nat. Genet. 50, 912–919 (2018).

78. Romero, C. et al. Exploring the genetic overlap between twelve psychiatric disorders. Nat. Genet. 54, 1795–1802 (2022).

79. Sharma, E. et al. Comorbidities in obsessive-compulsive disorder across the lifespan: a systematic review and meta-analysis. Front. Psychiatry 12, 703701 (2021).

80. Lee, P. H. et al. Genomic relationships, novel loci, and pleiotropic mechanisms across eight psychiatric disorders. Cell 179, 1469–1482.e11 (2019).

81. Grotzinger, A. D. Shared genetic architecture across psychiatric disorders. Psychol. Med. 51, 2210–2216 (2021).

82. Walters, R. K. et al. Transancestral GWAS of alcohol dependence reveals common genetic underpinnings with psychiatric disorders. Nat. Neurosci. 21, 1656–1669 (2018).

83. Virtanen, S. et al. Association of obsessive-compulsive disorder and obsessive-compulsive symptoms with substance misuse in 2 longitudinal cohorts in Sweden. JAMA Network Open 5, e2214779 (2022).

84. Sullivan, P. F. et al. Psychiatric genomics: an update and an agenda. Am. J. Psychiatry 175, 15–27 (2018).

85. Stewart, S. E. et al. Genome-wide association study of obsessive-compulsive disorder. Mol. Psychiatry 18, 788– 798 (2013).

86. Piantadosi SC, McClain LL, Klei L, Wang J, Chamberlain BL, Springer SA, et al. (2021): Transcriptome alterations are enriched for synapse-associated genes in the striatum of subjects with obsessive-compulsive disorder [no. 1]. Transl Psychiatry 11: 1–11.

87. Cappi, C. et al. De novo damaging DNA coding mutations are associated with obsessive-compulsive disorder and overlap with Tourette’s disorder and autism. Biol. Psychiatry 87, 1035–1044 (2020).

88. Halvorsen, M. et al. Exome sequencing in obsessive-compulsive disorder reveals a burden of rare damaging coding variants. Nat. Neurosci. 24, 1071–1076 (2021).

89. Knox, C. et al. DrugBank 6.0: the DrugBank Knowledgebase for 2024. Nucleic Acids Res. 52, D1265–D1275 (2024).

90. Wang, J. et al. CAUSALdb: a database for disease/trait causal variants identified using summary statistics of genome-wide association studies. Nucleic Acids Res. 48, D807–D816 (2020).

91. Buniello, A. et al. The NHGRI-EBI GWAS Catalog of published genome-wide association studies, targeted arrays and summary statistics 2019. Nucleic Acids Res. 47, D1005–D1012 (2019).

92. Elsworth, B. et al. The MRC IEU OpenGWAS data infrastructure. bioRxiv (10.1101/2020.08.10.244293).

## Methods-only references

93. DSM-5. Diagnostic and statistical manual of mental disorders: DSM-5. 237-242. ISBN 978-0-89042-555–8 (2013).

94. World Health Organization. ICD-11: International classification of diseases (11th revision), https://icd.who.int/ (2019).

95. McCarthy, S. et al. A reference panel of 64,976 haplotypes for genotype imputation. Nat. Genet. 48, 1279–1283 (2016).

96. The 1000 Genomes Project Consortium. A global reference for human genetic variation. Nature 526, 68–74 (2015).

97. Lam, M. et al. RICOPILI: Rapid Imputation for COnsortias PIpeLIne. Bioinformatics 36, 930–933 (2020).

98. Willer, C. J., Li, Y. & Abecasis, G. R. METAL: fast and efficient meta-analysis of genomewide association scans. Bioinformatics 26, 2190–2191 (2010).

99. Higgins, J. P. T. & Thompson, S. G. Quantifying heterogeneity in a meta-analysis. Stat.in Med. 21, 1539–1558 (2002).

100. Higgins, J. P. T., Thompson, S. G., Deeks, J. J. & Altman, D. G. Measuring inconsistency in meta-analyses. BMJ 327, 557–560 (2003).

101. Turley, P. et al. Multi-trait analysis of genome-wide association summary statistics using MTAG. Nat. Genet. 50, 229–237 (2018).

102. Altshuler, D. M. et al. Integrating common and rare genetic variation in diverse human populations. Nature 467, 52–58 (2010).

103. Grotzinger, A. D., de la Fuente, J., Privé, F., Nivard, M. G. & Tucker-Drob, E. M. Pervasive downward bias in estimates of liability-scale heritability in genome-wide association study meta-analysis: a simple solution. Biol. Psychiatry 93, 29–36 (2023).

104. Wang, D. et al. Comprehensive functional genomic resource and integrative model for the human brain. Science 362, eaat8464 (2018).

105. de Klein, N. et al. Brain expression quantitative trait locus and network analyses reveal downstream effects and putative drivers for brain-related diseases. Nat. Genet. 55, 377–388 (2023).

106. de Leeuw, C. A., Mooij, J. M., Heskes, T. & Posthuma, D. MAGMA: generalized gene-set analysis of GWAS data. PLoS Comput. Biol. 11, e1004219 (2015).

107. Finucane, H. K. et al. Partitioning heritability by functional annotation using genome-wide association summary statistics. Nat. Genet. 47, 1228–1235 (2015).

108. Benner, C., Spencer, C. C. A., Havulinna, A. S., Salomaa, V., Ripatti, S. & Pirinen, M. FINEMAP: efficient variable selection using summary data from genome-wide association studies. Bioinformatics 32, 1493– 1501 (2016).

109. Chen, W. et al. Fine mapping causal variants with an approximate Bayesian method using marginal test statistics. Genetics 200, 719–736 (2015).

110. Kichaev, G. et al. Integrating functional data to prioritize causal variants in statistical fine-mapping studies. PLoS Genet. 10, e1004722 (2014).

111. Kichaev, G. & Pasaniuc, B. Leveraging functional-annotation data in trans-ethnic fine-mapping studies. Am J Hum Genet 97, 260–271 (2015).

112. Lek, M. et al. Analysis of protein-coding genetic variation in 60,706 humans. Nature 536, 285–291 (2016).

