## Supplementary Notes and Figures for "Genome-wide analyses identify 30 loci associated with obsessive-compulsive disorder"

### Supplementary Notes

#### Supplementary Note 1: Author Byline – Consortias

##### *23andMe*

Stella Aslibekyan, Adam Auton, Elizabeth Babalola, Robert K. Bell, Jessica Bielenberg, Jonathan Bowes, Katarzyna Bryc, Ninad S. Chaudhary, Daniella Coker, Sayantan Das, Emily DelloRusso, Sarah L. Elson, Nicholas Eriksson, Teresa Filshtein, Pierre Fontanillas, Will Freyman, Zach Fuller, Chris German, Julie M. Granka, Karl Heilbron, Alejandro Hernandez, Barry Hicks, David A. Hinds, Ethan M. Jewett, Yunxuan Jiang, Katelyn Kukar, Alan Kwong, Yanyu Liang, Keng-Han Lin, Bianca A. Llamas, Matthew H. McIntyre, Steven J. Micheletti, Meghan E. Moreno, Priyanka Nandakumar, Dominique T. Nguyen, Jared O'Connell, Aaron A. Petrakovitz, G. David Poznik, Alexandra Reynoso, Shubham Saini, Morgan Schumacher, Leah Selcer, Anjali J. Shastri, Janie F. Shelton, Jingchunzi Shi, Suyash Shringarpure, Qiaojuan Jane Su, Susana A. Tat, Vinh Tran, Joyce Y. Tung, Xin Wang, Wei Wang, Catherine H. Weldon, Peter Wilton, Corinna D. Wong.

##### *COGA authors*

The Collaborative Study on the Genetics of Alcoholism (COGA), Principal Investigators B. Porjesz, V. Hesselbrock, T. Foroud; Scientific Director, A. Agrawal; Translational Director, D. Dick, includes ten different centers: University of Connecticut (V. Hesselbrock); Indiana University (H.J. Edenberg, T. Foroud, Y. Liu, M.H. Plawecki); University of Iowa Carver College of Medicine (S. Kuperman, J. Kramer); SUNY Downstate Health Sciences University (B. Porjesz, J. Meyers, C. Kamarajan, A. Pandey); Washington University in St. Louis (L. Bierut, J. Rice, K. Bucholz, A. Agrawal); University of California at San Diego (M. Schuckit); Rutgers University (J. Tischfield, D. Dick, R. Hart, J. Salvatore); The Children's Hospital of Philadelphia, University of Pennsylvania (L. Almasy); Icahn School of Medicine at Mount Sinai (A. Goate, P. Slesinger); and Howard University (D. Scott). Other COGA collaborators include: L. Bauer (University of Connecticut); J. Nurnberger Jr., L. Wetherill, X., Xuei, D. Lai, S. O'Connor, (Indiana University); G. Chan (University of Iowa; University of Connecticut); D.B. Chorlian, J. Zhang, P. Barr, S. Kinreich, G. Pandey (SUNY Downstate); N. Mullins (Icahn School of Medicine at Mount Sinai); A. Anokhin, S. Hartz, E. Johnson, V. McCutcheon, S. Saccone (Washington University); J. Moore, F. Aliev, Z. Pang, S. Kuo (Rutgers University); A. Merikangas (The Children's Hospital of Philadelphia and University of Pennsylvania); H. Chin and A. Parsian are the NIAAA Staff Collaborators. We continue to be inspired by our memories of Henri Begleiter and Theodore Reich, founding PI and Co-PI of COGA, and also owe a debt of gratitude to other past organizers of COGA, including Ting- Kai Li, P. Michael Conneally, Raymond Crowe, and Wendy Reich, for their critical contributions. This national collaborative study is supported by NIH Grant U10AA008401 from the National Institute on Alcohol Abuse and Alcoholism (NIAAA) and the National Institute on Drug Abuse (NIDA).

#### *EstBB authors*

Andres Metspalu[1], Tõnu Esko[1], Reedik Mägi[1], Mari Nelis[1], Georgi Hudjashov[1]

1: Estonian Genome Centre, Institute of Genomics, University of Tartu

#### *Hunt authors*

Laurent F. Thomas[1,2,3,4], Anne H. Skogholt[1], Bendik Winsvold[1,5,6], Maiken Elvestad Gabrielsen[1], Sigrid Børte[1,7,8], Ole A Andreassen MD, PhD[9,10], Ottar Bjerkeset MD, PhD[11,12], Ole Kristian Drange MD, PhD[11,13], Grete Dyb MD, PhD[14], Katrine Kveli Fjukstad MD, PhD[15,16], Lars G Fritsche PhD[17], Ingrid Heuch MD, PhD[5], Marit Sæbø Indredavik MD, PhD[2], Håvard Kallestad MD[11,13], Amy E Martinsen MSc[1,2,5], Gunnar Morken MD, PhD[11,13], Jonas Bille Nielsen PhD[1,18], Torunn Stene Nøvik MD, PhD[19], Linda M Pedersen PhD[5], Sara Germans Selvik MD, PhD[20], Børge Sivertsen PhD[11,21,22], Marit Skrove MD, PhD[23], Synne Øien Stensland MD, PhD[7,14,], Eystein Stordal MD, PhD[11,20], Cristen J Willer PhD[18], Wei Zhou PhD[24,25], Kristian Hveem[1,26,27], John-Anker Zwart[1,5,8]

1: K. G. Jebsen Center for Genetic Epidemiology, Department of Public Health and Nursing, Faculty of Medicine and Health Sciences, Norwegian University of Science and Technology (NTNU), Trondheim, Norway.
2: Department of Clinical and Molecular Medicine, Faculty of Medicine and Health Sciences, Norwegian University of Science and Technology (NTNU), Trondheim, Norway.
3: BioCore - Bioinformatics Core Facility, Norwegian University of Science and Technology, Trondheim. Norway.
4: Clinic of Laboratory Medicine, St.Olavs Hospital, Trondheim University Hospital, Trondheim, Norway.
5: Department of Research and Innovation and Education, Division of Clinical Neuroscience, Oslo University Hospital, Oslo, Norway.
6: Department of Neurology, Oslo University Hospital, Oslo, Norway.
7: Research and Communication Unit for Musculoskeletal Health (FORMI), Department of Research and Innovation, Division of Clinical Neuroscience, Oslo University Hospital, Oslo, Norway.
8: Institute of Clinical Medicine, Faculty of Medicine, University of Oslo, Oslo, Norway.
9: Division of Mental Health and Addiction, Oslo University Hospital, Oslo, Norway.
10: NORMENT, University of Oslo, Oslo, Norway.
11: Department of Mental Health, Faculty of Medicine and Health Sciences, Norwegian University of Science and Technology (NTNU), Trondheim, Norway.
12: Faculty of Nursing and Health Sciences, NORD University, Levanger, Norway.
13: Division of Mental Health Care, St. Olavs Hospital, Trondheim University Hospital, Trondheim, Norway.
14: Norwegian Centre for Violence and Traumatic Stress Studies, Oslo, Norway.
15: Department of Psychiatry, Nord-Trøndelag Hospital Trust, Levanger Hospital, Levanger, Norway.
16: Department of Laboratory Medicine, Children's and Women's Health, Norwegian University of Science and Technology (NTNU), Trondheim, Norway.
17: Center for Statistical Genetics, Department of Biostatistics, University of Michigan, Ann Arbor, MI, 48109, USA.

18: Department of Internal Medicine, Division of Cardiovascular Medicine, University of Michigan, Ann Arbor, MI, 48109, USA.
19: Department of Child and Adolescent Psychiatry, St. Olavs Hospital, Trondheim University Hospital, Trondheim, Norway.
20: Department of Psychiatry, Hospital Namsos, Nord-Trøndelag Health Trust, Namsos, Norway.
21: Department of Health Promotion, Norwegian Institute of Public Health, Bergen, Norway.
22: Department of Research and Innovation, Helse-Fonna HF, Haugesund, Norway.
23: Regional Centre for Child and Youth Mental Health and Child Welfare, Department of Mental Health, Faculty of Medicine and Health Sciences, Norwegian University of Science and Technology (NTNU), Trondheim, Norway.
24: Department of Computational Medicine and Bioinformatics, University of Michigan, Ann Arbor, MI, 48109, USA.
25: Analytic and Translational Genetics Unit, Massachusetts General Hospital, Boston, MA, USA.
26: HUNT Research Center, Department of Public Health and Nursing, Faculty of Medicine and Health Sciences, Norwegian University of Science and Technology, Trondheim, Norway.
27: Department of Research, Innovation and Education, St. Olavs Hospital, Trondheim University Hospital, Trondheim, Norway.

#### *VA Million Veteran Program*

MVP Program Office

- Sumitra Muralidhar, Ph.D., Program Director
  US Department of Veterans Affairs, 810 Vermont Avenue NW, Washington, DC 20420

- Jennifer Moser, Ph.D., Associate Director, Scientific Programs
  US Department of Veterans Affairs, 810 Vermont Avenue NW, Washington, DC 20420

- Jennifer E. Deen, B.S., Associate Director, Cohort & Public Relations
  US Department of Veterans Affairs, 810 Vermont Avenue NW, Washington, DC 20420

MVP Executive Committee

- Co-Chair: Philip S. Tsao, Ph.D.
  VA Palo Alto Health Care System, 3801 Miranda Avenue, Palo Alto, CA 94304

- Co-Chair: Sumitra Muralidhar, Ph.D.
  US Department of Veterans Affairs, 810 Vermont Avenue NW, Washington, DC 20420

- J. Michael Gaziano, M.D., M.P.H.
  VA Boston Healthcare System, 150 S. Huntington Avenue, Boston, MA 02130

- Elizabeth Hauser, Ph.D.
  Durham VA Medical Center, 508 Fulton Street, Durham, NC 27705

- Amy Kilbourne, Ph.D., M.P.H.
VA HSR&D, 2215 Fuller Road, Ann Arbor, MI 48105

- Shiuh-Wen Luoh, M.D., Ph.D.
VA Portland Health Care System, 3710 SW US Veterans Hospital Rd, Portland, OR 97239

- Michael Matheny, M.D., M.S., M.P.H.
VA Tennessee Valley Healthcare System, 1310 24th Ave. South, Nashville, TN 37212

- Dave Oslin, M.D.
Philadelphia VA Medical Center, 3900 Woodland Avenue, Philadelphia, PA 19104

MVP Co-Principal Investigators

- J. Michael Gaziano, M.D., M.P.H.
VA Boston Healthcare System, 150 S. Huntington Avenue, Boston, MA 02130

- Philip S. Tsao, Ph.D.
VA Palo Alto Health Care System, 3801 Miranda Avenue, Palo Alto, CA 94304

MVP Core Operations

- Lori Churby, B.S., Director, MVP Regulatory Affairs
VA Palo Alto Health Care System, 3801 Miranda Avenue, Palo Alto, CA 94304

- Stacey B. Whitbourne, Ph.D., Director, MVP Cohort Management
VA Boston Healthcare System, 150 S. Huntington Avenue, Boston, MA 02130

- Jessica V. Brewer, M.P.H., Director, MVP Recruitment & Enrollment
VA Boston Healthcare System, 150 S. Huntington Avenue, Boston, MA 02130

- Shahpoor (Alex) Shayan, M.S., Director, MVP Recruitment and Enrollment Informatics
VA Boston Healthcare System, 150 S. Huntington Avenue, Boston, MA 02130

- Luis E. Selva, Ph.D., Executive Director, MVP Biorepositories
VA Boston Healthcare System, 150 S. Huntington Avenue, Boston, MA 02130

- Saiju Pyarajan Ph.D., Director, Data and Computational Sciences
VA Boston Healthcare System, 150 S. Huntington Avenue, Boston, MA 02130

- Kelly Cho, M.P.H, Ph.D., Director, MVP Phenomics Data Core
VA Boston Healthcare System, 150 S. Huntington Avenue, Boston, MA 02130

- Scott L. DuVall, Ph.D., Director, VA Informatics and Computing Infrastructure (VINCI)
VA Salt Lake City Health Care System, 500 Foothill Drive, Salt Lake City, UT 84148

- Mary T. Brophy M.D., M.P.H., Director, VA Central Biorepository
VA Boston Healthcare System, 150 S. Huntington Avenue, Boston, MA 02130

MVP Coordinating Centers

- MVP Coordinating Center, Boston - J. Michael Gaziano, M.D., M.P.H.
  VA Boston Healthcare System, 150 S. Huntington Avenue, Boston, MA 02130

- MVP Coordinating Center, Palo Alto – Philip S. Tsao, Ph.D.
  VA Palo Alto Health Care System, 3801 Miranda Avenue, Palo Alto, CA 94304

- MVP Information Center, Canandaigua – Brady Stephens, M.S.
  Canandaigua VA Medical Center, 400 Fort Hill Avenue, Canandaigua, NY 14424

- Cooperative Studies Program Clinical Research Pharmacy Coordinating Center, Albuquerque – Todd Connor, Pharm.D.; Dean P. Argyres, B.S., M.S.

New Mexico VA Health Care System, 1501 San Pedro Drive SE, Albuquerque, NM 87108

MVP Publications and Presentations Committee

- Co-Chair: Themistocles L. Assimes, M.D., Ph. D
  VA Palo Alto Health Care System, 3801 Miranda Avenue, Palo Alto, CA 94304
- Co-Chair: Adriana Hung, M.D.; M.P.H
  VA Tennessee Valley Healthcare System, 1310 24th Ave. South, Nashville, TN 37212
- Co-Chair: Henry Kranzler, M.D.
  Philadelphia VA Medical Center, 3900 Woodland Avenue, Philadelphia, PA 19104

MVP Local Site Investigators

- Samuel Aguayo, M.D., Phoenix VA Health Care System
  650 E. Indian School Road, Phoenix, AZ 85012
- Sunil Ahuja, M.D., South Texas Veterans Health Care System
  7400 Merton Minter Boulevard, San Antonio, TX 78229
- Kathrina Alexander, M.D., Veterans Health Care System of the Ozarks
  1100 North College Avenue, Fayetteville, AR 72703
- Xiao M. Androulakis, M.D., Columbia VA Health Care System
  6439 Garners Ferry Road, Columbia, SC 29209
- Prakash Balasubramanian, M.D., William S. Middleton Memorial Veterans Hospital
  2500 Overlook Terrace, Madison, WI 53705
- Zuhair Ballas, M.D., Iowa City VA Health Care System
  601 Highway 6 West, Iowa City, IA 52246-2208
- Jean Beckham, Ph.D., Durham VA Medical Center
  508 Fulton Street, Durham, NC 27705
- Sujata Bhushan, M.D., VA North Texas Health Care System
  4500 S. Lancaster Road, Dallas, TX 75216

- Edward Boyko, M.D., VA Puget Sound Health Care System
  1660 S. Columbian Way, Seattle, WA 98108-1597

- David Cohen, M.D., Portland VA Medical Center
  3710 SW U.S. Veterans Hospital Road, Portland, OR 97239

- Louis Dellitalia, M.D., Birmingham VA Medical Center
  700 S. 19th Street, Birmingham AL 35233

- L. Christine Faulk, M.D., Robert J. Dole VA Medical Center
  5500 East Kellogg Drive, Wichita, KS 67218-1607

- Joseph Fayad, M.D., VA Southern Nevada Healthcare System
  6900 North Pecos Road, North Las Vegas, NV 89086

- Daryl Fujii, Ph.D., VA Pacific Islands Health Care System
  459 Patterson Rd, Honolulu, HI 96819

- Saib Gappy, M.D., John D. Dingell VA Medical Center
  4646 John R Street, Detroit, MI 48201

- Frank Gesek, Ph.D., White River Junction VA Medical Center
  163 Veterans Drive, White River Junction, VT 05009

- Jennifer Greco, M.D., Sioux Falls VA Health Care System
  2501 W 22nd Street, Sioux Falls, SD 57105

- Michael Godschalk, M.D., Richmond VA Medical Center
  1201 Broad Rock Blvd., Richmond, VA 23249

- Todd W. Gress, M.D., Ph.D., Hershel “Woody” Williams VA Medical Center
  1540 Spring Valley Drive, Huntington, WV 25704

- Samir Gupta, M.D., M.S.C.S., VA San Diego Healthcare System
  3350 La Jolla Village Drive, San Diego, CA 92161

- Salvador Gutierrez, M.D., Edward Hines, Jr. VA Medical Center
  5000 South 5th Avenue, Hines, IL 60141

- John Harley, M.D., Ph.D., Cincinnati VA Medical Center
  3200 Vine Street, Cincinnati, OH 45220

- Kimberly Hammer, Ph.D., Fargo VA Health Care System
  2101 N. Elm, Fargo, ND 58102

- Mark Hamner, M.D., Ralph H. Johnson VA Medical Center
  109 Bee Street, Mental Health Research, Charleston, SC 29401

- Adriana Hung, M.D., M.P.H., VA Tennessee Valley Healthcare System
  1310 24th Avenue, South Nashville, TN 37212

- Robin Hurley, M.D., W.G. (Bill) Hefner VA Medical Center
  1601 Brenner Ave, Salisbury, NC 28144

- Pran Iruvanti, D.O., Ph.D., Hampton VA Medical Center
  100 Emancipation Drive, Hampton, VA 23667

- Frank Jacono, M.D., VA Northeast Ohio Healthcare System
  10701 East Boulevard, Cleveland, OH 44106

- Darshana Jhala, M.D., Philadelphia VA Medical Center
  3900 Woodland Avenue, Philadelphia, PA 19104

- Scott Kinlay, M.B.B.S., Ph.D., VA Boston Healthcare System
  150 S. Huntington Avenue, Boston, MA 02130

- Jon Klein, M.D., Ph.D., Louisville VA Medical Center
  800 Zorn Avenue, Louisville, KY 40206

- Michael Landry, Ph.D., Southeast Louisiana Veterans Health Care System
  2400 Canal Street, New Orleans, LA 70119

- Peter Liang, M.D., M.P.H., VA New York Harbor Healthcare System
  423 East 23rd Street, New York, NY 10010

- Suthat Liangpunsakul, M.D., M.P.H., Richard Roudebush VA Medical Center
  1481 West 10th Street, Indianapolis, IN 46202

- Jack Lichy, M.D., Ph.D., Washington DC VA Medical Center
  50 Irving St, Washington, D. C. 20422

- C. Scott Mahan, M.D., Charles George VA Medical Center
  1100 Tunnel Road, Asheville, NC 28805

- Ronnie Marrache, M.D., VA Maine Healthcare System
  1 VA Center, Augusta, ME 04330

- Stephen Mastorides, M.D., James A. Haley Veterans' Hospital
  13000 Bruce B. Downs Blvd, Tampa, FL 33612

- Elisabeth Mates M.D., Ph.D., VA Sierra Nevada Health Care System
  975 Kirman Avenue, Reno, NV 89502

- Kristin Mattocks, Ph.D., M.P.H., Central Western Massachusetts Healthcare System
  421 North Main Street, Leeds, MA 01053

- Paul Meyer, M.D., Ph.D., Southern Arizona VA Health Care System
  3601 S 6th Avenue, Tucson, AZ 85723

- Jonathan Moorman, M.D., Ph.D., James H. Quillen VA Medical Center
  Corner of Lamont & Veterans Way, Mountain Home, TN 37684

- Timothy Morgan, M.D., VA Long Beach Healthcare System
  5901 East 7th Street Long Beach, CA 90822

- Maureen Murdoch, M.D., M.P.H., Minneapolis VA Health Care System
  One Veterans Drive, Minneapolis, MN 55417

- James Norton, Ph.D., VA Health Care Upstate New York
  113 Holland Avenue, Albany, NY 12208

- Olaoluwa Okusaga, M.D., Michael E. DeBakey VA Medical Center
  2002 Holcombe Blvd, Houston, TX 77030

- Kris Ann Oursler, M.D., Salem VA Medical Center
  1970 Roanoke Blvd, Salem, VA 24153

- Ana Palacio, M.D., M.P.H., Miami VA Health Care System
  1201 NW 16th Street, 11 GRC, Miami FL 33125

- Samuel Poon, M.D., Manchester VA Medical Center
  718 Smyth Road, Manchester, NH 03104

- Emily Potter, Pharm.D., VA Eastern Kansas Health Care System
  4101 S 4th Street Trafficway, Leavenworth, KS 66048

- Michael Rauchman, M.D., St. Louis VA Health Care System
  915 North Grand Blvd, St. Louis, MO 63106

- Richard Servatius, Ph.D., Syracuse VA Medical Center
  800 Irving Avenue, Syracuse, NY 13210

- Satish Sharma, M.D., Providence VA Medical Center
  830 Chalkstone Avenue, Providence, RI 02908

- River Smith, Ph.D., Eastern Oklahoma VA Health Care System
  1011 Honor Heights Drive, Muskogee, OK 74401

- Peruvemba Sriram, M.D., N. FL/S. GA Veterans Health System
  1601 SW Archer Road, Gainesville, FL 32608

- Patrick Strollo, Jr., M.D., VA Pittsburgh Health Care System
  University Drive, Pittsburgh, PA 15240

- Neeraj Tandon, M.D., Overton Brooks VA Medical Center
  510 East Stoner Ave, Shreveport, LA 71101

- Philip Tsao, Ph.D., VA Palo Alto Health Care System
  3801 Miranda Avenue, Palo Alto, CA 94304-1290

- Gerardo Villareal, M.D., New Mexico VA Health Care System
  1501 San Pedro Drive, S.E. Albuquerque, NM 87108

- Agnes Wallbom, M.D., M.S., VA Greater Los Angeles Health Care System
  11301 Wilshire Blvd, Los Angeles, CA 90073

- Jessica Walsh, M.D., VA Salt Lake City Health Care System
  500 Foothill Drive, Salt Lake City, UT 84148

- John Wells, Ph.D., Edith Nourse Rogers Memorial Veterans Hospital
  200 Springs Road, Bedford, MA 01730

- Jeffrey Whittle, M.D., M.P.H., Clement J. Zablocki VA Medical Center
  5000 West National Avenue, Milwaukee, WI 53295

- Mary Whooley, M.D., San Francisco VA Health Care System
  4150 Clement Street, San Francisco, CA 94121

- Allison E. Williams, N.D., Ph.D., R.N, Bay Pines VA Healthcare System
  10,000 Bay Pines Blvd Bay Pines, FL 33744

- Peter Wilson, M.D., Atlanta VA Medical Center
  1670 Clairmont Road, Decatur, GA 30033

- Junzhe Xu, M.D., VA Western New York Healthcare System
  3495 Bailey Avenue, Buffalo, NY 14215-1199

- Shing Shing Yeh, Ph.D., M.D., Northport VA Medical Center
  79 Middleville Road, Northport, NY 11768

### Supplementary Note 2: Ethics and sample descriptions

GWAS results based on some of the included cases and controls have been published previously by the International OCD Foundation (IOCDF-GC; Stewart et al., 2013, IOCDF & OCGAS et al., 2018) and the OCD Collaborative Genetics Association Study (OCGAS; Mattheisen et al., 2015, IOCDF & OCGAS et al., 2018). These data were re-analyzed for the current publication using newly matched control participants that were genotyped with the same microarrays as the cases, making up 2,828 cases and 4,887 controls. GWAS results based on a subset of the cohorts are currently available as preprints ($N_{cases} = 14{,}140$, $N_{controls} = 562{,}117$, Strom et al., 2021 and $N_{cases} = 37{,}015$, $N_{controls} = 948{,}616$, Strom et al., 2024). Of those cohorts, three (EstBB, FinnGen, iPSYCH) were updated to include additional OCD cases and controls compared to the samples in the preprints. Seven cohorts are new to this study and were not included in any of the previously published GWAS ($N_{cases} = 6{,}120$, $N_{controls} = 430{,}999$). Here we describe each individual sample that was included in the OCD meta-analysis, in alphabetical order. The header of each sample lists study identifier, study principal investigator(s) (PI(s)), country or site name, and if there has been a previous publication in connection with the data, also PubMed ID(s) or doi. See **Supplementary Table 1** for an overview of sample sizes, number of included SNPs, reference panel, GWAS analysis tool, and Lambda1000 estimate for each individual cohort.

##### *Ethics approvals*

All relevant ethics approvals have been obtained by the respective cohort's. 23andMe: Participants provided informed consent and volunteered to participate in the research online, under a protocol approved by the external AAHRPP-accredited IRB, Ethical & Independent (E&I) Review Services. As of 2022, E&I Review Services is part of Salus IRB (https://www.versiticlinicaltrials.org/salusirb). AGDS: All study protocols were approved by the QIMR Berghofer Medical Research Institute Human Research Ethics Committee. The protocol for approaching participants through the DHS, enrolling them in the study, and consenting for all phases of the study (including invitation to future related studies) and accessing MBS and PBS records was approved by the Ethics Department of the Department of Human Services. BioVU: The Vanderbilt University Medical Center Institutional Review Board oversees BioVU and approved this project (IRB201609). COGA: Institutional review boards at all sites approved the study and all participants provided informed consent. EGOS: Ethical approvals were obtained from the Institutional Review Board (IRB) at the Icahn School of Medicine at Mount Sinai, New York, NY, and the Regional Ethical Review Board in Stockholm. EPOC: The study was in accordance with the revised Declaration of Helsinki and approved by the local ethics committees of the Charite University Medicine Berlin and the University Hospital Bonn. EstBB: At recruitment, participants signed a consent allowing follow-up linkage of their electronic health records (EHRs), thereby providing a longitudinal collection of their phenotypic information. FinnGen: The Ethical Review Board of the Hospital District of Helsinki and Uusimaa approved the FinnGen study protocol Nr. HUS/990/2017. The FinnGen project was approved by Finnish Institute for Health and Welfare (THL), approval numbers THL/2031/6.02.00/2017, amendments THL/341/6.02.00/2018, THL/2222/6.02.00/2018, and THL/283/6.02.00/2019. HUNT: The HUNT study was approved by the Regional Committee for Medical and Health Research Ethics, Norway (2015/575). IOCDF/OCGAS: This work was approved by the relevant IRBs at all participating sites, and all participants provided written informed consent. iPSYCH: The study was approved by the Regional Scientific Ethics Committee in Denmark and the Danish Data Protection Agency. MVP: The U.S. Department of Veterans Affairs (VA) Million Veteran Program (MVP) is collecting genetic and electronic health record (EHR) data in the U.S with ethical approval given by the Central VA Institutional Review Board (IRB) and site-specific IRBs. All relevant ethical regulations for work with human subjects were followed in the conduct of the study, and informed consent was obtained from all participants. MoBa: The establishment of MoBa and initial data collection was based on a license from the Norwegian Data Protection Agency and approval from The Regional Committees for Medical and Health Research Ethics. The MoBa cohort is now based on regulations related to the Norwegian Health Registry Act. NORDiC-SWE: This study was approved by the Regional Ethics Committee, Stockholm (EPN Stockholm) and the Institutional Review Board (IRB) at the University of North Carolina at Chapel Hill and all subjects provided informed consent. NORDiC-NOR: The NORDiC-NOR study was approved by the Norwegian Regional Committee for Medical and Health Research Ethics (IRB00001872 REK West) under project number 2018/52 REKVest (PI: Bjarne Hansen) and project number: 2014/75 REKVest (PI: Jan Haavik) and all subjects provided informed consent. OCGAS-all: Ethics approvals for the OCGAS study were obtained from the Hopkins Medicine Institutional Review Boards, the Butler Institutional Review Board, the UCLA Institutional Review Boards, the Mass General Brigham Human Research Committee, the Columbia University Institutional Review Boards, and the National Institutes of Health Institutional Review Board (NIH IRB). OCGAS-nestadt: Ethics approvals for the OCGAS study were obtained from the Hopkins Medicine Institutional Review Boards. OCGAS-ab: The study was approved by REB at Hospital for Sick Children. Ethics approvals for the OCGAS study were obtained from the Hopkins Medicine Institutional Review Boards. OCGAS-gh: Informed written consent was obtained in all cases by the participants or their parents. The study was approved by the ethical commissions of all involved universities in accordance with the latest version of the Declaration of Helsinki, including an ethical permission granted by the Ethic Committees from Aachen, Wuerzburg, Marburg, Freiburg, and the Cantonal Ethic Commission of Zuerich (Ref. Nr. 39/97, 140/3 and EK: KEK-ZHNr. 2010-0340/3). OCD-

WWF: The study was approved by the ethics committee of the University of Wuerzburg, Germany and was conducted according to the ethical principles of the Helsinki Declaration. All patients gave written informed consent prior to participation. Psych_Broad: Both studies have been approved by the Clinical Research Ethics Committee (CREC) of Hospital Universitari Vall d'Hebron. All methods were performed in accordance with the relevant guidelines and regulations and written informed consent was obtained from participant parents before inclusion into the study. UKBB: Research on the UK Biobank is conducted under a generic Research Tissue Bank approval from the UK North West Multi-centre Research Ethics Committee (MREC). This research was approved to be conducted under that approval by the governing Research Ethics Committee of the UK Biobank. The analyses in this paper were performed under an approved extension to project 16577. Yale-Penn: Participants were recruited from eastern U.S sites and provided written informed consent as approved by the institutional review board at each site.

We confirm that all necessary patient/participant consent has been obtained and the appropriate institutional forms have been archived, and that any patient/participant/sample identifiers included were not known to anyone (e.g., hospital staff, patients or participants themselves) outside the research group so cannot be used to identify individuals.

#### *23andMe | 23andMe | USA | https://doi.org/10.1101/2024.03.06.24303776*

Samples of European ancestry were drawn from the customer base of 23andMe Inc., a private consumer genetics company. The 23andMe cohort has been described in detail elsewhere (Hyde et al., 2016). Participants provided informed consent and volunteered to participate in the research online, under a protocol approved by the external AAHRPP-accredited IRB, Ethical & Independent (E&I) Review Services. As of 2022, E&I Review Services is part of Salus IRB (https://www.versiticlinicaltrials.org/salusirb). The cohort was selected from participant data available on 1st September 2017 and the analysis was reviewed and approved by a private institutional review board (www.eandireview.com). All individuals identified as cases have reported being diagnosed with OCD, while controls have reported not having been diagnosed with OCD. 30,167 OCD cases and 929,804 controls were included in the final GWAS. Of the cases, 10,808 (35.83%) were male and 19,359 (64.17%) were female. Of the controls, 443,336 (47.68%) were male and 486,468 (52.32%) were female. Of the cases, 7,040 (23.34%) individuals were below the age of 30, 11,087 (36.75%) between 30 and 45, 7,056 (23.39%) between 45 and 60, and 4,984 (16.52%) above the age of 60. Of the controls, 91,287 (9.82%) individuals were below the age of 30, 230,696 (24.81%) between 30 and 45, 257,793 (37.64%) between 45 and 60, and 350,028 (27.73%) above the age of 60. Extraction of DNA and genotyping was performed by the National Genetics Institute (NGI), a CLIA licensed clinical laboratory and a subsidiary of Laboratory Corporation of America. Individuals were genotyped on four different genotype platforms. Two (V1, V2) platforms were variants of the Illumina HumanHap550+ BeadChip, including 25,000 custom SNPs, one platform (V3) was the Illumina OmniExpress+ BeadChip, with custom SNPs to increase overlap with V2, and one platform (V4) is in current use and a fully customized array. Individuals that failed to meet a 98.5% call rate were re-analyzed. Only individuals with > 97% European ancestry were included in the analysis. European ancestry was determined through an analysis of local ancestry using a support vector machine to classify individual haplotypes into one of 31 reference populations. Those classifications were then fed into a hidden Markov model (HMM) which accounts for incorrect assignments and switch errors, thereby giving probabilities for each reference population per window (100 SNPs). Simulated admixed individuals were then used to re-calibrate the HMM probabilities so the assigned ancestries were consistent with the simulated individuals. Publicly available datasets (Human Genome Diversity Project, HapMap (Altshuler et al., 2010), and 1000 Genomes (The 1000 Genomes Project Consortium, 2015)) and 23andMe customers with four grandparents from the same country served as the reference population. Identity-by-descent (IBD) estimation was used to define a maximal set of unrelated individuals for each analysis. Individuals who shared less than 700 cM IBD were defined as unrelated (e.g., approximately less related than first cousins).

The merged UK10K and 1000 Genomes Phase 3 panel was used for imputation. Finch, an internally developed tool that implements the Beagle haplotype graph-based phasing algorithm, modified to separate the steps of graph construction and phasing, was used for phasing. Imputation was then performed as an estimated allele dosage averaged over a set of possible imputed haplotypes for each individual. Genetic association testing was performed using logistic regression, assuming an additive model for allelic effects, including age, sex, first five principal components (PCs), and the genotype platform as covariates. For imputed data, the imputed dosages rather than the best-guess genotypes were used. The association p-value was computed using a likelihood ratio test.

##### *AGDS | Martin, N. | QIMR, Brisbane, Australia*

The Australian Genetics of Depression study (AGDS) was established to recruit a large cohort of individuals who have been diagnosed with depression at some point in their lifetime. The purpose of establishing this cohort is to investigate genetic and environmental risk factors for depression and response to commonly prescribed antidepressants. All cases and controls were diagnosed with Major Depressive Disorder. OCD cases were identified based on self-reported clinical diagnosis for OCD while OCD controls had no self-reported clinical diagnosis for OCD and obtained a score below 10 on the Obsessive-Compulsive Inventory-Revised (OCI-R). A total of 20,689 participants were recruited through the Australian Department of Human Services and a media campaign, 75% of whom were female. Participants were recruited to the Australian Genetics of Depression Study (www.geneticsofdepression.org.au) between 2016 and 2019. All study protocols were approved by the QIMR Berghofer Medical Research Institute Human Research Ethics Committee. The protocol for approaching participants through the DHS, enrolling them in the study, and consenting for all phases of the study (including invitation to future related studies) and accessing MBS and PBS records was approved by the Ethics Department of the Department of Human Services. The average age of participants was 43 years ± 15 years. Participants completed an online questionnaire that consisted of a compulsory module that assessed self-reported psychiatric history, clinical depression using the Composite Interview

Diagnostic Interview Short Form, and experiences of using commonly prescribed antidepressants. Further voluntary modules assessed a wide range of traits of relevance to psychopathology. Participants who reported they were willing to provide a DNA sample (75%) were sent a saliva kit in the mail. In the present study, we included 757 cases and 5,368 controls for whom genotype data were available. Samples from the AGDS were genotyped in three different genotyping centers using the same array (GSAMD-24v1-0_20011747). Genotype calling was performed with GenomeStudio. A common set of high QC markers between the different genotyping batches was obtained prior to joint imputation. Marker exclusion criteria (prior to imputation) included: unknown or ambiguous map position and strand alignment in a BLAST search, missingness > 5%, HWE test $P < 1x10^{-06}$, MAF < 1%, GenTrain score < 0.6. The Michigan imputation server (Das et al., 2016) was used to impute the genotypes using the HRCr1.1 as a reference panel. Individuals were excluded based on a high missingness (missing rate > 3%), inconsistent (and unresolvable) sex, or if deemed ancestry outliers from the European population (6 standard deviations from the first two genetic PCs. The GWAS was done employing a logistic regression using PLINK 1.9 (Purcell et al., 2007) and imputed dosage genotypes while correcting for the genotyping center and the first twenty ancestry PCs as covariates.

##### *BioVU | Davis, L. K. | Nashville, Tennessee, USA*

Vanderbilt University Medical Center (VUMC) is a tertiary care center that provides inpatient and outpatient care in Nashville, TN. The VUMC electronic health record (EHR) system was established in 1990 and includes data on billing codes from the International Classification of Diseases, 9th and 10th editions (ICD-9 and ICD-10), Current Procedural Terminology (CPT) codes, laboratory values, reports, and clinical documentation. In 2007, VUMC launched a biobank, BioVU, which links a patient’s DNA sample to their EHR. The BioVU Consent form is provided to patients in the outpatient clinic environments at VUMC. The form states policies on data sharing and privacy, and should a signature be obtained, makes any blood leftover from clinical care eligible for BioVU banking. The Vanderbilt University Medical Center Institutional Review Board oversees BioVU and approved this project (IRB201609). OCD case status was determined through a combination of ICD codes, medications, and natural language processing for EHR notes. Using ICD codes, cases were defined as individuals with any codes for OCD (ICD9: 300.3, ICD10: F42, F63.3, F45.22). Additional cases were gathered by first finding individuals with “obsessive-compulsive”, “obsessive compulsive”, or “OCD" in clinic notes, problem lists, discharge summaries, or clinical communications, where the first instance occurred before 55 years of age. Mentions of “OCD” were excluded if the keyword “osteochondritis” or “osteocondritis” occurred in the same document, or if certain ICD codes related to pervasive developmental disorders, osteochondritis dissecans, and bariatric surgery status (ICD9: 299*, 732.7, 649.2, 649.20, 649.21, 649.23, 649.24, V45.86, ICD10: F84*, M93.20, O99.840, O99.841, O99.842, O99.843, O99.844, O99.845, Z98.84) occurred on the same date as the mention. Keyword mentions were excluded if negating terms or terms related to familial relationships occurred within 30 characters on either side of the mention. Next, these individuals were required to have OCD medication or cognitive behavioral therapy in their EHR. Finally, individuals with metabolic disorder codes (ICD9: 277.89, 277.9, ICD10: E88.9, E88.89) were excluded. Controls were defined as any individual without OCD codes, metabolic disorder codes, “obsessive-compulsive”, “obsessive compulsive”, or “OCD" in their clinical notes, and without evidence of OCD medication. The initial sample included 1062 cases (median age in years across EHR: 32.75, 58.6% female) and 40,316 controls (median age in years across EHR: 53.57% female). We obtained genotype information on 94,474 BioVU individuals genotyped on the Illumina MEGA EX array. Using PLINK v1.95 (Purcell et al., 2007), genotypes were filtered for SNP and individual call rates, sex discrepancies, and excessive heterozygosity. We selected individuals of European ancestry using principal component analysis (PCA) implemented in FlashPCA (Abraham & Inouye, 2014) and confirmed the absence of genotyping batch effects through logistic regression with batch as the phenotype. Autosomes were imputed to the HRC panel using Michigan Imputation Server (Das et al., 2016) in five batches. After imputation, genotypes were converted to hard calls with PLINK using the default threshold settings. SNPs with multiple alleles or imputation quality less than $R^2$ of 0.3 were excluded. Next, SNPs with minor allele frequency (MAF) less than 0.005 or genotyping rates less than 0.98 were excluded. Individuals with call rates less than 0.98 were excluded. We ran a series of PCA to determine BioVU individuals of European genetic ancestry. First, we performed PCA using FlashPCA on BioVU combined with CEU, YRI, and CHB reference sets from 1000 Genomes Project Phase 3 (The 1000 Genomes Project Consortium, 2015). PCs were scaled so that the axes could be interpreted as proportions of genetic ancestry. We selected BioVU individuals who were within 40% of the CEU cluster along the CEU-CHB axis and within 30% of the CEU cluster on the CEU-YRI axis, generating a once-PCA filtered European set. To ensure subsequent steps would remove SNPs associated with reduced quality rather than cryptic population substructure, we filtered the previously identified BioVU European cluster to identify individuals falling within the CEU, TSI, and GIH 1000 genomes populations, producing a twice filtered European set. Using the twice-filtered European set we conducted a series of SNP checks. First, we filtered individuals with IBS greater than 0.2 and calculated PCs to use as covariates. Next, we checked for imputation batch effects by conducting pairwise logistic regression of the five imputation batches using sex and top 10 PCs as covariates. SNPs with p-values less than 0.001 in the additive model were flagged. We then compared MAF between BioVU and the CEU reference population. Any SNPs with a MAF difference greater than 0.1 were flagged. SNPs with a Hardy-Weinberg Equilibrium (HWE) p-value less than $1x10^{-10}$ were flagged. Finally, the flagged SNPs from the batch effect, MAF difference, and HWE were excluded from the once-PCA filtered BioVU European set, resulting in 9,386,383 SNPs for analysis. The final dataset consisted of 1041 cases and 38,613 controls. To account for the large case-control imbalance, we used SAIGE (Zhou et al., 2018) for the GWAS. Covariates included were sex, median age across medical records, and top 10 PCs.

##### *Children's Hospital of Philadelphia (CHOP) | Hakonarson, H; Gur, R. E. | USA | 25840117, 2588255*

Data were collected in the Philadelphia Neurodevelopmental Cohort (PNC) from the Children's Hospital of Philadelphia (CHOP) (Satterthwaite et al., 2016). 9428 participants aged 8-21 years completed a computerized structured screener based on the Kiddie-Schedule for Affective

Disorders and Schizophrenia (K-SADS) called GO-ASSESS (Calkins et al., 2014, 2015), which asked about the lifetime presence of any obsessive or compulsive symptoms as well as the severity, level of impairment and age of onset of symptoms. Samples were genotyped using Illumina Human610-Quadv1_B BeadChip array. As previously described (Burton et al., 2021), QC filtering was done using standard methods including removing SNPs with MAF < 0.01, and imputation quality < 0.6. GWAS was conducted using R (v3.5.1) with a logistic regression and using age, sex, and 3 PC scores as covariates.

##### *COGA | Porjesz, B.; Foroud, T.; Agrawal, A. | USA | 31270906, 31090166*

Participants of the Collaborative Study on the Genetics of Alcoholism (COGA) were recruited from 7 sites across the U.S. Institutional review boards at all sites approved the study and all participants provided informed consent. OCD diagnosis was determined using DSM-III-R or DSM-IV. COGA European ancestry (EA) data were genotyped using three arrays: Illumina Human1M array, Illumina Human OmniExpress V1 array, Smokescreen genotyping array (two different batches) (Lai et al., 2019, 2020). To confirm pedigree structure and estimate PCs of population stratification, we used a set of independent (defined as linkage disequilibrium (LD) r2 < 0.5) and high-quality variants (MAF > 10%, HWE p-value > 0.001, missing rate < 2%, 47,000 variants) genotyped on all arrays. Pairwise identity by descent was computed using PLINK (Chang et al., 2015; Purcell et al., 2007) to confirm family relationships and family structures were updated if necessary. PCs were calculated using Eigenstrat (Price et al., 2006). Based on the first two PCs, each individual was assigned an ancestry (i.e., African ancestry, European ancestry, or other) if they clustered with the corresponding 1000 Genomes project populations. Before imputation, Mendelian errors were detected using Pedcheck (O'Connell & Weeks, 1998) and inconsistencies were set to missing. Palindromic variants were excluded to avoid strand ambiguities. Then variants with missing rates > 5%, MAF < 3%, and HWE P values < 0.0001 were excluded. SHAPEIT2 (Delaneau et al., 2013) was used to phase haplotypes of each sample then Minimac 3.32 (Das et al., 2016) was used for imputation. 1000 genomes project samples were used as the reference panel (Phase 3, version 5; (The 1000 Genomes Project Consortium, 2015)). Samples on each array were imputed separately due to the different array content. After imputation, variants with $R^2 < 0.60$ were excluded. Genotype probabilities were converted to genotypes if they were > 0.90, then Mendelian error checking was performed again. All genotype and imputed variants with missing rates less than 20%, MAF ≥ 1%, and HWE P values > 0.000001 were kept in analyses. Unrelated samples selected from COGA EA datasets were included in analysis (131 cases and 1358 controls) using PLINK. Sex, age, the first 10 PCs, and array indicator were included as covariates.

##### *EGOS | Grice, D. E. | Sweden | 31907560*

The EGOS source population consists of individuals born in Sweden between January 1954 and December 1998 that presented at least two diagnoses of OCD or chronic tic disorders (CTD) at different time points in the Swedish National Patient Register (NPR) and were followed between January 1997 and December 2012 (N = 20,374). The International Classification of Disease (ICD) was used for the identification of OCD cases (F49 ICD version 10). Detailed information for each individual was obtained through linkage to the Swedish national registers, e.g., family relatedness, identification of additional psychiatric diagnoses, medical diagnoses, birth-related variables, and relevant demographic and social data. To create an epidemiologically valid subset of the source cohort that also includes biospecimens and additional phenotyping, individuals were contacted from within the source population. Study participants could elect whether to donate blood or saliva. Individuals are aged 16-64 years. All samples were genotyped using the Global Screening Array (GSA). Further description of the EGOS cohort has been represented by Mahjani et al. (Mahjani et al., 2020) The dataset that was used in this meta-analysis consists of 1026 OCD cases from EGOS and 1208 controls from LifeGene. EGOS was supported by a grant from the Beatrice and Samuel A. Seaver Foundation to DEG. Ethical approvals were obtained from the Institutional Review Board (IRB) at the Icahn School of Medicine at Mount Sinai, New York, NY, and the Regional Ethical Review Board in Stockholm.

##### *EPOC | Wagner, M.; Kathmann, N. | Germany | 30744714, 30008679, 29890378, 29721727, 29159055, 28541065, 28481032, 28160276*

The EPOC (Endophenotypes of OCD) sample comprises (epi)genetic and deep phenotype data of OCD patients, unaffected first-degree relatives of OCD patients and healthy controls that were collected at two sites in Germany (Berlin and Bonn) between 2014 and 2017. In the present analysis, data from 195 patients with OCD (55.9 % female, 44.1 % male) and 204 controls (63.2 % female, 36.8 % male) were included. Mean age was 33.37 (SD = 10.76; range: 18–64) for patients and 34.72 (SD = 12.64; range: 18–64) for controls. Lifetime comorbidity rates of patients were 60.0% for depression; 11.3% for panic disorder or agoraphobia; 7.7% for tic disorder; 7.7% for specific phobia; 6.7% for social phobia; 4.6% for generalized anxiety disorder; 4.1% for PTSD; 4.1% for anorexia; 3.1% for hypochondria; and 2.6% for hoarding disorder. OCD patients were recruited via the outpatient clinics at the Department of Psychology of Humboldt University, Berlin and at the Department of Psychiatry and Psychotherapy of the University Hospital Bonn. Healthy volunteers were recruited from the general population via public advertisements in the same cities. All participants were examined by trained clinical psychologists using the Structured Clinical Interview for DSM-IV (SCIDI) to assess OCD diagnosis and potential comorbidities. To establish cross-site reliability of clinical ratings, all instructions were standardized, and raters completed assessments of four training videos. Patients were only included if they: (a) met diagnostic criteria of OCD based on the SCID-I interview; (b) were free of past or present psychotic, bipolar or substance-related disorders; (c) did not take neuroleptic medication for the previous four weeks; and (d) did not use benzodiazepines in the prior two weeks. Healthy controls were excluded if they: (a) took any psychoactive medication in the previous three months; (b) had a current Axis I disorder; (c) had a lifetime diagnosis of OCD or tic disorder; or (d) had a family history of OCD. Written informed consent was obtained and participants were compensated for their time. The study was in accordance with the revised Declaration of Helsinki and approved by the local ethics committees of the Charité University Medicine Berlin and the University Hospital Bonn. Genotyping of cases and controls was performed on the Illumina Global Screen Array (GSA) at the Life Brain Center, Bonn. Genotype quality control was done using Plink-1.9 (Purcell et al., 2007), and R (version 3.5.1). We checked the data for sex inconsistencies and grossly failing markers (call rate < 0.5). Individuals with a call rate of < 0.95 were removed. The heterozygosity rate for each subject was calculated; outliers (±3 SD from the mean heterozygosity rate) were identified and removed. On marker level, SNPs were removed if at least one of the following conditions was true: significant difference of missing rate between cases and controls, call rate < 0.95; deviation of HWE ($P < 1x10^{-06}$); and MAF < 0.05 (computed separately in cases and controls). Furthermore, all A/T or C/G SNPs were removed. To check for population stratification, PCA was performed, and the first two PCs were checked for outliers. Seven individuals, whose non-European ethnicity was additionally validated based on their demographic data, were excluded. The genotyped data were imputed on the Michigan Imputation Server (Das et al., 2016) using the 1000 Genomes Phase 3 (version 5) reference panel (The 1000 Genomes Project

Consortium, 2015). GWAS was conducted using SNPTEST (version 2.5.2) (Marchini et al., 2007) with the first four PCs as covariates.

##### *EstBB | Metspalu, A. | Estonia*

The Estonian Biobank (EstBB) is a population-based cohort with a rich variety of phenotypic and health-related information collected for each participant (Leitsalu et al., 2015). At recruitment, participants signed a consent allowing follow-up linkage of their electronic health records (EHRs), thereby providing a longitudinal collection of their phenotypic information. The EstBB database includes health records from the national Health Insurance Fund Treatment Bills (from 2004 onwards), Tartu University Hospital (from 2008), and North Estonia Medical Center (from 2005), and data from different registries (causes of death, cancer, etc.). For all the participants EstBB provides information on the diagnoses in ICD-10 coding and information on drug dispensing data, including drug ATC codes, prescription status and purchase date (if available).

Genotyping of DNA samples from the Estonian Biobank was done at the Core Genotyping Lab of the Institute of Genomics, University of Tartu using the Illumina Global Screening Arrays (GSAv1.0, GSAv2.0, and GSAv2.0_EST). Samples were genotyped and then PLINK format files were created using Illumina GenomeStudio v2.0.4. During the quality control all individuals with call-rate < 95% or mismatching sex that was defined based on the heterozygosity of X chromosome and sex in the phenotype data, were excluded from the analysis. Variants were filtered by call-rate < 95% and HWE p-value $< 1x10^{-04}$ (autosomal variants only). Variant positions were updated to Genome Reference Consortium Human Build 37 and all variants were changed to be from TOP strand using reference information provided by Dr. Will Rayner from the University of Oxford (https://www.well.ox.ac.uk/~wrayner/strand/). After QC the dataset contained 202,910 samples for imputation. Before imputation variants with MAF < 1% and Indels were removed. Prephasing was done using the Eagle v2.3 software (Loh, Danecek, et al., 2016; Loh, Palamara, et al., 2016). The number of conditioning haplotypes Eagle2 uses when phasing each sample was set to: –Kpbwt=20000. Imputation was done using Beagle v.18May20.d20 (B. L. Browning et al., 2018; S. R. Browning & Browning, 2007) with effective population size $N_{Effective} = 20{,}000$. As a reference, Estonian population specific imputation reference of 2297 WGS samples was used (Mitt et al., 2017). Further, EstBB samples were combined with the 1000 genomes phase 3 dataset for ancestry analysis. Genetic principal components were calculated using a subset of quality controlled and pruned genotyped SNPs. This was further used to identify and remove samples that deviated from the main cluster.

For the genome-wide study of OCD, EstBB cases were defined as participants with F42* ICD10 diagnosis codes in their EHRs. We conducted a GWAS on individuals of European ancestry, including 772 cases and 196,079 controls. The analysis was performed with the REGENIE software (2.2.4) (Mbatchou et al., 2021) including related individuals and adjusting for the first 10 PCs of the genotype matrix, as well as for birth year, birth year squared and sex.

##### *FinnGen | Kaprio, J. A. | Finland*

Finnish samples are population-based biobank samples collected between August 2017 and August 2019 (collection further ongoing), including legacy samples collected since the 1980s. The Ethical Review Board of the Hospital District of Helsinki and Uusimaa approved the FinnGen study protocol Nr. HUS/990/2017. The FinnGen project was approved by Finnish Institute for Health and Welfare (THL), approval numbers THL/2031/6.02.00/2017, amendments THL/341/6.02.00/2018, THL/2222/6.02.00/2018, and THL/283/6.02.00/2019. The here included data has not been published before but a general description of the FinnGen study can be found elsewhere (Mars et al., 2020). Cases were recruited through hospital records (inpatient and outpatients) from 1970 onwards, based on clinical diagnoses used in patient care, supplemented by cause of death diagnoses (if clinical diagnosis was underlying and/or contributing cause). Case definitions were based on a diagnosis of ICD-10 F42 and ICD-8 3003, there were no case exclusion criteria. The mean age of cases was 45.36 $(SD = 15.44)$ years with a minimum age of

16.5 and a maximum age of 91.4, including 330 males and 460 females. Controls were defined as all other participants in FinnGen with GWAS data. The mean age of controls was 60.02 ($SD = 17.28$) years with a minimum age of 0 and a maximum age of 105.73, including 71,252 males and 90,138 females. Genotyping of cases and controls was performed in multiple batches on multiple arrays in multiple centers. For genotype quality control the following filters were applied to the data: exclusion of chromosome X, exclusion of variants with INFO score < 0.95, with missingness > 0.01, or with MAF < 0.05. To check for population stratification, PCA was performed, and a Bayesian algorithm was used to spot outliers. Individuals were excluded if there was a mismatch between imputed sex and sex in registry data. For calculating the genetic relationship matrix, we used the genotype dataset where genotypes with GP < 0.95 have been set missing. Only variants imputed with an INFO score > 0.95 in all batches were used. Variants with > 3% missing genotypes were excluded as well as variants with MAF < 1%. The remaining variants were LD pruned with a 1Mb window and $r^2$ threshold of 0.1. Imputation was performed using Eagle 2.4/Beagle 4.1 (B. L. Browning, 2017; Loh, Danecek, et al., 2016) using Finnish WGS (depth up to 30x) samples as reference with a total amount of 16962023 variants. GWAS was performed using SAIGE (v0.35.8.8) (Zhou et al., 2018) including age, sex, the first 10 PCs, genotyping batch and kinship matrix as covariates.

##### *HUNT | Zwart, J.-A. | Norway | 22879362*

The Trøndelag Health Study (HUNT) consists of three different population-based health surveys conducted in the county of Nord-Trøndelag, Norway over approximately 20 years (HUNT1: 1984-1986, HUNT2: 1995-1997 and HUNT3: 2006-2008). The HUNT study was approved by the Regional Committee for Medical and Health Research Ethics, Norway (2015/575). For each survey, the entire adult population (≥ 20 years) was invited to participate by completing questionnaires, attending clinical examinations and interviews. Participation rates in HUNT1,

HUNT2 and HUNT3 were 89.4% (N = 77,212), 69.5% (N = 65 237) and 54.1% (N = 50 807), respectively. Taken together, the study included more than 120,000 different individuals from NordTrøndelag County. Biological samples including DNA have been collected for approximately 70000 participants. The entire HUNT Study has been described in more detail elsewhere (Krokstad et al., 2013). For the present study, we included participants from HUNT2 and HUNT3. Cases and controls were defined by linkage to hospital diagnostic registries from the time period 1987-2017. Cases were defined as those with a hospital diagnosis of obsessive-compulsive disorder (ICD10 code F42). Controls were defined as those without ICD10 code F42 and without ICD-9 code 300 ("anxiety, dissociative and somatoform disorders"). The study was approved by the Regional Committee for Medical and Health Research Ethics (ref. 2015/575). In total, DNA from 71,860 HUNT samples was genotyped at the Genomics Core Facility at the Norwegian University of Science and Technology using one of three different Illumina HumanCoreExome arrays (HumanCoreExome12 v1.0, HumanCoreExome12 v1.1 and UM HUNT Biobank v1.0). Samples that failed to reach a 99% call rate, had contamination >2.5% as estimated with BAF Regress (Jun et al., 2012), large chromosomal copy number variants, lower call rate of a technical duplicate pair and twins, gonosomal constellations other than XX and XY, or whose inferred sex contradicted the reported gender, were excluded. Samples that passed quality control were analyzed in a second round of genotype calling following the Genome Studio quality control protocol described elsewhere (Guo et al., 2014). Genomic position, strand orientation and the reference allele of genotyped variants were determined by aligning their probe sequences against the human genome (Genome Reference Consortium Human genome build 37 and revised Cambridge Reference Sequence of the human mitochondrial DNA; http://genome.ucsc.edu) using BLAT (Dunham et al., 2012). PLINK v1.90 (Purcell et al., 2007) was then used to exclude variants if their probe sequences could not be perfectly mapped, cluster separation was < 0.3, Gentrain score < 0.15, showed deviations from HWE in unrelated samples of European ancestry with p-value < 0.0001), had a call rate < 99%, or another assay with higher call rate genotyped the same variant. Ancestry of all samples was inferred by projecting all genotyped samples into the space of the PCs of the Human Genome Diversity Project (HGDP) reference panel (938 unrelated individuals; downloaded from http://csg.sph.umich.edu/chaolong/LASER/) (Li et al., 2008; Wang et al., 2014) using PLINK. Recent European ancestry was defined as samples that fell into an ellipsoid spanning exclusively European populations of the HGDP panel. The different arrays were harmonized by reducing to a set of overlapping variants and excluding variants that showed frequency differences > 15% between data sets, or that were monomorphic in one and had MAF > 1% in another data set. The resulting genotype data were phased using Eagle2 v2.3 (Loh, Danecek, et al., 2016). Imputation was performed on the 69,716 samples of recent European ancestry using Minimac3 (v2.0.1, http://genome.sph.umich.edu/wiki/Minimac3, Das et al., 2016) with default settings (2.5 Mb reference based chunking with 500kb windows) and a customized Haplotype Reference consortium release 1.1 (HRC v1.1) for autosomal variants and HRC v1.1 for chromosome X variants (McCarthy et al., 2016). The customized reference panel represented the merged panel of two reciprocally imputed reference panels: (a) 2201 lowcoverage whole genome sequences samples from the HUNT study and (b) HRC v1.1 with 1023 HUNT WGS samples removed before merging. We excluded imputed variants with Rsq < 0.3 or minor allele count < 1, resulting in 24.2 million well-imputed variants. After restricting to those with available phenotype information 66,476 individuals (284 cases and 66,192 controls) were included in the analysis. Association analyses were conducted using SAIGE (Zhou et al., 2018), a generalized mixed effects model approach, to account for cryptic population structure and relatedness when modeling the association between genotype probabilities (dosages) and OCD. Models were adjusted for sex, birth year, genotyping batch and four PCs. PCs were computed using PLINK.

##### *IOCDF-GC and 610k_trio | Multiple | Multiple | 22889921, 28761083*

The results of the International OCD Foundation-Genetics Consortium (IOCDF-GC) study was previously published (Arnold et al., 2018; Stewart et al., 2013). The IOCDF-GC case-control cohort (IOCDF-GC) consists of 1519 European ancestry cases and 3541 matching controls from IOCDF-GC and three cohorts previously genotyped, including the Alzheimer’s Disease Genetics Initiative (Lee et al., 2008), the Center for Applied Genomics (CAG) at Children’s Hospital of Philadelphia (CHOP) (Gur et al., 2012), and the Breast and Prostate Cancer Cohort Consortium (BPC3; (Schumacher et al., 2011)) The IOCDF-GC trio sample (610k_trio) consists of 323 European ancestry complete trios. All cases and trios were recruited predominantly from OCD specialty clinics, and controls were recruited from Bonn, Germany and from Cape Town, South Africa. This work was approved by the relevant IRBs at all participating sites, and all participants provided written informed consent. For study inclusion, all cases and trio probands were required to have a DSM-IV diagnosis of OCD. The controls from Bonn had an absent lifetime history of all axis I disorders and the South African controls were diagnostically unscreened. The sample description of the three cohorts refer to the primary article. All samples were genotyped on Illumina Human610-Quadv1_B SNP array (Illumina, San Diego, CA, USA). Standard quality control (QC) protocol was conducted with PLINK (Purcell et al., 2007). Samples were removed for call rates < 98%, sex discrepancy and ambiguous genomic sex, related samples with pihat > 0.2. SNP QC included removing monomorphic SNPs, CNV-targeted SNP probes, SNPs with genotyping rate < 98%, SNPs with MAF < 0.01, strand-ambiguous SNPs with significant allele frequency differences or aberrant LD correlations with adjacent SNPs based on the entire HapMap2 reference panel, SNPs with $P < 1x10^{-06}$ in HWE test among controls or $P < 1x10^{-10}$ among cases, SNPs with differential missing rate between cases and controls (> 0.02), and SNPs with batch effect ($P < 1x10^{-05}$) between different control cohorts. Multidimensional scaling (MDS) analyses were performed in PLINK2 (Chang et al., 2015) and samples were removed when they were significant outliers in the first five MDS dimensions or when there were no matching cases or controls on these MDS dimensions. The genotyped data were phased by SHAPEIT2 (Delaneau et al., 2013) and imputed by Minimac3 (Das et al., 2016) using the HRC (McCarthy et al., 2016) release 1.1 as the reference panel. GWAS was performed on SNPs with INFO score > 0.8 and MAF > 0.01, using a logistic regression model in PLINK2 (Chang et al., 2015) with the first five and the 7th MDS components as covariates. Same QC was conducted on the 323 trios, with an additional filter of removing SNPs with Mendelian errors. In each trio, the transmitted alleles and untransmitted alleles were converted into one case and one pseudo-control. Phasing and imputation were conducted on the cases and pseudo-controls in the same way as the case-control cohort. GWAS was performed without covariates.

##### *iPSYCH | Borglum, A.D.; Mors, O.; Mattheisen, M. | Denmark*

In the scope of the Danish OCD and Tourette Study (DOTS) within The Lundbeck Foundation Initiative for Integrative Psychiatric Research (iPSYCH), Danish nation-wide population-based case-cohort samples were collected and genotyped. The study was approved by the Regional Scientific Ethics Committee in Denmark and the Danish Data Protection Agency. All analyses of the samples were performed on the secured national GenomeDK high performance computing cluster in Denmark (https://genome.au.dk). Samples stem from the newly updated baseline cohort iPSYCH2015 including singletons born between 1981 and 2008 who were born to a known mother and resided in Denmark on their first birthday. Genetic information was obtained by the

Statens Serum Institut (SSI) at the Danish Neonatal Screening Biobank (DNSB) from heel prick blood samples that had been collected from all newborn babies in Denmark. The genetic information was linked with the Danish Civil Registration System and thereby coupled with the Danish Psychiatric Central Research Register which collects patient data of individuals treated in psychiatric hospitals (from 1969 onwards) or in outpatient psychiatric clinics (from 1995 onwards). See (Bybjerg-Grauholm et al., 2020; C. B. Pedersen et al., 2018) for a detailed description of the overall cohort, array genotyping and quality control. Cases included in the present study were not primarily ascertained for OCD; OCD cases were drawn from cases that also presented a diagnosis of one of the core disorders iPSYCH primarily collected for and from randomly ascertained population “controls” with a diagnosis of OCD. OCD cases were diagnosed by a healthcare professional and met ICD10 (F42) criteria, controls were randomly from the controls and excluded individuals with an F42 diagnosis. Samples for iPSYCH2012 were genotyped on the PsychChip v 1.0 array (Illumina, San Diego, CA, USA), while samples for iPSYCH2015i were genotyped on the Illumina Global Screening (GSA) v2 Array, both at the Broad Institute of MIT and Harvard (Cambridge, MA, USA). Genotype calling of markers with MAF > 0.01 was performed by merging call sets from GenCal (Illumina: Illumina GenCall Data Analysis Software) and Birdseed (Korn et al., 2008), and less frequent variants were called with zCall (Goldstein et al., 2012). Genotyping and data analysis was performed in 23 waves. Genotype data were processed using the Rapid Imputation and COmputational PIpeLIne for Genome-Wide Association Studies (RICOPILI; (Lam et al., 2020) to perform stringent QC, imputation, PC analysis, and primary association analysis. SHAPEIT (Delaneau et al., 2011) was used for phasing, imputation was conducted with IMPUTE2 (Howie et al., 2009), using the HRC as a reference panel (McCarthy et al., 2016). We removed samples with a call rate below 95%, with a sex mismatch, between the sex obtained from genotype data and from the register data, as well as related individuals. PC analysis was used to exclude ancestral outliers of non-European descent, excluding all individuals exceeding eight standard deviations from the mean on the first three PCs. The GWAS was performed using RICOPILI and included 10 PCs as covariates. The final dataset included 4509

OCD cases and 38,392 controls, 7,544,427 SNPs, had a Lambda of 1.074 and a Lambda1000 of 1.009.

##### *Michigan/Toronto OCD Imaging Genomics Study | Noam Soreni; Gregory L. Hanna; Kate D. Fitzgerald; David Rosenberg; Paul D. Arnold | USA and Canada*

Individuals from the Michigan/Toronto OCD Imaging Genomics Study (Michigan/Toronto IGS) were recruited from four different academic child psychiatry sites: The Hospital for Sick Children, McMaster University, University of Michigan, and Wayne State University (Gazzellone et al., 2016). Cases were defined by a clinical investigator according to DSM-IV criteria. Samples were genotyped using three different arrays: HumanCoreExome, PsychArray and Omni2.5. Each genotyping array was processed separately; then imputed data from all arrays were combined. As previously described (Burton et al., 2021), QC filtering was done using standard methods including removing SNPs with MAF < 0.01, and imputation quality < 0.6. The GWAS was conducted using R (v3.5.1) with a logistic regression and using age, sex, and 3 PCs as covariates.

##### *Million Veteran Program | Stein, M.B.; Gelernter, J. | USA | 26441289*

The U.S. Department of Veterans Affairs (VA) Million Veteran Program (MVP) is collecting genetic and electronic health record (EHR) data in the U.S with ethical approval given by the Central VA Institutional Review Board (IRB) and site-specific IRBs (Gaziano et al., 2016; Harrington et al., 2019). All relevant ethical regulations for work with human subjects were followed in the conduct of the study, and informed consent was obtained from all participants. We used data release version 4; briefly, genotyping was performed with Affymetrix Axiom Biobank Array and quality control (Hunter-Zinck et al., 2020), phasing chromosomes with EAGLE2 (Loh, Danecek, et al., 2016) and imputation with Minimac3 (Das et al., 2016), using the 1000 Genomes Project reference panel, phase 3 (The 1000 Genomes Project Consortium, 2015). We selected European (EUR) ancestry data which were defined using PCA (Gaziano et al., 2016). For QC filtering, we set up imputation quality scores > 0.6, HWE filtering $> 5x10^{-05}$, MAF > 0.001, missing call rates for variants < 0.1, and missing call rates for samples < 0.1. Data were aligned to the GRCh37 reference genome. Cases were defined having at least one International Classification of Disease (ICD) outpatient code for OCD (ICD-9: 300.3; ICD-10: F42.2, F42.3, F42.8, F42.9) by any provider, excluding individuals suffering from schizophrenia. To optimize the number of cases to keep when removing individuals with kinship, which was calculated by KING (Manichaikul et al., 2010) for a minimum threshold of 0.0884 corresponding to a second-degree relationship, we implemented the following algorithm where cases have priority compared to controls: between a case and a control, we remove the individual as control; if there is relatedness between two cases, we remove the one that has the highest number of relationships with other individuals; we do the same between two controls. In conclusion, we obtained 5129 cases and 422,860 controls. GWAS was performed using PLINK 2.0 (Chang et al., 2015) setting age, sex, and the first ten PCs scores as covariates.

##### *MoBa | Ask, H. | Norway | 27063603*

The Norwegian mother, father and child cohort study (MoBa) is a population-based pregnancy cohort study conducted by the Norwegian Institute of Public Health. The establishment of MoBa and initial data collection was based on a license from the Norwegian Data Protection Agency and approval from The Regional Committees for Medical and Health Research Ethics. The MoBa cohort is now based on regulations related to the Norwegian Health Registry Act. Participants were recruited from all over Norway between 1999 and 2008. The women consented to participation in 41% of the pregnancies. The cohort now includes 114,500 children, 95,200 mothers and 75,200 fathers. Blood samples were obtained from the mothers and fathers at 17–18 weeks of gestation and from mothers and children (umbilical cord) at birth. For the current study we used genotype data from 17,000 randomly selected trios, genotyped in three batches on three different arrays (harvest12: Illumina HumanCoreExome12v1.1, harvest24: Illumina HumanCoreExome24v1.0, rotterdam1: Illumina Global Screening Array MD v.1.0.). harvest12 and harvest24 were genotyped at the Genomics core facility in Trondheim, Norway while the rotterdam1 samples were genotyped at ERASMUS MC, Rotterdam, Netherlands. PLINK version

1.90 beta 3.365 (Purcell et al., 2007) was used to conduct quality control, details of QC have been previously described by Helgeland et al. (Helgeland et al., 2019). Individuals were excluded if they had a genotyping call rate below 95% or autosomal heterozygosity greater than four standard deviations from the sample mean. SNPs were excluded if they were ambiguous (A/T and C/G), had a genotyping call rate below 98%, MAF of less than 1%, or HWE P-value less than $1x10^{-06}$. Population stratification was assessed using the HapMap phase 3 release 3 as a reference, by PCA using EIGENSTRAT (Price et al., 2006) version 6.1.4. Visual inspection identified a homogenous population of European ethnicity and individuals of non-European ethnicity were removed. Phasing was conducted using Shapeit (Delaneau et al., 2013) release 837 and the duoHMM approach was used to account for the pedigree structure. Imputation was conducted using the Haplotype reference consortium (HRC) release 1-1 as the genetic reference panel. The Sanger Imputation Server was used to perform the imputation with the Positional Burrows Wheeler Transform (PBWT). The phasing and imputation were conducted separately for each genotyping batch. A core homogeneous sample of European ethnicity across all batches and arrays were available for use in analysis (totals prior to analysis-specific exclusions for relatedness: $N_{Mothers} = 14{,}804$; $N_{Fathers} = 15{,}198$). OCD diagnosis was ascertained through linkage to the Norwegian Patient Registry (ICD-10 codes from specialist health care registered from 2008-2018). Case inclusion criteria were ICD-10 code F42 diagnosed at least once. Control inclusion criteria were no F-diagnosis. Control individuals were excluded if they were related to any of the cases (pihat > 0.2). Sex ratio in the cases was 40:60 (male:female) and in the controls 50:50. GWA analysis was based on 104 cases and 2193 controls. GWAS was performed using SAIGE (Zhou et al., 2018), including the first 10 PCs and genotyping batch as covariates.

##### *NORDiC-SWE | Crowley, J; Mataix-Cols, D.; Rück, C. | Sweden | 31424634*

A paper describing the rationale, design, and methods of the NORDiC study has been published previously (Mataix-Cols et al., 2020). NORDiC-SWE is the Swedish case-control arm of the study, and all samples were collected in Sweden between 2015 and 2019. This study was approved by the Regional Ethics Committee, Stockholm (EPN Stockholm) and the Institutional Review Board

(IRB) at the University of North Carolina at Chapel Hill and all subjects provided informed consent. OCD cases have a primary ICD-10 and/or DSM-5 diagnosis of OCD from a multidisciplinary specialist OCD team (established with a semi-structured instrument such as the MINI or the SCID). All patients were included in the study regardless of psychiatric comorbidity, as long as they fulfilled strict diagnostic criteria for OCD. Patients were excluded in cases of diagnostic uncertainty, such as OCD secondary to a neurological disorder or CNS insult, or where the differential diagnosis between OCD and an alternative condition was unclear. Our cases had a mean age at symptom debut of 12.1 years and 59% were female. Approximately 58% of patients had a documented psychiatric comorbidity. Controls were unrelated to any OCD case to the third degree and unaffected with OCD. Controls were excluded if they had a lifetime history of anorexia nervosa (controls were inherited from an anorexia GWAS). The controls included 95% females. The NORDiC-SWE cohort consists of 971 OCD cases and 2735 controls (LIFEGENE control batch 1: N = 1026 and LIFEGENE control batch 2: N = 1389). Subjects provided either blood or saliva for DNA extraction. All samples were genotyped on the Illumina Global Screening Array (GSA) at LIFE&BRAIN in Bonn, Germany. Samples were filtered for duplicates ($pi_{hat}$ ≥ 0.95) and cryptic relatedness ($pi_{hat}$ ≥ 0.2). We selected individuals of European ancestry using PCA implemented in PEDDY v0.4.3 (B. S. Pedersen & Quinlan, 2017). We defined variants as QC-failing if they met one of the following criteria: 1) maximum genotype missingness in a cohort > 0.02; 2) allele frequency < 0.001 in at least one cohort; 3) max – min allele frequency > 0.1 across all five cohorts; 4) max – min allele frequency > 0.03 across all three control cohorts; 5) genome-wide significant in a control vs. control synthetic GWAS. A total of 154,791 variants that met at least one of these criteria were excluded. We used the RICOPILI v2018_Dec_7.001 (Lam et al., 2020) pipeline to run an automated round of pre-imputation QC. The pre-imputation QC step involved a series of hard filters on variant and sample level data, including removing variants with pre-sample pruning call rate < 0.95, samples with call rate < 0.98, FHET outside of ± 0.20, samples with discrepancies between reported and derived sex, and post sample-pruning variants that meet any of the following : 1) call rate < 0.98, 2) missing difference > 0.02, 3) invariant positions, 4) MAF > 0.01, 5) HWE $p < 1x10^{-06}$ in controls, and 6) HWE $p < 1x10^{-10}$ in cases. The final dataset consisted of 647,335 variant calls across a total of 1997 cases and 3943 controls. RICOPILI's impute_dirsub module was used to conduct imputation using the Haplotype Reference Consortium (HRC) reference panel. In our imputation run we used EAGLE v2.3.5 (Loh, Danecek, et al., 2016) for pre-phasing, and minimac3 v2.0.1 (Das et al., 2016) for imputation. We derived 3 different imputed callsets from this process: 1) a set of high confidence imputed genotypes (2,771,425 SNPs), 2) 7,112,906 imputed best-guess genotypes with medium level accuracy, and 3) genotypes for variants where imputation accuracy is lowered in order to increase the total number of variants included in the imputation (resulting in 8,995,398 SNPs). We elected to run our GWAS across the largest dataset of imputed variants that were generated during the imputation process on a subset of samples that were of European ancestry. We conducted PCA on these samples across high-confidence imputed genotypes using the pacer_sub RICOPILI module and tested the first 20 PCs for significant association with sample case/control status (significant = p-value < 0.05/20). We identified PCs 1, 3 and 14 as significant predictors and used them as covariates in the GWAS analysis. We used RICOPILI's postimp_navi module to conduct the final GWAS. Summary statistics were well controlled across the separate GWAS. We noted a lambda of 1.01 and a lambda1000 of 1.01 across a total of 7,679,714 tested SNPs.

##### *NORDiC-NOR | Crowley, J.; Kvale, G.; Hansen, S. | Norway | 31424634*

NORDiC-NOR is the Norwegian case-control arm of the NORDiC study and all samples were collected in Norway between 2016 and 2019. The NORDiC-NOR study was approved by the

Norwegian Regional Committee for Medical and Health Research Ethics (IRB00001872 REK West) under project number 2018/52 REKVest (PI: Bjarne Hansen) and project number: 2014/75 REKVest (PI: Jan Haavik) and all subjects provided informed consent. OCD Cases have a primary ICD-10 and/or DSM-5 diagnosis of OCD from a multidisciplinary specialist OCD team (established with a semi-structured instrument such as the MINI or the SCID). All patients were included in the study regardless of psychiatric comorbidity, as long as they fulfilled strict diagnostic criteria for OCD. Patients were excluded in cases of diagnostic uncertainty, such as OCD secondary to a neurological disorder or CNS insult, or where the differential diagnosis between OCD and an alternative condition was unclear. The mean age at symptom debut was 17 years and 65% were female. Approximately 51% of patients had a documented psychiatric comorbidity. Controls reported no OCD or first-degree family members with OCD. Among the controls, 50% were female. Subjects provided either blood or saliva for DNA extraction. All samples were genotyped on the Illumina Global Screening Array at LIFE&BRAIN in Bonn, Germany. The pre-GWAS QC applied to the NORDiC-NOR dataset (482 cases, 343 controls in the raw data) was nearly identical to that applied to NORDIC-SWE data after merging of separate genotype data, consisting of pruning of cryptic relatedness, marking of samples that are of likely European ancestry and pruning of the dataset for single variants where there was suggestive evidence of technical biases or batch effects. In our cryptic relatedness QC step, we identified 4 samples that had a mean $pi_{hat}$ with other samples of ≥ 0.1, 74 samples that had evidence of being a sample duplicate ($pi_{hat}$ ≥ 0.95) and 8 samples from the remaining cohort with evidence of cryptic relatedness ($pi_{hat}$ ≥ 0.2). We found a total of 6263 variants overlapped between the merged PLINK fileset and the 1000 genomes data included in PEDDY (B. S. Pedersen & Quinlan, 2017) and identified a total of 368 cases and 315 controls with likely European ancestry. We performed variant-level QC on a PC-pruned subset of 340 cases and 307 controls, defining variants as failing if they met one of the following criteria: 1) maximum genotype missingness in a cohort > 0.02; 2) allele frequency of 0 in at least one cohort; 3) max – min allele frequency > 0.1. A total of 136,525 variants that met at least one of these criteria were excluded, leaving us with a final case/control dataset consisting of genotype calls across 479,358 variants. We used calls across these variants in samples that had been pruned for relatedness issues (not including standard pairwise relatedness issues as RICOPILI can detect these) as input for the GWAS (407 cases, 340 controls). Imputation and GWAS was performed analogously to the NORDiC-SWE and EGOS analysis. The final data set consisted of 365 cases and 315 controls. Imputed callsets resulted in 1) 3,043,464 high confidence imputed genotypes 2) 7,277,174 genotypes with medium level, and 3) 8,964,589 genotypes with a low imputation accuracy. We identified PCs 1, 2, 3 and 4 as significant predictors and used them as covariates in the NORDiC-NOR GWAS. The analysis resulted in a lambda of 1.00 and a lambda1000 of 1.01 across a total of 7,518,582 tested SNPs.

##### *OCGAS_all | Multiple | USA | 22889921*

Results of The OCD Collaborative Genetics Association Study (OCGAS) have been published previously (Mattheisen et al., 2015). For detailed sample- and analysis description refer to the primary article. In brief: The OCGAS sample consists of 986 cases from OCGAS, a family based sample, and 1023 controls from the Genomic Psychiatry Cohort (GPC; (Pato et al., 2013)), a population based sample. The inclusion of control samples was slightly altered in this publication compared to the original study. For study inclusion, probands were required to meet DSM-IV criteria for OCD with onset of obsessions and/or compulsions before the age of 18 years (mean

= 9.4 years; SD = 6.35), as evaluated by a PhD-level clinical psychologist using the Structured Clinical Interview for DSM-IV modified and extended to include additional symptom and diagnostic information. Several mental- and brain disorders were reasons for exclusion. Genotyping was performed at the Johns Hopkins SNP Center using Illumina's HumanOmniExpress bead chips (Illumina, San Diego, CA, USA). A stringent quality control protocol was followed, including checking the relatedness of samples, sex comparison, Mendelian inconsistencies etc. One sample was removed per related pair with pihat > 0.2. Multidimensional scaling analyses were performed on singleton OCD cases and unselected controls, as implemented in PLINK (Purcell et al., 2007). Samples were removed when they significantly deviated in the first two multi-dimensional scaling dimensions (> 4 SD from the mean). The genotyped data were phased by SHAPEIT2 (Delaneau et al., 2013) and imputed by Minimac3 (Das et al., 2016) using the HRC release 1.1 (McCarthy et al., 2016) as the reference panel. GWAS was performed on SNPs with info score > 0.8 and MAF > 0.01, using logistic regression model in PLINK2 (Chang et al., 2015) with the first four and the 9th MDS components as covariates. Ethics approvals for the OCGAS study were obtained from the Hopkins Medicine Institutional Review Boards, the Butler Institutional Review Board, the UCLA Institutional Review Boards, the Mass General Brigham Human Research Committee, the Columbia University Institutional Review Boards, and the National Institutes of Health Institutional Review Board (NIH IRB).

##### *OCGAS-nestadt | Nestadt G., OCGAS Consortium | USA*

The OCGAS-nestadt study was conducted at one of the five participating recruitment sites of the National Institute of Mental Health. The study was approved by the IRB boards at: Johns Hopkins University School of Medicine, Brown Medical School, New York State Psychiatric Institute and College of Physicians and Surgeons at Columbia University, University of California Los Angeles (UCLA) School of Medicine, Massachusetts General Hospital and Harvard Medical School, National Institute of Mental Health, and Keck School of Medicine at the University of Southern California. Samples were collected between 2007 and 2014. The sample comprised of trios (including an affected proband and both parents) or in some cases a proband and an unaffected sibling. Each case was evaluated by a MD- or PhD-level clinical psychologist using the Structured Clinical Interview for DSM-IV (SCID). The checklist of obsessions and compulsions from the Y-BOCS, refined to include the age of onset, offset, and severity of each symptom, as well as the Y-BOCS scores for the worst episode (lifetime) was recorded. Course and treatment response variables were also included. A similar model was used for evaluating tics and Tourette disorder. Axis I disorder diagnoses were assigned using the JHU Diagnostic Assignment Checklist, an instrument that documents the criteria for over 20 DSM-IV disorders; this instrument also was the primary tool for the diagnostic consensus procedure. The SCID-II was used to evaluate four personality disorders (schizotypic, obsessive-compulsive, avoidant, and dependent), and the FISC was used to obtain additional information about each participant from a knowledgeable informant. Children over the age of eight were assessed in the same way, except that the Kiddie-SADS was used in place of the SCID. Final diagnostic status was assigned based on the consensus of two psychiatrists or psychologists reviewing the case independently. The agreement between diagnosticians using the Diagnostic Assignment Checklist has been studied and found to be excellent for variables such as age at onset of OCD. The chance-corrected percent agreement between the diagnosticians for the diagnosis of OCD was K = 0.92; for age at onset of OCD, K = 0.88 (for age +/- 5 years), and Pearson's r = 0.71. The diagnostic information from each site was reviewed by one of the five members of the JHU diagnostic consensus committee to ensure comparability across sites. For study inclusion, probands were required to meet DSM-IV criteria for OCD - with onset of obsessions and/or compulsions before the age of 18 years (mean = 9.4 years; SD = 6.35). Subjects with an age-of-onset > 17 years, schizophrenia, severe mental retardation that does not permit an evaluation to characterize the psychiatric disorder, Tourette disorder or OCD occurring exclusively in the context of depression (secondary OCD) were excluded. In addition, individuals were removed from the sample if they were previously diagnosed with brain pathology including brain tumors, Huntington's disease, Parkinson's disease, or Alzheimer's disease. This resulted in a final sample of 212 cases and 212 controls. Samples were genotyped on the Illumina PsychChip array at USC. Ethics approvals for the OCGAS study were obtained from the Hopkins Medicine Institutional Review Boards. The

OCD Collaborative Genetics Association Study (OCGAS) is a collaborative research study and was funded by the following NIMH Grant Numbers: MH071507, MH079489, MH079487, MH079488 and MH079494.

##### *OCGAS-ab | Arnold P., Burton C. | Canada | 27777633, 31772171*

Samples in the OCGAS-ab cohort were collected between 2008-2015 and consist of trios. The study was approved by REB at Hospital for Sick Children. Cases were required to meet DSM-IV criteria for OCD and were selected based on the K-SADS and CY-BOCS questionnaires. Exclusion criteria for cases were age of onset < 18 years of age, psychosis, and history of severe neurological disorders other than Tourette's disorder. Of the 55 cases and 55 controls, 50% are female, the mean age was 16.38 (SD = 3.78). One individual presented a co-morbid diagnosis of ASD, 11 of ADHD, six of Tics/Tourette's syndrome, five of anxiety disorders, one of a eating disorder, two of depression, and two presented learning difficulties. Samples were genotyped on the Illumina PsychChip array at USC. Ethics approvals for the OCGAS study were obtained from the Hopkins Medicine Institutional Review Boards.

##### *OCGAS-gh | Grünblatt E., Walitza S. | Switzerland and Germany | 28065182, 29102815*

Participants in the early-onset OCD cohort were recruited at the Departments of Child and Adolescent Psychiatry of the Universities of Würzburg, Marburg, Aachen, and Freiburg in Germany and Zurich in Switzerland. Informed written consent was obtained in all cases by the participants or their parents. The study was approved by the ethical commissions of all involved universities in accordance with the latest version of the Declaration of Helsinki, including an ethical permission granted by the Ethic Committees from Aachen, Würzburg, Marburg, Freiburg, and the Cantonal Ethic Commission of Zürich (Ref. Nr. 39/97, 140/3 and EK: KEK-ZHNr. 2010-0340/3). Samples were collected between 2000 and 2016 and resulted in 56 cases and 56 controls in trios or case-control samples. Patients were included if they fulfilled the diagnostic criteria for current OCD according to DSM-4 and ICD-10. To assess OCD diagnostic criteria, early-onset OCD patients and parents were interviewed separately by senior clinicians with a semi-structured diagnostic interview of psychiatric disorders in children and adolescents (Kinder-DIPS; children and parents version) (Margraf et al., 2017); the patients and parents located in Zürich underwent the German version of a semi-structured clinical interview (K-SADS-PL) (Kaufman et al., 2000) . In addition, severity and additional characteristics of OCD symptoms were evaluated with the Children's Yale-Brown Obsessive Compulsive Scale (CY-BOCS) (Scahill et al., 1997). Kinder-DIPS or K-SADS-PL was used to screen for the existence of comorbid disorders (affective-, anxiety-, eating- and tic disorders, attention-deficit/hyperactivity disorder, conduct-, oppositional disorder, as well as substance use, abuse, psychosis and somatic diseases) in children and adolescents. CASCAP-D was used to screen for autistic spectrum disorders (Schmidt et al., 2000). Present and lifetime Tourette's syndrome and tic disorders were assessed with the adapted German version of the Child and Adult Schedule for Tourette and Other Behavioral Syndromes (STOBS) and the Yale Global Tic Severity Scale (YGTSS) (Leckman et al., 1989) in the Zürich patients. Case exclusion criteria were a lifetime history of Tourette's syndrome, psychotic disorder, autism spectrum disorder, mental retardation (IQ < 70) or alcohol dependence. Patients with comorbid disorders were only included if OCD was the primary diagnosis. Controls were excluded if they presented a major psychiatric disorder or an IQ < 70. All pediatric OCD patients received cognitive behavior therapy, while when insufficient, drug treatment was added, most commonly with an SSRI. Samples were genotyped on the ILMN PsychChip array and genotyped at USC in one batch.

##### *OCD-WWF | Domschke, K; Berberich G. | Germany*

For the obsessive-compulsive disorder - Windach Würzburg Freiburg (OCD-WWF) study, 129 inpatients with OCD (mean age ± SD: 34.47±11.86 years; 66 female) were recruited at the

Psychosomatic Hospital Windach, Windach, Germany, between 2014 and 2017. OCD diagnosis was ascertained on the basis of a structured clinical interview according to DSMIV criteria (SCID-I) by experienced psychiatrists and/or clinical psychologists. Inclusion criteria were age at inclusion between 18 and 80 years and European descent (self-report up to third generation). Exclusion criteria comprised severe somatic and neurological disorders, the consumption of illegal drugs, and pregnancy. Comorbid tic disorder, trichotillomania, skin-picking disorder or other current axis I diagnoses except for depression (n = 71), specific phobias (n = 10), generalized anxiety disorder (n = 1), social phobia (n = 5), panic disorder (n = 2), agoraphobia (n = 5) or posttraumatic stress disorder (n = 3) were excluded. The study was approved by the ethics committee of the University of Würzburg, Germany and was conducted according to the ethical principles of the Helsinki Declaration. All patients gave written informed consent prior to participation. Cases and controls were genotyped on Illumina's Global screening array. GWAS analysis was performed using RICOPILI (Lam et al., 2020), employing standard parameters. First, we ran an automated round of pre-imputation QC. The pre-imputation QC step involved a series of hard filters on variant and sample level data, including removing variants with pre-sample pruning call rate $< 0.95$, samples with call rate $< 0.98$, FHET outside of $\pm$ 0.20, samples with discrepancies between reported and derived sex, and post sample-pruning variants that meet any of the following : 1) call rate $< 0.98$, 2) missing difference $> 0.02$, 3) invariant positions, 4) MAF $>$

0.01, 5) HWE $p < 1E-6$ in controls, and 6) HWE $p < 1E-10$ in cases. RICOPILI's impute_dirsub module was used to conduct imputation using the Haplotype Reference Consortium (HRC) reference panel. RICOPILI's impute_dirsub module was used to conduct imputation using the 1000's genomes (1000G) reference panel (The 1000 Genomes Project Consortium, 2015). We conducted PCA on these samples across high-confidence imputed genotypes using the pacer_sub RICOPILI module and tested the first 20 PCs for significant association with sample case/control status. In a final step, we used RICOPILI's postimp_navi module to conduct the GWAS analysis.

##### *Psych_Broad | Mathews C | Netherlands, Italy, USA, Spain*

Psych_Broad sample consists of 1396 European ancestry cases and 4009 population matched controls. The cases were recruited predominantly from OCD specialty clinics in the US, Spain, The Netherlands, and Italy. The Spanish controls were part of the Mental-Cat clinical sample or the INSchool population-based cohort. A total of 1757 controls from the Mental-Cat cohort (60.3% males) were evaluated and recruited prospectively from a restricted geographic area at the Hospital Universitari Vall d'Hebron of Barcelona (Spain) and consisted of unrelated healthy blood donors. The INSchool sample consists of 765 children (76.2% males) from schools in Catalonia. Both studies have been approved by the Clinical Research Ethics Committee (CREC) of Hospital Universitari Vall d'Hebron. All methods were performed in accordance with the relevant guidelines and regulations and written informed consent was obtained from participant parents before inclusion into the study. Detailed information has been published previously (Bosch et al., 2021; Rovira et al., 2020). Genomic DNA samples were obtained either from peripheral blood lymphocytes by the salting out procedure or from saliva using the Oragene DNA Self-Collection

Kit (DNA Genotek, Kanata, Ontario Canada). DNA concentrations were determined using the Pico- Green dsDNA Quantitation Kit (Molecular Probes, Eugene, OR). All cases were required to have a DSM-IV diagnosis of OCD. The population based unscreened controls were recruited from the same countries. All samples were genotyped on Infinium PsychArray-24 at Broad Institute (Cambridge, MA, USA). Standard quality control (QC) protocol was conducted with PLINK2 (Chang et al., 2015). Samples were removed for call rates < 98%, sex discrepancy and ambiguous genomic sex, related samples with pihat > 0.2. SNP QC included removing monomorphic SNPs, SNPs with genotyping rate < 98%, SNPs with MAF < 0.01, SNPs with $P < 1x10^{-06}$ in HWE test among controls or $P < 1x10^{-10}$ among cases, SNPs with differential missing rate between cases and controls (> 0.02), and SNPs with batch effect ($P < 1x10^{-05}$). Multidimensional scaling (MDS) analyses were performed in PLINK2 and samples were removed when they were significant outliers in the first six MDS dimensions or when there were no matching cases or controls on the MDS dimensions. The genotyped data were phased by SHAPEIT2 (Delaneau et al., 2013) and imputed by Minimac3 (Das et al., 2016) using the HRC release 1.1 (McCarthy et al., 2016) as the reference panel. GWAS was performed on SNPs with INFO score > 0.8 and MAF > 0.01, using a logistic regression model in PLINK2 with the first six and the 8th MDS PCA components as covariates. The GWAS result of this data set has not been previously published.

##### *UKBB | Breen, G. | United Kingdom | 30305743*

The UK Biobank sample consists of 776 OCD cases and 125,729 controls, see (Bycroft et al., 2018) for a general description of the UK Biobank resource. The data was derived from an online mental health questionnaire, completed by participants between July 2016 and July 2017. Research on the UK Biobank is conducted under a generic Research Tissue Bank approval from the UK North West Multi-centre Research Ethics Committee (MREC). This research was approved to be conducted under that approval by the governing Research Ethics Committee of the UK Biobank. The analyses in this paper were performed under an approved extension to project 16577. Cases were defined by self-report of a professional diagnosis of OCD ("Have you been diagnosed with [Obsessive compulsive disorder (OCD)] by a professional, even if you don't have it currently?"). The median age of cases at the time of report (not age at diagnosis) was 62 (interquartile range (IQR) = 55-68), 59% of cases are female. All participants who did not report an OCD diagnosis were included as controls. The median age of controls at the time of report was 65 (IQR = 58-70), 56% of controls are female. Genotyping of cases and controls was performed on the Affymetrix Axiom UK Biobank Array / Affymetrix Axiom UK BiLEVE array and genotyped in the Affymetrix Research Services Laboratory, Santa Clara, CA in several batches. For genotype quality control the following filters were applied to the data: MAF > 0.01 and call-rate > 98%. 4-means clustering was applied on the first two PCs to determine participants of European ancestries (as described by (Warren et al., 2017)). Further, quality assurance outliers marked by UKBB were removed. Relatives (KING relatedness > 0.044) greedily (e.g. keeping the parents in a parent-child trio) and participants with mismatched sex were removed (reported females with FX ≥ 0.6, reported males with FX ≤ 0.85; FX determined using chrX SNPs in approximate linkage equilibrium, r2 < 0.2). Imputation was performed with IMPUTE4 (Bycroft et al., 2018) using a combined HRC/UK-10k reference panel. Imputed data was filtered on MAF ≥

0.01 and INFO ≥ 0.4. GWAS was performed using SAIGE (Zhou et al., 2018), including the first six PCs from participants of European ancestries, factors for genotyping batch and assessment center as covariates.

##### *Yale-Penn | Gelernter, J.; CO-PI: Kranzler, H. | USA | 24166409, 32492095*

Yale-Penn data consists of three cohorts, called Yale-Penn 1 (also abbreviated GWCIDR), 2 (also abbreviated EXHCE), and 3 (also abbreviated MEGA123). Participants were recruited from eastern U.S sites and provided written informed consent as approved by the institutional review board at each site. Certificates of confidentiality were issued by the National Institute on Drug Abuse and the National Institute on Alcohol Abuse and Alcoholism. As previously described (Cheng et al., 2018), Yale-Penn 1 samples were genotyped using Illumina HumanOmni1-Quad v1.0 microarray, targeting approximately one million SNPs. Yale-Penn 2 samples were genotyped using Illumina HumanCore Exome array, including approximately 0.5 million SNPs. Yale-Penn 3 samples were genotyped using Illumina Multi-Ethnic Genotyping Array, including approximately 1.7 million SNPs. Genotyping was performed separately in each cohort using the 1000 Genomes Project reference panel, phase 1 (Durbin et al., 2010). First 10 PCs, calculated with EIGENSOFT (Price et al., 2006), were used to differentiate European and African ancestries through k-means clustering (Sherva et al., 2016; Solovieff et al., 2010). Imputation was performed with Minimac3 (Das et al., 2016), using the 1000 Genomes Project reference panel, phase 3 (The 1000 Genomes Project Consortium, 2015). For QC filtering, we kept SNPs with imputation quality scores > 0.6, HWE filtering $> 5x10^{-05}$, MAF > 0.05, missing call rates for variants < 0.5, and missing call rates for samples < 0.5. Imputation score was > 0.8. Data were aligned to the GRCh37 reference genome. Cases were defined according to the Diagnostic and Statistical Manual of mental disorders 4th edition (DSM-IV) by interviewers. As described for the MVP dataset, to optimize the number of cases to keep when removing individuals with kinship, which was calculated by KING (Manichaikul et al., 2010) for a minimum threshold of 0.0884 corresponding to a second-degree relationship, we implemented the following algorithm where cases have priority compared to controls: between a case and a control, we remove the control; if there is relatedness between two cases, we remove the one that has the highest number of relationships with other individuals; we do the same between two controls. In conclusion, we obtained 74 cases and 1635 controls for Yale-Penn 1, 52 cases and 1646 controls for Yale-Penn 2, and 53 cases and 1926 controls for Yale-Penn 3, making up a total of 179 cases and 5207 controls. GWAS was performed separately on each of the three cohorts using PLINK 2.0 (Chang et al., 2015) setting age, sex, and the first ten PCs as covariates.

#### Supplementary Note 3: Definition of independent significant loci

In total, the main meta-analysis contained 1672 SNPs with a p-value smaller than $5 \times 10^{-8}$. To identify independent genome-wide significant loci, we first performed automated LD-based ‘clumping’ of genome-wide SNPs using PLINK within the RICOPILI pipeline. A locus was defined by considering genomic regions harboring one or more genome-wide significant SNPs and testing for LD with neighboring SNPs in a 500-kb range. If a locus contained several genome-wide significant SNPs in LD ($r^2 > 0.1$), the SNP with the lowest p-value was selected as lead SNP. We then used GCTA-COJO to determine if a locus contained multiple independent SNPs (see also **Online Methods**). For this approach, we used the stepwise conditional regression approach to correct the betas and p-values of neighboring SNPs (in a sliding window of 10 Mb) based on the LD between the SNPs. We used the default p-value threshold of $5 \times 10^{-8}$ to define a genome-wide significant hit. The LD reference panel was created from 73,005 individuals from the QIMR Berghofer Medical Research Institute genetic epidemiology cohort. GCTA-COJO has the advantage that it can identify multiple independent signals within a single locus, which the standard PLINK clumping (as implemented into Ricopili) alone might miss.

### Supplementary Note 4: X-chromosome analysis in 23andMe

Results for the association between X-chromosome SNPs and OCD are only reported for a subset of our study samples (23andMe), as this information was only available for a limited number of samples. We are currently working in parallel on a manuscript that focuses on sex-stratified analyses and will include the full meta-analysis of all samples that have information for the X-chromosome available in that manuscript. For the 23andMe sample analysis of the X-chromosome was equivalent to the analysis of autosomes (see cohort description above), with the difference that for phasing 23andMe built separate haplotype graphs for the non-pseudoautosomal region and each pseudoautosomal region, and then phased these regions separately. Association tests were computed with male genotypes coded as if they were homozygous diploid for the observed allele. No SNP on the X-chromosome reached genome-wide significance.

### Supplementary Note 5: Subgroup, MTAG & common-factor analyses (GenomicSEM)

Heterogeneity was assessed with Cochran's Q statistic and $I^2$ statistic (Higgins et al., 2002; 2003). Cochran's Q is calculated as the weighted sum of squared differences between individual study effects and the pooled effect across studies, with the weights being those used in the pooling method. Q is distributed as a chi-square statistic with k (number of studies) minus 1 degrees of freedom. The $I^2$ statistic describes the percentage of variation across studies that is due to heterogeneity rather than chance. Unlike Q, it does not inherently depend upon the number of studies considered. As mentioned in the main article, we did not find evidence of significant heterogeneity in the results of our 30 genome-wide SNPs. Nevertheless, this does not rule out heterogeneity in study results at other loci or across the entire genome. Therefore, we conducted several analyses to explore whether the heterogeneity in sample ascertainment methods influenced genetic findings. In total, we found 39 independent loci that were associated with either our meta-analysis, a common factor GenomicSEM analysis, subgroup analyses, MTAG analyses, or more than one of the analyses mentioned (see further below for details). An overview of these results and loci can be found in **Supplementary Table 3**.

#### *Subgroup GWASs and meta analysis without 23andMe*

To investigate loci that are predominantly associated with the individual subgroups (clinical, comorbid, biobanks, 23andMe), we conducted separate GWASs for each subgroup, identifying seven SNPs associated with OCD in 23andMe (rs2836950 uniquely in this analysis; see **Supplementary Figure 37A and 37B** for Manhattan-plots and QQ-plots), and two in the clinical data (rs11856716 uniquely in this analysis; see **Supplementary Figure 34A and 34B** for Manhattan-plots and QQ-plots, and **Supplementary Table 3** for a list of all significant SNPs, as well as **Supplementary Figure 35A, 35B** and **36A and 36B** for Manhattan and QQ-plots of the comorbid and biobank specific GWASs). When conducting a combined GWAS of clinical, comorbid, and biobank cohorts (excluding 23andMe), two SNPs reached significance (**Supplementary Figure 33**). Genetic correlations ($rG$) between the four sub-groups ranged from 0.63 ($SE = 0.11, P = 2.1x10^{-09}$) between the comorbid and biobank sub-groups to 0.93 ($SE = 0.066, P = 9.89x10^{-45}$) between the comorbid and 23andMe sub-groups. The genetic correlation between all clinically diagnosed cases (clinical, comorbid and biobank) and the 23andme cases was 0.92 ($SE = 0.040, P = 2.58x10^{-113}$; See **Supplementary Table 7** for all correlation estimates).

#### *Multi-Trait Analysis of GWAS (MTAG)*

As the sample sizes for the different subgroups in our study varied, some of the subgroup analyses had limited power to detect subgroup-specific genetic variations. To address this, we conducted four multi-trait analyses to increase our power to detect these variations while still being able to identify subgroup-specific signals. MTAG is more commonly used to combine different but related traits into one meta-analysis by leveraging the shared heritability among the different traits and thereby gaining power. While we do not have different traits, our aim was to generate ascertainment-specific estimates, while boosting power by leveraging the high shared heritability among the subgroups. In addition to stronger evidence (meaning lower p-value) for several of the OCD hits defined in the main GWAS, 11 SNPs were associated with the biobank samples, of which one SNP (rs138354) was not found in any of the other analyses (including the meta-analysis, one-factor GWAS (see further below for details), subgroup-specific GWASs, or other MTAG analyses), 19 SNPs were associated with the clinical subgroup, of which one SNP (rs4838077) was not found in any of the other analyses, 24 SNPs were associated with the self-report subgroup, with one SNP (rs56346215) uniquely found in this analysis, and 20 SNPs associated with the comorbid subgroup (see **Supplementary Table 3** for a full list of results, and **Supplementary Figures S34-37** for Manhattan-plots).

##### *Genomic structural equation modeling (GenomicSEM)*

First, we ran a common-factor model without individual SNP effects, following the tutorial 'Models without individual SNP effects' on the GenomicSEM github website (Grotzinger et al., 2019, see web resources). In brief, the summary statistics were first harmonized and filtered (with the munge-function), using HapMap3 as the reference file, using the effective sample size (clinical: Neff=21,,562, comorbid: Neff=18,794, biobanks: Neff=36,124, 23andMe: Neff=116,876) as the input sample size, and filtering SNPs to INFO > 0.9 and MAF > 0.01. In a second step, multivariable LDSC was run to obtain the genetic covariance matrix and corresponding sampling covariance matrix, using precomputed European-ancestry LD scores, a sample prevalence of 0.5 and a population prevalence of 0.02. In a third step we ran a confirmatory factor analysis (CFA) using the pre-packaged common factor model in GenomicSEM using diagonally weighted least squares (DWLS) estimation. Second, we ran a multivariate GWAS of the common factor. In multivariate GWAS, the common factor defined by genetic indicators is regressed on each SNP, thereby generating summary statistics for the common factor (details can be found in the tutorial "GenomicSEM for Common Factor GWAS", see web resources). First, summary statistics for all four subgroups were prepared for multivariate GWAS with the 'sumstats' function in

GenomicSEM, which aligns and merges all files. Next, with the ‘userGWAS’ function, the S (genetic covariance) and V (corresponding sampling covariance) matrices from the LDSC output (from the model without SNP effects) and the summary statistics were combined to create a separate S and V matrix for each SNP containing the effect estimate. The function also transforms the effect estimates from the summary statistics and their SEs into covariances and SEs of covariances by taking the product of the regression coefficient and SNP variance from the reference file (1000 genomes phase 3). The CFA showed some evidence of sample ascertainment impacting our results at a genome-wide scale, however, not beyond what has been observed with closely related psychiatric illnesses (Wray et al., 2018; Strom et al., 2024) and yielded a satisfactory fit for a one-factor model $(Chisq = 3.46, df = 2, P_{chisq} = 0.18, CFI = 0.998, SRMR = 0.056)$, with high and similar loadings across all four subgroups (**Supplementary Figure 39** and **Supplementary Table 8**). Conducting a GWAS of this common factor model, we identified 20 genome-wide significant loci, all of which also reached significance in the main meta-analysis GWAS. Notably, the Manhattan-plot for the GenomicSEM GWAS closely resembles that of the main meta-analysis, though the common-factor GWAS shows reduced power to detect associated loci. This overall reduced power may partly stem from GenomicSEM’s exclusion of subgroup-specific unique variances. Additionally, GenomicSEM's modeling approach may overlook localized SNP-heritability deviations, which are preserved in a straightforward meta-analysis, allowing for greater power. While these unique variances of the subgroups may introduce heterogeneity, this heterogeneity may still be OCD-related, potentially capturing variations in OCD symptom profiles across samples (e.g., one sample may overrepresent checking symptoms, while another has a higher prevalence of intrusive thoughts or religious OCD cases). If such sample-specific variations align with distinct genetic factors, the unique variances in GenomicSEM could reflect genuine OCD-related variance, which a simple meta-analysis might retain, leading to stronger associations.

##### *Genetic correlations of subgroups with published GWASs*

We also conducted bivariate LDSC analyses for the 112 published GWASs (see Supplementary Note 5 for details) and our four subgroup GWASs to study potential heterogeneity between our subgroups. As expected, the comorbid subgroup showed stronger genetic correlations for the traits for which these studies were primarily ascertained (for AGDS and iPSYCH MDD, for iPSYCH alone also ADHD and ASD). All three traits (MDD, ASD, ADHD) in contrast were less strongly associated with the clinical and biobank cohorts, with the 23andMe sub-group showing estimates in between (**Supplementary Figure 40** and **Supplementary Table 19**).

#### Supplementary Note 6: Cross-trait genetic correlations

As mentioned in the main paper, we used bivariate LDSC to investigate the extent of genetic correlations between OCD and a curated list of 112 previously published GWASs of psychiatric, substance use, cognition/socioeconomic status (SES), personality, psychological, neurological, autoimmune, cardiovascular, anthropomorphic/diet, fertility, and other traits. We conducted the same analysis for the four OCD sub-groups (**Supplementary Figure 41** and **Supplementary Table 19**) as well as comparing the OCD GWAS of only 23andMe to the OCD GWAS excluding 23andMe (**Supplementary Figure 42** and **Supplementary Table 19)**. The co-morbid sub-group, as expected because the iPSYCH samples are enriched for MDD, ADHD, and ASD cases, tends to correlate higher with most psychiatric disorders they were primarily ascertained for, while clinical and biobank cohorts tend to correlate less strongly, with the 23andMe sub-group in between.

Beyond the strong genetic correlations of the main GWAS with psychiatric disorders described in the main text, high genetic correlations were also shown for several smoking phenotypes (positively with nicotine dependence, cigarettes per day, smoking initiation, and negatively with age smoking initiation), as well as a negative genetic correlation with alcohol dependence ($r_G = -0.1458,\ 95\%\,CI\,[-0.22, -0.29]$, $P_{adj} = 0.0001)$ was observed. We also found a negative correlation with educational attainment (EA, $r_G = -0.1015,\ 95\%\ CI\ [-0{,}14, -0.203],\ P_{adj} = 7.32x10^{-06}$) and other cognition traits, though there was some heterogeneity across the four subgroups with the biobank sub-cohort correlating positively with EA ($r_G = 0.1675$, $95\%\ CI\ [0.084{,}0.251]$, $P_{adj} = 0.0004$), while the 23andMe and co-morbid sub-groups correlated negatively (23andMe: $r_G = -0.1608,\ 95\%\ CI\ [-0.211, -0.111],\ P_{adj} = 7.61x10^{-10}$; Comorbid: $r_G = -0.1298,\ 95\%\ CI\ [-0.205, -0.054]$ , $P_{adj} = 0.0018$ ), and the clinical cohorts not showing a significant correlation ($r_G = -0.0412,\ 95\%\ CI\ [-0.041, 0.022],\ P_{adj} = 0.3191$). Moreover, some of the anthropomorphic traits (BMI, body fat, waist-hip-circumference, hip-circumference, and diet including fat) and auto-immune disorders (Crohn's disease, ulcerative colitis, and inflammatory bowel disease) showed a significant negative correlation with OCD, while asthma (adult onset) showed a positive correlation. In addition, significant correlations of substantial effect ($r_G > |0.25|$) were found with suicide attempt, tiredness, migraine (all positive), and with subjective well-being, childhood maltreatment, self-rated health, and risk taking-auto speeding (negative).

### Supplementary Note 7: Gene-based results.

#### *mBAT-combo*

A gene-based analysis was conducted using multivariate Set-Based Association Test (mBAT-combo) (Li et al., 2023) within GCTA version 1.94.1 (Yang et al., 2011). mBAT-combo has the advantage of being better powered than other gene-based association test methods to detect multi-SNP associations in the context of masking effects (i.e., when the product of the true SNP effect sizes and the LD correlation is negative). To ensure that the overall power is maximized independent of masking effects at specific loci, mBAT-combo combines mBAT and fastBAT test statistics through a Cauchy combination method, which allows the combination of different test statistics without a priori knowledge of the correlation structure. mBAT-combo identified 207 protein-coding genes significantly associated with OCD at a Bonferroni-corrected threshold of $P =$

$2.67x10^{-06}$ (**Supplementary Table 10**). Of these 104 were located on chromosome 6 in the extended major histocompatibility (MHC) locus (25MB - 35MB).

##### *Transcriptome-wide association study (TWAS)*

The transcriptome-wide association study (TWAS) using human prefrontal cortex gene expression weights generated by the psychENCODE consortium identified 24 significant protein-coding genes (Bonferroni-corrected $P = 4.76x10^{-06}$), of which 15 showed strong evidence for a co-localised signal (COLOC PP4 > 0.8; i.e., a single SNP associated with both OCD and gene expression implicating a causal role for this association) (**Supplementary Table 11**). Conditional analyses of each locus identified 14 genes with statistically independent TWAS signals, 11 of which also had evidence of a co-localized signal (COLOC PP4 > 0.8) (**Supplementary Table 11**).

##### *Summary-based Mendelian Randomization (SMR)*

We complemented the TWAS analysis with summary-based Mendelian Randomisation (SMR) using eQTL meta-analysis results from the eQTLGen (Võsa et al., 2021) (whole blood) and MetaBrain consortia (de Klein et al., 2023). A total of 46 unique protein-coding genes (21 of which located in the MHC region) reached significance (**Supplementary Table 12**), of which seven were significant in both the MetaBrain (14 genes in total) and eQTLGen (39 genes in total) datasets. Of the 46 genes, 24 (six in MHC region) had a HEIDI (heterogeneity in dependent instruments) P > 0.05 in blood or brain tissue, indicating that a single causal variant underlies the GWAS and eQTL association (**Supplementary Table 12**). We found three significant genes with HEIDI P > 0.05 in both the MetaBrain and eQTLGen datasets (*WDR6*, *ARIH2*, and *FLOT1*), one of which (WDR6) also had a significant and colocalized signal in the TWAS analysis.

##### *Psychiatric Omnilocus Prioritisation Score (PsyOPS)*

We used the Psychiatric Omnilocus Prioritisation Score (PsyOPS) (Wainberg et al., 2022) in a supervised approach that uses both positional mapping and biological annotations to prioritize putative causal genes for psychiatric traits. This approach expanded the list of candidate genes within OCD loci. For example, *CSRNP3* is nearest the index SNP rs9287859 on chromosome 2 and had a significant mBAT-combo association ($P = 3.99x10^{-08}$). However, PsyOPS prioritized the sodium channel protein subunit genes *SCN1A*, *SCN2A*, and *SCN3A*, which are located further away from the index SNP but are given greater weight by PsyOPS than other genes in the locus due to their loss-of-function intolerance, brain-specific expression, and involvement in neurodevelopmental disorders.

#### *TWAS-COLOC and SMR-HEIDI*

Colocalization of the significant TWAS results (TWAS-COLOC) identified 15 genes (PPH4 > 0.8), 11 of which were conditionally independent, while HEIDI prioritized 22 significant SMR associations (HEIDI P > 0.05). In total, 35 genes significantly associated with OCD passed colocalization and/or HEIDI criteria.

#### *Convergence across methods*

From the individual results for the four approaches, the following overlap in significant genes was observed: For the positional mapping approaches 123 genes showed significance in both the mBAT-combo and PsyOPS analyses. For the functional eQTL approaches, ten genes showed significance in both approaches (TWAS and SMR). For the group of 37 genes that was identified by at least one positional approach (i.e., either mBAT-combo or PsyOPS) and one functional eQTL approach (i.e., either TWAS or SMR), 27 genes were identified by mBAT-combo and either TWAS (four genes) or SMR (23 genes), and 27 genes were identified by PsyOPS and either TWAS (nine genes) or SMR (18 genes). See the main manuscript for the genes that were either conditional independent (TWAS-COLOC) or were significant in both tested SMR tissues (whole blood and brain). Out of the 123 genes identified in both positional mapping approaches (see above), 24 genes overlapped with significant results for the SMR analysis using eQTLs from whole blood, and eight genes overlapped with significant results for the SMR analysis using eQTLs from brain tissue. Out of these, five genes showed significance across the positional mapping approaches and SMR analyses for both tissues. These included the aforementioned *WDR6*, *ARIH2*, *NCKIPSD* genes from the previously identified chromosome 3 region, *BTN3A2* in the extended MHC region, and the Poly(A) Binding Protein Cytoplasmic 1 Like (*PABPC1L*) gene on chromosome 20. Out of the 123 genes identified in both positional mapping approaches (see above), 12 genes were also significant in the TWAS analysis. Of these, two genes were conditionally independent (*CTNND1*, and *WDR6*).

##### *Overlap of gene-based results with rare-coding variant results*

We performed a bi-directional look-up to assess the overlap of gene-findings from common-variance GWAS and rare-variance studies. We assessed 1) whether gene-findings from our GWAS showed evidence for rare variant involvement and 2) vice versa, whether findings from rare variant testing showed evidence of common variant association in our GWAS, using data from Halvorsen et al. (2021). We did not observe any evidence for differential case/control burden in the 251 GWAS-derived genes (**Supplementary Table 14**) for de novo synonymous variants ($p$ = 0.42), missense non-damaging variants ($p$ = 0.58) or missense damaging variants ($p$ = 0.58). We observed the lowest p-value for loss of function variants (p = 0.08), which was driven by two such variants in cases and none in controls. These de novo variants impact the genes *QRICH1* and *ZKSCAN3*. We note that *QRICH1* in particular is loss of function intolerant (pLI > 0.995) and was observed to carry a separate loss of function de novo variant in the largest de novo variant study published for Tourette Syndrome (Wang et al., 2018).

Next, we looked up 200 genes from Halvorsen et al. (2021) with a probability of loss of function intolerance > 0.995 (defined by Lek et al., 2016) and effect size estimate > 1. We found varying degrees of evidence for four different genes: *QRICH1*, *CTNND1*, *PTPRD*, *CHD3* (all look-ups can be found in **Supplementary Table 6**). Nevertheless, we found no evidence for unequal proportions of the 251 GWAS highlighted genes in the exome summary statistics (OR = 1.25, p = 0.76), most likely due to the exome study being severely underpowered (around 1300 total cases).

### Supplementary Note 8: Sample size and number of GWS loci

For OCD, like with other complex traits, there seems to be a relationship between the number of genome-wide significant loci that were identified in the GWAS and the sample size in the analysis. To highlight this relationship, we conducted additional meta-analyses combining different sub-samples of previously published GWASs (e.g., combining the PGC OCD2 GWAS with the 23andMe sample from this GWAS). A graphical representation can be found in **Supplementary Figure 43**. Between 25,000 (PGC OCD2 GWAS) and 30,000 cases (23andMe sample), the relationship between genome-wide significant loci and sample size seems to become linear, and (at least within European ancestry samples) each additional ~ 1000 cases will add one additional genome-wide significant SNP on average (see **Supplementary Table 20**).

### Supplementary Note 9: Assumptions and Limitations of SMR (and TWAS)

Transcriptome-wide association studies and SMR are commonly used for the identification and prioritization of target genes from GWAS of complex traits. A key advantage of these approaches is their use of summary-based GWAS and QTL datasets, meaning they can be applied across a range of experimental settings. In addition, TWAS and SMR only consider genetically regulated gene expression and are therefore not confounded by non-genetic factors that may influence expression (e.g., medication use). Nonetheless, the approaches are associated with several important assumptions and limitations (PMID: 30926968). For TWAS, the prediction accuracy of gene expression relies on the heritability of gene expression and therefore requires well-powered test SNP genotype and expression datasets for the development of expression weights. Meanwhile, SMR using a single causal variant is unable to distinguish between pleiotropy, where the same causal variant affects the trait and gene expression, and causality, where the effect of a causal variant on a trait is mediated by gene expression. Both approaches also assume the genotype is associated with gene expression and genes with a higher cis-genetic correlation to trait are more likely to be causal. In addition, they ignore long-range regulatory effects (trans-eQTLs) that may explain a large proportion of heritability, lack eQTL data from different ancestry groups, and cannot identify causal tissues of cell types. Despite these limitations, TWAS and SMR can identify important relationships between QTL datasets and GWAS to find target genes (PMID:36778001). We combined information from these approaches and other gene-based methods with different underlying assumptions (e.g., mBAT-combo or PsyOPS) to increase confidence in our gene-based associations and prioritize target genes within OCD loci.

#### Supplementary Note 10: A note on liability scale heritability estimates and sample ascertainment

When calculating liability scale heritability, both sample prevalence and population prevalence are important considerations. The sample prevalence is typically estimated from the number of cases and controls, but in biobank-scale studies, specialized analytical approaches (e.g., REGENIE) may obscure the true sample prevalence underlying the analysis. For population prevalence, robust estimates from entire populations are often used (e.g., from meta-analysis or national disease registers); however, ascertainment differences can complicate the use of precise population prevalences, as the specific fraction of the overall patient spectrum in individual cohorts may be unknown. Our study is limited by the use of a uniform prevalence estimate of 1% across all cohorts, which is on the lower end compared to previous studies and likely does not reflect the true prevalence across all enrolled cohorts. We adopted this approach due to the lack of reliable population prevalence data from prior meta-analyses or national registries, especially in Nordic countries, where available data is biased toward more severe and chronic cases based on hospital diagnoses. Consequently, liability scale heritability estimates from this manuscript should be interpreted with caution. Furthermore, when comparing estimates of (liability scale) heritability across studies, it is essential to consider the analytical approach and software that was used; comparisons should only be made when the same approaches are employed. For example, previous (higher) heritability estimates for OCD from GWASs studies (e.g., Mattheisen et al., 2015, or Stewart et al., 2013) used GCTA (Yang et al., 2011) for estimations while the current study uses LDSC. It is known that heritability estimates from these two (and other) approaches do not compare sufficiently (Evans et al., 2018).

### Supplementary Figures

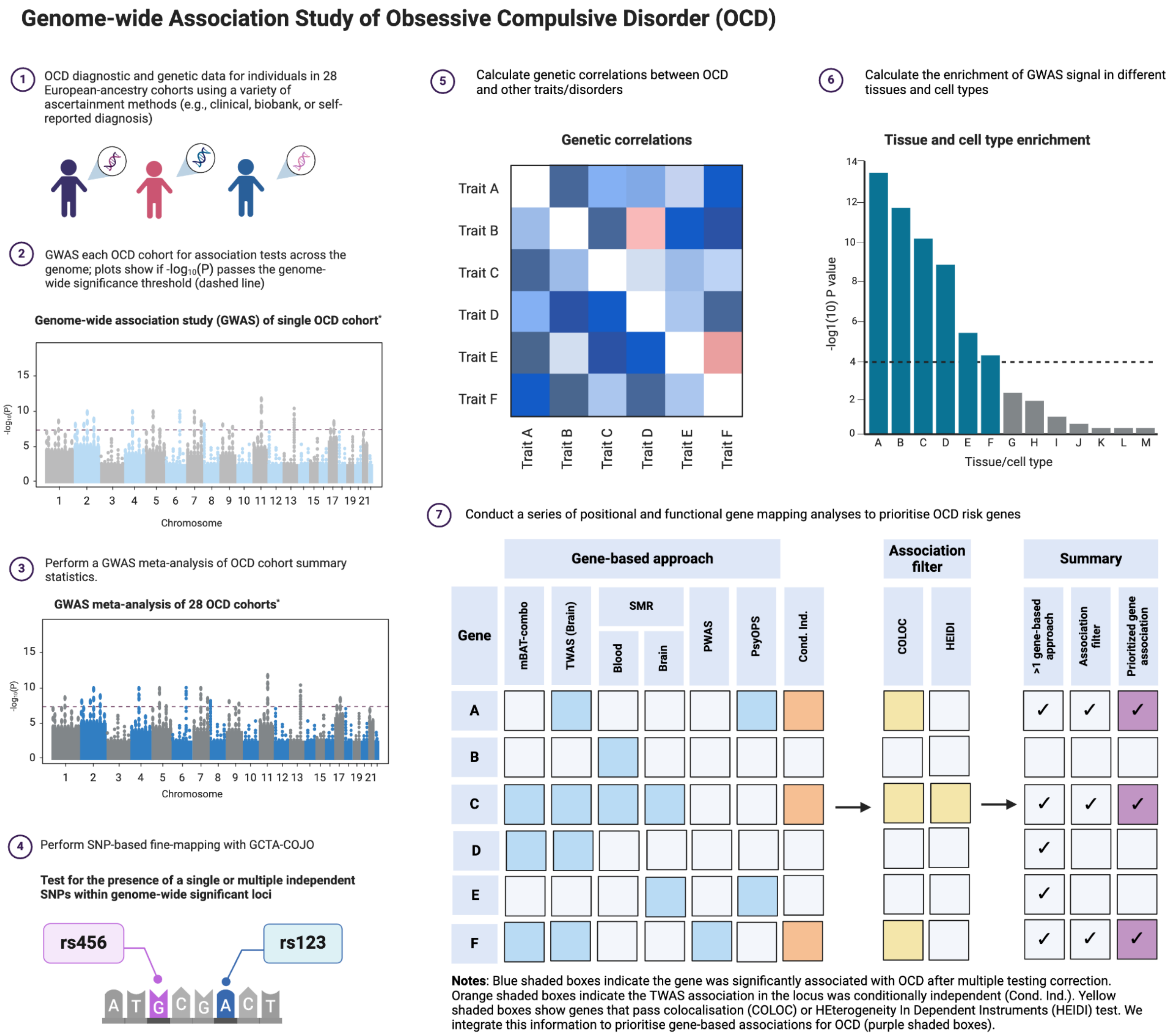

**Supplementary Figure 1: Schematic overview of the main bioinformatic analyses presented in the manuscript.** *Steps 1-4 outline the process for identifying SNP-based association signals*: 1) Collecting clinical and genetic data from OCD patients and controls across 28 different cohorts at various institutes; 2) Conducting individual GWAS for each OCD cohort; 3) Meta-analyzing the individual GWAS results to obtain combined summary statistics; 4) Characterizing the SNP findings through fine-mapping to determine whether each locus contains an independent signal. *Steps 5-7 detail additional analyses performed to further characterize the SNP signals identified in steps 1-4*, including: 5) Genetic correlation analysis between OCD and other traits/disorders; 6) Analyzing tissue and cell-type enrichment of the OCD-associated variants; and 7) A series of positional and functional gene mapping analyses to prioritize OCD risk genes. * GWAS Manhattan plots are not based on real data. Figure was created in BioRender

### Regional association plots and forest plots of the 30 significant SNPs

A

| rs78587207 | T/G | | 11:57654991 | | | |
|---|---|---|---|---|---|---|
| --?+---++---------??-?----?-- |  |  | het_P: | 0.46 | het_I: | 0.0 |
| | **info** | **p_value** | **f_ca(n)** | **f_co(n)** | **ln(OR)** | **STDerr** |
| dan_daner_23andMe. | 0.99 | 1.16e−06 | 0.688(30167) | 0.688(929804) | −0.0441 | 0.00906 |
| dan_daner_AGDS. | 0.96 | 0.619 | 0.679(757) | 0.684(5368) | −0.0301 | 0.0607 |
| dan_daner_Chop. | 1.00 | 0.991 | 0.701(406) | 0.701(1369) | 0.000963 | 0.0902 |
| dan_daner_Coga. | 1.00 | 0.545 | 0.695(131) | 0.695(1358) | −0.0872 | 0.144 |
| dan_daner_EGOS. | 0.94 | 0.853 | 0.678(926) | 0.676(1047) | −0.0143 | 0.0771 |
| dan_daner_EPOC. | 0.89 | 0.0374 | 0.652(195) | 0.727(204) | −0.336 | 0.163 |
| dan_daner_EstBB. | 0.99 | 0.431 | 0.637(772) | 0.637(196079) | 0.0422 | 0.0536 |
| dan_daner_FinnGen. | 1.00 | 0.806 | 0.641(1059) | 0.637(198110) | 0.0116 | 0.0472 |
| dan_daner_Hunt. | 0.99 | 0.218 | 0.652(284) | 0.677(66192) | −0.112 | 0.0914 |
| dan_daner_MVP. | 0.98 | 0.0942 | 0.686(5129) | 0.686(422860) | −0.0362 | 0.0216 |
| dan_daner_MoBa. | 0.98 | 0.852 | 0.659(104) | 0.664(2193) | −0.0277 | 0.149 |
| dan_daner_NORDiC−NOR. | 1.02 | 0.0591 | 0.646(599) | 0.693(619) | −0.175 | 0.0926 |
| dan_daner_NORDiC−SWE. | 0.95 | 0.282 | 0.656(1691) | 0.671(1323) | −0.0622 | 0.0578 |
| dan_daner_Psych_Broad. | 1.00 | 0.561 | 0.688(1396) | 0.686(4009) | −0.0282 | 0.0484 |
| dan_daner_UKBB. | 1.00 | 0.0051 | 0.685(776) | 0.685(125729) | −0.147 | 0.0525 |
| dan_daner_Wue. | 0.94 | 0.0924 | 0.637(140) | 0.685(526) | −0.247 | 0.147 |
| dan_daner_YalePenn_GWCIDR. | 1.05 | 0.749 | 0.663(74) | 0.663(1635) | −0.0595 | 0.186 |
| dan_daner_YalePenn_MEGA123. | 1.04 | 0.594 | 0.683(53) | 0.683(1926) | −0.114 | 0.215 |
| dan_daner_iPSYCH. | 1.00 | 0.000723 | 0.657(4509) | 0.675(38392) | −0.0797 | 0.0236 |
| dan_daner_iocd. | 1.01 | 0.212 | 0.690(1519) | 0.694(3541) | −0.0601 | 0.0482 |
| dan_daner_iocdf_trio. | 0.93 | 0.344 | 0.672(323) | 0.696(323) | −0.118 | 0.124 |
| dan_daner_ocgas. | 1.03 | 0.0649 | 0.680(986) | 0.709(1023) | −0.125 | 0.0678 |
| dan_daner_sickkids. | 1.00 | 0.115 | 0.692(275) | 0.692(205) | −0.292 | 0.185 |
| **Meta** | **0.99** | **5.28e−12** | **0.681(53660)** | **0.677(2044417)** | **−0.049** | **0.0071** |

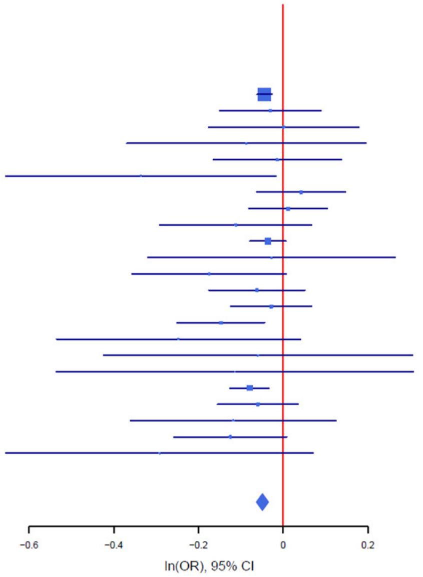

B

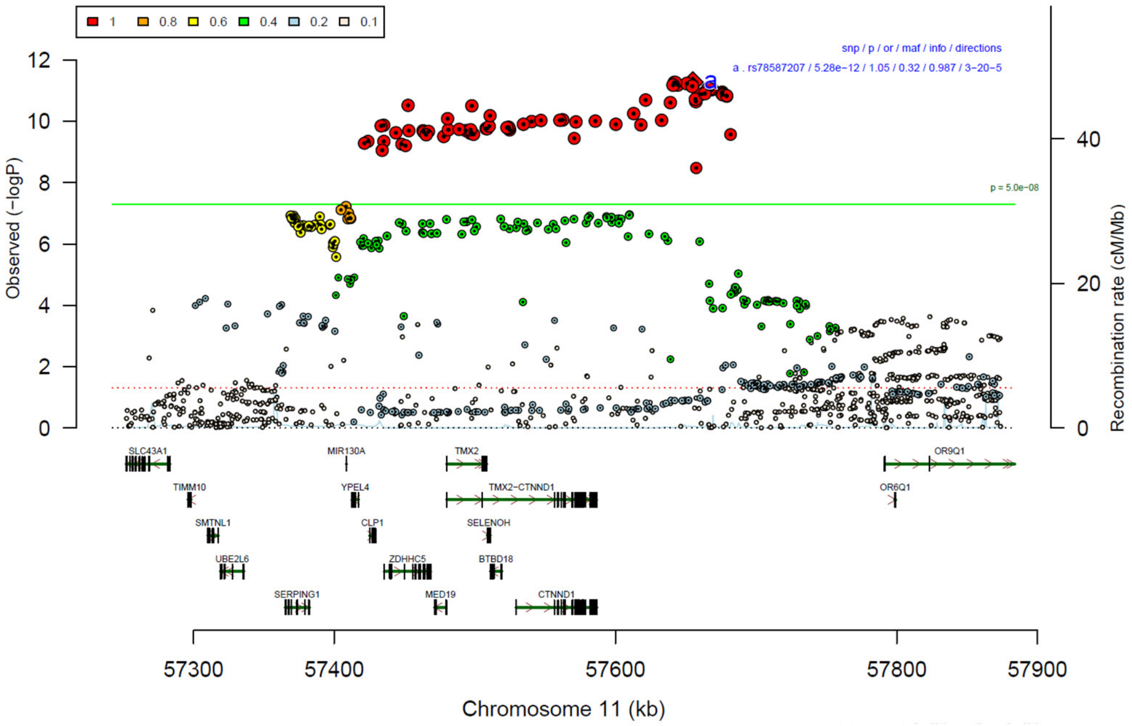

**Supplementary Figure 2: Forest plot (A) and regional association plot (B) of SNP rs78587207 in the meta-analysis.** The forest plot (A) shows the imputation quality (INFO) score, p-value of the SNP association (two-sided and not corrected for multiple testing, derived from a fixed-effects inverse-variance weighted meta-analysis), allele frequency in cases with case sample size (f_ca(n)), allele frequency in controls and control sample size (f_co(n)), beta estimates (ln(OR)) and standard error (STDerr) for each study as well as for the combined meta-analysis. Each cohort's sample size is detailed in supplementary table 1. Data on the right show the ln(OR) value as listed per cohort with 95% confidence intervals. In the regional association plot (B), the –log10 p-values (from two-sided association tests) are shown on the left y-axis. Recombination rates expressed in centimorgans (cM) per Mb (Megabase) (blue line) are shown on the right y-axis. Position in Mb is on the x-axis, with genes shown below the regional association plot. Only SNPs with an association p-value less than 0.1 were plotted. The most associated SNP is shown as a diamond and marked with the letter a.

A

| rs13262595 | G/A | | 8:143316970 | | | |
|---|---|---|---|---|---|---|
| ++++-++++-+++-+-+-++-+-++?-+ | | | het_P: | 0.0819 | het_I: | 28.9 |
| | info | p_value | f_ca(n) | f_co(n) | ln(OR) | STDerr |
| dan_daner_23andMe. | 1.00 | 0.000187 | 0.555(30167) | 0.555(929804) | −0.0313 | 0.00838 |
| dan_daner_AGDS. | 0.99 | 0.338 | 0.555(757) | 0.567(5368) | −0.0538 | 0.0561 |
| dan_daner_Chop. | 1.00 | 0.805 | 0.570(406) | 0.570(1369) | −0.0202 | 0.0817 |
| dan_daner_Coga. | 1.00 | 0.5 | 0.549(131) | 0.549(1358) | 0.0904 | 0.134 |
| dan_daner_EGOS. | 0.98 | 0.0192 | 0.538(926) | 0.585(1047) | −0.167 | 0.0714 |
| dan_daner_EPOC. | 0.99 | 0.858 | 0.577(195) | 0.576(204) | −0.0253 | 0.142 |
| dan_daner_EstBB. | 0.98 | 0.884 | 0.583(772) | 0.583(196079) | −0.0076 | 0.0522 |
| dan_daner_FinnGen. | 1.00 | 0.0689 | 0.572(1059) | 0.598(198110) | −0.0837 | 0.046 |
| dan_daner_Hunt. | 0.99 | 0.77 | 0.545(284) | 0.540(66192) | 0.0248 | 0.0847 |
| dan_daner_MVP. | 0.96 | 0.0445 | 0.550(5129) | 0.550(422860) | −0.0409 | 0.0204 |
| dan_daner_MoBa. | 0.98 | 0.265 | 0.601(104) | 0.563(2193) | 0.164 | 0.147 |
| dan_daner_NORDiC−NOR. | 0.95 | 0.984 | 0.548(599) | 0.556(619) | 0.0018 | 0.0904 |
| dan_daner_NORDiC−SWE. | 1.02 | 0.0613 | 0.559(1691) | 0.582(1323) | −0.0996 | 0.0532 |
| dan_daner_Psych_Broad. | 0.98 | 0.0247 | 0.523(1396) | 0.542(4009) | −0.102 | 0.0456 |
| dan_daner_UKBB. | 1.00 | 0.727 | 0.566(776) | 0.566(125729) | −0.0181 | 0.0519 |
| dan_daner_Wue. | 0.94 | 0.625 | 0.546(140) | 0.560(526) | −0.0688 | 0.141 |
| dan_daner_YalePenn_GWCIDR. | 0.99 | 0.352 | 0.538(74) | 0.538(1635) | 0.165 | 0.177 |
| dan_daner_YalePenn_MEGA123. | 1.02 | 0.287 | 0.559(53) | 0.559(1926) | −0.214 | 0.201 |
| dan_daner_bioVU. | 1.00 | 0.478 | 0.543(1014) | 0.552(38613) | −0.0324 | 0.0457 |
| dan_daner_iPSYCH. | 0.99 | 0.000314 | 0.543(4509) | 0.562(38392) | −0.081 | 0.0225 |
| dan_daner_iocd. | 1.01 | 0.000164 | 0.534(1519) | 0.576(3541) | −0.169 | 0.0448 |
| dan_daner_iocdf_trio. | 1.04 | 0.0626 | 0.520(323) | 0.573(323) | −0.205 | 0.11 |
| dan_daner_ocgas−ab. | 0.90 | 0.618 | 0.581(55) | 0.534(55) | 0.172 | 0.345 |
| dan_daner_ocgas−gh. | 1.17 | 0.6 | 0.501(56) | 0.554(56) | −0.135 | 0.257 |
| dan_daner_ocgas−nes. | 1.04 | 0.307 | 0.552(212) | 0.522(212) | 0.14 | 0.137 |
| dan_daner_ocgas. | 1.03 | 0.00314 | 0.518(986) | 0.566(1023) | −0.185 | 0.0628 |
| dan_daner_sickkids. | 1.00 | 0.52 | 0.561(275) | 0.561(205) | 0.108 | 0.168 |
| **Meta** | **0.99** | **1.31e−11** | **0.552(53660)** | **0.561(2044417)** | **−0.0444** | **0.0066** |

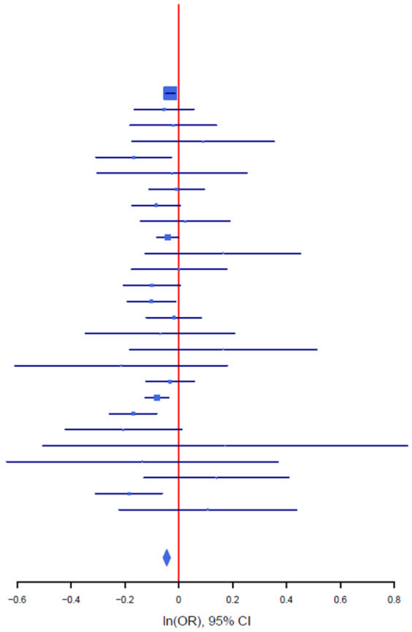

B

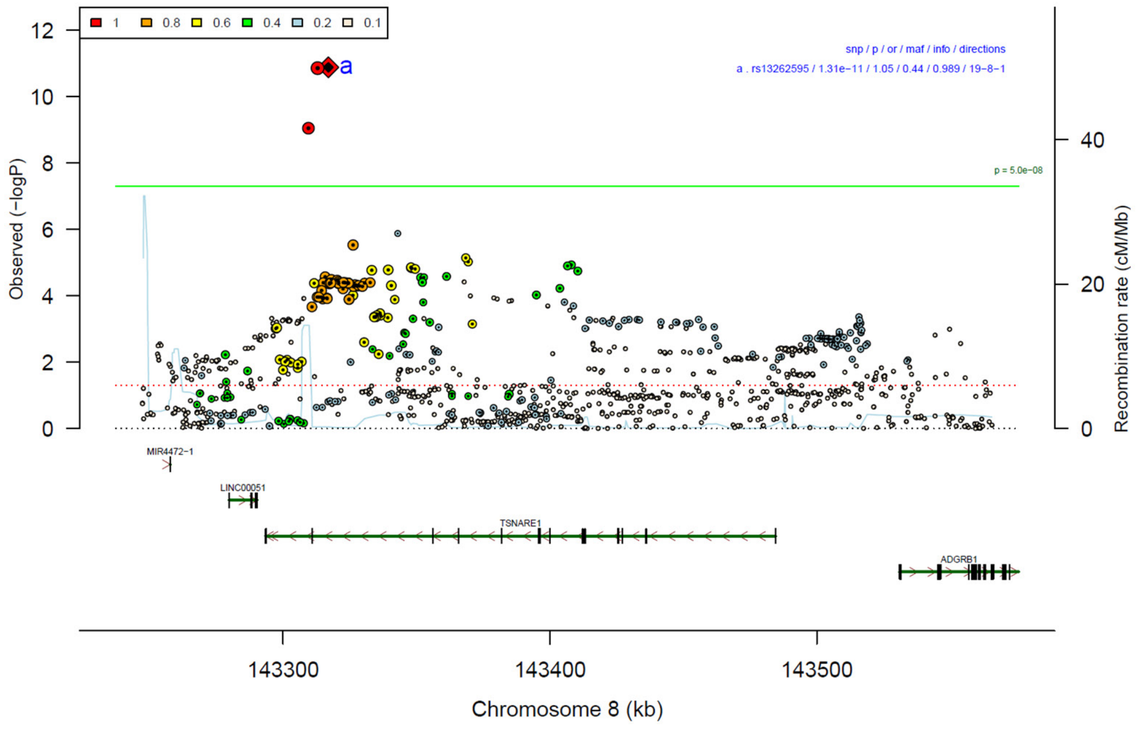

**Supplementary Figure 3: Forest plot (A) and regional association plot (B) of SNP rs13262595 in the meta-analysis.** The forest plot (A) shows the imputation quality (INFO) score, p-value of the SNP association (two-sided and not corrected for multiple testing, derived from a fixed-effects inverse-variance weighted meta-analysis), allele frequency in cases with case sample size (f_ca(n)), allele frequency in controls and control sample size (f_co(n)), beta estimates (ln(OR)) and standard error (STDerr) for each study as well as for the combined meta-analysis. Each cohort's sample size is detailed in supplementary table 1. Data on the right show the ln(OR) value as listed per cohort with 95% confidence intervals. In the regional association plot (B), the –log10 p-values (from two-sided association tests) are shown on the left y-axis. Recombination rates expressed in centimorgans (cM) per Mb (Megabase) (blue line) are shown on the right y-axis. Position in Mb is on the x-axis, with genes shown below the regional association plot. Only SNPs with an association p-value less than 0.1 were plotted. The most associated SNP is shown as a diamond and marked with the letter a.

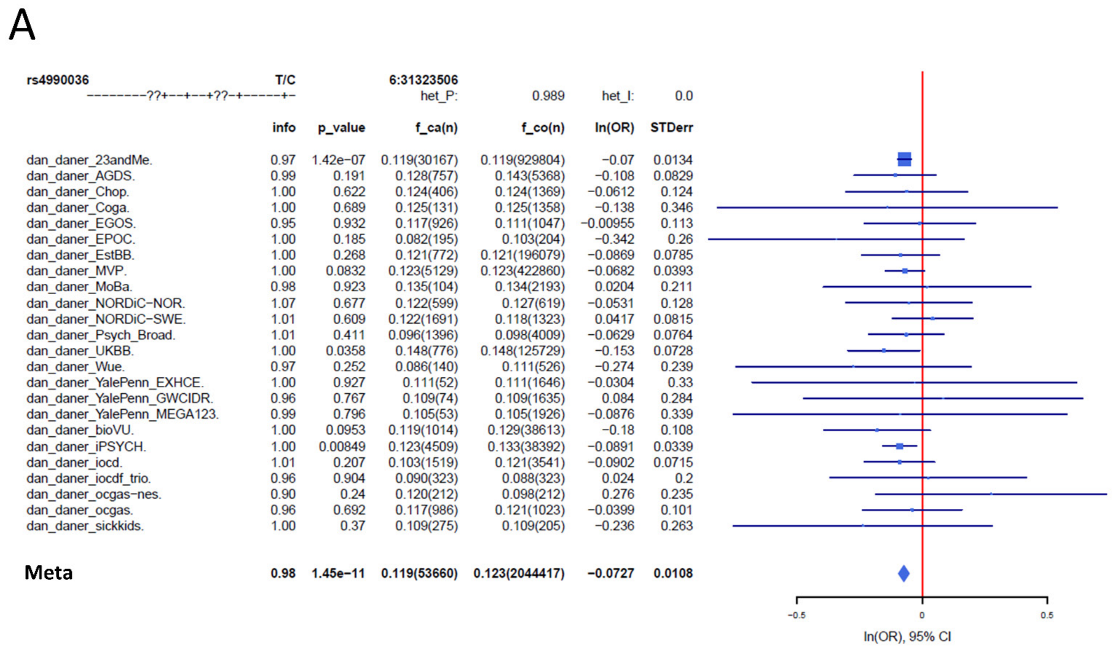

| | info | p_value | f_ca(n) | f_co(n) | ln(OR) | STDerr |
|---|---|---|---|---|---|---|
| dan_daner_23andMe. | 0.97 | 1.42e−07 | 0.119(30167) | 0.119(929804) | −0.07 | 0.0134 |
| dan_daner_AGDS. | 0.99 | 0.191 | 0.128(757) | 0.143(5368) | −0.108 | 0.0829 |
| dan_daner_Chop. | 1.00 | 0.622 | 0.124(406) | 0.124(1369) | −0.0612 | 0.124 |
| dan_daner_Coga. | 1.00 | 0.689 | 0.125(131) | 0.125(1358) | −0.138 | 0.346 |
| dan_daner_EGOS. | 0.95 | 0.932 | 0.117(926) | 0.111(1047) | −0.00955 | 0.113 |
| dan_daner_EPOC. | 1.00 | 0.185 | 0.082(195) | 0.103(204) | −0.342 | 0.26 |
| dan_daner_EstBB. | 1.00 | 0.268 | 0.121(772) | 0.121(196079) | −0.0869 | 0.0785 |
| dan_daner_MVP. | 1.00 | 0.0832 | 0.123(5129) | 0.123(422860) | −0.0682 | 0.0393 |
| dan_daner_MoBa. | 0.98 | 0.923 | 0.135(104) | 0.134(2193) | 0.0204 | 0.211 |
| dan_daner_NORDiC−NOR. | 1.07 | 0.677 | 0.122(599) | 0.127(619) | −0.0531 | 0.128 |
| dan_daner_NORDiC−SWE. | 1.01 | 0.609 | 0.122(1691) | 0.118(1323) | 0.0417 | 0.0815 |
| dan_daner_Psych_Broad. | 1.01 | 0.411 | 0.096(1396) | 0.098(4009) | −0.0629 | 0.0764 |
| dan_daner_UKBB. | 1.00 | 0.0358 | 0.148(776) | 0.148(125729) | −0.153 | 0.0728 |
| dan_daner_Wue. | 0.97 | 0.252 | 0.086(140) | 0.111(526) | −0.274 | 0.239 |
| dan_daner_YalePenn_EXHCE. | 1.00 | 0.927 | 0.111(52) | 0.111(1646) | −0.0304 | 0.33 |
| dan_daner_YalePenn_GWCIDR. | 0.96 | 0.767 | 0.109(74) | 0.109(1635) | 0.084 | 0.284 |
| dan_daner_YalePenn_MEGA123. | 0.99 | 0.796 | 0.105(53) | 0.105(1926) | −0.0876 | 0.339 |
| dan_daner_bioVU. | 1.00 | 0.0953 | 0.119(1014) | 0.129(38613) | −0.18 | 0.108 |
| dan_daner_iPSYCH. | 1.00 | 0.00849 | 0.123(4509) | 0.133(38392) | −0.0891 | 0.0339 |
| dan_daner_iocd. | 1.01 | 0.207 | 0.103(1519) | 0.121(3541) | −0.0902 | 0.0715 |
| dan_daner_iocdf_trio. | 0.96 | 0.904 | 0.090(323) | 0.088(323) | 0.024 | 0.2 |
| dan_daner_ocgas−nes. | 0.90 | 0.24 | 0.120(212) | 0.098(212) | 0.276 | 0.235 |
| dan_daner_ocgas. | 0.96 | 0.692 | 0.117(986) | 0.121(1023) | −0.0399 | 0.101 |
| dan_daner_sickkids. | 1.00 | 0.37 | 0.109(275) | 0.109(205) | −0.236 | 0.263 |
| **Meta** | **0.98** | **1.45e−11** | **0.119(53660)** | **0.123(2044417)** | **−0.0727** | **0.0108** |

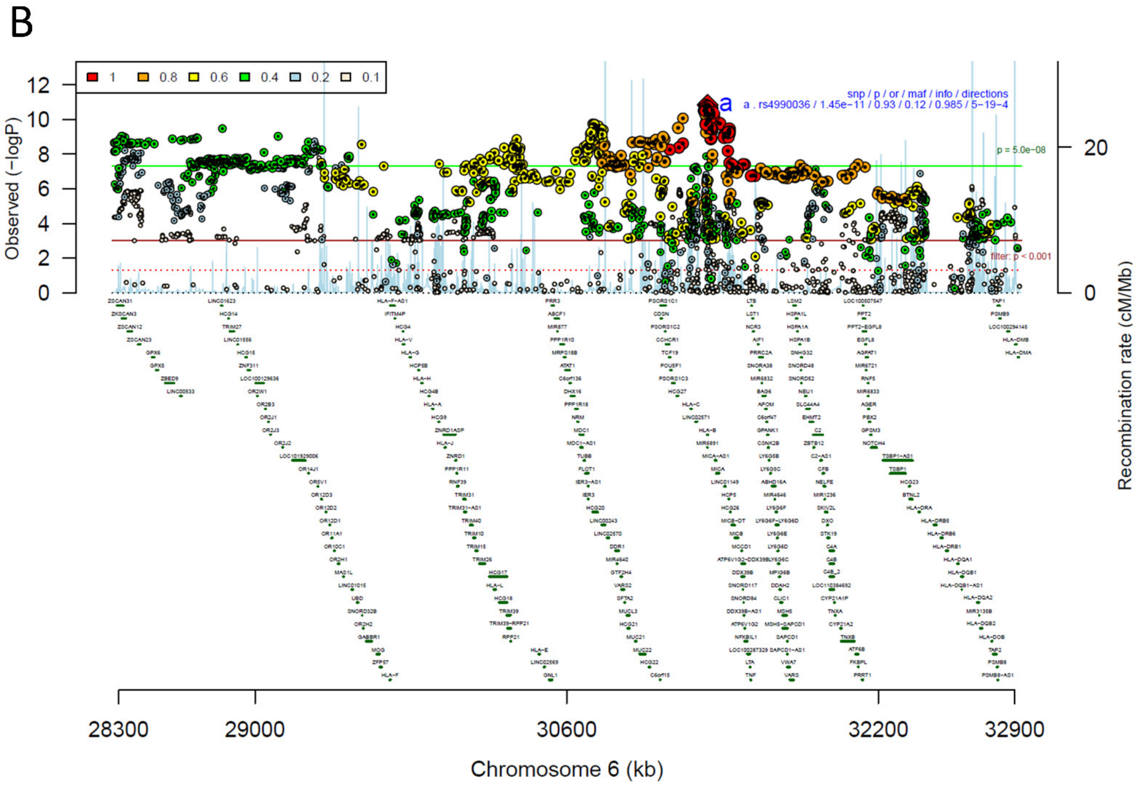

**Supplementary Figure 4: Forest plot (A) and regional association plot (B) of SNP rs4990036 in the meta-analysis:** The forest plot (A) shows the imputation quality (INFO) score, p-value of the SNP association (two-sided and not corrected for multiple testing, derived from a fixed-effects inverse-variance weighted meta-analysis), allele frequency in cases with case sample size (f_ca(n)), allele frequency in controls and control sample size (f_co(n)), beta estimates (ln(OR)) and standard error (STDerr) for each study as well as for the combined meta-analysis. Each cohort's sample size is detailed in supplementary table 1. Data on the right show the ln(OR) value as listed per cohort with 95% confidence intervals. In the regional association plot (B), the –log10 p-values (from two-sided association tests) are shown on the left y-axis. Recombination rates expressed in centimorgans (cM) per Mb (Megabase) (blue line) are shown on the right y-axis. Position in Mb is on the x-axis, with genes shown below the regional association plot. Only SNPs with an association p-value less than 0.1 were plotted. The most associated SNP is shown as a diamond and marked with the letter a.

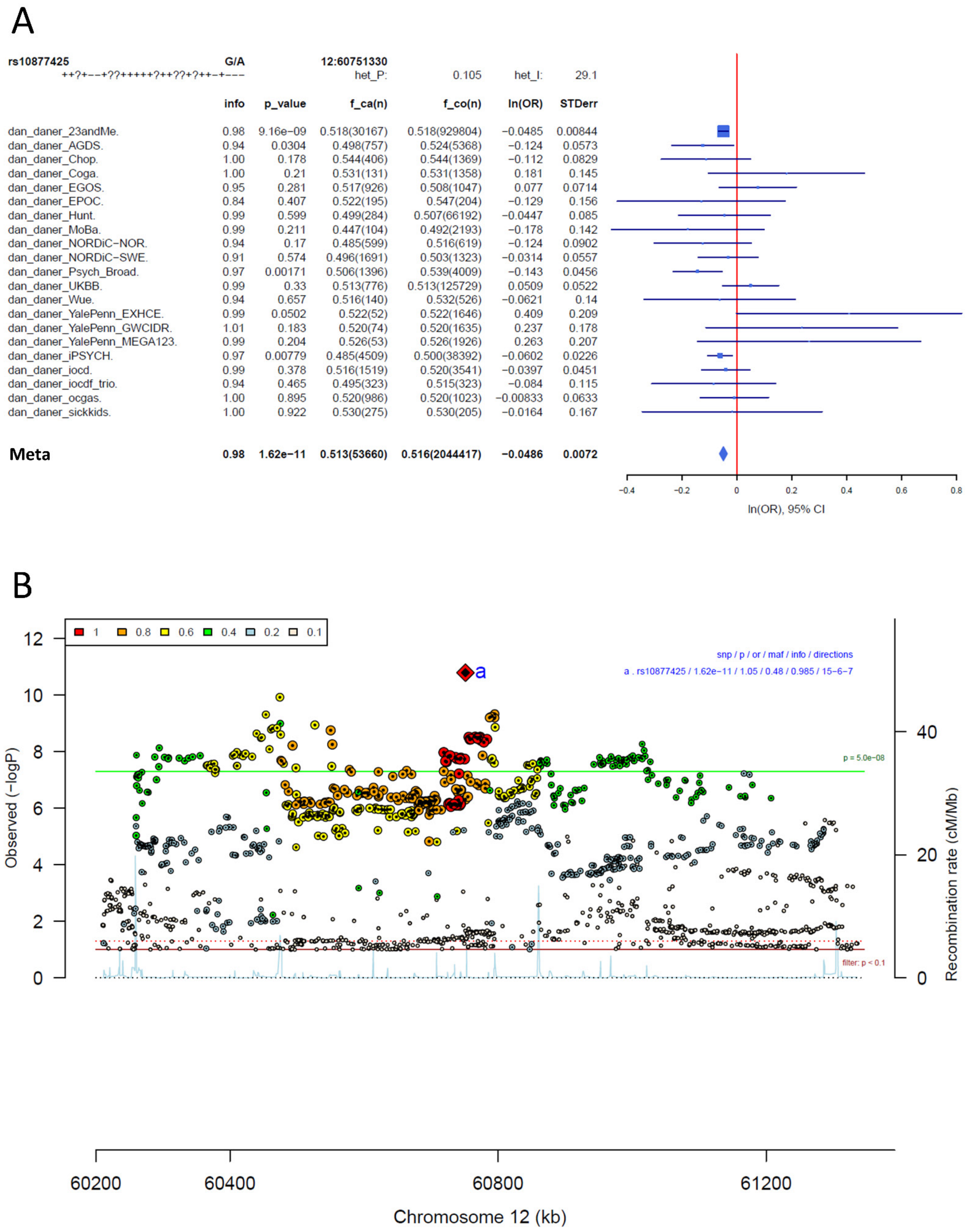

**Supplementary Figure 5: Forest plot (A) and regional association plot (B) of SNP rs10877425 in the meta-analysis.** The forest plot (A) shows the imputation quality (INFO) score, p-value of the SNP association (two-sided and not corrected for multiple testing, derived from a fixed-effects inverse-variance weighted meta-analysis), allele frequency in cases with case sample size (f_ca(n)), allele frequency in controls and control sample size (f_co(n)), beta estimates (ln(OR)) and standard error (STDerr) for each study as well as for the combined meta-analysis. Each cohort's sample size is detailed in supplementary table 1. Data on the right show the ln(OR) value as listed per cohort with 95% confidence intervals. In the regional association plot (B), the –log10 p-values (from two-sided association tests) are shown on the left y-axis. Recombination rates expressed in centimorgans (cM) per Mb (Megabase) (blue line) are shown on the right y-axis. Position in Mb is on the x-axis, with genes shown below the regional association plot. Only SNPs with an association p-value less than 0.1 were plotted. The most associated SNP is shown as a diamond and marked with the letter a.

A

rs7626445 T/C 3:49114544
----?+++-+-------??+?----+-- het_P: 0.958 het_I: 0.0

| | info | p_value | f_ca(n) | f_co(n) | ln(OR) | STDerr |
|---|---|---|---|---|---|---|
| dan_daner_23andMe. | 1.00 | 1.25e−08 | 0.653(30167) | 0.653(929804) | −0.0499 | 0.00874 |
| dan_daner_AGDS. | 0.98 | 0.276 | 0.642(757) | 0.655(5368) | −0.0637 | 0.0585 |
| dan_daner_Chop. | 1.00 | 0.97 | 0.650(406) | 0.650(1369) | −0.00316 | 0.0853 |
| dan_daner_EGOS. | 0.97 | 0.79 | 0.630(926) | 0.641(1047) | 0.0196 | 0.0737 |
| dan_daner_EPOC. | 0.95 | 0.673 | 0.648(195) | 0.639(204) | 0.0645 | 0.153 |
| dan_daner_EstBB. | 0.99 | 0.781 | 0.663(772) | 0.663(196079) | 0.0151 | 0.0544 |
| dan_daner_FinnGen. | 1.00 | 0.862 | 0.666(1059) | 0.666(198110) | −0.0083 | 0.0477 |
| dan_daner_Hunt. | 1.00 | 0.873 | 0.658(284) | 0.651(66192) | 0.0143 | 0.089 |
| dan_daner_MVP. | 0.98 | 0.171 | 0.646(5129) | 0.646(422860) | −0.0286 | 0.0209 |
| dan_daner_MoBa. | 1.00 | 0.425 | 0.630(104) | 0.659(2193) | −0.122 | 0.153 |
| dan_daner_NORDiC−NOR. | 1.04 | 0.216 | 0.637(599) | 0.660(619) | −0.111 | 0.09 |
| dan_daner_NORDiC−SWE. | 1.02 | 0.756 | 0.628(1691) | 0.634(1323) | −0.0169 | 0.0544 |
| dan_daner_Psych_Broad. | 1.02 | 0.0919 | 0.640(1396) | 0.653(4009) | −0.0783 | 0.0465 |
| dan_daner_UKBB. | 1.00 | 0.0902 | 0.658(776) | 0.658(125729) | −0.0927 | 0.0547 |
| dan_daner_Wue. | 0.98 | 0.734 | 0.616(140) | 0.625(526) | −0.0476 | 0.14 |
| dan_daner_YalePenn_EXHCE. | 1.02 | 0.845 | 0.652(52) | 0.652(1646) | 0.0432 | 0.221 |
| dan_daner_YalePenn_GWCIDR. | 1.00 | 0.555 | 0.646(74) | 0.646(1635) | −0.109 | 0.185 |
| dan_daner_YalePenn_MEGA123. | 1.00 | 0.19 | 0.665(53) | 0.665(1926) | −0.271 | 0.207 |
| dan_daner_bioVU. | 1.00 | 0.155 | 0.632(1014) | 0.647(38613) | −0.068 | 0.0478 |
| dan_daner_iPSYCH. | 0.98 | 0.00289 | 0.621(4509) | 0.637(38392) | −0.0691 | 0.0232 |
| dan_daner_iocd. | 0.98 | 0.892 | 0.639(1519) | 0.650(3541) | −0.00638 | 0.047 |
| dan_daner_iocdf_trio. | 0.97 | 0.752 | 0.650(323) | 0.658(323) | −0.0375 | 0.119 |
| dan_daner_ocgas. | 1.01 | 0.443 | 0.636(986) | 0.622(1023) | 0.0501 | 0.0655 |
| dan_daner_sickkids. | 1.00 | 0.662 | 0.646(275) | 0.646(205) | −0.0773 | 0.177 |
| **Meta** | **0.99** | **1.74e−11** | **0.647(53660)** | **0.654(2044417)** | **−0.0459** | **0.0068** |

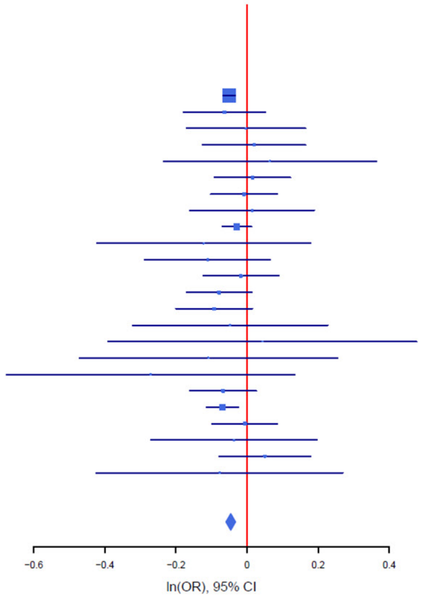

B

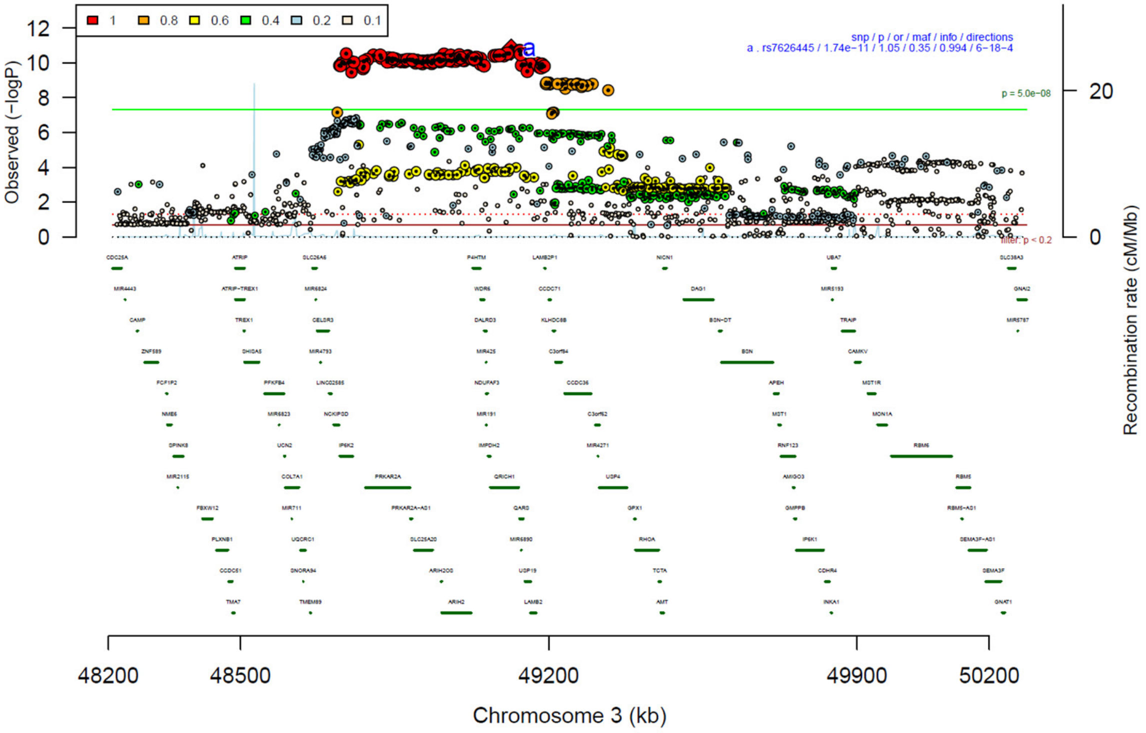

**Supplementary Figure 6: Forest plot (A) and regional association plot (B) of SNP rs7626445 in the meta-analysis.** The forest plot (A) shows the imputation quality (INFO) score, p-value of the SNP association (two-sided and not corrected for multiple testing, derived from a fixed-effects inverse-variance weighted meta-analysis), allele frequency in cases with case sample size (f_ca(n)), allele frequency in controls and control sample size (f_co(n)), beta estimates (ln(OR)) and standard error (STDerr) for each study as well as for the combined meta-analysis. Each cohort's sample size is detailed in supplementary table 1. Data on the right show the ln(OR) value as listed per cohort with 95% confidence intervals. In the regional association plot (B), the –log10 p-values (from two-sided association tests) are shown on the left y-axis. Recombination rates expressed in centimorgans (cM) per Mb (Megabase) (blue line) are shown on the right y-axis. Position in Mb is on the x-axis, with genes shown below the regional association plot. Only SNPs with an association p-value less than 0.1 were plotted. The most associated SNP is shown as a diamond and marked with the letter a.

A

rs2564930 T/C 3:53017307

--?+-----+---+-++??-?------- het_P: 0.111 het_I: 27.0

| | info | p_value | f_ca(n) | f_co(n) | ln(OR) | STDerr |
|---|---|---|---|---|---|---|
| dan_daner_23andMe. | 1.00 | 2.33e−06 | 0.341(30167) | 0.341(929804) | −0.0419 | 0.00889 |
| dan_daner_AGDS. | 0.98 | 0.968 | 0.342(757) | 0.341(5368) | −0.0023 | 0.0593 |
| dan_daner_Chop. | 1.00 | 0.158 | 0.352(406) | 0.352(1369) | 0.12 | 0.0851 |
| dan_daner_Coga. | 1.00 | 0.841 | 0.329(131) | 0.329(1358) | −0.0288 | 0.143 |
| dan_daner_EGOS. | 0.98 | 0.264 | 0.320(926) | 0.325(1047) | −0.0844 | 0.0755 |
| dan_daner_EPOC. | 0.95 | 0.427 | 0.352(195) | 0.381(204) | −0.125 | 0.158 |
| dan_daner_EstBB. | 0.99 | 0.758 | 0.377(772) | 0.377(196079) | −0.0163 | 0.0531 |
| dan_daner_FinnGen. | 1.00 | 0.113 | 0.298(1059) | 0.324(198110) | −0.0764 | 0.0482 |
| dan_daner_Hunt. | 1.00 | 0.235 | 0.349(284) | 0.323(66192) | 0.107 | 0.0901 |
| dan_daner_MVP. | 0.96 | 0.153 | 0.354(5129) | 0.354(422860) | −0.0305 | 0.0214 |
| dan_daner_MoBa. | 1.00 | 0.522 | 0.332(104) | 0.311(2193) | 0.1 | 0.156 |
| dan_daner_NORDiC−NOR. | 1.01 | 0.691 | 0.322(599) | 0.329(619) | 0.0368 | 0.0927 |
| dan_daner_NORDiC−SWE. | 1.02 | 0.358 | 0.322(1691) | 0.311(1323) | 0.052 | 0.0566 |
| dan_daner_Psych_Broad. | 0.99 | 0.00454 | 0.350(1396) | 0.387(4009) | −0.134 | 0.047 |
| dan_daner_UKBB. | 1.00 | 0.037 | 0.339(776) | 0.339(125729) | −0.111 | 0.0533 |
| dan_daner_Wue. | 0.97 | 0.102 | 0.295(140) | 0.345(526) | −0.245 | 0.15 |
| dan_daner_YalePenn_EXHCE. | 1.01 | 0.902 | 0.353(52) | 0.353(1646) | −0.0265 | 0.215 |
| dan_daner_YalePenn_GWCIDR. | 1.04 | 0.55 | 0.354(74) | 0.354(1635) | −0.11 | 0.184 |
| dan_daner_YalePenn_MEGA123. | 0.99 | 0.563 | 0.355(53) | 0.355(1926) | −0.123 | 0.213 |
| dan_daner_iPSYCH. | 0.99 | 0.0129 | 0.329(4509) | 0.343(38392) | −0.0593 | 0.0238 |
| dan_daner_iocd. | 0.99 | 0.00407 | 0.314(1519) | 0.345(3541) | −0.138 | 0.0481 |
| dan_daner_iocdf_trio. | 1.00 | 0.113 | 0.321(323) | 0.363(323) | −0.187 | 0.118 |
| dan_daner_ocgas. | 1.01 | 0.00347 | 0.322(986) | 0.366(1023) | −0.196 | 0.067 |
| dan_daner_sickkids. | 1.00 | 0.478 | 0.338(275) | 0.338(205) | −0.13 | 0.184 |
| **Meta** | **0.99** | **3.41e−11** | **0.339(53660)** | **0.345(2044417)** | **−0.0465** | **0.007** |

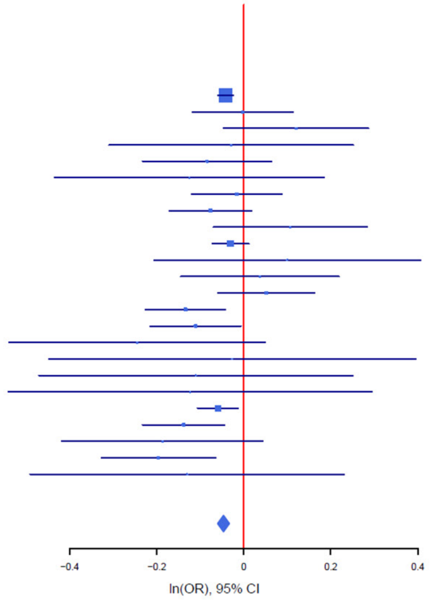

B

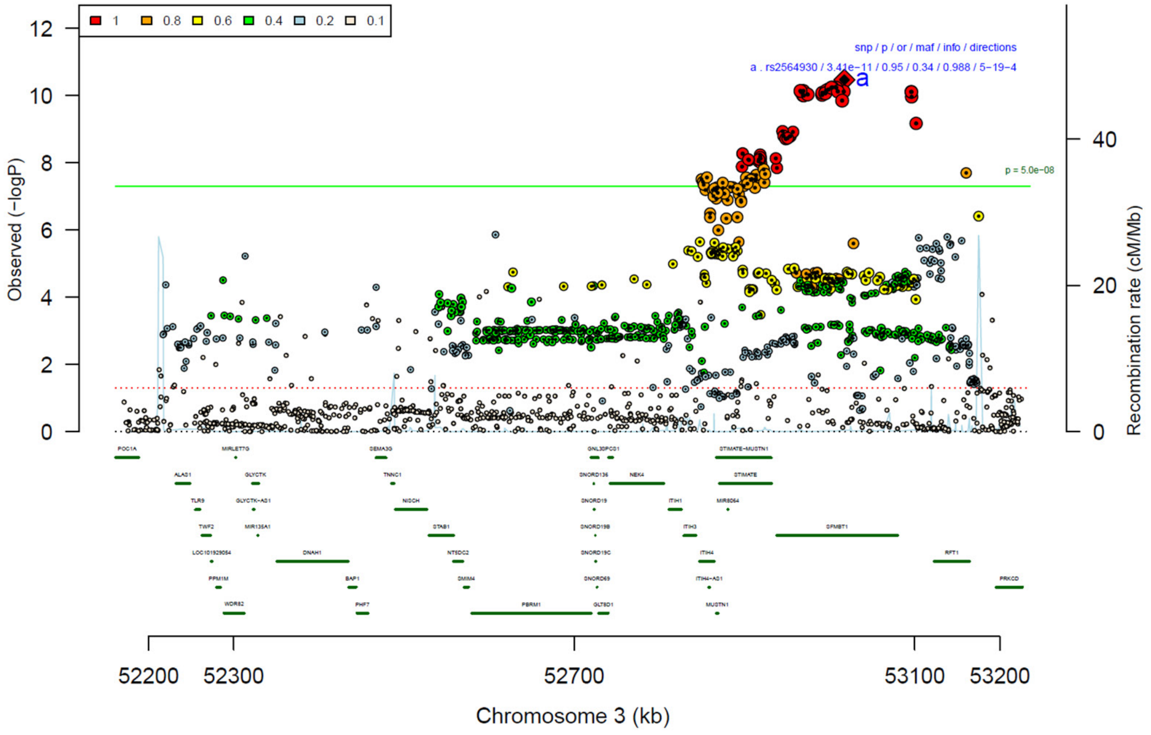

**Supplementary Figure 7: Forest plot (A) and regional association plot (B) of SNP rs2564930 in the meta-analysis.** The forest plot (A) shows the imputation quality (INFO) score, p-value of the SNP association (two-sided and not corrected for multiple testing, derived from a fixed-effects inverse-variance weighted meta-analysis), allele frequency in cases with case sample size (f_ca(n)), allele frequency in controls and control sample size (f_co(n)), beta estimates (ln(OR)) and standard error (STDerr) for each study as well as for the combined meta-analysis. Each cohort's sample size is detailed in supplementary table 1. Data on the right show the ln(OR) value as listed per cohort with 95% confidence intervals. In the regional association plot (B), the –log10 p-values (from two-sided association tests) are shown on the left y-axis. Recombination rates expressed in centimorgans (cM) per Mb (Megabase) (blue line) are shown on the right y-axis. Position in Mb is on the x-axis, with genes shown below the regional association plot. Only SNPs with an association p-value less than 0.1 were plotted. The most associated SNP is shown as a diamond and marked with the letter a.

A

| rs4702 | G/A | | 15:91426560 | | | |
|---|---|---|---|---|---|---|
| --?+--+--++--+---+--+--+-?+? | | | het_P: | 0.418 | het_I: | 3.1 |
| | info | p_value | f_ca(n) | f_co(n) | ln(OR) | STDerr |
| dan_daner_23andMe. | 1.00 | 1.56e−07 | 0.456(30167) | 0.456(929804) | 0.0439 | 0.00837 |
| dan_daner_AGDS. | 0.97 | 0.554 | 0.471(757) | 0.461(5368) | 0.0333 | 0.0563 |
| dan_daner_Chop. | 1.00 | 0.177 | 0.462(406) | 0.462(1369) | −0.11 | 0.0817 |
| dan_daner_Coga. | 1.00 | 0.0595 | 0.450(131) | 0.450(1358) | 0.256 | 0.136 |
| dan_daner_EGOS. | 0.91 | 0.276 | 0.470(926) | 0.452(1047) | 0.0801 | 0.0736 |
| dan_daner_EPOC. | 1.00 | 0.261 | 0.402(195) | 0.437(204) | −0.166 | 0.148 |
| dan_daner_EstBB. | 0.89 | 0.478 | 0.396(772) | 0.396(196079) | 0.0395 | 0.0557 |
| dan_daner_FinnGen. | 1.00 | 0.0317 | 0.496(1059) | 0.470(198110) | 0.0973 | 0.0453 |
| dan_daner_Hunt. | 0.96 | 0.701 | 0.430(284) | 0.444(66192) | −0.0336 | 0.0875 |
| dan_daner_MVP. | 0.99 | 0.176 | 0.451(5129) | 0.451(422860) | 0.0272 | 0.0201 |
| dan_daner_MoBa. | 0.89 | 0.392 | 0.418(104) | 0.444(2193) | −0.122 | 0.143 |
| dan_daner_NORDiC−NOR. | 0.91 | 0.472 | 0.442(599) | 0.440(619) | 0.0663 | 0.0922 |
| dan_daner_NORDiC−SWE. | 0.91 | 0.362 | 0.471(1691) | 0.456(1323) | 0.0508 | 0.0557 |
| dan_daner_Psych_Broad. | 0.99 | 0.524 | 0.466(1396) | 0.455(4009) | 0.0288 | 0.0451 |
| dan_daner_UKBB. | 1.00 | 0.432 | 0.445(776) | 0.445(125729) | −0.041 | 0.0522 |
| dan_daner_Wue. | 0.96 | 0.31 | 0.460(140) | 0.426(526) | 0.141 | 0.139 |
| dan_daner_YalePenn_GWCIDR. | 0.98 | 0.878 | 0.458(74) | 0.458(1635) | −0.0274 | 0.179 |
| dan_daner_iPSYCH. | 0.96 | 0.00591 | 0.447(4509) | 0.432(38392) | 0.0629 | 0.0229 |
| dan_daner_iocd. | 0.98 | 0.507 | 0.454(1519) | 0.445(3541) | 0.03 | 0.0453 |
| dan_daner_iocdf_trio. | 0.94 | 0.489 | 0.440(323) | 0.458(323) | −0.0797 | 0.115 |
| dan_daner_ocgas−ab. | 1.00 | 0.114 | 0.407(55) | 0.500(55) | −0.539 | 0.341 |
| dan_daner_ocgas−gh. | 0.86 | 0.444 | 0.481(56) | 0.437(56) | 0.228 | 0.298 |
| dan_daner_ocgas−nes. | 0.99 | 0.2 | 0.472(212) | 0.515(212) | −0.179 | 0.14 |
| dan_daner_ocgas. | 1.00 | 0.6 | 0.456(986) | 0.450(1023) | 0.0334 | 0.0637 |
| dan_daner_sickkids. | 1.00 | 0.173 | 0.457(275) | 0.457(205) | 0.226 | 0.166 |
| **Meta** | **0.98** | **9.07e−10** | **0.455(53660)** | **0.449(2044417)** | **0.0406** | **0.0066** |

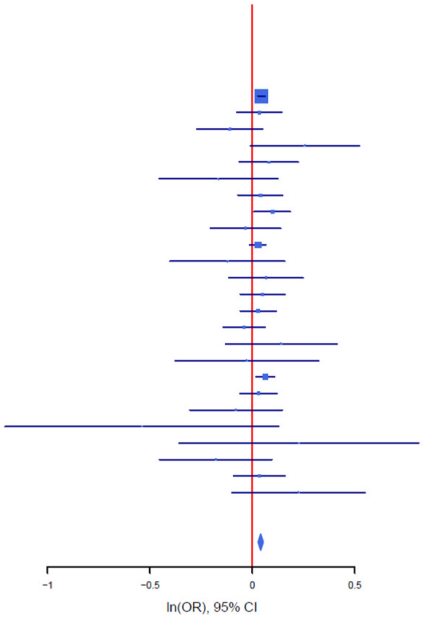

B

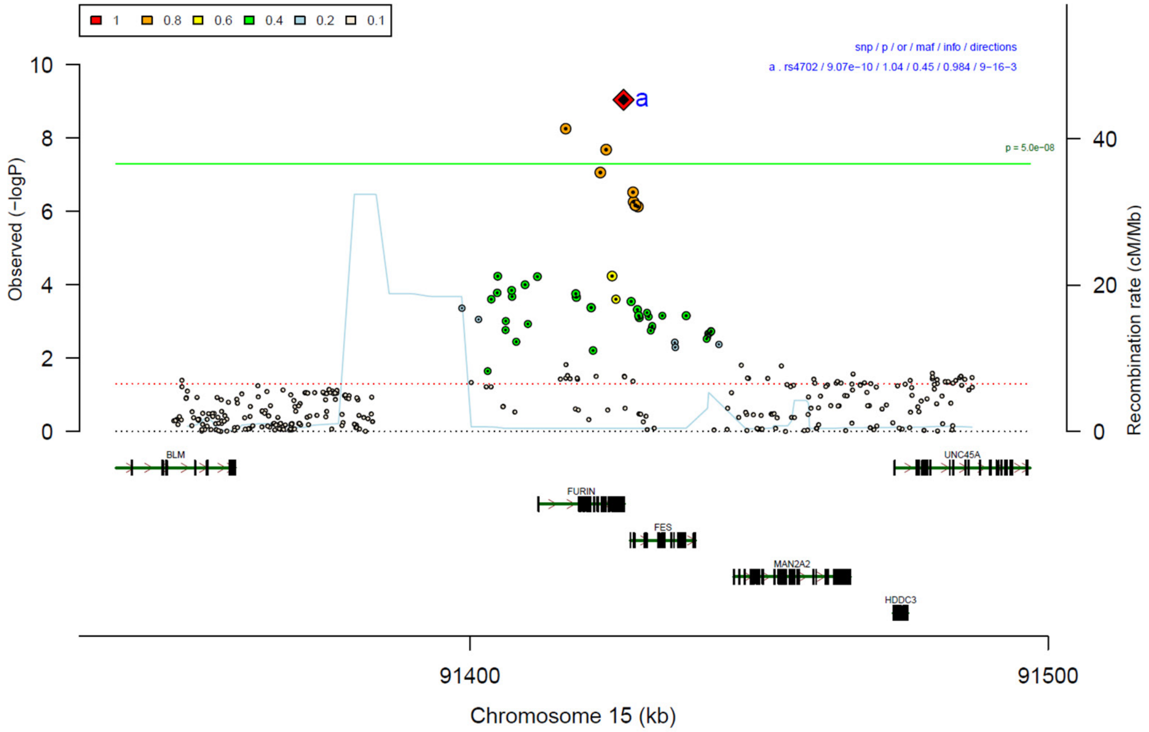

**Supplementary Figure 8: Forest plot (A) and regional association plot (B) of SNP rs4702 in the meta-analysis.** The forest plot (A) shows the imputation quality (INFO) score, p-value of the SNP association (two-sided and not corrected for multiple testing, derived from a fixed-effects inverse-variance weighted meta-analysis), allele frequency in cases with case sample size (f_ca(n)), allele frequency in controls and control sample size (f_co(n)), beta estimates (ln(OR)) and standard error (STDerr) for each study as well as for the combined meta-analysis. Each cohort's sample size is detailed in supplementary table 1. Data on the right show the ln(OR) value as listed per cohort with 95% confidence intervals. In the regional association plot (B), the –log10 p-values (from two-sided association tests) are shown on the left y-axis. Recombination rates expressed in centimorgans (cM) per Mb (Megabase) (blue line) are shown on the right y-axis. Position in Mb is on the x-axis, with genes shown below the regional association plot. Only SNPs with an association p-value less than 0.1 were plotted. The most associated SNP is shown as a diamond and marked with the letter a.

A

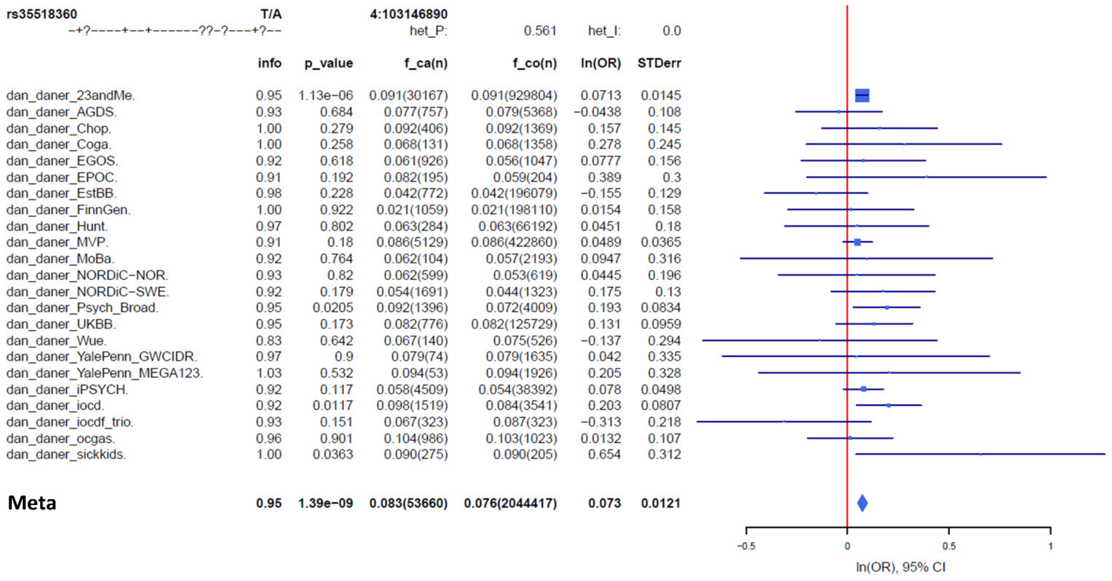

| | info | p_value | f_ca(n) | f_co(n) | ln(OR) | STDerr |
|---|---|---|---|---|---|---|
| dan_daner_23andMe. | 0.95 | 1.13e−06 | 0.091(30167) | 0.091(929804) | 0.0713 | 0.0145 |
| dan_daner_AGDS. | 0.93 | 0.684 | 0.077(757) | 0.079(5368) | −0.0438 | 0.108 |
| dan_daner_Chop. | 1.00 | 0.279 | 0.092(406) | 0.092(1369) | 0.157 | 0.145 |
| dan_daner_Coga. | 1.00 | 0.258 | 0.068(131) | 0.068(1358) | 0.278 | 0.245 |
| dan_daner_EGOS. | 0.92 | 0.618 | 0.061(926) | 0.056(1047) | 0.0777 | 0.156 |
| dan_daner_EPOC. | 0.91 | 0.192 | 0.082(195) | 0.059(204) | 0.389 | 0.3 |
| dan_daner_EstBB. | 0.98 | 0.228 | 0.042(772) | 0.042(196079) | −0.155 | 0.129 |
| dan_daner_FinnGen. | 1.00 | 0.922 | 0.021(1059) | 0.021(198110) | 0.0154 | 0.158 |
| dan_daner_Hunt. | 0.97 | 0.802 | 0.063(284) | 0.063(66192) | 0.0451 | 0.18 |
| dan_daner_MVP. | 0.91 | 0.18 | 0.086(5129) | 0.086(422860) | 0.0489 | 0.0365 |
| dan_daner_MoBa. | 0.92 | 0.764 | 0.062(104) | 0.057(2193) | 0.0947 | 0.316 |
| dan_daner_NORDiC−NOR. | 0.93 | 0.82 | 0.062(599) | 0.053(619) | 0.0445 | 0.196 |
| dan_daner_NORDiC−SWE. | 0.92 | 0.179 | 0.054(1691) | 0.044(1323) | 0.175 | 0.13 |
| dan_daner_Psych_Broad. | 0.95 | 0.0205 | 0.092(1396) | 0.072(4009) | 0.193 | 0.0834 |
| dan_daner_UKBB. | 0.95 | 0.173 | 0.082(776) | 0.082(125729) | 0.131 | 0.0959 |
| dan_daner_Wue. | 0.83 | 0.642 | 0.067(140) | 0.075(526) | −0.137 | 0.294 |
| dan_daner_YalePenn_GWCIDR. | 0.97 | 0.9 | 0.079(74) | 0.079(1635) | 0.042 | 0.335 |
| dan_daner_YalePenn_MEGA123. | 1.03 | 0.532 | 0.094(53) | 0.094(1926) | 0.205 | 0.328 |
| dan_daner_iPSYCH. | 0.92 | 0.117 | 0.058(4509) | 0.054(38392) | 0.078 | 0.0498 |
| dan_daner_iocd. | 0.92 | 0.0117 | 0.098(1519) | 0.084(3541) | 0.203 | 0.0807 |
| dan_daner_iocdf_trio. | 0.93 | 0.151 | 0.067(323) | 0.087(323) | −0.313 | 0.218 |
| dan_daner_ocgas. | 0.96 | 0.901 | 0.104(986) | 0.103(1023) | 0.0132 | 0.107 |
| dan_daner_sickkids. | 1.00 | 0.0363 | 0.090(275) | 0.090(205) | 0.654 | 0.312 |
| **Meta** | **0.95** | **1.39e−09** | **0.083(53660)** | **0.076(2044417)** | **0.073** | **0.0121** |

B

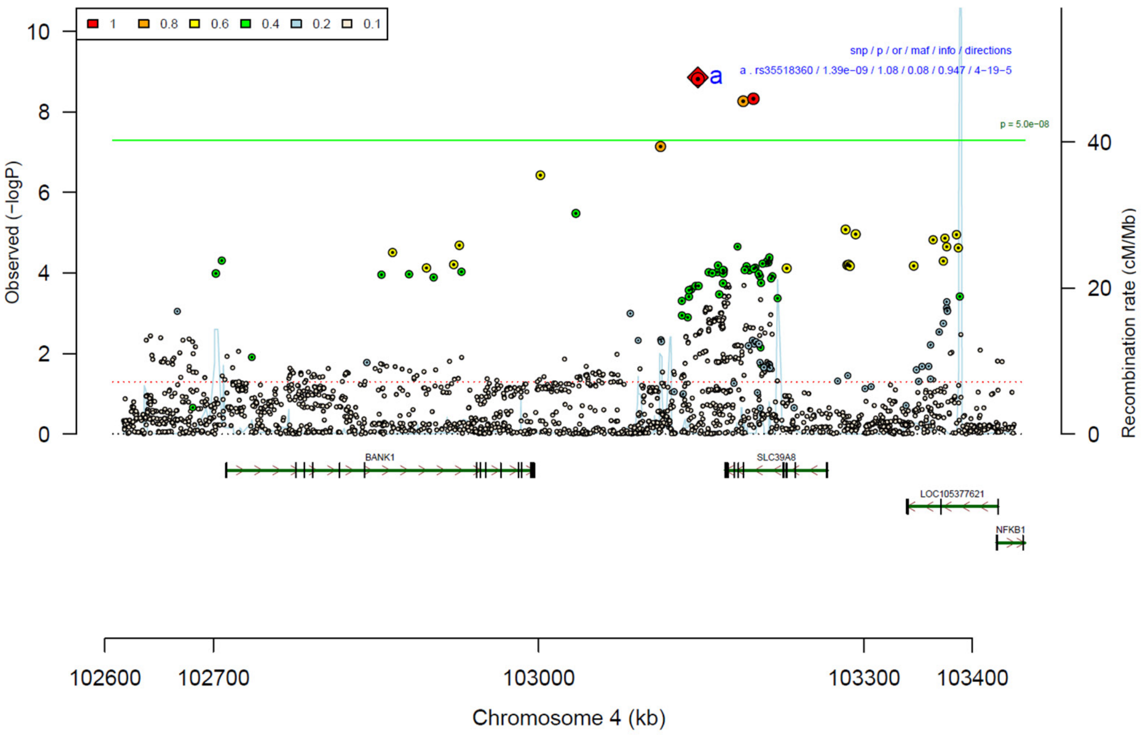

**Supplementary Figure 9: Forest plot (A) and regional association plot (B) of SNP rs35518360 in the meta-analysis.** The forest plot (A) shows the imputation quality (INFO) score, p-value of the SNP association (two-sided and not corrected for multiple testing, derived from a fixed-effects inverse-variance weighted meta-analysis), allele frequency in cases with case sample size (f_ca(n)), allele frequency in controls and control sample size (f_co(n)), beta estimates (ln(OR)) and standard error (STDerr) for each study as well as for the combined meta-analysis. Each cohort's sample size is detailed in supplementary table 1. Data on the right show the ln(OR) value as listed per cohort with 95% confidence intervals. In the regional association plot (B), the –log10 p-values (from two-sided association tests) are shown on the left y-axis. Recombination rates expressed in centimorgans (cM) per Mb (Megabase) (blue line) are shown on the right y-axis. Position in Mb is on the x-axis, with genes shown below the regional association plot. Only SNPs with an association p-value less than 0.1 were plotted. The most associated SNP is shown as a diamond and marked with the letter a.

A

| rs4904738 | T/C | | 14:42179732 | | | |
|---|---|---|---|---|---|---|
| --?---+----+------?---++-+-- |  |  | het_P: | 0.426 | het_I: | 2.6 |
| | **info** | **p_value** | **f_ca(n)** | **f_co(n)** | **ln(OR)** | **STDerr** |
| dan_daner_23andMe. | 1.00 | 1.83e−05 | 0.565(30167) | 0.565(929804) | −0.0362 | 0.00844 |
| dan_daner_AGDS. | 0.98 | 0.676 | 0.562(757) | 0.569(5368) | −0.0236 | 0.0564 |
| dan_daner_Chop. | 1.00 | 0.000593 | 0.567(406) | 0.567(1369) | −0.284 | 0.0827 |
| dan_daner_Coga. | 1.00 | 0.122 | 0.555(131) | 0.555(1358) | −0.212 | 0.137 |
| dan_daner_EGOS. | 1.01 | 0.907 | 0.527(926) | 0.527(1047) | −0.00813 | 0.0692 |
| dan_daner_EPOC. | 0.94 | 0.877 | 0.534(195) | 0.527(204) | 0.023 | 0.148 |
| dan_daner_EstBB. | 0.98 | 0.529 | 0.512(772) | 0.512(196079) | −0.0324 | 0.0516 |
| dan_daner_FinnGen. | 1.00 | 0.277 | 0.489(1059) | 0.501(198110) | −0.0491 | 0.0452 |
| dan_daner_Hunt. | 0.99 | 0.105 | 0.496(284) | 0.531(66192) | −0.139 | 0.0856 |
| dan_daner_MVP. | 0.95 | 0.065 | 0.564(5129) | 0.564(422860) | −0.038 | 0.0206 |
| dan_daner_MoBa. | 0.99 | 0.897 | 0.524(104) | 0.530(2193) | −0.0185 | 0.142 |
| dan_daner_NORDiC−NOR. | 0.98 | 0.462 | 0.517(599) | 0.521(619) | −0.0649 | 0.0882 |
| dan_daner_NORDiC−SWE. | 0.99 | 0.0801 | 0.531(1691) | 0.557(1323) | −0.0938 | 0.0535 |
| dan_daner_Psych_Broad. | 0.99 | 0.179 | 0.541(1396) | 0.553(4009) | −0.0607 | 0.0452 |
| dan_daner_UKBB. | 0.99 | 0.457 | 0.573(776) | 0.573(125729) | 0.0388 | 0.0522 |
| dan_daner_Wue. | 1.01 | 0.791 | 0.540(140) | 0.548(526) | −0.0359 | 0.136 |
| dan_daner_YalePenn_EXHCE. | 1.00 | 0.991 | 0.554(52) | 0.554(1646) | 0.0023 | 0.213 |
| dan_daner_YalePenn_GWCIDR. | 0.95 | 0.529 | 0.544(74) | 0.544(1635) | −0.114 | 0.181 |
| dan_daner_YalePenn_MEGA123. | 1.00 | 0.0781 | 0.548(53) | 0.548(1926) | −0.362 | 0.206 |
| dan_daner_iPSYCH. | 0.98 | 0.0352 | 0.558(4509) | 0.569(38392) | −0.0476 | 0.0226 |
| dan_daner_iocd. | 1.00 | 0.883 | 0.563(1519) | 0.560(3541) | 0.00658 | 0.0449 |
| dan_daner_iocdf_trio. | 1.01 | 0.316 | 0.539(323) | 0.567(323) | −0.112 | 0.112 |
| dan_daner_ocgas−ab. | 0.99 | 0.0771 | 0.483(55) | 0.612(55) | −0.598 | 0.339 |
| dan_daner_ocgas−nes. | 0.98 | 0.0717 | 0.510(212) | 0.572(212) | −0.254 | 0.141 |
| dan_daner_ocgas. | 1.03 | 0.599 | 0.550(986) | 0.557(1023) | −0.0329 | 0.0627 |
| dan_daner_sickkids. | 1.00 | 0.69 | 0.535(275) | 0.535(205) | 0.0701 | 0.176 |
| **Meta** | **0.98** | **1.48e−09** | **0.558(53660)** | **0.553(2044417)** | **−0.0403** | **0.0067** |

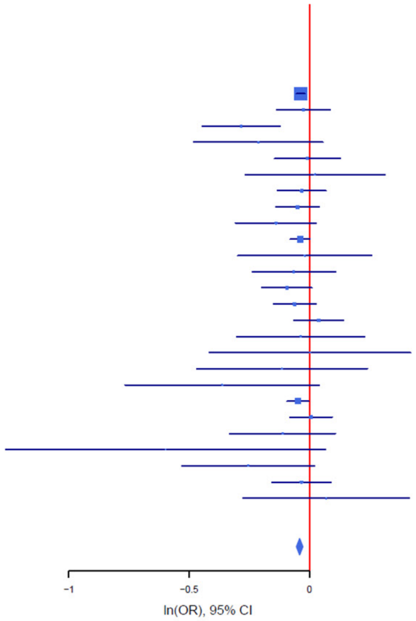

B

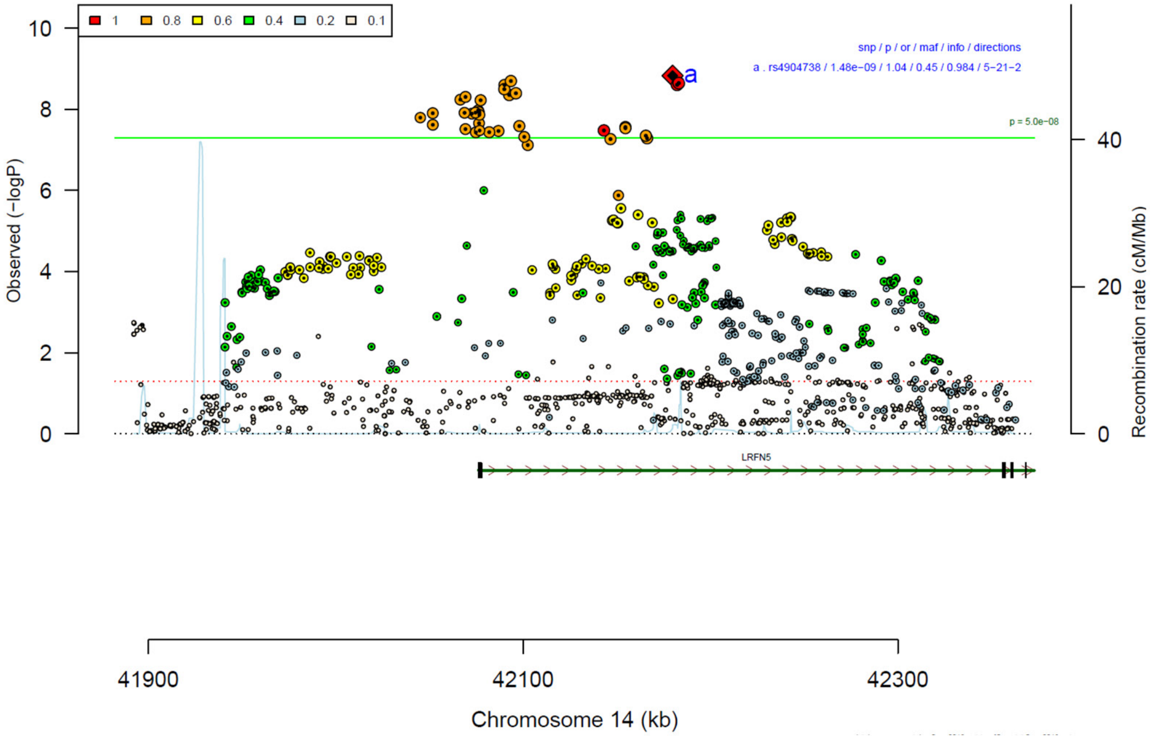

**Supplementary Figure 10: Forest plot (A) and regional association plot (B) of SNP rs4904738 in the meta-analysis.** The forest plot (A) shows the imputation quality (INFO) score, p-value of the SNP association (two-sided and not corrected for multiple testing, derived from a fixed-effects inverse-variance weighted meta-analysis), allele frequency in cases with case sample size (f_ca(n)), allele frequency in controls and control sample size (f_co(n)), beta estimates (ln(OR)) and standard error (STDerr) for each study as well as for the combined meta-analysis. Each cohort's sample size is detailed in supplementary table 1. Data on the right show the ln(OR) value as listed per cohort with 95% confidence intervals. In the regional association plot (B), the –log10 p-values (from two-sided association tests) are shown on the left y-axis. Recombination rates expressed in centimorgans (cM) per Mb (Megabase) (blue line) are shown on the right y-axis. Position in Mb is on the x-axis, with genes shown below the regional association plot. Only SNPs with an association p-value less than 0.1 were plotted. The most associated SNP is shown as a diamond and marked with the letter a.

A

| rs254779 | T/C | | 5:88019629 | | | |
|---|---|---|---|---|---|---|
| -------+-----+-+-??+-------- |  |  | het_P: | 0.664 | het_I: | 0.0 |
| | info | p_value | f_ca(n) | f_co(n) | ln(OR) | STDerr |
| dan_daner_23andMe. | 1.00 | 8.79e−06 | 0.425(30167) | 0.425(929804) | −0.0378 | 0.0085 |
| dan_daner_AGDS. | 0.96 | 0.391 | 0.422(757) | 0.433(5368) | −0.0492 | 0.0574 |
| dan_daner_Chop. | 1.00 | 0.278 | 0.431(406) | 0.431(1369) | −0.0892 | 0.0822 |
| dan_daner_Coga. | 1.00 | 0.916 | 0.429(131) | 0.429(1358) | −0.0138 | 0.131 |
| dan_daner_EGOS. | 0.94 | 0.99 | 0.394(926) | 0.395(1047) | −0.0009 | 0.0737 |
| dan_daner_EPOC. | 0.95 | 0.105 | 0.377(195) | 0.420(204) | −0.253 | 0.157 |
| dan_daner_EstBB. | 0.97 | 0.75 | 0.407(772) | 0.407(196079) | 0.0168 | 0.0528 |
| dan_daner_FinnGen. | 1.00 | 0.0939 | 0.385(1059) | 0.406(198110) | −0.0767 | 0.0458 |
| dan_daner_Hunt. | 0.99 | 0.549 | 0.392(284) | 0.401(66192) | −0.0519 | 0.0867 |
| dan_daner_MVP. | 0.96 | 0.0278 | 0.432(5129) | 0.432(422860) | −0.0453 | 0.0206 |
| dan_daner_MoBa. | 0.98 | 0.116 | 0.462(104) | 0.408(2193) | 0.23 | 0.146 |
| dan_daner_NORDiC−NOR. | 0.90 | 0.208 | 0.412(599) | 0.387(619) | 0.118 | 0.0938 |
| dan_daner_NORDiC−SWE. | 0.92 | 0.25 | 0.391(1691) | 0.406(1323) | −0.065 | 0.0565 |
| dan_daner_Psych_Broad. | 1.00 | 0.602 | 0.430(1396) | 0.429(4009) | −0.0238 | 0.0455 |
| dan_daner_UKBB. | 1.00 | 0.378 | 0.415(776) | 0.415(125729) | −0.0462 | 0.0524 |
| dan_daner_Wue. | 0.96 | 0.316 | 0.382(140) | 0.414(526) | −0.142 | 0.141 |
| dan_daner_YalePenn_EXHCE. | 0.99 | 0.195 | 0.407(52) | 0.407(1646) | −0.287 | 0.221 |
| dan_daner_YalePenn_GWCIDR. | 0.98 | 0.195 | 0.429(74) | 0.429(1635) | −0.24 | 0.185 |
| dan_daner_YalePenn_MEGA123. | 0.98 | 0.382 | 0.435(53) | 0.435(1926) | −0.183 | 0.209 |
| dan_daner_bioVU. | 0.99 | 0.54 | 0.409(1014) | 0.414(38613) | −0.0284 | 0.0464 |
| dan_daner_iPSYCH. | 0.98 | 0.0505 | 0.390(4509) | 0.401(38392) | −0.0451 | 0.023 |
| dan_daner_iocd. | 0.98 | 0.0869 | 0.405(1519) | 0.421(3541) | −0.0785 | 0.0459 |
| dan_daner_iocdf_trio. | 1.05 | 0.411 | 0.416(323) | 0.440(323) | −0.0904 | 0.11 |
| dan_daner_ocgas−nes. | 0.82 | 0.0201 | 0.454(212) | 0.523(212) | −0.364 | 0.156 |
| dan_daner_ocgas. | 0.99 | 0.542 | 0.417(986) | 0.406(1023) | 0.0395 | 0.0648 |
| dan_daner_sickkids. | 1.00 | 0.477 | 0.450(275) | 0.450(205) | −0.124 | 0.174 |
| **Meta** | **0.99** | **1.53e−09** | **0.419(53660)** | **0.421(2044417)** | **−0.0402** | **0.0067** |

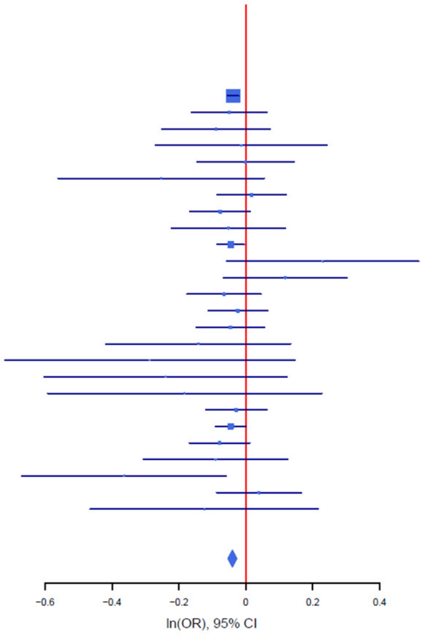

B

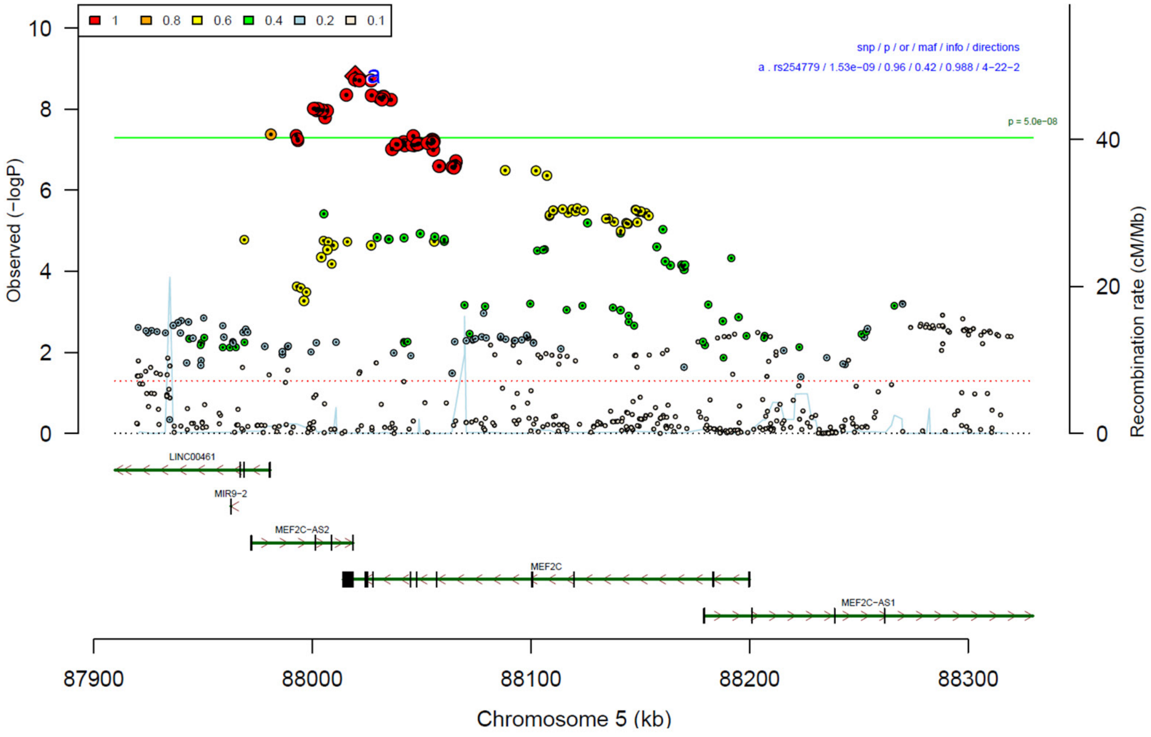

**Supplementary Figure 11: Forest plot (A) and regional association plot (B) of SNP rs254779 in the meta-analysis.** The forest plot (A) shows the imputation quality (INFO) score, p-value of the SNP association (two-sided and not corrected for multiple testing, derived from a fixed-effects inverse-variance weighted meta-analysis), allele frequency in cases with case sample size (f_ca(n)), allele frequency in controls and control sample size (f_co(n)), beta estimates (ln(OR)) and standard error (STDerr) for each study as well as for the combined meta-analysis. Each cohort's sample size is detailed in supplementary table 1. Data on the right show the ln(OR) value as listed per cohort with 95% confidence intervals. In the regional association plot (B), the –log10 p-values (from two-sided association tests) are shown on the left y-axis. Recombination rates expressed in centimorgans (cM) per Mb (Megabase) (blue line) are shown on the right y-axis. Position in Mb is on the x-axis, with genes shown below the regional association plot. Only SNPs with an association p-value less than 0.1 were plotted. The most associated SNP is shown as a diamond and marked with the letter a.

A

| rs2198140 | T/C | 8:35039646 | | | | |
|---|---|---|---|---|---|---|
| ------+--------+?-+-+----+-++- |  | het_P: | 0.911 | het_I: | 0.0 |  |
| | info | p_value | f_ca(n) | f_co(n) | ln(OR) | STDerr |
| dan_daner_23andMe. | 0.97 | 7.24e−08 | 0.494(30167) | 0.494(929804) | −0.0459 | 0.00853 |
| dan_daner_AGDS. | 0.98 | 0.408 | 0.493(757) | 0.505(5368) | −0.0464 | 0.056 |
| dan_daner_Chop. | 1.00 | 0.217 | 0.490(406) | 0.490(1369) | −0.105 | 0.085 |
| dan_daner_Coga. | 1.00 | 0.377 | 0.483(131) | 0.483(1358) | −0.126 | 0.143 |
| dan_daner_EGOS. | 0.95 | 0.804 | 0.505(926) | 0.512(1047) | 0.0177 | 0.0716 |
| dan_daner_EPOC. | 0.93 | 0.79 | 0.483(195) | 0.491(204) | −0.0407 | 0.153 |
| dan_daner_EstBB. | 1.00 | 0.131 | 0.564(772) | 0.564(196079) | −0.0778 | 0.0515 |
| dan_daner_FinnGen. | 1.00 | 0.592 | 0.579(1059) | 0.580(198110) | −0.0245 | 0.0457 |
| dan_daner_Hunt. | 1.00 | 0.59 | 0.478(284) | 0.491(66192) | −0.0459 | 0.0851 |
| dan_daner_MoBa. | 0.99 | 0.831 | 0.510(104) | 0.503(2193) | 0.0305 | 0.143 |
| dan_daner_NORDiC−NOR. | 0.98 | 0.862 | 0.498(599) | 0.499(619) | −0.0154 | 0.0883 |
| dan_daner_NORDiC−SWE. | 0.96 | 0.949 | 0.505(1691) | 0.501(1323) | 0.00349 | 0.0542 |
| dan_daner_Psych_Broad. | 1.00 | 0.0883 | 0.465(1396) | 0.481(4009) | −0.0766 | 0.0449 |
| dan_daner_UKBB. | 0.99 | 0.264 | 0.495(776) | 0.495(125729) | 0.0579 | 0.0518 |
| dan_daner_Wue. | 1.01 | 0.0874 | 0.441(140) | 0.499(526) | −0.231 | 0.135 |
| dan_daner_YalePenn_EXHCE. | 1.02 | 0.368 | 0.500(52) | 0.500(1646) | 0.184 | 0.204 |
| dan_daner_YalePenn_GWCIDR. | 0.95 | 0.737 | 0.497(74) | 0.497(1635) | 0.0605 | 0.181 |
| dan_daner_YalePenn_MEGA123. | 0.99 | 0.327 | 0.491(53) | 0.491(1926) | −0.207 | 0.211 |
| dan_daner_bioVU. | 0.99 | 0.548 | 0.493(1014) | 0.500(38613) | −0.0275 | 0.0457 |
| dan_daner_iPSYCH. | 0.99 | 0.436 | 0.501(4509) | 0.505(38392) | −0.0174 | 0.0224 |
| dan_daner_iocd. | 0.96 | 0.264 | 0.475(1519) | 0.493(3541) | −0.051 | 0.0456 |
| dan_daner_iocdf_trio. | 1.00 | 0.53 | 0.490(323) | 0.507(323) | −0.0699 | 0.111 |
| dan_daner_ocgas−ab. | 1.12 | 0.245 | 0.466(55) | 0.547(55) | −0.358 | 0.308 |
| dan_daner_ocgas−gh. | 0.85 | 0.439 | 0.487(56) | 0.445(56) | 0.234 | 0.303 |
| dan_daner_ocgas−nes. | 0.86 | 0.298 | 0.477(212) | 0.510(212) | −0.156 | 0.15 |
| dan_daner_ocgas. | 1.01 | 0.0822 | 0.465(986) | 0.494(1023) | −0.11 | 0.0632 |
| dan_daner_sickkids. | 1.00 | 0.787 | 0.540(275) | 0.540(205) | −0.0457 | 0.169 |
| **Meta** | **0.98** | **2.18e−09** | **0.496(53660)** | **0.513(2044417)** | **−0.0419** | **0.007** |

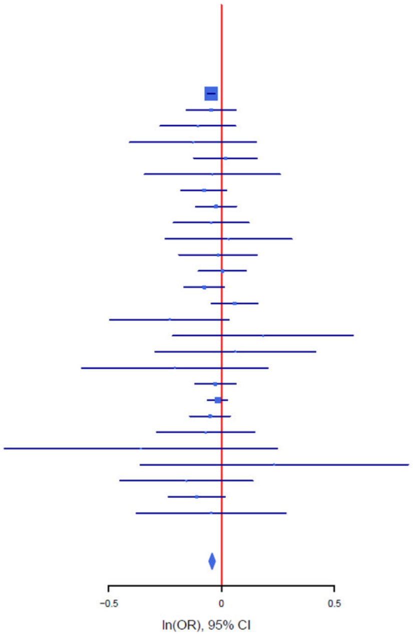

B

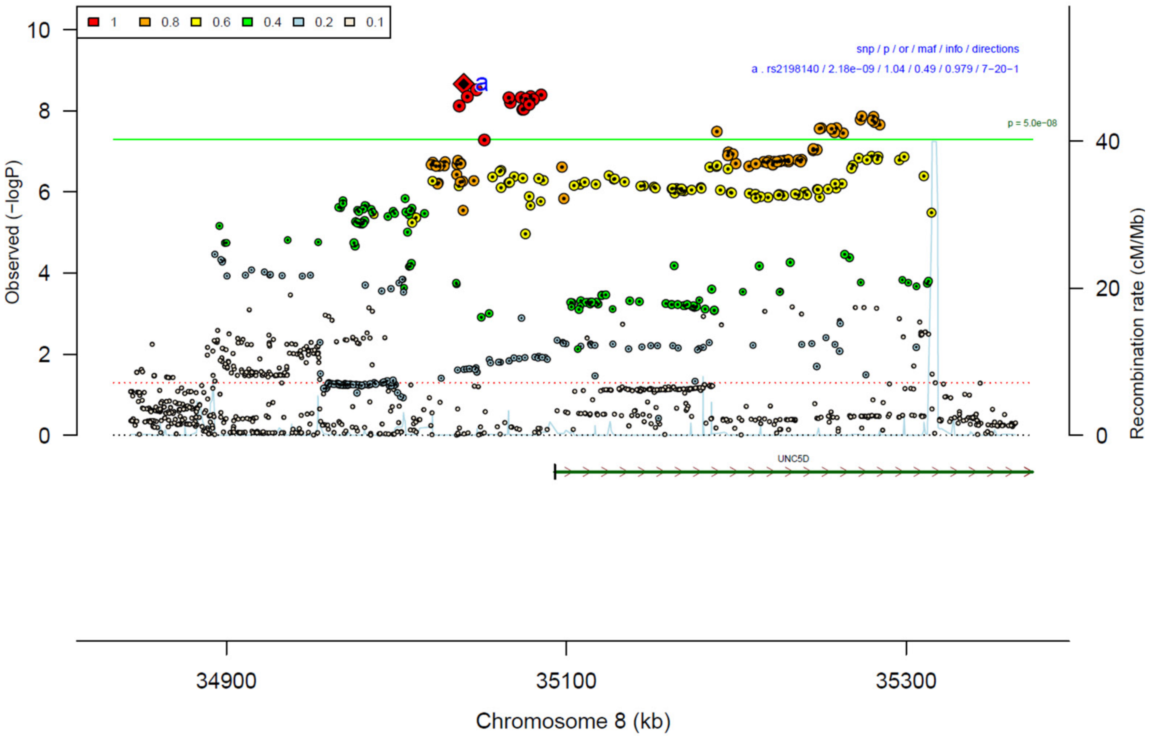

**Supplementary Figure 12: Forest plot (A) and regional association plot (B) of SNP rs2198140 in the meta-analysis.** The forest plot (A) shows the imputation quality (INFO) score, p-value of the SNP association (two-sided and not corrected for multiple testing, derived from a fixed-effects inverse-variance weighted meta-analysis), allele frequency in cases with case sample size (f_ca(n)), allele frequency in controls and control sample size (f_co(n)), beta estimates (ln(OR)) and standard error (STDerr) for each study as well as for the combined meta-analysis. Each cohort's sample size is detailed in supplementary table 1. Data on the right show the ln(OR) value as listed per cohort with 95% confidence intervals. In the regional association plot (B), the –log10 p-values (from two-sided association tests) are shown on the left y-axis. Recombination rates expressed in centimorgans (cM) per Mb (Megabase) (blue line) are shown on the right y-axis. Position in Mb is on the x-axis, with genes shown below the regional association plot. Only SNPs with an association p-value less than 0.1 were plotted. The most associated SNP is shown as a diamond and marked with the letter a.

A

rs12516488 G/A 5:45203199

---+-+--?-+--------++++-+--+- het_P: 0.731 het_I: 0.0

| | info | p_value | f_ca(n) | f_co(n) | ln(OR) | STDerr |
|---|---|---|---|---|---|---|
| dan_daner_23andMe. | 1.00 | 1.31e−08 | 0.817(30167) | 0.817(929804) | 0.0624 | 0.011 |
| dan_daner_AGDS. | 0.98 | 0.209 | 0.834(757) | 0.822(5368) | 0.0943 | 0.0751 |
| dan_daner_Chop. | 1.00 | 0.737 | 0.796(406) | 0.796(1369) | −0.0331 | 0.0984 |
| dan_daner_Coga. | 1.00 | 0.28 | 0.820(131) | 0.820(1358) | 0.193 | 0.179 |
| dan_daner_EGOS. | 1.02 | 0.636 | 0.853(926) | 0.856(1047) | −0.0463 | 0.0977 |
| dan_daner_EPOC. | 0.99 | 0.798 | 0.833(195) | 0.831(204) | 0.0491 | 0.191 |
| dan_daner_EstBB. | 1.00 | 0.298 | 0.861(772) | 0.861(196079) | 0.0774 | 0.0743 |
| dan_daner_Hunt. | 1.00 | 0.222 | 0.847(284) | 0.823(66192) | 0.137 | 0.112 |
| dan_daner_MVP. | 0.98 | 0.0683 | 0.831(5129) | 0.831(422860) | 0.0497 | 0.0273 |
| dan_daner_MoBa. | 1.00 | 0.24 | 0.870(104) | 0.839(2193) | 0.232 | 0.197 |
| dan_daner_NORDiC−NOR. | 0.99 | 0.623 | 0.855(599) | 0.852(619) | 0.0608 | 0.124 |
| dan_daner_NORDiC−SWE. | 1.00 | 0.307 | 0.860(1691) | 0.852(1323) | 0.0773 | 0.0757 |
| dan_daner_Psych_Broad. | 1.02 | 0.434 | 0.805(1396) | 0.822(4009) | −0.0445 | 0.0569 |
| dan_daner_UKBB. | 1.00 | 0.975 | 0.823(776) | 0.823(125729) | −0.00211 | 0.0679 |
| dan_daner_Wue. | 1.00 | 0.179 | 0.846(140) | 0.812(526) | 0.249 | 0.185 |
| dan_daner_YalePenn_EXHCE. | 1.01 | 0.554 | 0.804(52) | 0.804(1646) | 0.159 | 0.269 |
| dan_daner_YalePenn_GWCIDR. | 1.00 | 0.848 | 0.814(74) | 0.814(1635) | −0.0434 | 0.227 |
| dan_daner_YalePenn_MEGA123. | 0.99 | 0.54 | 0.800(53) | 0.800(1926) | 0.166 | 0.271 |
| dan_daner_bioVU. | 1.00 | 0.32 | 0.842(1014) | 0.832(38613) | 0.0609 | 0.0613 |
| dan_daner_iPSYCH. | 1.00 | 0.993 | 0.847(4509) | 0.847(38392) | 0.0003 | 0.031 |
| dan_daner_iocd. | 1.02 | 0.097 | 0.819(1519) | 0.810(3541) | 0.0959 | 0.0578 |
| dan_daner_iocdf_trio. | 1.02 | 0.439 | 0.805(323) | 0.822(323) | −0.11 | 0.142 |
| dan_daner_ocgas−ab. | 1.11 | 0.434 | 0.860(55) | 0.795(55) | 0.322 | 0.412 |
| dan_daner_ocgas−gh. | 1.10 | 0.148 | 0.786(56) | 0.866(56) | −0.523 | 0.361 |
| dan_daner_ocgas−nes. | 0.95 | 0.216 | 0.785(212) | 0.821(212) | −0.223 | 0.18 |
| dan_daner_ocgas. | 0.97 | 0.872 | 0.815(986) | 0.816(1023) | −0.0134 | 0.083 |
| dan_daner_sickkids. | 1.00 | 0.478 | 0.831(275) | 0.831(205) | 0.16 | 0.225 |
| **Meta** | **0.99** | **3.78e−09** | **0.825(53660)** | **0.826(2044417)** | **0.0517** | **0.0088** |

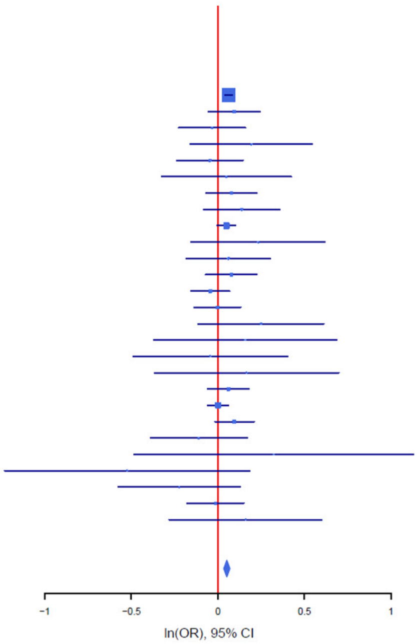

B

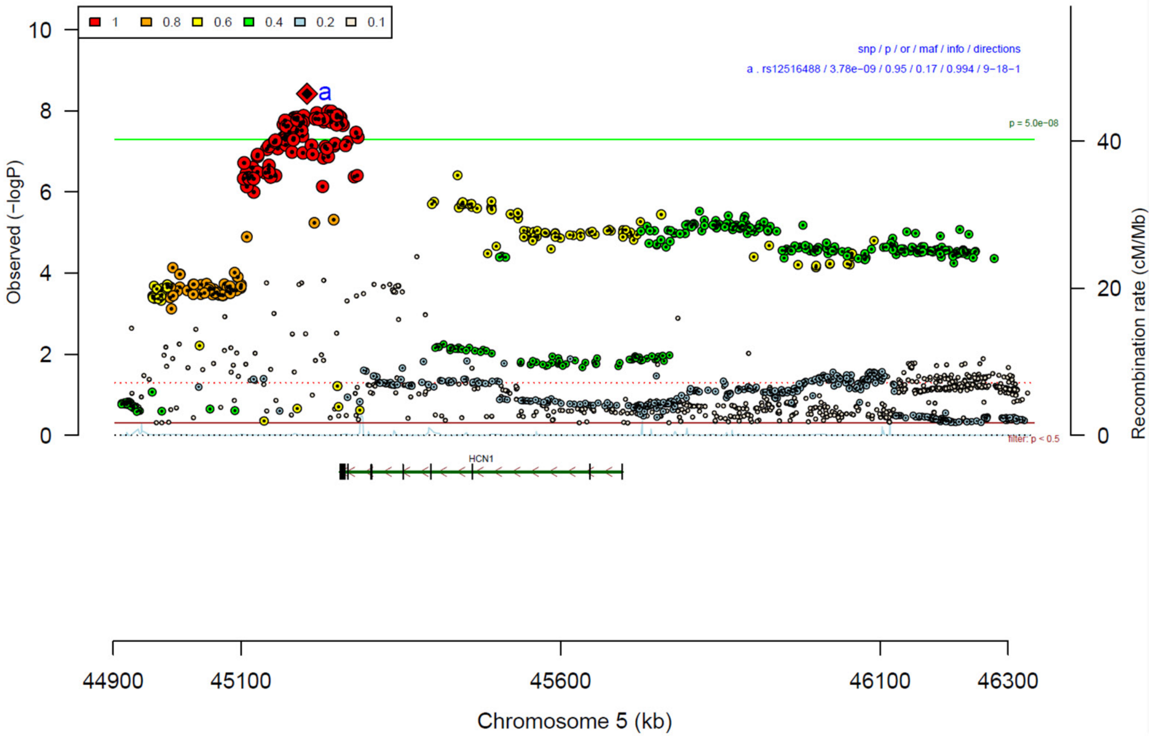

**Supplementary Figure 13: Forest plot (A) and regional association plot (B) of SNP rs12516488 in the meta-analysis.** The forest plot (A) shows the imputation quality (INFO) score, p-value of the SNP association (two-sided and not corrected for multiple testing, derived from a fixed-effects inverse-variance weighted meta-analysis), allele frequency in cases with case sample size (f_ca(n)), allele frequency in controls and control sample size (f_co(n)), beta estimates (ln(OR)) and standard error (STDerr) for each study as well as for the combined meta-analysis. Each cohort's sample size is detailed in supplementary table 1. Data on the right show the ln(OR) value as listed per cohort with 95% confidence intervals. In the regional association plot (B), the –log10 p-values (from two-sided association tests) are shown on the left y-axis. Recombination rates expressed in centimorgans (cM) per Mb (Megabase) (blue line) are shown on the right y-axis. Position in Mb is on the x-axis, with genes shown below the regional association plot. Only SNPs with an association p-value less than 0.1 were plotted. The most associated SNP is shown as a diamond and marked with the letter a.

A

rs3899258 G/A 5:50256551

--+----+--+--+?-----+--+++-- het_P: 0.711 het_I: 0.0

| | info | p_value | f_ca(n) | f_co(n) | ln(OR) | STDerr |
|---|---|---|---|---|---|---|
| dan_daner_23andMe. | 0.98 | 8.84e−09 | 0.777(30167) | 0.777(929804) | 0.0588 | 0.0103 |
| dan_daner_AGDS. | 0.99 | 0.513 | 0.776(757) | 0.770(5368) | 0.0434 | 0.0664 |
| dan_daner_Chop. | 1.00 | 0.00569 | 0.766(406) | 0.766(1369) | 0.283 | 0.102 |
| dan_daner_Coga. | 1.00 | 0.0813 | 0.793(131) | 0.793(1358) | 0.309 | 0.177 |
| dan_daner_EGOS. | 0.99 | 0.293 | 0.794(926) | 0.773(1047) | 0.0897 | 0.0853 |
| dan_daner_EPOC. | 0.97 | 0.215 | 0.800(195) | 0.771(204) | 0.233 | 0.188 |
| dan_daner_EstBB. | 1.00 | 0.988 | 0.817(772) | 0.817(196079) | −0.000975 | 0.0663 |
| dan_daner_FinnGen. | 1.00 | 0.439 | 0.863(1059) | 0.855(198110) | 0.0498 | 0.0644 |
| dan_daner_Hunt. | 1.00 | 0.768 | 0.798(284) | 0.791(66192) | 0.0308 | 0.105 |
| dan_daner_MoBa. | 0.99 | 0.345 | 0.750(104) | 0.776(2193) | −0.162 | 0.172 |
| dan_daner_NORDiC−NOR. | 0.97 | 0.853 | 0.797(599) | 0.797(619) | 0.0206 | 0.111 |
| dan_daner_NORDiC−SWE. | 0.97 | 0.0637 | 0.814(1691) | 0.797(1323) | 0.126 | 0.0681 |
| dan_daner_Psych_Broad. | 1.02 | 0.616 | 0.758(1396) | 0.745(4009) | 0.026 | 0.0516 |
| dan_daner_UKBB. | 0.98 | 0.873 | 0.777(776) | 0.777(125729) | −0.01 | 0.0625 |
| dan_daner_Wue. | 1.03 | 0.741 | 0.769(140) | 0.777(526) | −0.0525 | 0.159 |
| dan_daner_YalePenn_EXHCE. | 1.01 | 0.678 | 0.774(52) | 0.774(1646) | −0.0976 | 0.235 |
| dan_daner_YalePenn_GWCIDR. | 0.99 | 0.726 | 0.782(74) | 0.782(1635) | 0.0769 | 0.22 |
| dan_daner_YalePenn_MEGA123. | 1.02 | 0.328 | 0.769(53) | 0.769(1926) | 0.249 | 0.255 |
| dan_daner_bioVU. | 0.99 | 0.815 | 0.779(1014) | 0.781(38613) | −0.0129 | 0.0554 |
| dan_daner_iPSYCH. | 0.99 | 0.644 | 0.782(4509) | 0.780(38392) | 0.0125 | 0.0271 |
| dan_daner_iocd. | 0.98 | 0.498 | 0.778(1519) | 0.771(3541) | 0.0367 | 0.0541 |
| dan_daner_iocdf_trio. | 0.97 | 0.514 | 0.768(323) | 0.783(323) | −0.0886 | 0.136 |
| dan_daner_ocgas−ab. | 0.99 | 0.922 | 0.787(55) | 0.790(55) | 0.039 | 0.399 |
| dan_daner_ocgas−gh. | 1.11 | 0.598 | 0.779(56) | 0.769(56) | 0.167 | 0.316 |
| dan_daner_ocgas−nes. | 1.00 | 0.313 | 0.765(212) | 0.797(212) | −0.17 | 0.168 |
| dan_daner_ocgas. | 0.96 | 0.811 | 0.783(986) | 0.779(1023) | 0.0186 | 0.0781 |
| dan_daner_sickkids. | 1.00 | 0.752 | 0.771(275) | 0.771(205) | 0.0653 | 0.207 |
| **Meta** | **0.99** | **4.94e−09** | **0.782(53660)** | **0.792(2044417)** | **0.0496** | **0.0085** |

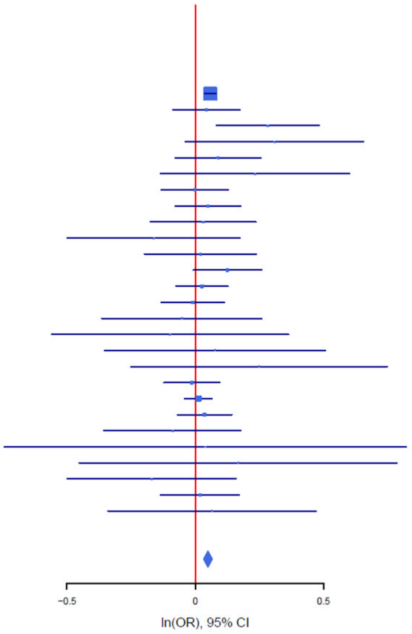

B

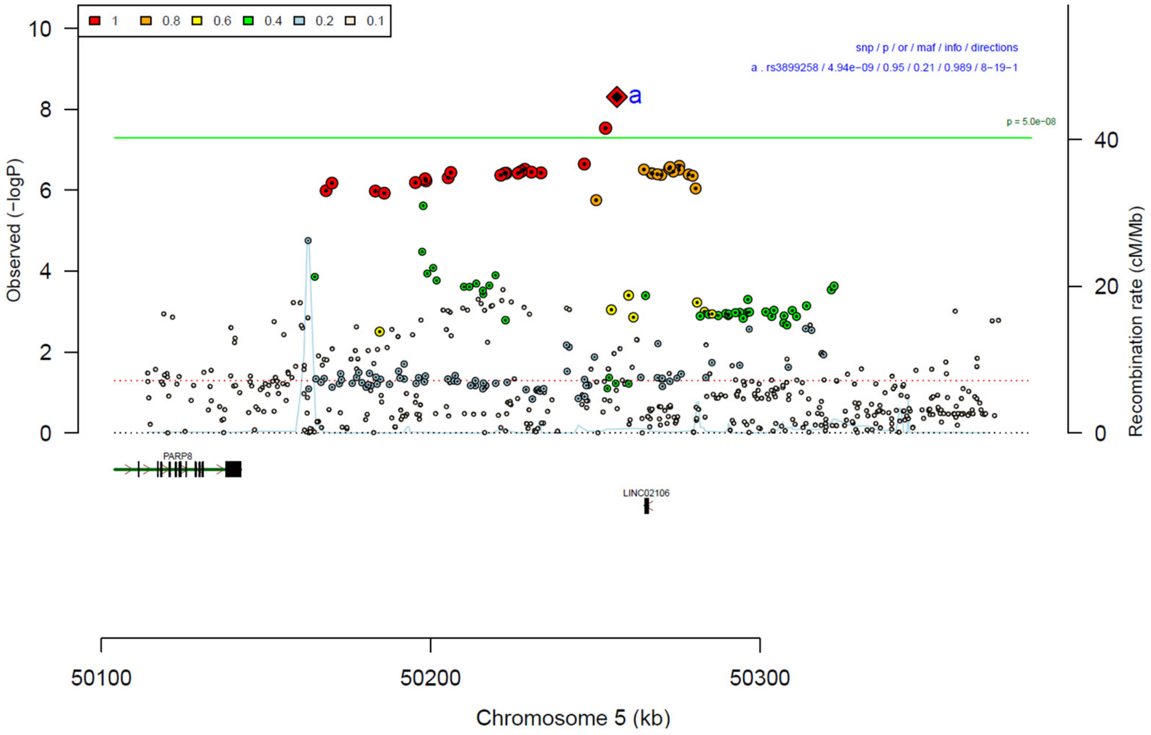

**Supplementary Figure 14: Forest plot (A) and regional association plot (B) of SNP rs3899258 in the meta-analysis.** The forest plot (A) shows the imputation quality (INFO) score, p-value of the SNP association (two-sided and not corrected for multiple testing, derived from a fixed-effects inverse-variance weighted meta-analysis), allele frequency in cases with case sample size (f_ca(n)), allele frequency in controls and control sample size (f_co(n)), beta estimates (ln(OR)) and standard error (STDerr) for each study as well as for the combined meta-analysis. Each cohort's sample size is detailed in supplementary table 1. Data on the right show the ln(OR) value as listed per cohort with 95% confidence intervals. In the regional association plot (B), the –log10 p-values (from two-sided association tests) are shown on the left y-axis. Recombination rates expressed in centimorgans (cM) per Mb (Megabase) (blue line) are shown on the right y-axis. Position in Mb is on the x-axis, with genes shown below the regional association plot. Only SNPs with an association p-value less than 0.1 were plotted. The most associated SNP is shown as a diamond and marked with the letter a.

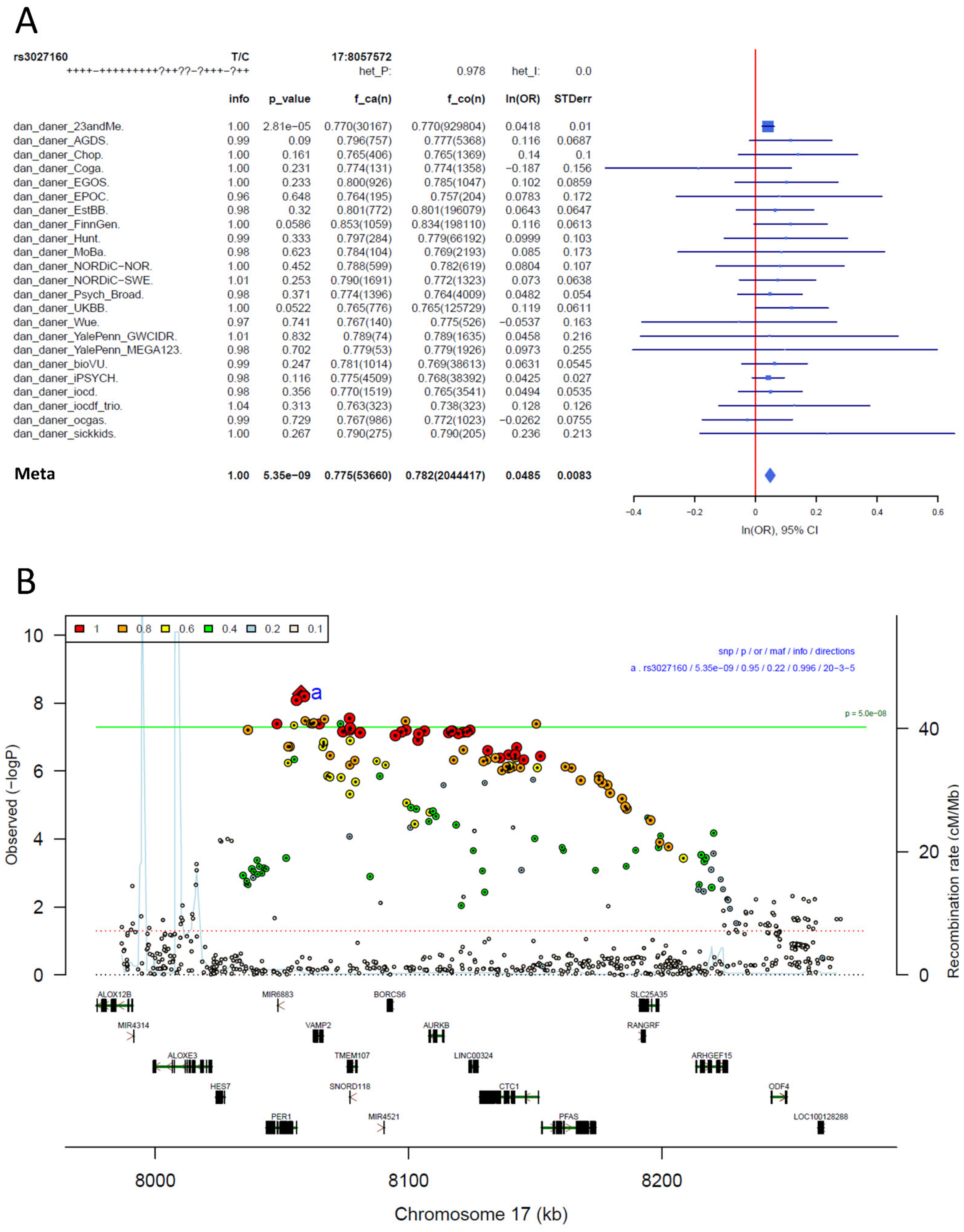

**Supplementary Figure 15: Forest plot (A) and regional association plot (B) of SNP rs3027160 in the meta-analysis.** The forest plot (A) shows the imputation quality (INFO) score, p-value of the SNP association (two-sided and not corrected for multiple testing, derived from a fixed-effects inverse-variance weighted meta-analysis), allele frequency in cases with case sample size (f_ca(n)), allele frequency in controls and control sample size (f_co(n)), beta estimates (ln(OR)) and standard error (STDerr) for each study as well as for the combined meta-analysis. Each cohort's sample size is detailed in supplementary table 1. Data on the right show the ln(OR) value as listed per cohort with 95% confidence intervals. In the regional association plot (B), the –log10 p-values (from two-sided association tests) are shown on the left y-axis. Recombination rates expressed in centimorgans (cM) per Mb (Megabase) (blue line) are shown on the right y-axis. Position in Mb is on the x-axis, with genes shown below the regional association plot. Only SNPs with an association p-value less than 0.1 were plotted. The most associated SNP is shown as a diamond and marked with the letter a.

A

rs203768 T/C 2:200901427

−+−−−−−−−−++−+−−−??−?−+−−−++ het_P: 0.335 het_I: 9.1

| | info | p_value | f_ca(n) | f_co(n) | ln(OR) | STDerr |
|---|---|---|---|---|---|---|
| dan_daner_23andMe. | 1.00 | 0.00069 | 0.831(30167) | 0.831(929804) | −0.0376 | 0.011 |
| dan_daner_AGDS. | 1.03 | 0.305 | 0.834(757) | 0.820(5368) | 0.0747 | 0.0729 |
| dan_daner_Coga. | 1.00 | 0.54 | 0.823(131) | 0.823(1358) | −0.11 | 0.179 |
| dan_daner_EGOS. | 0.96 | 0.77 | 0.808(926) | 0.804(1047) | −0.0263 | 0.0901 |
| dan_daner_EPOC. | 1.00 | 0.595 | 0.826(195) | 0.846(204) | −0.104 | 0.195 |
| dan_daner_EstBB. | 1.00 | 0.273 | 0.798(772) | 0.798(196079) | −0.0698 | 0.0636 |
| dan_daner_FinnGen. | 1.00 | 0.00262 | 0.713(1059) | 0.745(198110) | −0.155 | 0.0516 |
| dan_daner_Hunt. | 1.00 | 0.816 | 0.826(284) | 0.833(66192) | −0.0264 | 0.113 |
| dan_daner_MVP. | 0.97 | 0.00398 | 0.828(5129) | 0.828(422860) | −0.0755 | 0.0262 |
| dan_daner_MoBa. | 1.00 | 0.758 | 0.841(104) | 0.832(2193) | 0.0582 | 0.189 |
| dan_daner_NORDiC−NOR. | 1.00 | 0.00598 | 0.810(599) | 0.853(619) | −0.324 | 0.118 |
| dan_daner_NORDiC−SWE. | 1.00 | 0.603 | 0.803(1691) | 0.807(1323) | −0.0349 | 0.0671 |
| dan_daner_Psych_Broad. | 1.00 | 0.443 | 0.837(1396) | 0.841(4009) | −0.0468 | 0.061 |
| dan_daner_UKBB. | 1.00 | 0.962 | 0.817(776) | 0.817(125729) | −0.00315 | 0.0668 |
| dan_daner_Wue. | 0.98 | 0.919 | 0.843(140) | 0.841(526) | −0.0191 | 0.188 |
| dan_daner_YalePenn_EXHCE. | 1.07 | 0.519 | 0.825(52) | 0.825(1646) | −0.163 | 0.252 |
| dan_daner_YalePenn_GWCIDR. | 1.01 | 0.954 | 0.843(74) | 0.843(1635) | 0.0143 | 0.245 |
| dan_daner_YalePenn_MEGA123. | 0.99 | 0.248 | 0.848(53) | 0.848(1926) | 0.37 | 0.321 |
| dan_daner_bioVU. | 1.00 | 0.00366 | 0.789(1014) | 0.813(38613) | −0.17 | 0.0586 |
| dan_daner_iPSYCH. | 0.99 | 0.00916 | 0.812(4509) | 0.822(38392) | −0.0748 | 0.0287 |
| dan_daner_iocd. | 1.00 | 0.856 | 0.846(1519) | 0.835(3541) | 0.0111 | 0.0615 |
| dan_daner_iocdf_trio. | 1.00 | 0.484 | 0.852(323) | 0.837(323) | 0.108 | 0.153 |
| dan_daner_ocgas. | 1.00 | 0.305 | 0.841(986) | 0.853(1023) | −0.0903 | 0.0881 |
| dan_daner_sickkids. | 1.00 | 0.892 | 0.822(275) | 0.822(205) | 0.0323 | 0.238 |
| **Meta** | **0.99** | **6.14e−09** | **0.824(53254)** | **0.817(2043048)** | **−0.0499** | **0.0086** |

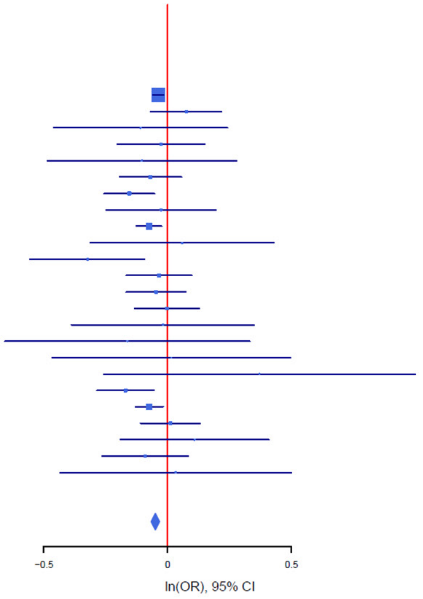

B

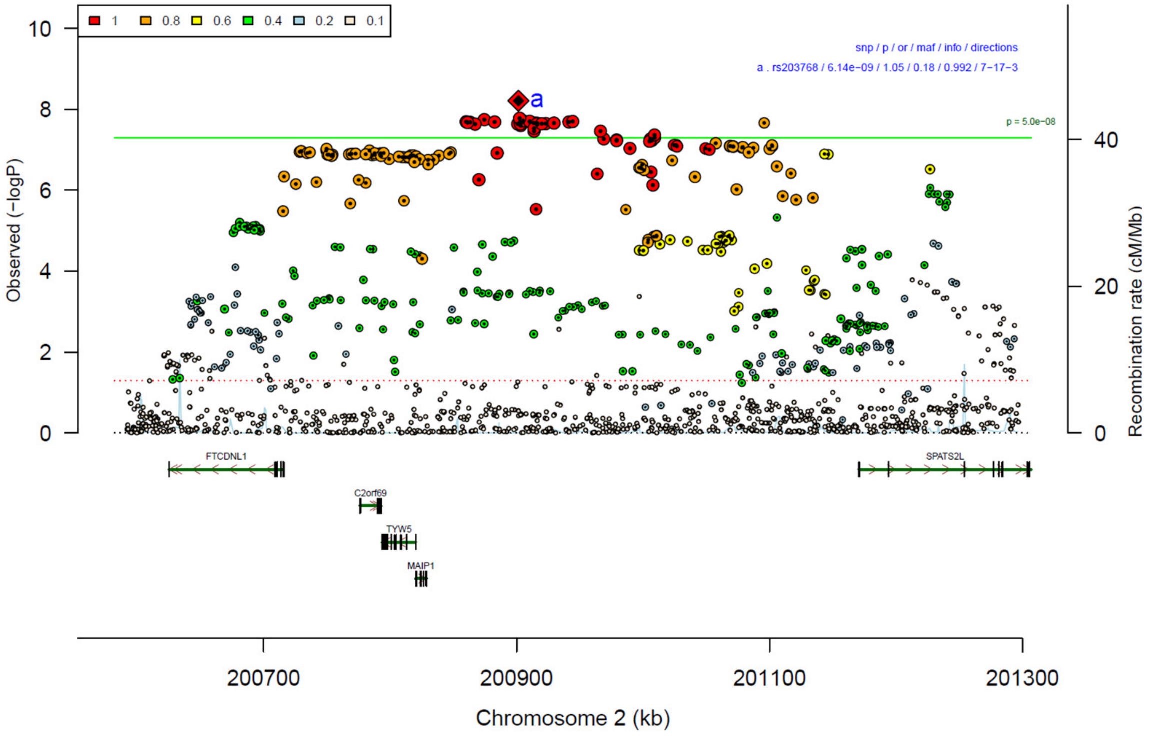

**Supplementary Figure 16: Forest plot (A) and regional association plot (B) of SNP rs203768 in the meta-analysis.** The forest plot (A) shows the imputation quality (INFO) score, p-value of the SNP association (two-sided and not corrected for multiple testing, derived from a fixed-effects inverse-variance weighted meta-analysis), allele frequency in cases with case sample size (f_ca(n)), allele frequency in controls and control sample size (f_co(n)), beta estimates (ln(OR)) and standard error (STDerr) for each study as well as for the combined meta-analysis. Each cohort's sample size is detailed in supplementary table 1. Data on the right show the ln(OR) value as listed per cohort with 95% confidence intervals. In the regional association plot (B), the –log10 p-values (from two-sided association tests) are shown on the left y-axis. Recombination rates expressed in centimorgans (cM) per Mb (Megabase) (blue line) are shown on the right y-axis. Position in Mb is on the x-axis, with genes shown below the regional association plot. Only SNPs with an association p-value less than 0.1 were plotted. The most associated SNP is shown as a diamond and marked with the letter a.

A

| rs11263940 | T/C | | 1:37182486 | | | |
|---|---|---|---|---|---|---|
| −++−+−−−+−−−−+?−−−−−−−−−−−−++ | | | het_P: | 0.138 | het_I: | 23.2 |
| | info | p_value | f_ca(n) | f_co(n) | ln(OR) | STDerr |
| dan_daner_23andMe. | 0.99 | 5.23e−07 | 0.687(30167) | 0.687(929804) | −0.0452 | 0.00898 |
| dan_daner_AGDS. | 0.93 | 0.674 | 0.687(757) | 0.683(5368) | 0.0261 | 0.0621 |
| dan_daner_Chop. | 1.00 | 0.204 | 0.689(406) | 0.689(1369) | −0.111 | 0.0876 |
| dan_daner_Coga. | 1.00 | 0.906 | 0.704(131) | 0.704(1358) | 0.0175 | 0.148 |
| dan_daner_EGOS. | 0.98 | 0.00834 | 0.699(926) | 0.739(1047) | −0.207 | 0.0786 |
| dan_daner_EPOC. | 0.93 | 0.236 | 0.648(195) | 0.686(204) | −0.189 | 0.16 |
| dan_daner_EstBB. | 0.98 | 0.292 | 0.684(772) | 0.684(196079) | −0.0585 | 0.0556 |
| dan_daner_FinnGen. | 1.00 | 0.899 | 0.680(1059) | 0.681(198110) | 0.0061 | 0.0479 |
| dan_daner_Hunt. | 1.00 | 0.688 | 0.736(284) | 0.744(66192) | −0.0393 | 0.0977 |
| dan_daner_MoBa. | 1.00 | 0.299 | 0.745(104) | 0.714(2193) | 0.168 | 0.162 |
| dan_daner_NORDiC−NOR. | 0.97 | 0.905 | 0.713(599) | 0.720(619) | −0.0117 | 0.0987 |
| dan_daner_NORDiC−SWE. | 0.99 | 0.555 | 0.719(1691) | 0.723(1323) | −0.0351 | 0.0594 |
| dan_daner_Psych_Broad. | 1.01 | 0.0249 | 0.647(1396) | 0.666(4009) | −0.106 | 0.0473 |
| dan_daner_UKBB. | 1.00 | 0.907 | 0.699(776) | 0.699(125729) | −0.00658 | 0.0564 |
| dan_daner_Wue. | 0.94 | 0.632 | 0.677(140) | 0.692(526) | −0.0716 | 0.149 |
| dan_daner_YalePenn_EXHCE. | 0.98 | 0.183 | 0.685(52) | 0.685(1646) | −0.294 | 0.221 |
| dan_daner_YalePenn_GWCIDR. | 1.01 | 0.933 | 0.686(74) | 0.686(1635) | 0.0158 | 0.189 |
| dan_daner_YalePenn_MEGA123. | 0.99 | 0.568 | 0.683(53) | 0.683(1926) | 0.128 | 0.224 |
| dan_daner_bioVU. | 1.00 | 0.88 | 0.685(1014) | 0.683(38613) | 0.00742 | 0.049 |
| dan_daner_iPSYCH. | 0.99 | 0.871 | 0.717(4509) | 0.718(38392) | −0.004 | 0.0248 |
| dan_daner_iocd. | 0.99 | 0.0444 | 0.673(1519) | 0.693(3541) | −0.0973 | 0.0484 |
| dan_daner_iocdf_trio. | 0.95 | 0.33 | 0.650(323) | 0.675(323) | −0.118 | 0.121 |
| dan_daner_ocgas−ab. | 0.83 | 0.000747 | 0.563(55) | 0.768(55) | −1.65 | 0.488 |
| dan_daner_ocgas−gh. | 0.82 | 0.372 | 0.692(56) | 0.743(56) | −0.307 | 0.344 |
| dan_daner_ocgas−nes. | 0.97 | 0.525 | 0.666(212) | 0.692(212) | −0.0967 | 0.152 |
| dan_daner_ocgas. | 0.99 | 0.426 | 0.674(986) | 0.687(1023) | −0.0544 | 0.0683 |
| dan_daner_sickkids. | 1.00 | 0.06 | 0.668(275) | 0.668(205) | −0.365 | 0.194 |
| **Meta** | **0.99** | **7.23e−09** | **0.689(53660)** | **0.690(2044417)** | **−0.0431** | **0.0074** |

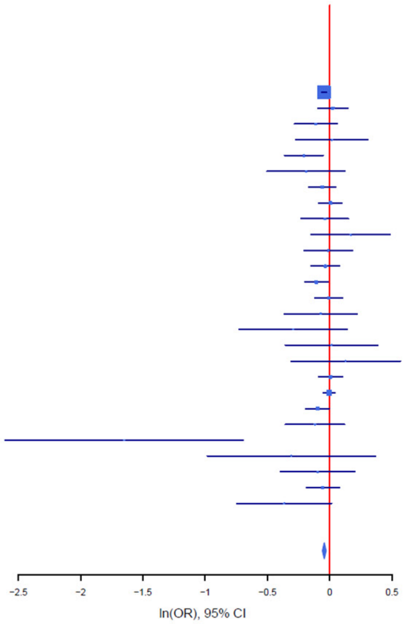

B

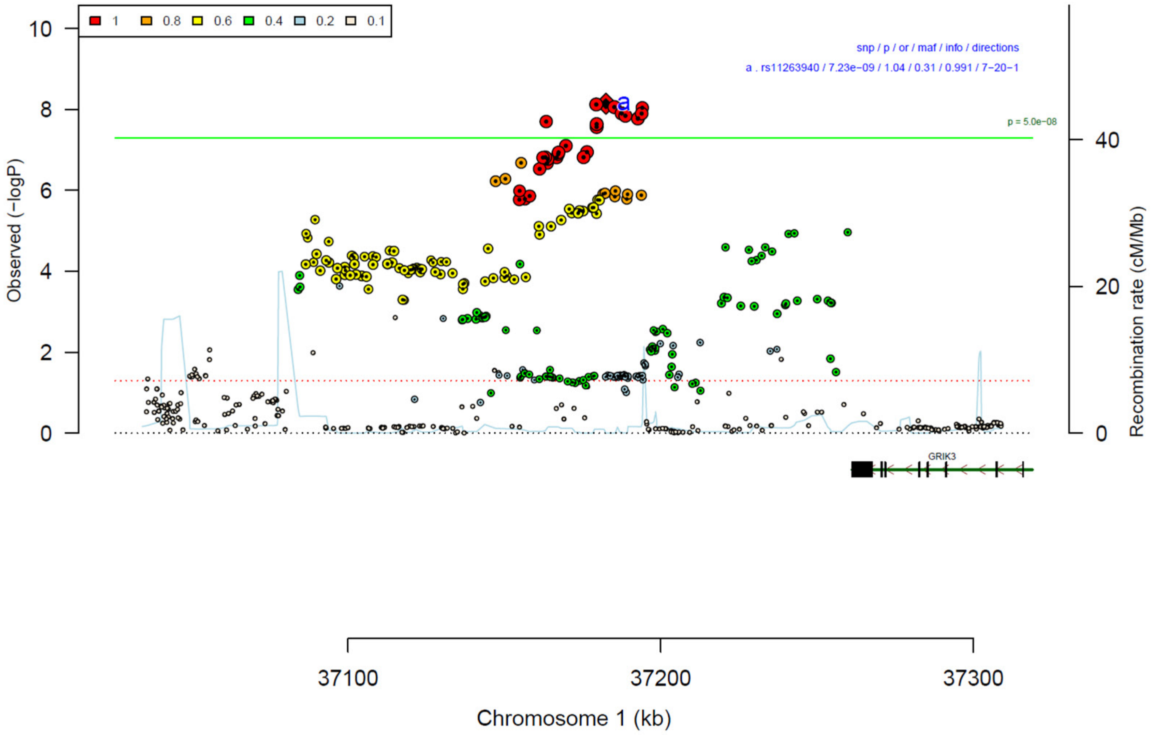

**Supplementary Figure 17: Forest plot (A) and regional association plot (B) of SNP rs11263940 in the meta-analysis.** The forest plot (A) shows the imputation quality (INFO) score, p-value of the SNP association (two-sided and not corrected for multiple testing, derived from a fixed-effects inverse-variance weighted meta-analysis), allele frequency in cases with case sample size (f_ca(n)), allele frequency in controls and control sample size (f_co(n)), beta estimates (ln(OR)) and standard error (STDerr) for each study as well as for the combined meta-analysis. Each cohort's sample size is detailed in supplementary table 1. Data on the right show the ln(OR) value as listed per cohort with 95% confidence intervals. In the regional association plot (B), the –log10 p-values (from two-sided association tests) are shown on the left y-axis. Recombination rates expressed in centimorgans (cM) per Mb (Megabase) (blue line) are shown on the right y-axis. Position in Mb is on the x-axis, with genes shown below the regional association plot. Only SNPs with an association p-value less than 0.1 were plotted. The most associated SNP is shown as a diamond and marked with the letter a.

A

rs67839857 G/A 5:91013369

--------?-+--------++++---+-++ het_P: 0.825 het_I: 0.0

| | info | p_value | f_ca(n) | f_co(n) | ln(OR) | STDerr |
|---|---|---|---|---|---|---|
| dan_daner_23andMe. | 1.00 | 3.3e−06 | 0.689(30167) | 0.689(929804) | 0.0422 | 0.0091 |
| dan_daner_AGDS. | 0.99 | 0.723 | 0.698(757) | 0.695(5368) | 0.0215 | 0.0607 |
| dan_daner_Chop. | 1.00 | 0.0724 | 0.698(406) | 0.698(1369) | 0.161 | 0.0893 |
| dan_daner_Coga. | 1.00 | 0.58 | 0.690(131) | 0.690(1358) | 0.0789 | 0.142 |
| dan_daner_EGOS. | 0.99 | 0.304 | 0.722(926) | 0.712(1047) | 0.0797 | 0.0776 |
| dan_daner_EPOC. | 0.99 | 0.308 | 0.698(195) | 0.665(204) | 0.159 | 0.157 |
| dan_daner_FinnGen. | 1.00 | 0.863 | 0.683(1059) | 0.683(198110) | 0.0083 | 0.0481 |
| dan_daner_Hunt. | 1.00 | 0.456 | 0.692(284) | 0.705(66192) | −0.0695 | 0.0933 |
| dan_daner_MVP. | 0.98 | 0.0388 | 0.697(5129) | 0.697(422860) | 0.0456 | 0.0221 |
| dan_daner_MoBa. | 1.00 | 0.598 | 0.721(104) | 0.701(2193) | 0.0819 | 0.155 |
| dan_daner_NORDiC−NOR. | 1.00 | 0.0792 | 0.739(599) | 0.696(619) | 0.171 | 0.0974 |
| dan_daner_NORDiC−SWE. | 0.98 | 0.278 | 0.707(1691) | 0.690(1323) | 0.0634 | 0.0584 |
| dan_daner_Psych_Broad. | 0.97 | 0.663 | 0.698(1396) | 0.701(4009) | 0.0217 | 0.0498 |
| dan_daner_UKBB. | 1.00 | 0.056 | 0.689(776) | 0.689(125729) | 0.107 | 0.0559 |
| dan_daner_Wue. | 0.99 | 0.22 | 0.654(140) | 0.690(526) | −0.177 | 0.144 |
| dan_daner_YalePenn_EXHCE. | 1.01 | 0.705 | 0.704(52) | 0.704(1646) | 0.0861 | 0.227 |
| dan_daner_YalePenn_GWCIDR. | 1.00 | 0.302 | 0.712(74) | 0.712(1635) | −0.194 | 0.188 |
| dan_daner_YalePenn_MEGA123. | 0.99 | 0.781 | 0.690(53) | 0.690(1926) | −0.0606 | 0.218 |
| dan_daner_bioVU. | 1.00 | 0.329 | 0.708(1014) | 0.697(38613) | 0.0485 | 0.0497 |
| dan_daner_iPSYCH. | 1.00 | 0.18 | 0.689(4509) | 0.682(38392) | 0.0323 | 0.0241 |
| dan_daner_iocd. | 0.96 | 0.153 | 0.691(1519) | 0.683(3541) | 0.0704 | 0.0492 |
| dan_daner_iocdf_trio. | 0.91 | 0.516 | 0.698(323) | 0.682(323) | 0.0818 | 0.126 |
| dan_daner_ocgas−ab. | 0.85 | 0.834 | 0.663(55) | 0.636(55) | −0.0769 | 0.367 |
| dan_daner_ocgas−gh. | 0.91 | 0.14 | 0.634(56) | 0.738(56) | −0.481 | 0.326 |
| dan_daner_ocgas−nes. | 1.00 | 0.203 | 0.670(212) | 0.707(212) | −0.19 | 0.15 |
| dan_daner_ocgas. | 0.97 | 0.6 | 0.684(986) | 0.691(1023) | −0.0364 | 0.0694 |
| dan_daner_sickkids. | 1.00 | 0.93 | 0.692(275) | 0.692(205) | 0.0164 | 0.187 |
| **Meta** | **0.99** | **7.63e−09** | **0.692(53660)** | **0.691(2044417)** | **0.0414** | **0.0072** |

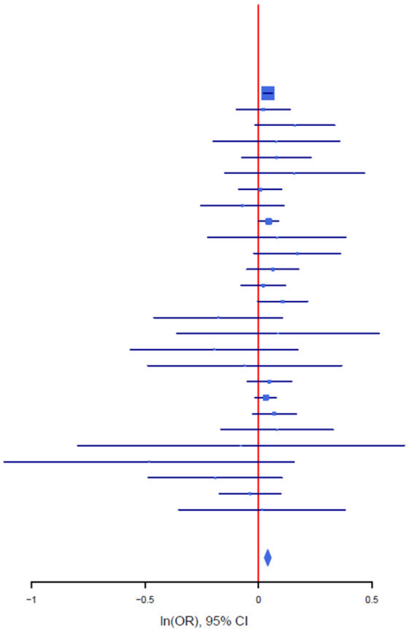

B

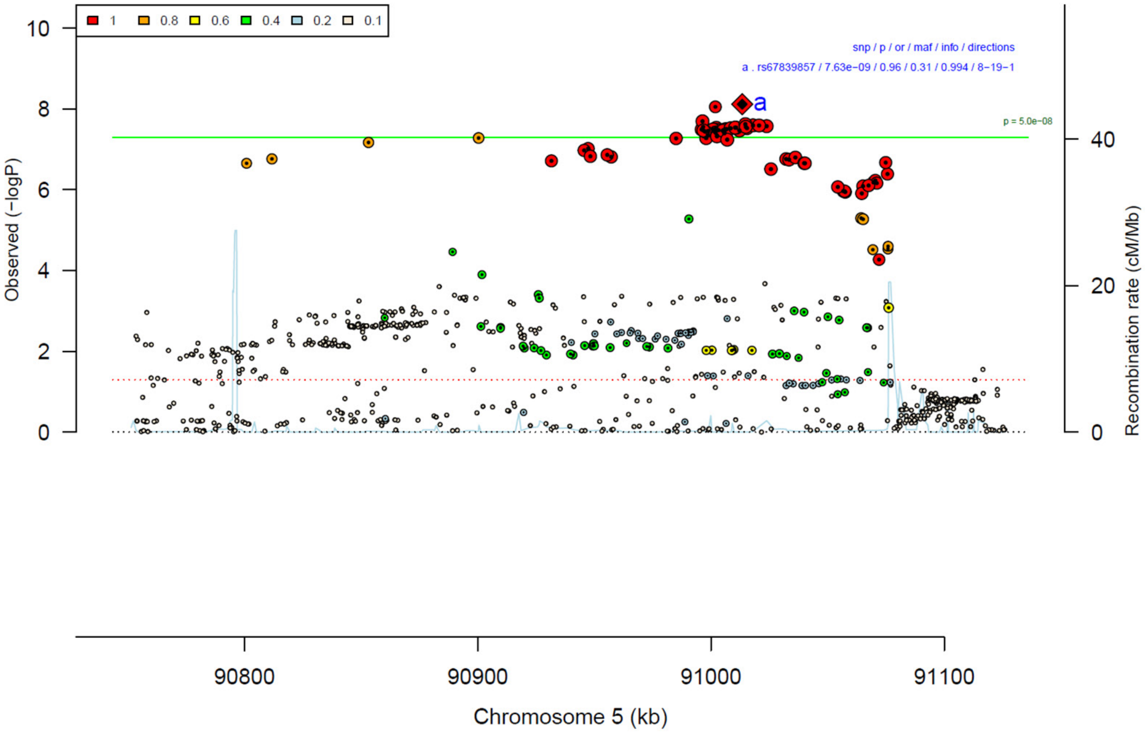

**Supplementary Figure 18: Forest plot (A) and regional association plot (B) of SNP rs67839857 in the meta-analysis.** The forest plot (A) shows the imputation quality (INFO) score, p-value of the SNP association (two-sided and not corrected for multiple testing, derived from a fixed-effects inverse-variance weighted meta-analysis), allele frequency in cases with case sample size (f_ca(n)), allele frequency in controls and control sample size (f_co(n)), beta estimates (ln(OR)) and standard error (STDerr) for each study as well as for the combined meta-analysis. Each cohort's sample size is detailed in supplementary table 1. Data on the right show the ln(OR) value as listed per cohort with 95% confidence intervals. In the regional association plot (B), the –log10 p-values (from two-sided association tests) are shown on the left y-axis. Recombination rates expressed in centimorgans (cM) per Mb (Megabase) (blue line) are shown on the right y-axis. Position in Mb is on the x-axis, with genes shown below the regional association plot. Only SNPs with an association p-value less than 0.1 were plotted. The most associated SNP is shown as a diamond and marked with the letter a.

A

| rs1555466 | T/C | | 20:20965369 | | | |
|---|---|---|---|---|---|---|
| | +++++---++++++?+++++-++--?++ | | het_P: | 0.269 | het_I: | 13.4 |
| | info | p_value | f_ca(n) | f_co(n) | ln(OR) | STDerr |
| dan_daner_23andMe. | 0.99 | 1.02e−06 | 0.213(30167) | 0.213(929804) | 0.0496 | 0.0101 |
| dan_daner_AGDS. | 0.99 | 0.532 | 0.233(757) | 0.224(5368) | 0.0412 | 0.0659 |
| dan_daner_Chop. | 1.00 | 0.209 | 0.215(406) | 0.215(1369) | 0.124 | 0.0985 |
| dan_daner_Coga. | 1.00 | 0.101 | 0.207(131) | 0.207(1358) | 0.262 | 0.16 |
| dan_daner_EGOS. | 1.00 | 0.112 | 0.218(926) | 0.244(1047) | −0.133 | 0.0833 |
| dan_daner_EPOC. | 1.00 | 0.488 | 0.208(195) | 0.233(204) | −0.116 | 0.168 |
| dan_daner_EstBB. | 1.00 | 0.187 | 0.241(772) | 0.241(196079) | −0.0794 | 0.0602 |
| dan_daner_FinnGen. | 1.00 | 0.333 | 0.277(1059) | 0.263(198110) | 0.0494 | 0.051 |
| dan_daner_Hunt. | 0.99 | 0.151 | 0.251(284) | 0.227(66192) | 0.146 | 0.101 |
| dan_daner_MoBa. | 0.98 | 0.369 | 0.245(104) | 0.221(2193) | 0.157 | 0.175 |
| dan_daner_NORDiC−NOR. | 0.99 | 0.0627 | 0.232(599) | 0.198(619) | 0.2 | 0.107 |
| dan_daner_NORDiC−SWE. | 1.00 | 0.386 | 0.228(1691) | 0.224(1323) | 0.0551 | 0.0634 |
| dan_daner_Psych_Broad. | 1.01 | 0.0778 | 0.212(1396) | 0.195(4009) | 0.097 | 0.0551 |
| dan_daner_UKBB. | 1.00 | 0.78 | 0.215(776) | 0.215(125729) | −0.0177 | 0.0633 |
| dan_daner_Wue. | 1.11 | 0.644 | 0.189(140) | 0.201(526) | −0.0758 | 0.164 |
| dan_daner_YalePenn_GWCIDR. | 1.02 | 0.361 | 0.209(74) | 0.209(1635) | 0.191 | 0.209 |
| dan_daner_YalePenn_MEGA123. | 1.02 | 0.765 | 0.213(53) | 0.213(1926) | 0.0717 | 0.24 |
| dan_daner_bioVU. | 1.00 | 0.663 | 0.212(1014) | 0.208(38613) | 0.0244 | 0.056 |
| dan_daner_iPSYCH. | 0.99 | 0.255 | 0.228(4509) | 0.223(38392) | 0.0303 | 0.0266 |
| dan_daner_iocd. | 0.99 | 0.199 | 0.215(1519) | 0.210(3541) | 0.0701 | 0.0546 |
| dan_daner_iocdf_trio. | 1.05 | 0.0341 | 0.216(323) | 0.169(323) | 0.296 | 0.139 |
| dan_daner_ocgas−ab. | 0.91 | 0.806 | 0.259(55) | 0.231(55) | 0.0957 | 0.389 |
| dan_daner_ocgas−gh. | 0.80 | 0.0973 | 0.229(56) | 0.146(56) | 0.671 | 0.405 |
| dan_daner_ocgas−nes. | 0.91 | 0.464 | 0.207(212) | 0.224(212) | −0.129 | 0.176 |
| dan_daner_ocgas. | 1.02 | 0.073 | 0.237(986) | 0.214(1023) | 0.135 | 0.0751 |
| dan_daner_sickkids. | 1.00 | 0.157 | 0.217(275) | 0.217(205) | 0.295 | 0.208 |
| **Meta** | **1.00** | **8.42e−09** | **0.218(53660)** | **0.224(2044417)** | **0.0478** | **0.0083** |

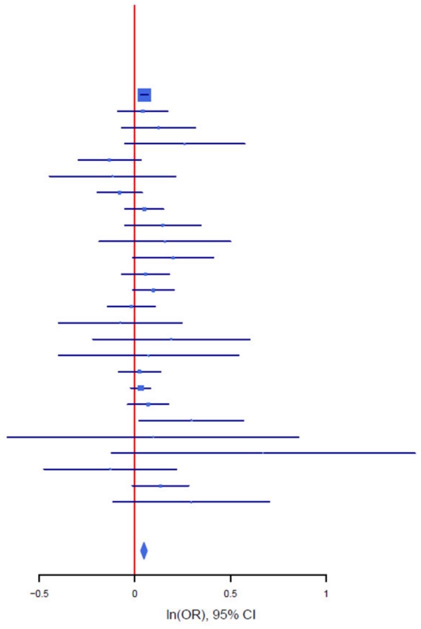

B

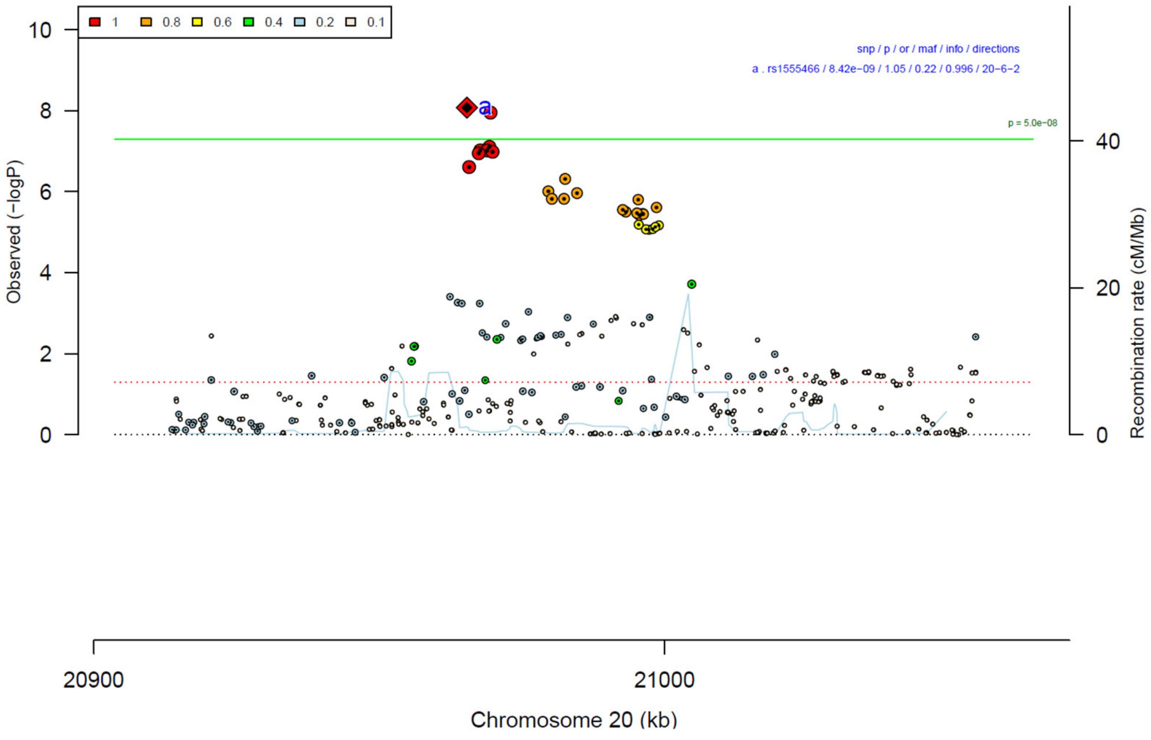

**Supplementary Figure 19: Forest plot (A) and regional association plot (B) of SNP rs1555466 in the meta-analysis.** The forest plot (A) shows the imputation quality (INFO) score, p-value of the SNP association (two-sided and not corrected for multiple testing, derived from a fixed-effects inverse-variance weighted meta-analysis), allele frequency in cases with case sample size (f_ca(n)), allele frequency in controls and control sample size (f_co(n)), beta estimates (ln(OR)) and standard error (STDerr) for each study as well as for the combined meta-analysis. Each cohort's sample size is detailed in supplementary table 1. Data on the right show the ln(OR) value as listed per cohort with 95% confidence intervals. In the regional association plot (B), the –log10 p-values (from two-sided association tests) are shown on the left y-axis. Recombination rates expressed in centimorgans (cM) per Mb (Megabase) (blue line) are shown on the right y-axis. Position in Mb is on the x-axis, with genes shown below the regional association plot. Only SNPs with an association p-value less than 0.1 were plotted. The most associated SNP is shown as a diamond and marked with the letter a.

A

| rs9886111 | G/C | | 7:88418451 | | | |
|---|---|---|---|---|---|---|
| +++−+++++−+++++−+??+?+−++−+− | | | het_P: | 0.0274 | het_I: | 38.4 |
| | info | p_value | f_ca(n) | f_co(n) | ln(OR) | STDerr |
| dan_daner_23andMe. | 1.00 | 0.000426 | 0.702(30167) | 0.702(929804) | −0.0322 | 0.00911 |
| dan_daner_AGDS. | 1.00 | 0.953 | 0.704(757) | 0.708(5368) | −0.00359 | 0.0609 |
| dan_daner_Chop. | 1.00 | 0.556 | 0.699(406) | 0.699(1369) | 0.0533 | 0.0905 |
| dan_daner_Coga. | 1.00 | 0.323 | 0.703(131) | 0.703(1358) | −0.146 | 0.148 |
| dan_daner_EGOS. | 0.96 | 0.0279 | 0.691(926) | 0.711(1047) | −0.172 | 0.0785 |
| dan_daner_EPOC. | 1.00 | 0.753 | 0.712(195) | 0.724(204) | −0.0522 | 0.166 |
| dan_daner_EstBB. | 1.00 | 0.0275 | 0.743(772) | 0.743(196079) | −0.126 | 0.057 |
| dan_daner_FinnGen. | 1.00 | 0.941 | 0.723(1059) | 0.728(198110) | −0.0037 | 0.0501 |
| dan_daner_Hunt. | 1.00 | 0.845 | 0.713(284) | 0.710(66192) | 0.0181 | 0.0929 |
| dan_daner_MVP. | 0.97 | 0.171 | 0.708(5129) | 0.708(422860) | −0.0303 | 0.0221 |
| dan_daner_MoBa. | 0.99 | 0.157 | 0.663(104) | 0.713(2193) | −0.226 | 0.159 |
| dan_daner_NORDiC−NOR. | 0.96 | 0.414 | 0.724(599) | 0.700(619) | 0.0804 | 0.0985 |
| dan_daner_NORDiC−SWE. | 0.99 | 0.94 | 0.691(1691) | 0.696(1323) | −0.00441 | 0.0579 |
| dan_daner_Psych_Broad. | 1.02 | 0.641 | 0.676(1396) | 0.681(4009) | −0.0221 | 0.0476 |
| dan_daner_UKBB. | 1.00 | 0.49 | 0.714(776) | 0.714(125729) | −0.0396 | 0.0574 |
| dan_daner_Wue. | 0.99 | 0.721 | 0.700(140) | 0.709(526) | −0.053 | 0.148 |
| dan_daner_YalePenn_EXHCE. | 1.01 | 0.698 | 0.685(52) | 0.685(1646) | 0.087 | 0.224 |
| dan_daner_YalePenn_GWCIDR. | 0.98 | 0.788 | 0.701(74) | 0.701(1635) | −0.0525 | 0.195 |
| dan_daner_YalePenn_MEGA123. | 1.02 | 0.301 | 0.701(53) | 0.701(1926) | 0.243 | 0.235 |
| dan_daner_bioVU. | 1.00 | 0.47 | 0.703(1014) | 0.711(38613) | −0.0364 | 0.0504 |
| dan_daner_iPSYCH. | 0.99 | 1.18e−06 | 0.697(4509) | 0.721(38392) | −0.119 | 0.0244 |
| dan_daner_iocd. | 1.01 | 0.313 | 0.679(1519) | 0.695(3541) | −0.0484 | 0.0479 |
| dan_daner_iocdf_trio. | 1.08 | 0.000566 | 0.644(323) | 0.737(323) | −0.408 | 0.118 |
| dan_daner_ocgas. | 0.99 | 0.354 | 0.680(986) | 0.693(1023) | −0.0637 | 0.0687 |
| dan_daner_sickkids. | 1.00 | 0.0484 | 0.665(275) | 0.665(205) | 0.389 | 0.197 |
| **Meta** | **0.99** | **8.59e−09** | **0.701(53660)** | **0.711(2044417)** | **−0.041** | **0.0071** |

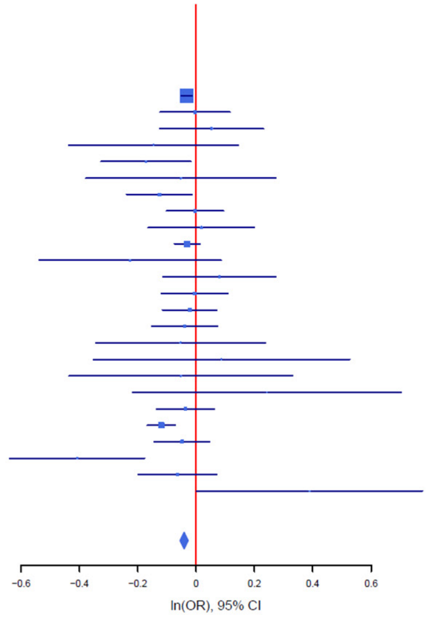

B

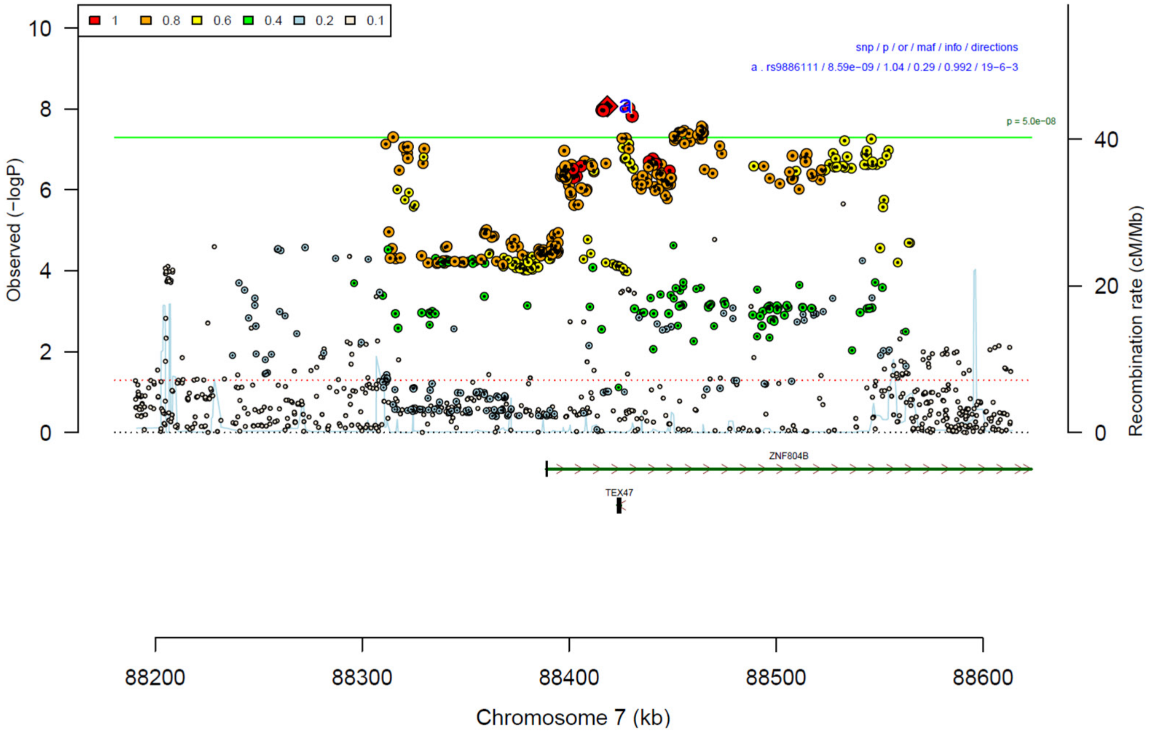

**Supplementary Figure 20: Forest plot (A) and regional association plot (B) of SNP rs9886111 in the meta-analysis.** The forest plot (A) shows the imputation quality (INFO) score, p-value of the SNP association (two-sided and not corrected for multiple testing, derived from a fixed-effects inverse-variance weighted meta-analysis), allele frequency in cases with case sample size (f_ca(n)), allele frequency in controls and control sample size (f_co(n)), beta estimates (ln(OR)) and standard error (STDerr) for each study as well as for the combined meta-analysis. Each cohort's sample size is detailed in supplementary table 1. Data on the right show the ln(OR) value as listed per cohort with 95% confidence intervals. In the regional association plot (B), the –log10 p-values (from two-sided association tests) are shown on the left y-axis. Recombination rates expressed in centimorgans (cM) per Mb (Megabase) (blue line) are shown on the right y-axis. Position in Mb is on the x-axis, with genes shown below the regional association plot. Only SNPs with an association p-value less than 0.1 were plotted. The most associated SNP is shown as a diamond and marked with the letter a.

A

rs9287859 G/A 2:166450259
++?++-++--++++?++-++-++-+?-? het_P: 0.548 het_I: 0.0

| | info | p_value | f_ca(n) | f_co(n) | ln(OR) | STDerr |
|---|---|---|---|---|---|---|
| dan_daner_23andMe. | 1.00 | 5.47e−05 | 0.391(30167) | 0.391(929804) | −0.0348 | 0.00863 |
| dan_daner_AGDS. | 0.98 | 0.0152 | 0.356(757) | 0.390(5368) | −0.142 | 0.0583 |
| dan_daner_Chop. | 1.00 | 0.0768 | 0.383(406) | 0.383(1369) | −0.149 | 0.0839 |
| dan_daner_Coga. | 1.00 | 0.917 | 0.391(131) | 0.391(1358) | −0.0143 | 0.138 |
| dan_daner_EGOS. | 0.96 | 0.484 | 0.404(926) | 0.401(1047) | 0.0507 | 0.0725 |
| dan_daner_EPOC. | 0.92 | 0.447 | 0.326(195) | 0.362(204) | −0.118 | 0.156 |
| dan_daner_EstBB. | 0.99 | 0.266 | 0.368(772) | 0.368(196079) | −0.0594 | 0.0535 |
| dan_daner_FinnGen. | 1.00 | 0.497 | 0.407(1059) | 0.401(198110) | 0.0313 | 0.0461 |
| dan_daner_Hunt. | 0.99 | 0.576 | 0.412(284) | 0.400(66192) | 0.0484 | 0.0865 |
| dan_daner_MoBa. | 0.97 | 0.708 | 0.404(104) | 0.416(2193) | −0.054 | 0.144 |
| dan_daner_NORDiC−NOR. | 0.92 | 0.271 | 0.400(599) | 0.425(619) | −0.102 | 0.0926 |
| dan_daner_NORDiC−SWE. | 1.00 | 0.0371 | 0.396(1691) | 0.422(1323) | −0.112 | 0.0539 |
| dan_daner_Psych_Broad. | 0.96 | 0.0383 | 0.367(1396) | 0.382(4009) | −0.0983 | 0.0475 |
| dan_daner_UKBB. | 0.99 | 0.727 | 0.390(776) | 0.390(125729) | 0.0186 | 0.0533 |
| dan_daner_Wue. | 0.96 | 0.548 | 0.375(140) | 0.396(526) | −0.0855 | 0.142 |
| dan_daner_YalePenn_GWCIDR. | 1.01 | 0.538 | 0.392(74) | 0.392(1635) | 0.111 | 0.179 |
| dan_daner_iPSYCH. | 0.98 | 0.000976 | 0.403(4509) | 0.421(38392) | −0.0757 | 0.023 |
| dan_daner_iocd. | 1.01 | 0.398 | 0.383(1519) | 0.389(3541) | −0.0386 | 0.0457 |
| dan_daner_iocdf_trio. | 0.94 | 0.078 | 0.367(323) | 0.413(323) | −0.208 | 0.118 |
| dan_daner_ocgas−ab. | 0.81 | 0.807 | 0.378(55) | 0.363(55) | 0.088 | 0.36 |
| dan_daner_ocgas−gh. | 0.80 | 0.913 | 0.382(56) | 0.398(56) | −0.0344 | 0.315 |
| dan_daner_ocgas−nes. | 0.80 | 0.709 | 0.386(212) | 0.377(212) | 0.0594 | 0.159 |
| dan_daner_ocgas. | 1.03 | 0.884 | 0.383(986) | 0.385(1023) | −0.00936 | 0.0643 |
| dan_daner_sickkids. | 1.00 | 0.553 | 0.380(275) | 0.380(205) | −0.107 | 0.18 |
| **Meta** | **0.99** | **9.83e−09** | **0.390(53660)** | **0.390(2044417)** | **−0.0413** | **0.0072** |

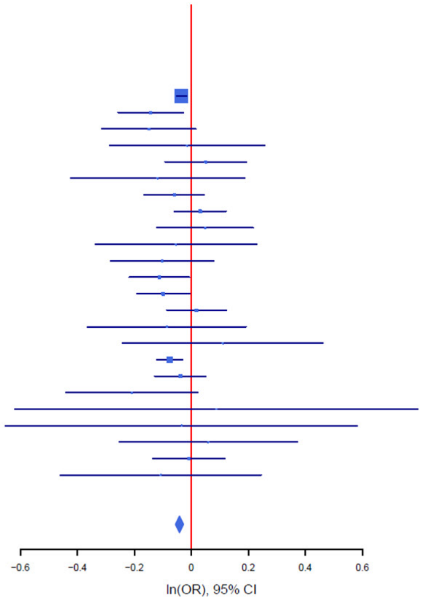

B

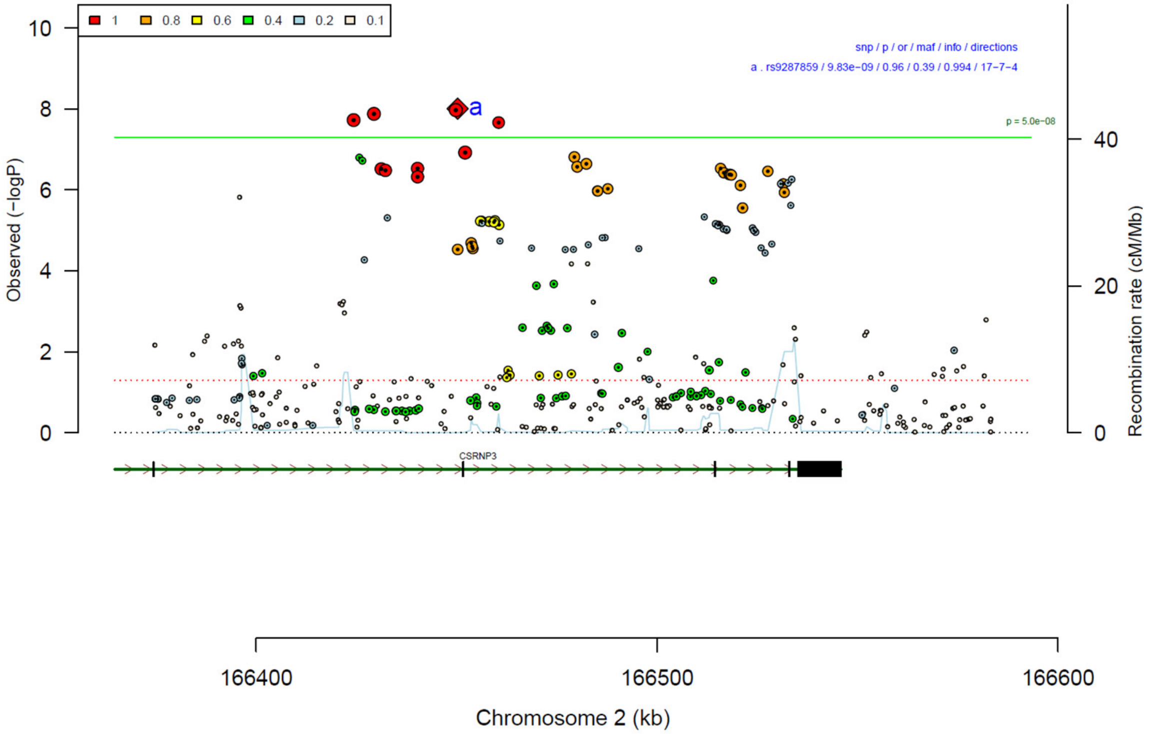

**Supplementary Figure 21: Forest plot (A) and regional association plot (B) of SNP rs9287859 in the meta-analysis.** The forest plot (A) shows the imputation quality (INFO) score, p-value of the SNP association (two-sided and not corrected for multiple testing, derived from a fixed-effects inverse-variance weighted meta-analysis), allele frequency in cases with case sample size (f_ca(n)), allele frequency in controls and control sample size (f_co(n)), beta estimates (ln(OR)) and standard error (STDerr) for each study as well as for the combined meta-analysis. Each cohort's sample size is detailed in supplementary table 1. Data on the right show the ln(OR) value as listed per cohort with 95% confidence intervals. In the regional association plot (B), the –log10 p-values (from two-sided association tests) are shown on the left y-axis. Recombination rates expressed in centimorgans (cM) per Mb (Megabase) (blue line) are shown on the right y-axis. Position in Mb is on the x-axis, with genes shown below the regional association plot. Only SNPs with an association p-value less than 0.1 were plotted. The most associated SNP is shown as a diamond and marked with the letter a.

A

rs2087319 C/A 4:56461365

++?+++-+-++-+-+++-++++--?--+ het_P: 0.596 het_I: 0.0

| | info | p_value | f_ca(n) | f_co(n) | ln(OR) | STDerr |
|---|---|---|---|---|---|---|
| dan_daner_23andMe. | 0.97 | 1.94e−06 | 0.740(30167) | 0.740(929804) | −0.0459 | 0.00962 |
| dan_daner_AGDS. | 0.94 | 0.751 | 0.744(757) | 0.749(5368) | −0.0208 | 0.0656 |
| dan_daner_Chop. | 1.00 | 0.897 | 0.725(406) | 0.725(1369) | −0.0124 | 0.0952 |
| dan_daner_Coga. | 1.00 | 0.269 | 0.762(131) | 0.762(1358) | −0.175 | 0.159 |
| dan_daner_EGOS. | 0.98 | 0.021 | 0.725(926) | 0.759(1047) | −0.186 | 0.0807 |
| dan_daner_EPOC. | 0.80 | 0.323 | 0.757(195) | 0.731(204) | 0.183 | 0.186 |
| dan_daner_EstBB. | 0.98 | 0.155 | 0.742(772) | 0.742(196079) | −0.0842 | 0.0592 |
| dan_daner_FinnGen. | 1.00 | 0.871 | 0.745(1059) | 0.744(198110) | 0.0084 | 0.0515 |
| dan_daner_Hunt. | 1.00 | 0.158 | 0.706(284) | 0.736(66192) | −0.136 | 0.0962 |
| dan_daner_MVP. | 0.93 | 0.0586 | 0.753(5129) | 0.753(422860) | −0.0448 | 0.0237 |
| dan_daner_MoBa. | 0.99 | 0.248 | 0.779(104) | 0.741(2193) | 0.192 | 0.166 |
| dan_daner_NORDiC−NOR. | 0.92 | 0.695 | 0.744(599) | 0.754(619) | −0.0411 | 0.105 |
| dan_daner_NORDiC−SWE. | 0.95 | 0.452 | 0.737(1691) | 0.750(1323) | −0.0469 | 0.0624 |
| dan_daner_Psych_Broad. | 0.97 | 0.245 | 0.739(1396) | 0.757(4009) | −0.061 | 0.0526 |
| dan_daner_UKBB. | 0.99 | 0.321 | 0.753(776) | 0.753(125729) | 0.0595 | 0.0599 |
| dan_daner_YalePenn_EXHCE. | 1.01 | 0.252 | 0.746(52) | 0.746(1646) | 0.28 | 0.244 |
| dan_daner_YalePenn_GWCIDR. | 1.01 | 0.544 | 0.754(74) | 0.754(1635) | 0.128 | 0.21 |
| dan_daner_YalePenn_MEGA123. | 0.97 | 0.249 | 0.747(53) | 0.747(1926) | −0.27 | 0.234 |
| dan_daner_iPSYCH. | 0.97 | 0.0742 | 0.736(4509) | 0.745(38392) | −0.0457 | 0.0256 |
| dan_daner_iocd. | 0.98 | 0.834 | 0.744(1519) | 0.740(3541) | 0.0108 | 0.0518 |
| dan_daner_iocdf_trio. | 0.96 | 0.475 | 0.713(323) | 0.731(323) | −0.0907 | 0.127 |
| dan_daner_ocgas−ab. | 0.90 | 0.783 | 0.776(55) | 0.748(55) | 0.108 | 0.391 |
| dan_daner_ocgas−gh. | 0.96 | 0.129 | 0.701(56) | 0.786(56) | −0.496 | 0.327 |
| dan_daner_ocgas−nes. | 0.92 | 0.418 | 0.684(212) | 0.713(212) | −0.127 | 0.157 |
| dan_daner_ocgas. | 0.99 | 0.511 | 0.725(986) | 0.732(1023) | −0.0474 | 0.0722 |
| dan_daner_sickkids. | 1.00 | 0.161 | 0.734(275) | 0.734(205) | 0.271 | 0.194 |
| **Meta** | **0.97** | **1.59e−08** | **0.740(53660)** | **0.744(2044417)** | **−0.043** | **0.0076** |

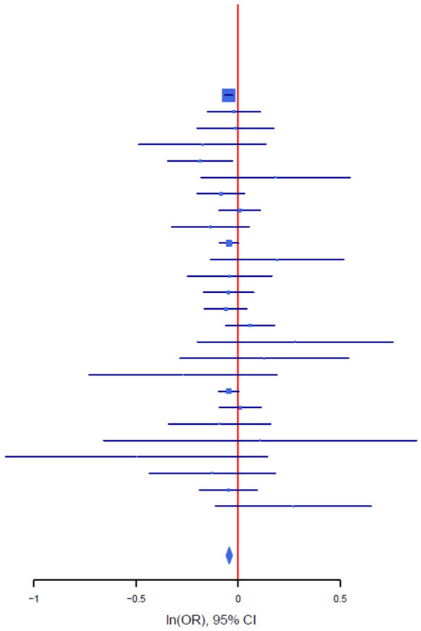

B

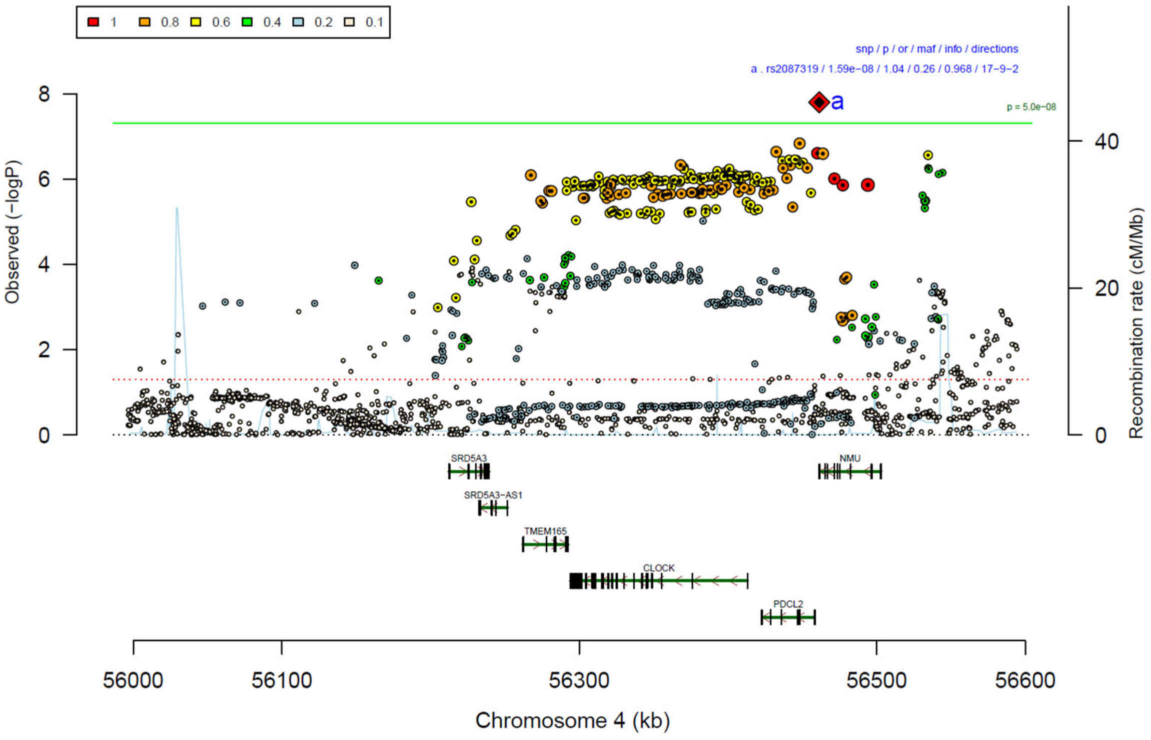

**Supplementary Figure 22: Forest plot (A) and regional association plot (B) of SNP rs2087319 in the meta-analysis.** The forest plot (A) shows the imputation quality (INFO) score, p-value of the SNP association (two-sided and not corrected for multiple testing, derived from a fixed-effects inverse-variance weighted meta-analysis), allele frequency in cases with case sample size (f_ca(n)), allele frequency in controls and control sample size (f_co(n)), beta estimates (ln(OR)) and standard error (STDerr) for each study as well as for the combined meta-analysis. Each cohort's sample size is detailed in supplementary table 1. Data on the right show the ln(OR) value as listed per cohort with 95% confidence intervals. In the regional association plot (B), the –log10 p-values (from two-sided association tests) are shown on the left y-axis. Recombination rates expressed in centimorgans (cM) per Mb (Megabase) (blue line) are shown on the right y-axis. Position in Mb is on the x-axis, with genes shown below the regional association plot. Only SNPs with an association p-value less than 0.1 were plotted. The most associated SNP is shown as a diamond and marked with the letter a.

A

| rs11125759 | G/A | | 2:58913399 | | | |
|---|---|---|---|---|---|---|
| ++?+--+-+++++-?++-++++++++++ | | | het_P: | 0.882 | het_I: | 0.0 |
| | **info** | **p_value** | **f_ca(n)** | **f_co(n)** | **ln(OR)** | **STDerr** |
| dan_daner_23andMe. | 0.99 | 1.69e−05 | 0.573(30167) | 0.573(929804) | −0.0366 | 0.00849 |
| dan_daner_AGDS. | 0.97 | 0.541 | 0.560(757) | 0.569(5368) | −0.0347 | 0.0568 |
| dan_daner_Chop. | 1.00 | 0.573 | 0.564(406) | 0.564(1369) | −0.0466 | 0.0828 |
| dan_daner_Coga. | 1.00 | 0.856 | 0.593(131) | 0.593(1358) | 0.0262 | 0.144 |
| dan_daner_EGOS. | 1.04 | 0.144 | 0.595(926) | 0.560(1047) | 0.101 | 0.0694 |
| dan_daner_EPOC. | 0.92 | 0.564 | 0.543(195) | 0.562(204) | −0.0869 | 0.15 |
| dan_daner_EstBB. | 0.97 | 0.349 | 0.510(772) | 0.510(196079) | 0.0488 | 0.0521 |
| dan_daner_FinnGen. | 1.00 | 0.482 | 0.481(1059) | 0.493(198110) | −0.0317 | 0.0451 |
| dan_daner_Hunt. | 1.00 | 0.452 | 0.567(284) | 0.588(66192) | −0.0646 | 0.086 |
| dan_daner_MoBa. | 1.00 | 0.317 | 0.611(104) | 0.578(2193) | 0.145 | 0.145 |
| dan_daner_NORDiC−NOR. | 1.00 | 0.0818 | 0.553(599) | 0.586(619) | −0.153 | 0.0877 |
| dan_daner_NORDiC−SWE. | 1.00 | 0.0203 | 0.560(1691) | 0.587(1323) | −0.125 | 0.054 |
| dan_daner_Psych_Broad. | 1.00 | 0.249 | 0.569(1396) | 0.582(4009) | −0.0522 | 0.0453 |
| dan_daner_UKBB. | 0.99 | 0.197 | 0.576(776) | 0.576(125729) | −0.0677 | 0.0525 |
| dan_daner_Wue. | 0.91 | 0.557 | 0.541(140) | 0.557(526) | −0.0839 | 0.143 |
| dan_daner_YalePenn_EXHCE. | 0.96 | 0.315 | 0.560(52) | 0.560(1646) | −0.214 | 0.213 |
| dan_daner_YalePenn_GWCIDR. | 1.02 | 0.615 | 0.559(74) | 0.559(1635) | −0.0886 | 0.176 |
| dan_daner_YalePenn_MEGA123. | 0.99 | 0.573 | 0.570(53) | 0.570(1926) | −0.116 | 0.205 |
| dan_daner_iPSYCH. | 0.99 | 0.0353 | 0.579(4509) | 0.591(38392) | −0.0477 | 0.0227 |
| dan_daner_iocd. | 1.00 | 0.0668 | 0.554(1519) | 0.573(3541) | −0.0826 | 0.0451 |
| dan_daner_iocdf_trio. | 0.96 | 0.238 | 0.542(323) | 0.574(323) | −0.135 | 0.115 |
| dan_daner_ocgas−ab. | 0.91 | 0.653 | 0.597(55) | 0.585(55) | 0.155 | 0.344 |
| dan_daner_ocgas−gh. | 0.96 | 0.633 | 0.567(56) | 0.605(56) | −0.138 | 0.29 |
| dan_daner_ocgas−nes. | 0.88 | 0.673 | 0.582(212) | 0.598(212) | −0.0634 | 0.15 |
| dan_daner_ocgas. | 1.02 | 0.36 | 0.563(986) | 0.578(1023) | −0.0581 | 0.0634 |
| dan_daner_sickkids. | 1.00 | 0.742 | 0.596(275) | 0.596(205) | −0.0571 | 0.173 |
| **Meta** | **0.99** | **1.79e−08** | **0.569(53660)** | **0.556(2044417)** | **−0.0398** | **0.0071** |

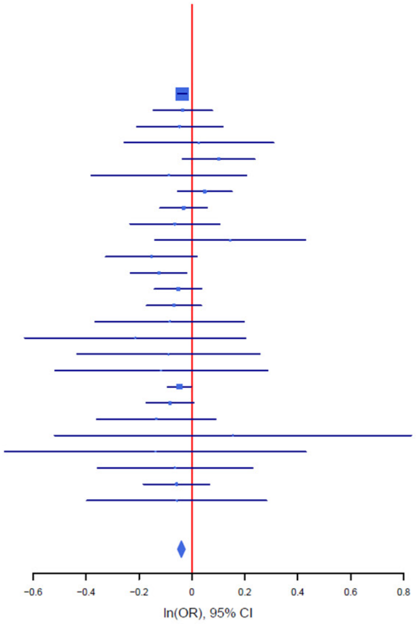

B

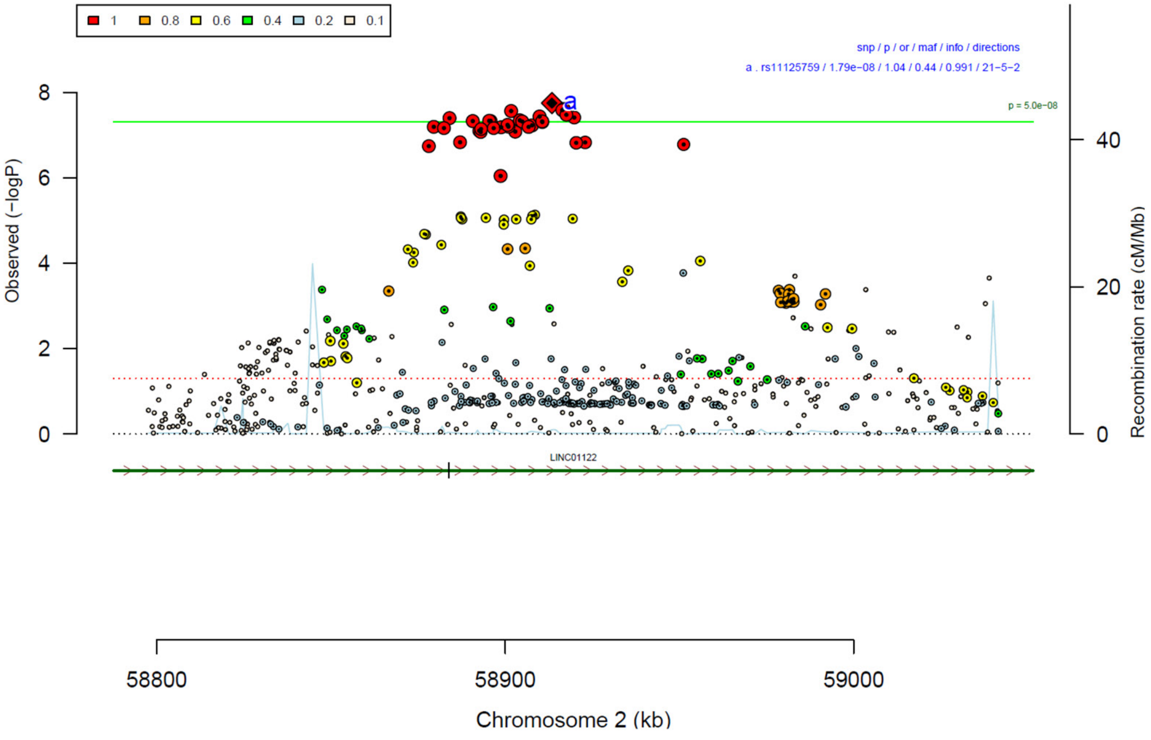

**Supplementary Figure 23: Forest plot (A) and regional association plot (B) of SNP rs11125759 in the meta-analysis.** The forest plot (A) shows the imputation quality (INFO) score, p-value of the SNP association (two-sided and not corrected for multiple testing, derived from a fixed-effects inverse-variance weighted meta-analysis), allele frequency in cases with case sample size (f_ca(n)), allele frequency in controls and control sample size (f_co(n)), beta estimates (ln(OR)) and standard error (STDerr) for each study as well as for the combined meta-analysis. Each cohort's sample size is detailed in supplementary table 1. Data on the right show the ln(OR) value as listed per cohort with 95% confidence intervals. In the regional association plot (B), the –log10 p-values (from two-sided association tests) are shown on the left y-axis. Recombination rates expressed in centimorgans (cM) per Mb (Megabase) (blue line) are shown on the right y-axis. Position in Mb is on the x-axis, with genes shown below the regional association plot. Only SNPs with an association p-value less than 0.1 were plotted. The most associated SNP is shown as a diamond and marked with the letter a.

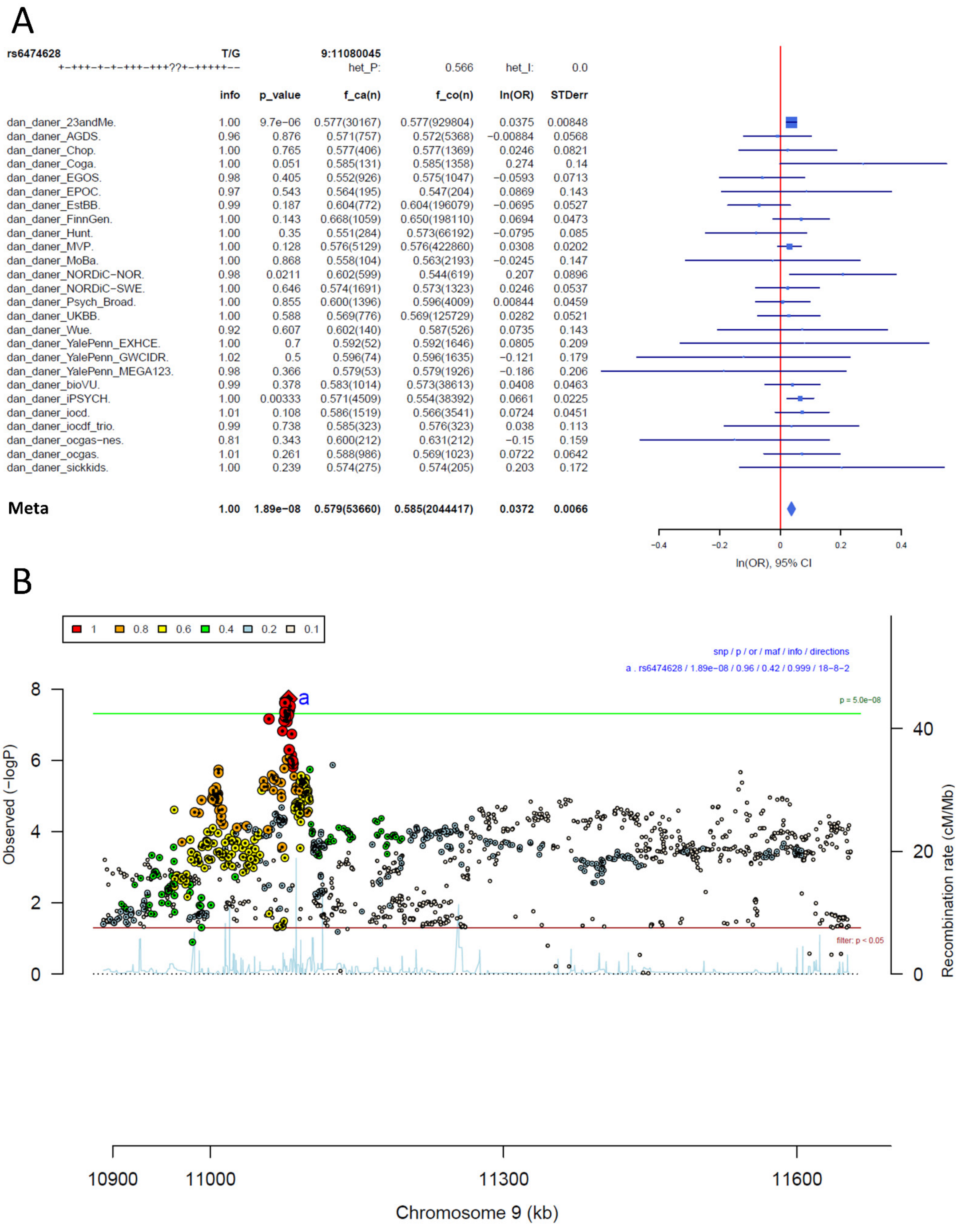

| | info | p_value | f_ca(n) | f_co(n) | ln(OR) | STDerr |
|---|---|---|---|---|---|---|
| dan_daner_23andMe. | 1.00 | 9.7e-06 | 0.577(30167) | 0.577(929804) | 0.0375 | 0.00848 |
| dan_daner_AGDS. | 0.96 | 0.876 | 0.571(757) | 0.572(5368) | -0.00884 | 0.0568 |
| dan_daner_Chop. | 1.00 | 0.765 | 0.577(406) | 0.577(1369) | 0.0246 | 0.0821 |
| dan_daner_Coga. | 1.00 | 0.051 | 0.585(131) | 0.585(1358) | 0.274 | 0.14 |
| dan_daner_EGOS. | 0.98 | 0.405 | 0.552(926) | 0.575(1047) | -0.0593 | 0.0713 |
| dan_daner_EPOC. | 0.97 | 0.543 | 0.564(195) | 0.547(204) | 0.0869 | 0.143 |
| dan_daner_EstBB. | 0.99 | 0.187 | 0.604(772) | 0.604(196079) | -0.0695 | 0.0527 |
| dan_daner_FinnGen. | 1.00 | 0.143 | 0.668(1059) | 0.650(198110) | 0.0694 | 0.0473 |
| dan_daner_Hunt. | 1.00 | 0.35 | 0.551(284) | 0.573(66192) | -0.0795 | 0.085 |
| dan_daner_MVP. | 1.00 | 0.128 | 0.576(5129) | 0.576(422860) | 0.0308 | 0.0202 |
| dan_daner_MoBa. | 1.00 | 0.868 | 0.558(104) | 0.563(2193) | -0.0245 | 0.147 |
| dan_daner_NORDiC-NOR. | 0.98 | 0.0211 | 0.602(599) | 0.544(619) | 0.207 | 0.0896 |
| dan_daner_NORDiC-SWE. | 1.00 | 0.646 | 0.574(1691) | 0.573(1323) | 0.0246 | 0.0537 |
| dan_daner_Psych_Broad. | 1.00 | 0.855 | 0.600(1396) | 0.596(4009) | 0.00844 | 0.0459 |
| dan_daner_UKBB. | 1.00 | 0.588 | 0.569(776) | 0.569(125729) | 0.0282 | 0.0521 |
| dan_daner_Wue. | 0.92 | 0.607 | 0.602(140) | 0.587(526) | 0.0735 | 0.143 |
| dan_daner_YalePenn_EXHCE. | 1.00 | 0.7 | 0.592(52) | 0.592(1646) | 0.0805 | 0.209 |
| dan_daner_YalePenn_GWCIDR. | 1.02 | 0.5 | 0.596(74) | 0.596(1635) | -0.121 | 0.179 |
| dan_daner_YalePenn_MEGA123. | 0.98 | 0.366 | 0.579(53) | 0.579(1926) | -0.186 | 0.206 |
| dan_daner_bioVU. | 0.99 | 0.378 | 0.583(1014) | 0.573(38613) | 0.0408 | 0.0463 |
| dan_daner_iPSYCH. | 1.00 | 0.00333 | 0.571(4509) | 0.554(38392) | 0.0661 | 0.0225 |
| dan_daner_iocd. | 1.01 | 0.108 | 0.586(1519) | 0.566(3541) | 0.0724 | 0.0451 |
| dan_daner_iocdf_trio. | 0.99 | 0.738 | 0.585(323) | 0.576(323) | 0.038 | 0.113 |
| dan_daner_ocgas-nes. | 0.81 | 0.343 | 0.600(212) | 0.631(212) | -0.15 | 0.159 |
| dan_daner_ocgas. | 1.01 | 0.261 | 0.588(986) | 0.569(1023) | 0.0722 | 0.0642 |
| dan_daner_sickkids. | 1.00 | 0.239 | 0.574(275) | 0.574(205) | 0.203 | 0.172 |
| **Meta** | **1.00** | **1.89e-08** | **0.579(53660)** | **0.585(2044417)** | **0.0372** | **0.0066** |

**Supplementary Figure 24: Forest plot (A) and regional association plot (B) of SNP rs6474628 in the meta-analysis.** The forest plot (A) shows the imputation quality (INFO) score, p-value of the SNP association (two-sided and not corrected for multiple testing, derived from a fixed-effects inverse-variance weighted meta-analysis), allele frequency in cases with case sample size (f_ca(n)), allele frequency in controls and control sample size (f_co(n)), beta estimates (ln(OR)) and standard error (STDerr) for each study as well as for the combined meta-analysis. Each cohort's sample size is detailed in supplementary table 1. Data on the right show the ln(OR) value as listed per cohort with 95% confidence intervals. In the regional association plot (B), the –log10 p-values (from two-sided association tests) are shown on the left y-axis. Recombination rates expressed in centimorgans (cM) per Mb (Megabase) (blue line) are shown on the right y-axis. Position in Mb is on the x-axis, with genes shown below the regional association plot. Only SNPs with an association p-value less than 0.1 were plotted. The most associated SNP is shown as a diamond and marked with the letter a.

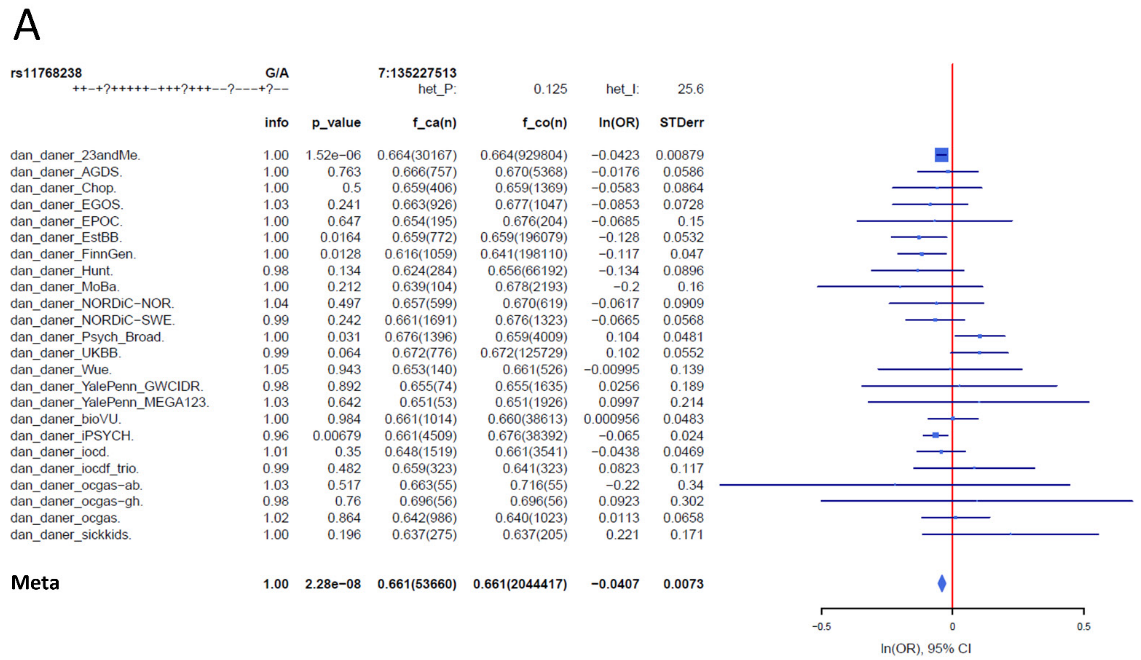

| | info | p_value | f_ca(n) | f_co(n) | ln(OR) | STDerr |
|---|---|---|---|---|---|---|
| dan_daner_23andMe. | 1.00 | 1.52e−06 | 0.664(30167) | 0.664(929804) | −0.0423 | 0.00879 |
| dan_daner_AGDS. | 1.00 | 0.763 | 0.666(757) | 0.670(5368) | −0.0176 | 0.0586 |
| dan_daner_Chop. | 1.00 | 0.5 | 0.659(406) | 0.659(1369) | −0.0583 | 0.0864 |
| dan_daner_EGOS. | 1.03 | 0.241 | 0.663(926) | 0.677(1047) | −0.0853 | 0.0728 |
| dan_daner_EPOC. | 1.00 | 0.647 | 0.654(195) | 0.676(204) | −0.0685 | 0.15 |
| dan_daner_EstBB. | 1.00 | 0.0164 | 0.659(772) | 0.659(196079) | −0.128 | 0.0532 |
| dan_daner_FinnGen. | 1.00 | 0.0128 | 0.616(1059) | 0.641(198110) | −0.117 | 0.047 |
| dan_daner_Hunt. | 0.98 | 0.134 | 0.624(284) | 0.656(66192) | −0.134 | 0.0896 |
| dan_daner_MoBa. | 1.00 | 0.212 | 0.639(104) | 0.678(2193) | −0.2 | 0.16 |
| dan_daner_NORDiC−NOR. | 1.04 | 0.497 | 0.657(599) | 0.670(619) | −0.0617 | 0.0909 |
| dan_daner_NORDiC−SWE. | 0.99 | 0.242 | 0.661(1691) | 0.676(1323) | −0.0665 | 0.0568 |
| dan_daner_Psych_Broad. | 1.00 | 0.031 | 0.676(1396) | 0.659(4009) | 0.104 | 0.0481 |
| dan_daner_UKBB. | 0.99 | 0.064 | 0.672(776) | 0.672(125729) | 0.102 | 0.0552 |
| dan_daner_Wue. | 1.05 | 0.943 | 0.653(140) | 0.661(526) | −0.00995 | 0.139 |
| dan_daner_YalePenn_GWCIDR. | 0.98 | 0.892 | 0.655(74) | 0.655(1635) | 0.0256 | 0.189 |
| dan_daner_YalePenn_MEGA123. | 1.03 | 0.642 | 0.651(53) | 0.651(1926) | 0.0997 | 0.214 |
| dan_daner_bioVU. | 1.00 | 0.984 | 0.661(1014) | 0.660(38613) | 0.000956 | 0.0483 |
| dan_daner_iPSYCH. | 0.96 | 0.00679 | 0.661(4509) | 0.676(38392) | −0.065 | 0.024 |
| dan_daner_iocd. | 1.01 | 0.35 | 0.648(1519) | 0.661(3541) | −0.0438 | 0.0469 |
| dan_daner_iocdf_trio. | 0.99 | 0.482 | 0.659(323) | 0.641(323) | 0.0823 | 0.117 |
| dan_daner_ocgas−ab. | 1.03 | 0.517 | 0.663(55) | 0.716(55) | −0.22 | 0.34 |
| dan_daner_ocgas−gh. | 0.98 | 0.76 | 0.696(56) | 0.696(56) | 0.0923 | 0.302 |
| dan_daner_ocgas. | 1.02 | 0.864 | 0.642(986) | 0.640(1023) | 0.0113 | 0.0658 |
| dan_daner_sickkids. | 1.00 | 0.196 | 0.637(275) | 0.637(205) | 0.221 | 0.171 |
| **Meta** | **1.00** | **2.28e−08** | **0.661(53660)** | **0.661(2044417)** | **−0.0407** | **0.0073** |

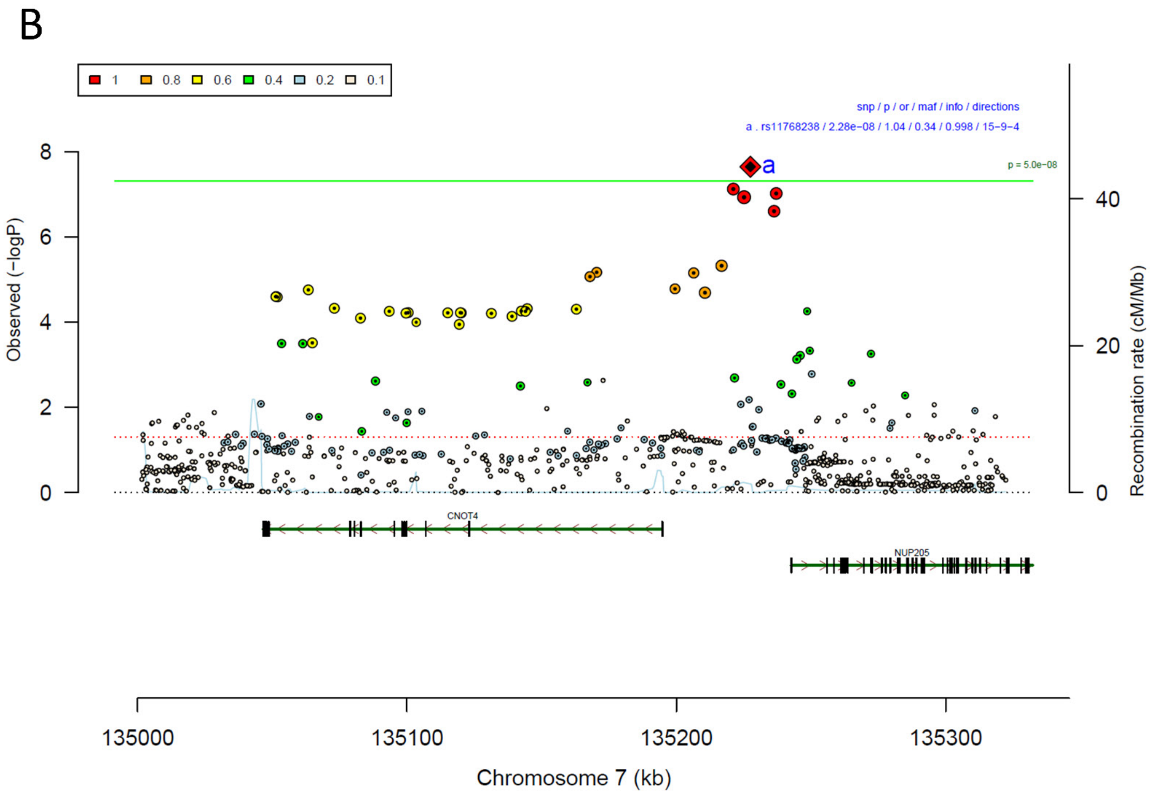

**Supplementary Figure 25: Forest plot (A) and regional association plot (B) of SNP rs11768238 in the meta-analysis.** The forest plot (A) shows the imputation quality (INFO) score, p-value of the SNP association (two-sided and not corrected for multiple testing, derived from a fixed-effects inverse-variance weighted meta-analysis), allele frequency in cases with case sample size (f_ca(n)), allele frequency in controls and control sample size (f_co(n)), beta estimates (ln(OR)) and standard error (STDerr) for each study as well as for the combined meta-analysis. Each cohort's sample size is detailed in supplementary table 1. Data on the right show the ln(OR) value as listed per cohort with 95% confidence intervals. In the regional association plot (B), the –log10 p-values (from two-sided association tests) are shown on the left y-axis. Recombination rates expressed in centimorgans (cM) per Mb (Megabase) (blue line) are shown on the right y-axis. Position in Mb is on the x-axis, with genes shown below the regional association plot. Only SNPs with an association p-value less than 0.1 were plotted. The most associated SNP is shown as a diamond and marked with the letter a.

A

| rs9479138 | T/G | | 6:152215199 | | | |
|---|---|---|---|---|---|---|
| ++?-++-++++-+++++++---+++--+ | | | het_P: | 0.74 | het_I: | 0.0 |
| | info | p_value | f_ca(n) | f_co(n) | ln(OR) | STDerr |
| dan_daner_23andMe. | 0.99 | 6.68e−06 | 0.333(30167) | 0.333(929804) | 0.0399 | 0.00884 |
| dan_daner_AGDS. | 0.99 | 0.221 | 0.386(757) | 0.371(5368) | 0.0704 | 0.0574 |
| dan_daner_Chop. | 1.00 | 0.55 | 0.343(406) | 0.343(1369) | −0.0531 | 0.0889 |
| dan_daner_Coga. | 1.00 | 0.757 | 0.358(131) | 0.358(1358) | 0.0424 | 0.137 |
| dan_daner_EGOS. | 0.99 | 0.708 | 0.333(926) | 0.322(1047) | 0.0279 | 0.0745 |
| dan_daner_EPOC. | 0.93 | 0.645 | 0.306(195) | 0.316(204) | −0.0735 | 0.159 |
| dan_daner_EstBB. | 0.94 | 0.902 | 0.298(772) | 0.298(196079) | 0.00707 | 0.0576 |
| dan_daner_FinnGen. | 1.00 | 0.435 | 0.376(1059) | 0.360(198110) | 0.0364 | 0.0467 |
| dan_daner_Hunt. | 1.00 | 0.609 | 0.360(284) | 0.348(66192) | 0.0454 | 0.0887 |
| dan_daner_MVP. | 0.93 | 0.0289 | 0.357(5129) | 0.357(422860) | 0.047 | 0.0215 |
| dan_daner_MoBa. | 0.99 | 0.78 | 0.346(104) | 0.337(2193) | 0.0426 | 0.153 |
| dan_daner_NORDiC−NOR. | 1.00 | 0.509 | 0.348(599) | 0.336(619) | 0.0613 | 0.0928 |
| dan_daner_NORDiC−SWE. | 0.96 | 0.854 | 0.318(1691) | 0.315(1323) | 0.0106 | 0.0582 |
| dan_daner_Psych_Broad. | 0.99 | 0.971 | 0.312(1396) | 0.308(4009) | −0.0018 | 0.049 |
| dan_daner_UKBB. | 1.00 | 0.714 | 0.361(776) | 0.361(125729) | 0.0198 | 0.054 |
| dan_daner_Wue. | 1.00 | 0.00793 | 0.398(140) | 0.311(526) | 0.374 | 0.141 |
| dan_daner_YalePenn_EXHCE. | 1.00 | 0.755 | 0.340(52) | 0.340(1646) | −0.0693 | 0.222 |
| dan_daner_YalePenn_GWCIDR. | 1.06 | 0.272 | 0.349(74) | 0.349(1635) | −0.206 | 0.188 |
| dan_daner_YalePenn_MEGA123. | 1.05 | 0.147 | 0.342(53) | 0.342(1926) | 0.295 | 0.203 |
| dan_daner_iPSYCH. | 1.00 | 0.0165 | 0.363(4509) | 0.351(38392) | 0.0556 | 0.0232 |
| dan_daner_iocd. | 1.00 | 0.365 | 0.328(1519) | 0.341(3541) | −0.043 | 0.0474 |
| dan_daner_iocdf_trio. | 0.95 | 0.173 | 0.330(323) | 0.296(323) | 0.168 | 0.123 |
| dan_daner_ocgas−ab. | 0.95 | 0.282 | 0.376(55) | 0.282(55) | 0.401 | 0.373 |
| dan_daner_ocgas−gh. | 0.91 | 0.335 | 0.298(56) | 0.257(56) | 0.32 | 0.332 |
| dan_daner_ocgas−nes. | 0.96 | 0.922 | 0.310(212) | 0.321(212) | −0.0149 | 0.153 |
| dan_daner_ocgas. | 1.01 | 0.965 | 0.344(986) | 0.344(1023) | −0.0029 | 0.0668 |
| dan_daner_sickkids. | 1.00 | 0.138 | 0.376(275) | 0.376(205) | 0.271 | 0.183 |
| **Meta** | **0.97** | **2.41e−08** | **0.339(53660)** | **0.340(2044417)** | **0.0389** | **0.007** |

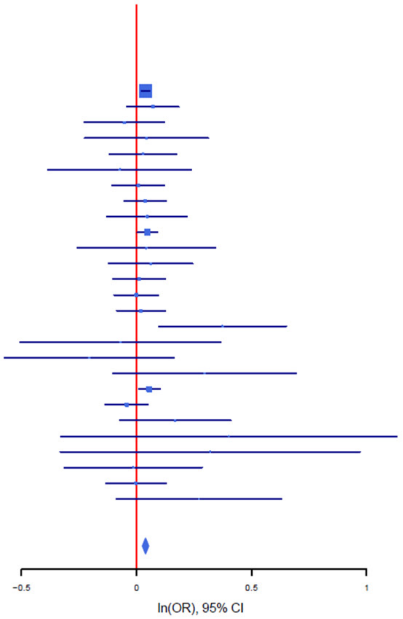

B

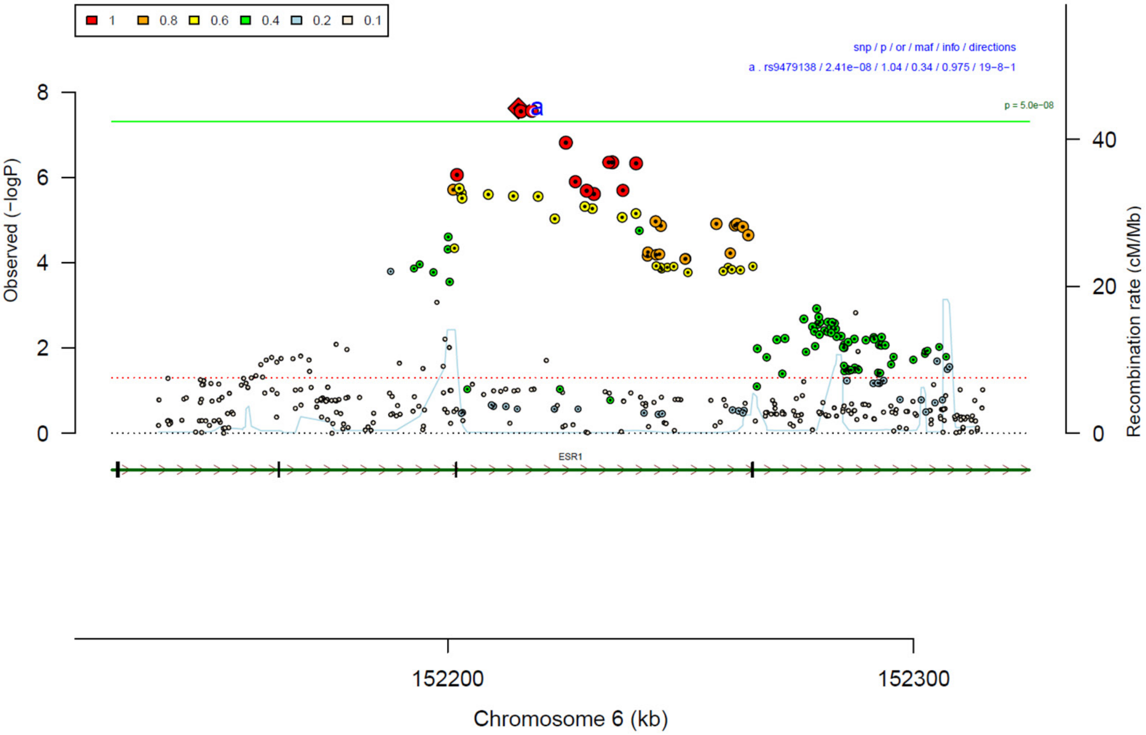

**Supplementary Figure 26: Forest plot (A) and regional association plot (B) of SNP rs9479138 in the meta-analysis.** The forest plot (A) shows the imputation quality (INFO) score, p-value of the SNP association (two-sided and not corrected for multiple testing, derived from a fixed-effects inverse-variance weighted meta-analysis), allele frequency in cases with case sample size (f_ca(n)), allele frequency in controls and control sample size (f_co(n)), beta estimates (ln(OR)) and standard error (STDerr) for each study as well as for the combined meta-analysis. Each cohort's sample size is detailed in supplementary table 1. Data on the right show the ln(OR) value as listed per cohort with 95% confidence intervals. In the regional association plot (B), the –log10 p-values (from two-sided association tests) are shown on the left y-axis. Recombination rates expressed in centimorgans (cM) per Mb (Megabase) (blue line) are shown on the right y-axis. Position in Mb is on the x-axis, with genes shown below the regional association plot. Only SNPs with an association p-value less than 0.1 were plotted. The most associated SNP is shown as a diamond and marked with the letter a.

A

| rs1567288 | G/A | | 4:98582985 | | | |
|---|---|---|---|---|---|---|
| +-++++?-++++++++++++++-+-+++ | | | het_P: | 0.276 | het_I: | 12.7 |
| | info | p_value | f_ca(n) | f_co(n) | ln(OR) | STDerr |
| dan_daner_23andMe. | 0.99 | 0.000863 | 0.555(30167) | 0.555(929804) | −0.0282 | 0.00846 |
| dan_daner_AGDS. | 0.90 | 0.393 | 0.550(757) | 0.535(5368) | 0.0501 | 0.0586 |
| dan_daner_Chop. | 1.00 | 0.889 | 0.559(406) | 0.559(1369) | −0.0115 | 0.0824 |
| dan_daner_Coga. | 1.00 | 0.605 | 0.568(131) | 0.568(1358) | −0.072 | 0.139 |
| dan_daner_EGOS. | 0.93 | 0.206 | 0.544(926) | 0.566(1047) | −0.0919 | 0.0728 |
| dan_daner_EstBB. | 0.99 | 0.838 | 0.558(772) | 0.558(196079) | 0.0107 | 0.052 |
| dan_daner_FinnGen. | 1.00 | 0.63 | 0.517(1059) | 0.523(198110) | −0.0216 | 0.0449 |
| dan_daner_Hunt. | 0.99 | 0.0679 | 0.544(284) | 0.581(66192) | −0.157 | 0.086 |
| dan_daner_MVP. | 0.95 | 0.0748 | 0.545(5129) | 0.545(422860) | −0.0367 | 0.0206 |
| dan_daner_MoBa. | 0.99 | 0.186 | 0.510(104) | 0.561(2193) | −0.188 | 0.142 |
| dan_daner_NORDiC-NOR. | 0.96 | 0.29 | 0.556(599) | 0.567(619) | −0.0949 | 0.0897 |
| dan_daner_NORDiC-SWE. | 0.96 | 0.121 | 0.536(1691) | 0.554(1323) | −0.0841 | 0.0543 |
| dan_daner_Psych_Broad. | 0.99 | 0.685 | 0.569(1396) | 0.571(4009) | −0.0184 | 0.0455 |
| dan_daner_UKBB. | 1.00 | 0.886 | 0.550(776) | 0.550(125729) | −0.0075 | 0.0522 |
| dan_daner_Wue. | 0.80 | 0.804 | 0.583(140) | 0.575(526) | 0.0382 | 0.154 |
| dan_daner_YalePenn_EXHCE. | 1.04 | 0.338 | 0.582(52) | 0.582(1646) | −0.197 | 0.205 |
| dan_daner_YalePenn_GWCIDR. | 0.97 | 0.00999 | 0.574(74) | 0.574(1635) | −0.476 | 0.185 |
| dan_daner_YalePenn_MEGA123. | 0.98 | 0.297 | 0.563(53) | 0.563(1926) | −0.217 | 0.208 |
| dan_daner_bioVU. | 0.99 | 0.00195 | 0.517(1014) | 0.545(38613) | −0.142 | 0.046 |
| dan_daner_iPSYCH. | 0.97 | 0.0196 | 0.514(4509) | 0.526(38392) | −0.0528 | 0.0226 |
| dan_daner_iocd. | 0.97 | 0.273 | 0.546(1519) | 0.559(3541) | −0.0499 | 0.0456 |
| dan_daner_iocdf_trio. | 0.98 | 0.315 | 0.548(323) | 0.576(323) | −0.114 | 0.113 |
| dan_daner_ocgas-ab. | 0.93 | 0.506 | 0.513(55) | 0.585(55) | −0.225 | 0.338 |
| dan_daner_ocgas-gh. | 0.87 | 0.19 | 0.528(56) | 0.606(56) | −0.398 | 0.304 |
| dan_daner_ocgas-nes. | 0.95 | 0.14 | 0.517(212) | 0.567(212) | −0.212 | 0.144 |
| dan_daner_ocgas. | 1.01 | 0.216 | 0.553(986) | 0.572(1023) | −0.0791 | 0.0639 |
| dan_daner_sickkids. | 1.00 | 0.134 | 0.556(275) | 0.556(205) | 0.261 | 0.174 |
| **Meta** | **0.98** | **3.8e−08** | **0.548(53660)** | **0.550(2044417)** | **−0.0364** | **0.0066** |

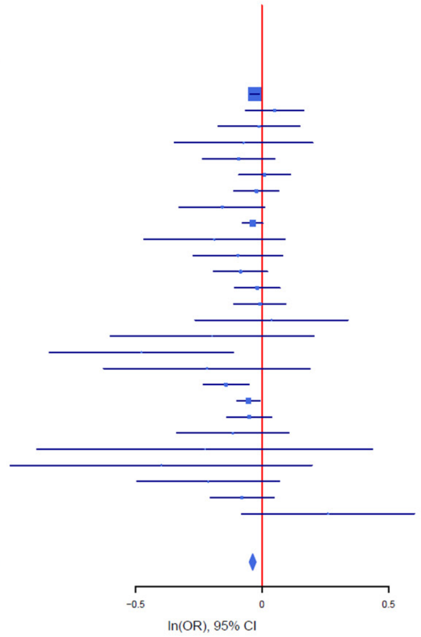

B

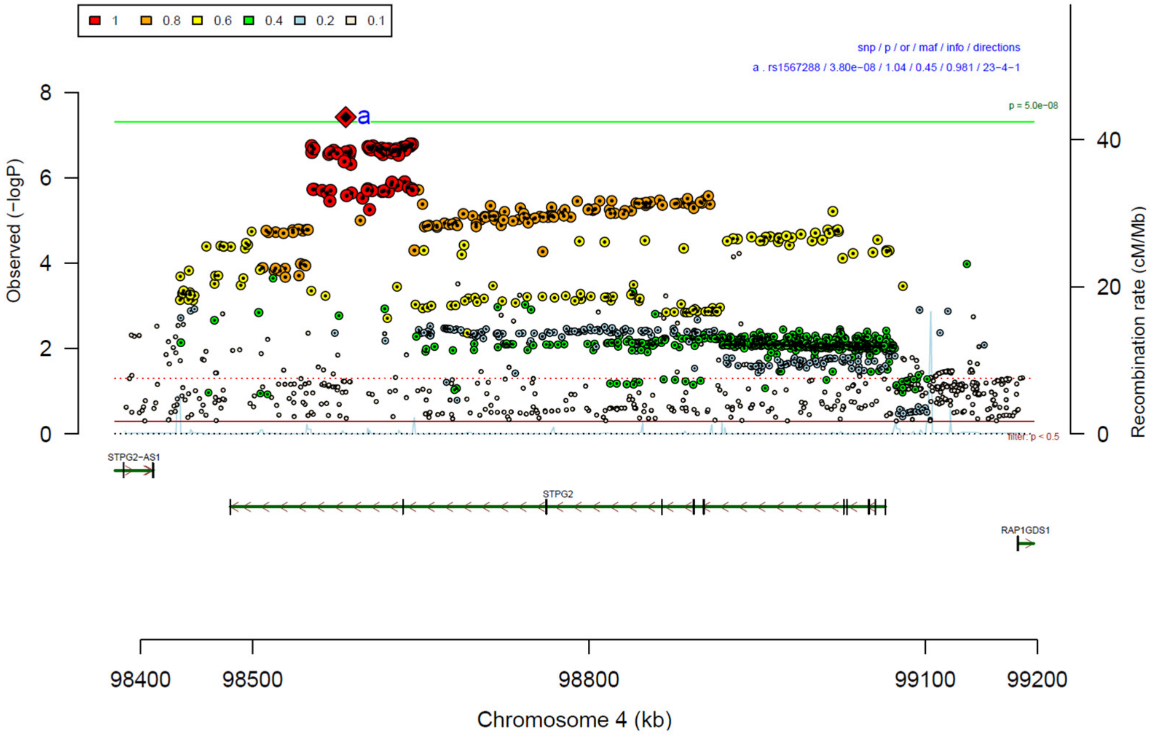

**Supplementary Figure 27: Forest plot (A) and regional association plot (B) of SNP rs1567288 in the meta-analysis.** The forest plot (A) shows the imputation quality (INFO) score, p-value of the SNP association (two-sided and not corrected for multiple testing, derived from a fixed-effects inverse-variance weighted meta-analysis), allele frequency in cases with case sample size (f_ca(n)), allele frequency in controls and control sample size (f_co(n)), beta estimates (ln(OR)) and standard error (STDerr) for each study as well as for the combined meta-analysis. Each cohort's sample size is detailed in supplementary table 1. Data on the right show the ln(OR) value as listed per cohort with 95% confidence intervals. In the regional association plot (B), the –log10 p-values (from two-sided association tests) are shown on the left y-axis. Recombination rates expressed in centimorgans (cM) per Mb (Megabase) (blue line) are shown on the right y-axis. Position in Mb is on the x-axis, with genes shown below the regional association plot. Only SNPs with an association p-value less than 0.1 were plotted. The most associated SNP is shown as a diamond and marked with the letter a.

A

rs4831130 T/G 3:116039842

+++-++++++-++++--+?++++++-++ het_P: 0.85 het_I: 0.0

| | info | p_value | f_ca(n) | f_co(n) | ln(OR) | STDerr |
|---|---|---|---|---|---|---|
| dan_daner_23andMe. | 1.00 | 8.54e−06 | 0.757(30167) | 0.757(929804) | 0.0435 | 0.0098 |
| dan_daner_AGDS. | 1.00 | 0.0121 | 0.790(757) | 0.764(5368) | 0.17 | 0.0677 |
| dan_daner_Chop. | 1.00 | 0.849 | 0.758(406) | 0.758(1369) | −0.0182 | 0.0954 |
| dan_daner_Coga. | 1.00 | 0.61 | 0.758(131) | 0.758(1358) | 0.082 | 0.161 |
| dan_daner_EGOS. | 0.95 | 0.524 | 0.727(926) | 0.725(1047) | 0.0511 | 0.0802 |
| dan_daner_EPOC. | 1.00 | 0.76 | 0.756(195) | 0.743(204) | 0.052 | 0.171 |
| dan_daner_EstBB. | 0.98 | 0.301 | 0.700(772) | 0.700(196079) | 0.0585 | 0.0565 |
| dan_daner_FinnGen. | 1.00 | 0.718 | 0.665(1059) | 0.662(198110) | 0.0171 | 0.0475 |
| dan_daner_Hunt. | 1.00 | 0.573 | 0.755(284) | 0.744(66192) | 0.0546 | 0.097 |
| dan_daner_MVP. | 0.94 | 0.242 | 0.748(5129) | 0.748(422860) | 0.028 | 0.0239 |
| dan_daner_MoBa. | 0.97 | 0.477 | 0.784(104) | 0.763(2193) | 0.118 | 0.166 |
| dan_daner_NORDiC−NOR. | 0.97 | 0.661 | 0.784(599) | 0.778(619) | −0.0469 | 0.107 |
| dan_daner_NORDiC−SWE. | 0.97 | 0.206 | 0.742(1691) | 0.758(1323) | −0.079 | 0.0624 |
| dan_daner_Psych_Broad. | 0.99 | 0.876 | 0.744(1396) | 0.747(4009) | 0.00803 | 0.0517 |
| dan_daner_UKBB. | 0.99 | 0.517 | 0.753(776) | 0.753(125729) | 0.039 | 0.0601 |
| dan_daner_Wue. | 1.00 | 0.243 | 0.761(140) | 0.729(526) | 0.185 | 0.158 |
| dan_daner_YalePenn_EXHCE. | 1.01 | 0.271 | 0.749(52) | 0.749(1646) | −0.243 | 0.221 |
| dan_daner_YalePenn_GWCIDR. | 0.98 | 0.738 | 0.760(74) | 0.760(1635) | 0.0707 | 0.211 |
| dan_daner_YalePenn_MEGA123. | 0.96 | 0.476 | 0.759(53) | 0.759(1926) | 0.179 | 0.252 |
| dan_daner_bioVU. | 1.00 | 0.0189 | 0.779(1014) | 0.757(38613) | 0.124 | 0.0529 |
| dan_daner_iPSYCH. | 1.00 | 0.0576 | 0.747(4509) | 0.738(38392) | 0.0485 | 0.0256 |
| dan_daner_iocd. | 1.01 | 0.748 | 0.754(1519) | 0.751(3541) | 0.0165 | 0.0514 |
| dan_daner_iocdf_trio. | 0.96 | 0.693 | 0.748(323) | 0.757(323) | −0.0518 | 0.132 |
| dan_daner_ocgas−ab. | 0.91 | 0.406 | 0.777(55) | 0.727(55) | 0.318 | 0.383 |
| dan_daner_ocgas−nes. | 0.81 | 0.911 | 0.770(212) | 0.768(212) | 0.0205 | 0.182 |
| dan_daner_ocgas. | 0.97 | 0.61 | 0.753(986) | 0.746(1023) | 0.0378 | 0.074 |
| dan_daner_sickkids. | 1.00 | 0.0866 | 0.750(275) | 0.750(205) | 0.321 | 0.187 |
| **Meta** | **0.98** | **3.92e−08** | **0.753(53660)** | **0.740(2044417)** | **0.0418** | **0.0076** |

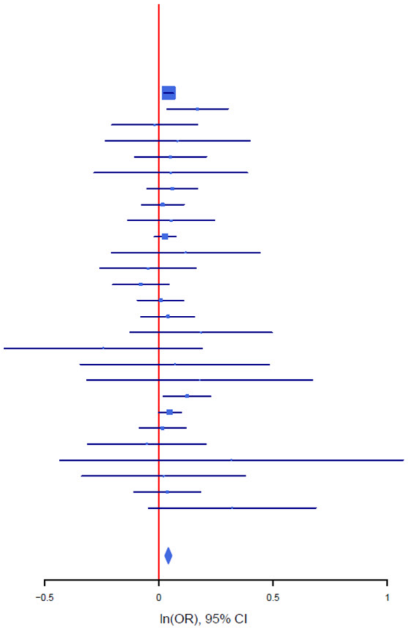

B

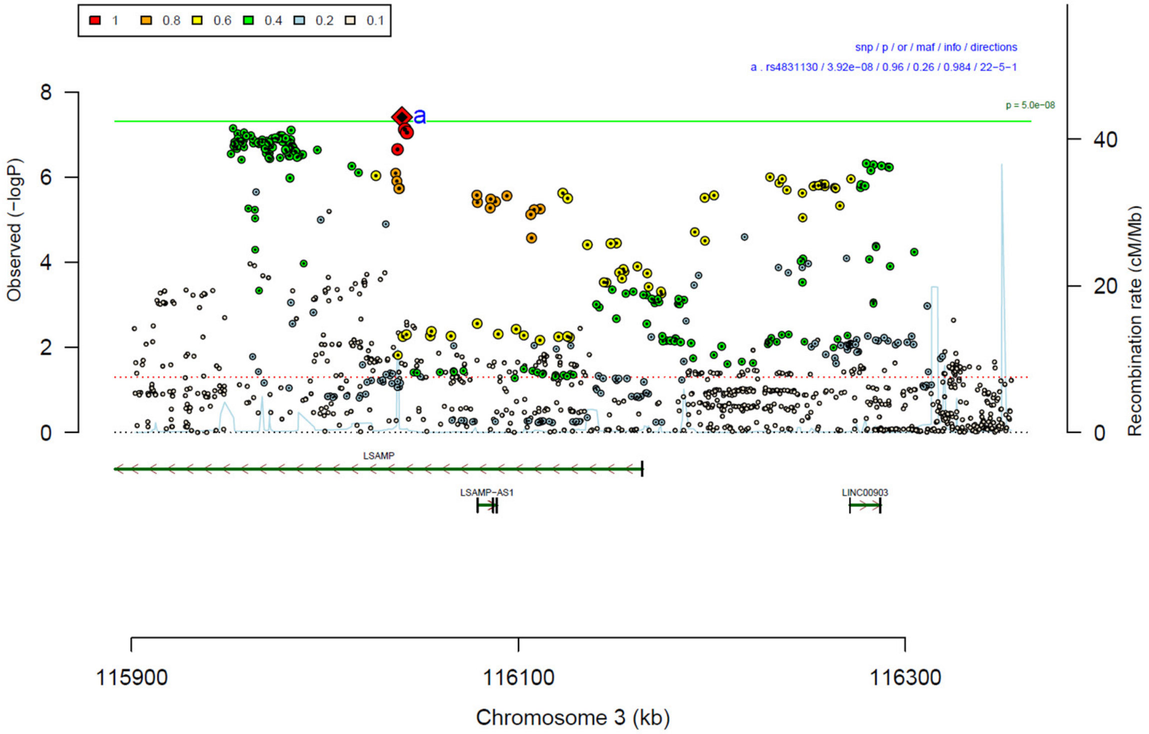

**Supplementary Figure 28: Forest plot (A) and regional association plot (B) of SNP rs4831130 in the meta-analysis.** The forest plot (A) shows the imputation quality (INFO) score, p-value of the SNP association (two-sided and not corrected for multiple testing, derived from a fixed-effects inverse-variance weighted meta-analysis), allele frequency in cases with case sample size (f_ca(n)), allele frequency in controls and control sample size (f_co(n)), beta estimates (ln(OR)) and standard error (STDerr) for each study as well as for the combined meta-analysis. Each cohort's sample size is detailed in supplementary table 1. Data on the right show the ln(OR) value as listed per cohort with 95% confidence intervals. In the regional association plot (B), the –log10 p-values (from two-sided association tests) are shown on the left y-axis. Recombination rates expressed in centimorgans (cM) per Mb (Megabase) (blue line) are shown on the right y-axis. Position in Mb is on the x-axis, with genes shown below the regional association plot. Only SNPs with an association p-value less than 0.1 were plotted. The most associated SNP is shown as a diamond and marked with the letter a.

A

| rs17718444 | T/C | | 3:71499401 | | | |
|---|---|---|---|---|---|---|
| --+-+--+-++--+-+------+-++-+ |  |  | het_P: | 0.131 | het_I: | 23.5 |
| | info | p_value | f_ca(n) | f_co(n) | ln(OR) | STDerr |
| dan_daner_23andMe. | 1.00 | 0.000119 | 0.334(30167) | 0.334(929804) | −0.0342 | 0.00892 |
| dan_daner_AGDS. | 0.93 | 0.0373 | 0.303(757) | 0.329(5368) | −0.13 | 0.0623 |
| dan_daner_Chop. | 1.00 | 0.0113 | 0.342(406) | 0.342(1369) | −0.224 | 0.0885 |
| dan_daner_Coga. | 1.00 | 0.951 | 0.314(131) | 0.314(1358) | 0.0091 | 0.148 |
| dan_daner_EGOS. | 0.99 | 0.31 | 0.286(926) | 0.304(1047) | −0.078 | 0.0767 |
| dan_daner_EPOC. | 0.92 | 0.967 | 0.316(195) | 0.325(204) | −0.00703 | 0.17 |
| dan_daner_EstBB. | 0.97 | 0.489 | 0.306(772) | 0.306(196079) | 0.0392 | 0.0566 |
| dan_daner_FinnGen. | 1.00 | 0.916 | 0.287(1059) | 0.289(198110) | −0.0052 | 0.0498 |
| dan_daner_Hunt. | 1.00 | 0.689 | 0.294(284) | 0.288(66192) | 0.0376 | 0.0939 |
| dan_daner_MVP. | 0.95 | 0.197 | 0.315(5129) | 0.315(422860) | −0.0286 | 0.0222 |
| dan_daner_MoBa. | 0.99 | 0.771 | 0.303(104) | 0.294(2193) | 0.0465 | 0.16 |
| dan_daner_NORDiC−NOR. | 0.96 | 0.68 | 0.287(599) | 0.273(619) | 0.0409 | 0.0992 |
| dan_daner_NORDiC−SWE. | 0.95 | 0.187 | 0.277(1691) | 0.290(1323) | −0.0797 | 0.0604 |
| dan_daner_Psych_Broad. | 1.02 | 1.87e−05 | 0.320(1396) | 0.366(4009) | −0.203 | 0.0474 |
| dan_daner_UKBB. | 1.00 | 0.00806 | 0.323(776) | 0.323(125729) | −0.143 | 0.054 |
| dan_daner_Wue. | 0.86 | 0.461 | 0.336(140) | 0.316(526) | 0.115 | 0.155 |
| dan_daner_YalePenn_EXHCE. | 0.98 | 0.224 | 0.333(52) | 0.333(1646) | 0.257 | 0.211 |
| dan_daner_YalePenn_GWCIDR. | 1.00 | 0.998 | 0.335(74) | 0.335(1635) | −0.000488 | 0.187 |
| dan_daner_YalePenn_MEGA123. | 1.01 | 0.546 | 0.338(53) | 0.338(1926) | 0.126 | 0.208 |
| dan_daner_bioVU. | 0.99 | 0.546 | 0.327(1014) | 0.319(38613) | 0.0297 | 0.0492 |
| dan_daner_iPSYCH. | 0.98 | 0.0467 | 0.290(4509) | 0.300(38392) | −0.0492 | 0.0247 |
| dan_daner_iocd. | 1.00 | 0.332 | 0.331(1519) | 0.338(3541) | −0.0461 | 0.0476 |
| dan_daner_iocdf_trio. | 1.03 | 0.423 | 0.357(323) | 0.336(323) | 0.0924 | 0.116 |
| dan_daner_ocgas−ab. | 1.00 | 0.773 | 0.298(55) | 0.355(55) | −0.1 | 0.346 |
| dan_daner_ocgas−gh. | 0.95 | 0.981 | 0.337(56) | 0.344(56) | −0.00727 | 0.297 |
| dan_daner_ocgas−nes. | 1.01 | 0.395 | 0.339(212) | 0.367(212) | −0.123 | 0.145 |
| dan_daner_ocgas. | 1.03 | 0.939 | 0.340(986) | 0.343(1023) | −0.00509 | 0.0663 |
| dan_daner_sickkids. | 1.00 | 0.895 | 0.328(275) | 0.328(205) | 0.0243 | 0.183 |
| **Meta** | **0.98** | **4.24e−08** | **0.323(53660)** | **0.320(2044417)** | **−0.0385** | **0.007** |

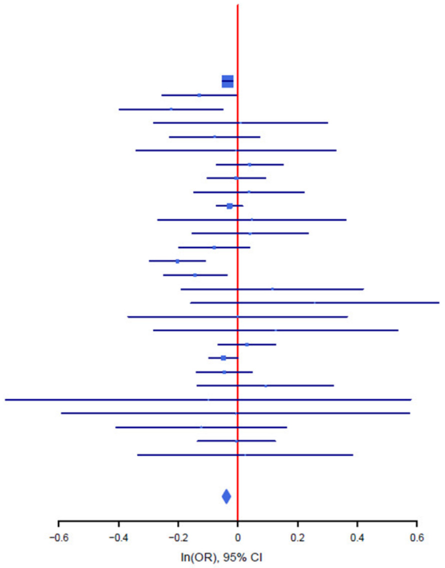

B

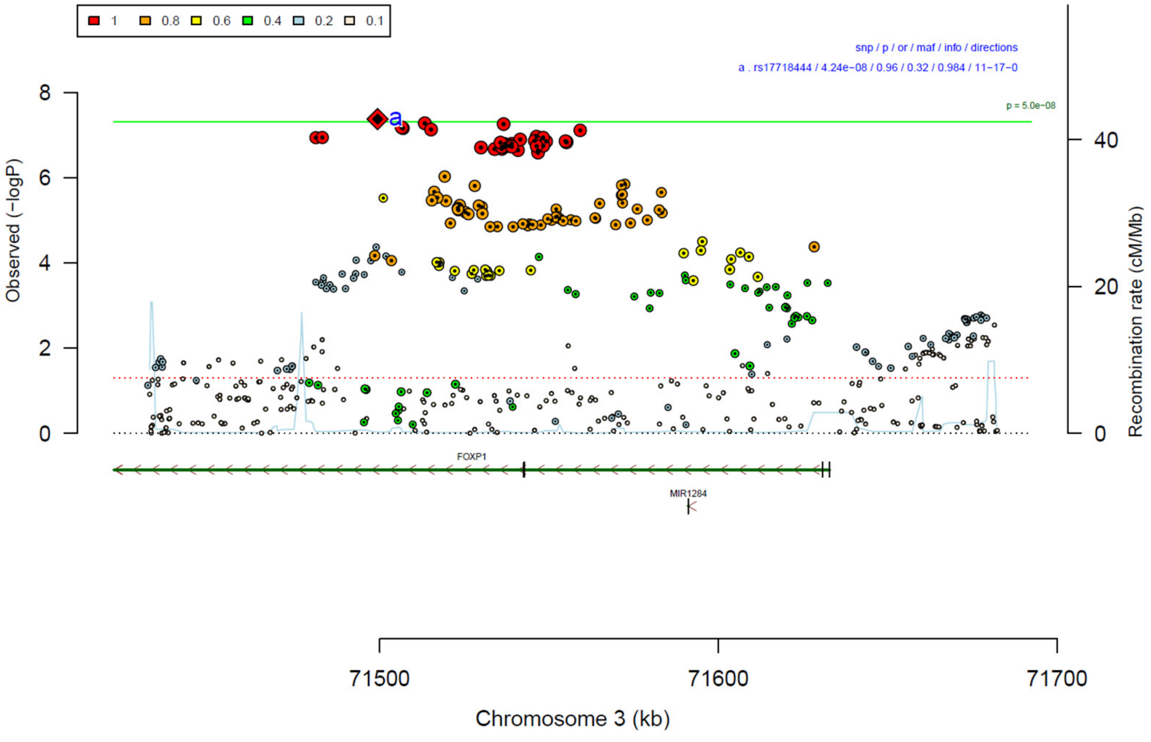

**Supplementary Figure 29: Forest plot (A) and regional association plot (B) of SNP rs17718444 in the meta-analysis.** The forest plot (A) shows the imputation quality (INFO) score, p-value of the SNP association (two-sided and not corrected for multiple testing, derived from a fixed-effects inverse-variance weighted meta-analysis), allele frequency in cases with case sample size (f_ca(n)), allele frequency in controls and control sample size (f_co(n)), beta estimates (ln(OR)) and standard error (STDerr) for each study as well as for the combined meta-analysis. Each cohort's sample size is detailed in supplementary table 1. Data on the right show the ln(OR) value as listed per cohort with 95% confidence intervals. In the regional association plot (B), the –log10 p-values (from two-sided association tests) are shown on the left y-axis. Recombination rates expressed in centimorgans (cM) per Mb (Megabase) (blue line) are shown on the right y-axis. Position in Mb is on the x-axis, with genes shown below the regional association plot. Only SNPs with an association p-value less than 0.1 were plotted. The most associated SNP is shown as a diamond and marked with the letter a.

A

rs6660196 T/G 1:168874798

++-+-+-+--+++-?-+--+++--++++

het_P: 0.0355 het_I: 35.7

| | info | p_value | f_ca(n) | f_co(n) | ln(OR) | STDerr |
|---|---|---|---|---|---|---|
| dan_daner_23andMe. | 0.99 | 4.32e−08 | 0.637(30167) | 0.637(929804) | 0.048 | 0.00879 |
| dan_daner_AGDS. | 1.00 | 0.773 | 0.627(757) | 0.621(5368) | 0.0165 | 0.0572 |
| dan_daner_Chop. | 1.00 | 0.293 | 0.647(406) | 0.647(1369) | 0.0902 | 0.0857 |
| dan_daner_Coga. | 1.00 | 0.041 | 0.637(131) | 0.637(1358) | −0.29 | 0.142 |
| dan_daner_EGOS. | 0.95 | 0.746 | 0.641(926) | 0.635(1047) | 0.0242 | 0.0746 |
| dan_daner_EPOC. | 1.00 | 0.636 | 0.613(195) | 0.630(204) | −0.0729 | 0.154 |
| dan_daner_EstBB. | 0.97 | 0.467 | 0.671(772) | 0.671(196079) | 0.0403 | 0.0555 |
| dan_daner_FinnGen. | 1.00 | 0.323 | 0.650(1059) | 0.662(198110) | −0.0469 | 0.0474 |
| dan_daner_Hunt. | 1.00 | 0.196 | 0.606(284) | 0.635(66192) | −0.113 | 0.0875 |
| dan_daner_MoBa. | 0.99 | 0.69 | 0.606(104) | 0.619(2193) | −0.0584 | 0.146 |
| dan_daner_NORDiC−NOR. | 1.04 | 0.978 | 0.610(599) | 0.615(619) | −0.0024 | 0.0879 |
| dan_daner_NORDiC−SWE. | 0.99 | 0.392 | 0.647(1691) | 0.636(1323) | 0.0476 | 0.0557 |
| dan_daner_Psych_Broad. | 0.96 | 0.729 | 0.642(1396) | 0.633(4009) | 0.0165 | 0.0477 |
| dan_daner_UKBB. | 0.99 | 0.31 | 0.620(776) | 0.620(125729) | −0.0545 | 0.0536 |
| dan_daner_Wue. | 0.97 | 0.212 | 0.682(140) | 0.643(526) | 0.184 | 0.147 |
| dan_daner_YalePenn_EXHCE. | 0.99 | 0.261 | 0.659(52) | 0.659(1646) | 0.261 | 0.232 |
| dan_daner_YalePenn_GWCIDR. | 1.00 | 0.104 | 0.646(74) | 0.646(1635) | 0.321 | 0.197 |
| dan_daner_YalePenn_MEGA123. | 1.01 | 0.537 | 0.662(53) | 0.662(1926) | 0.133 | 0.216 |
| dan_daner_bioVU. | 1.00 | 0.649 | 0.628(1014) | 0.628(38613) | −0.0216 | 0.0474 |
| dan_daner_iPSYCH. | 0.99 | 0.14 | 0.623(4509) | 0.614(38392) | 0.0341 | 0.0231 |
| dan_daner_iocd. | 1.03 | 0.243 | 0.652(1519) | 0.637(3541) | 0.0539 | 0.0462 |
| dan_daner_iocdf_trio. | 0.92 | 0.0128 | 0.688(323) | 0.624(323) | 0.308 | 0.124 |
| dan_daner_ocgas−ab. | 0.83 | 0.415 | 0.657(55) | 0.712(55) | −0.323 | 0.396 |
| dan_daner_ocgas−gh. | 0.98 | 0.108 | 0.605(56) | 0.692(56) | −0.487 | 0.303 |
| dan_daner_ocgas−nes. | 0.93 | 0.0131 | 0.718(212) | 0.640(212) | 0.39 | 0.157 |
| dan_daner_ocgas. | 1.00 | 0.102 | 0.657(986) | 0.633(1023) | 0.109 | 0.0665 |
| dan_daner_sickkids. | 1.00 | 0.217 | 0.650(275) | 0.650(205) | −0.226 | 0.183 |
| **Meta** | **0.99** | **4.86e−08** | **0.638(53660)** | **0.642(2044417)** | **0.0395** | **0.0072** |

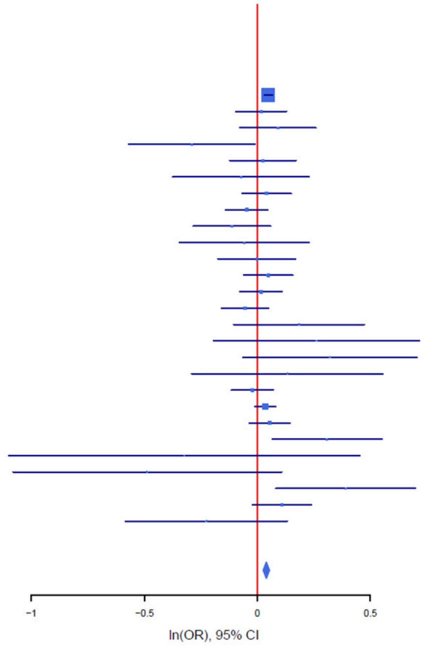

B

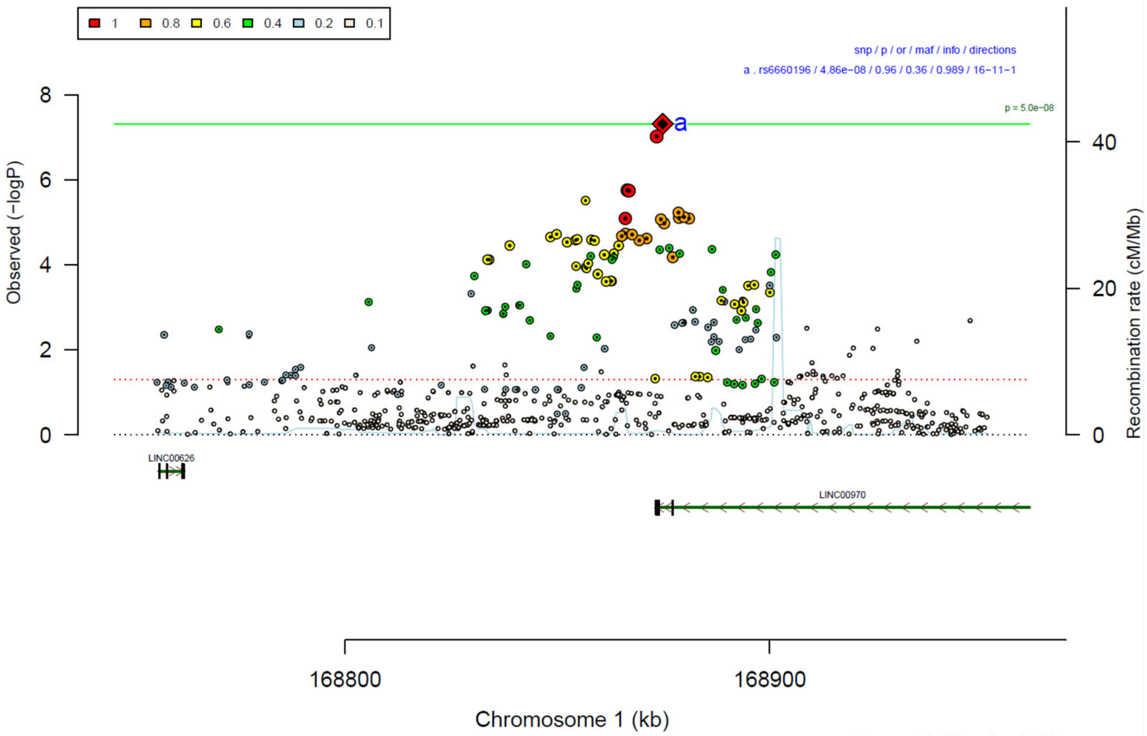

**Supplementary Figure 30: Forest plot (A) and regional association plot (B) of SNP rs6660196 in the meta-analysis.** The forest plot (A) shows the imputation quality (INFO) score, p-value of the SNP association (two-sided and not corrected for multiple testing, derived from a fixed-effects inverse-variance weighted meta-analysis), allele frequency in cases with case sample size (f_ca(n)), allele frequency in controls and control sample size (f_co(n)), beta estimates (ln(OR)) and standard error (STDerr) for each study as well as for the combined meta-analysis. Each cohort's sample size is detailed in supplementary table 1. Data on the right show the ln(OR) value as listed per cohort with 95% confidence intervals. In the regional association plot (B), the –log10 p-values (from two-sided association tests) are shown on the left y-axis. Recombination rates expressed in centimorgans (cM) per Mb (Megabase) (blue line) are shown on the right y-axis. Position in Mb is on the x-axis, with genes shown below the regional association plot. Only SNPs with an association p-value less than 0.1 were plotted. The most associated SNP is shown as a diamond and marked with the letter a.

A

| rs4931 | C/A | | 20:43530234 | | | |
|---|---|---|---|---|---|---|
| +++++−+++++++++++?++++−++?−+ | | | het_P: | 0.984 | het_I: | 0.0 |
| | **info** | **p_value** | **f_ca(n)** | **f_co(n)** | **ln(OR)** | **STDerr** |
| dan_daner_23andMe. | 1.00 | 0.000939 | 0.287(30167) | 0.287(929804) | −0.0307 | 0.00931 |
| dan_daner_AGDS. | 0.96 | 0.478 | 0.257(757) | 0.264(5368) | −0.0458 | 0.0647 |
| dan_daner_Chop. | 1.00 | 0.264 | 0.310(406) | 0.310(1369) | −0.101 | 0.0906 |
| dan_daner_Coga. | 1.00 | 0.858 | 0.291(131) | 0.291(1358) | −0.0267 | 0.15 |
| dan_daner_EGOS. | 0.98 | 0.513 | 0.254(926) | 0.247(1047) | 0.0532 | 0.0813 |
| dan_daner_EPOC. | 0.94 | 0.328 | 0.297(195) | 0.324(204) | −0.156 | 0.16 |
| dan_daner_EstBB. | 0.99 | 0.267 | 0.289(772) | 0.289(196079) | −0.0628 | 0.0566 |
| dan_daner_FinnGen. | 1.00 | 0.399 | 0.233(1059) | 0.246(198110) | −0.0443 | 0.0525 |
| dan_daner_Hunt. | 0.99 | 0.364 | 0.226(284) | 0.246(66192) | −0.0909 | 0.1 |
| dan_daner_MVP. | 0.99 | 0.284 | 0.283(5129) | 0.283(422860) | −0.024 | 0.0224 |
| dan_daner_MoBa. | 0.97 | 0.548 | 0.226(104) | 0.244(2193) | −0.103 | 0.171 |
| dan_daner_NORDiC−NOR. | 0.97 | 0.915 | 0.236(599) | 0.247(619) | −0.0109 | 0.103 |
| dan_daner_NORDiC−SWE. | 0.97 | 0.601 | 0.240(1691) | 0.243(1323) | −0.0329 | 0.0629 |
| dan_daner_Psych_Broad. | 0.96 | 0.0384 | 0.303(1396) | 0.326(4009) | −0.103 | 0.0498 |
| dan_daner_UKBB. | 1.00 | 0.0504 | 0.273(776) | 0.273(125729) | −0.114 | 0.0582 |
| dan_daner_Wue. | 0.97 | 0.893 | 0.284(140) | 0.283(526) | −0.0204 | 0.152 |
| dan_daner_YalePenn_GWCIDR. | 0.99 | 0.417 | 0.283(74) | 0.283(1635) | 0.155 | 0.191 |
| dan_daner_YalePenn_MEGA123. | 1.01 | 0.763 | 0.299(53) | 0.299(1926) | −0.0697 | 0.232 |
| dan_daner_bioVU. | 0.99 | 0.0549 | 0.260(1014) | 0.279(38613) | −0.0976 | 0.0509 |
| dan_daner_iPSYCH. | 0.98 | 0.00888 | 0.245(4509) | 0.257(38392) | −0.0684 | 0.0261 |
| dan_daner_iocd. | 0.98 | 0.0906 | 0.282(1519) | 0.291(3541) | −0.0849 | 0.0502 |
| dan_daner_iocdf_trio. | 0.97 | 0.544 | 0.308(323) | 0.324(323) | −0.0738 | 0.122 |
| dan_daner_ocgas−gh. | 0.88 | 0.87 | 0.270(56) | 0.270(56) | −0.0537 | 0.328 |
| dan_daner_ocgas−nes. | 0.94 | 0.899 | 0.274(212) | 0.272(212) | −0.0205 | 0.161 |
| dan_daner_ocgas. | 0.98 | 0.404 | 0.287(986) | 0.299(1023) | −0.0589 | 0.0706 |
| dan_daner_sickkids. | 1.00 | 0.858 | 0.285(275) | 0.285(205) | 0.0353 | 0.197 |
| **Meta** | **0.99** | **5e−08** | **0.278(53660)** | **0.279(2044417)** | **−0.0399** | **0.0073** |

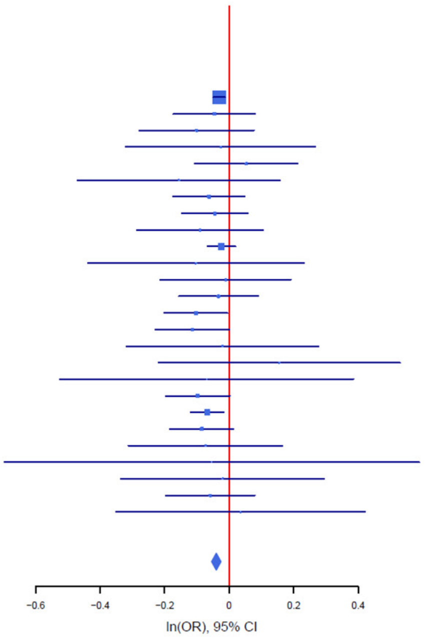

B

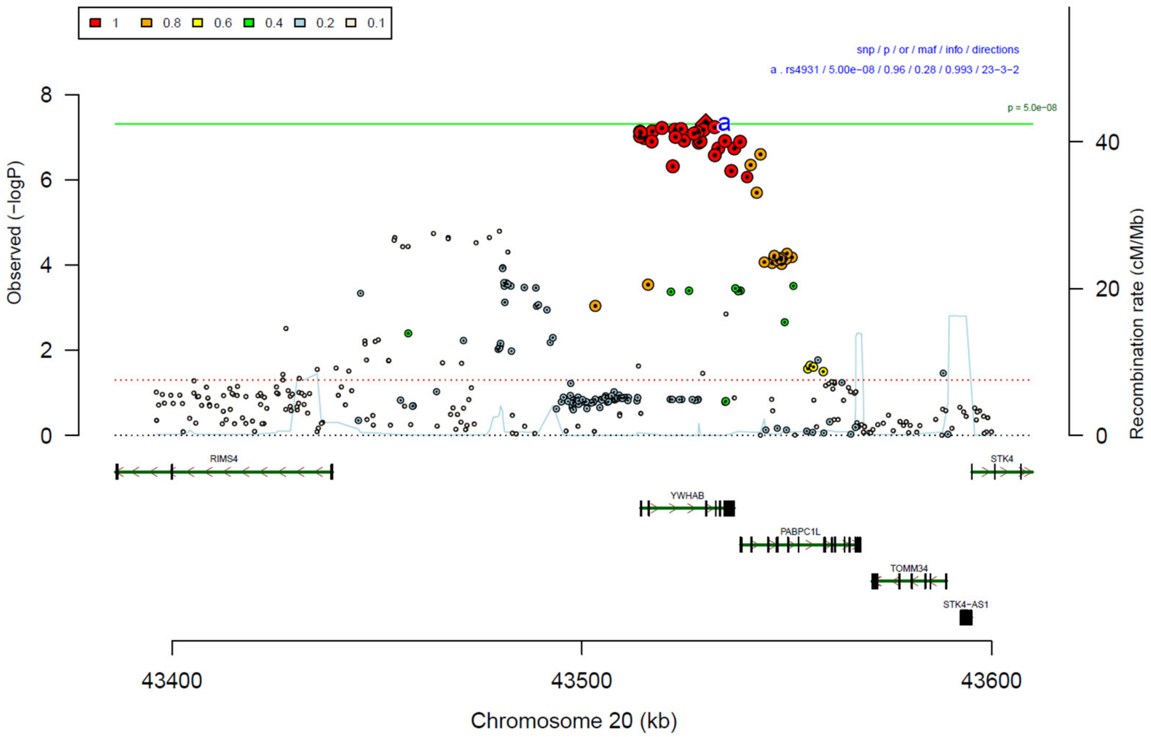

**Supplementary Figure 31: Forest plot (A) and regional association plot (B) of SNP rs4931 in the meta-analysis.** The forest plot (A) shows the imputation quality (INFO) score, p-value of the SNP association (two-sided and not corrected for multiple testing, derived from a fixed-effects inverse-variance weighted meta-analysis), allele frequency in cases with case sample size (f_ca(n)), allele frequency in controls and control sample size (f_co(n)), beta estimates (ln(OR)) and standard error (STDerr) for each study as well as for the combined meta-analysis. Each cohort's sample size is detailed in supplementary table 1. Data on the right show the ln(OR) value as listed per cohort with 95% confidence intervals. In the regional association plot (B), the –log10 p-values (from two-sided association tests) are shown on the left y-axis. Recombination rates expressed in centimorgans (cM) per Mb (Megabase) (blue line) are shown on the right y-axis. Position in Mb is on the x-axis, with genes shown below the regional association plot. Only SNPs with an association p-value less than 0.1 were plotted. The most associated SNP is shown as a diamond and marked with the letter a.

### Manhattan-plots and QQ-plots of sub-analyses

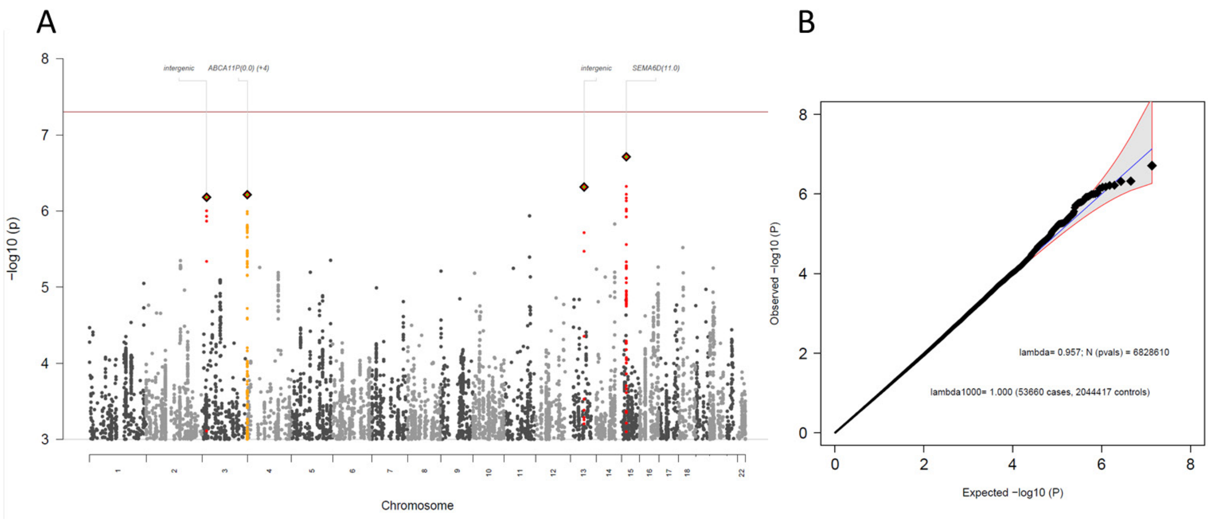

**Supplementary Figure 32: Manhattan-plot (A) and QQ-plot (B) of heterogeneity test**, indicating whether SNPs across the genome are significantly heterogeneously associated with some cohorts but not others. METAL implements Cochran's Q-test for heterogeneity based on a chi-square distribution; it generates a probability that, when large, indicates larger variation across studies rather than within subjects within a study.

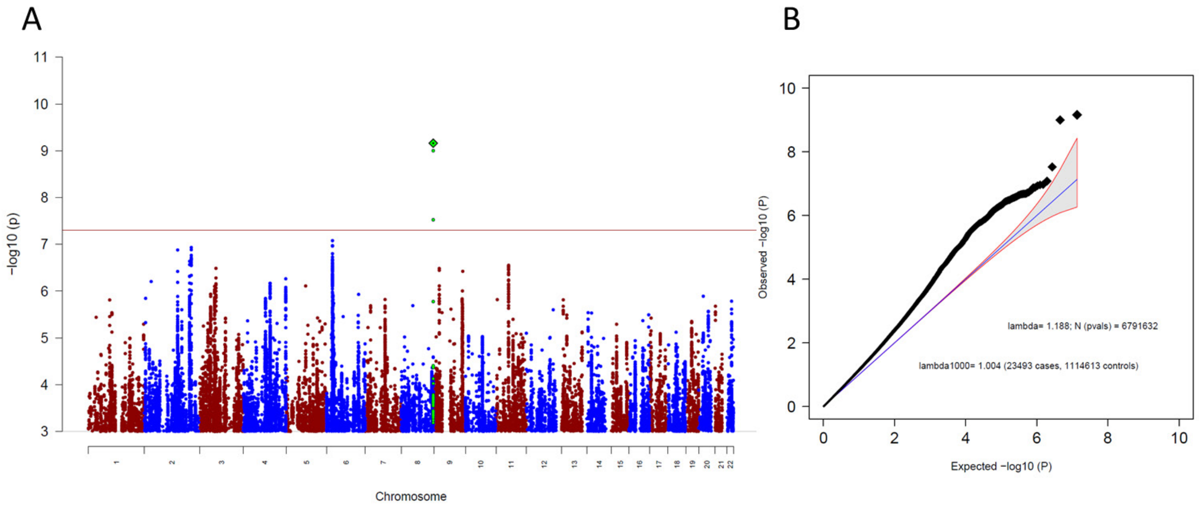

**Supplementary Figure 33: Manhattan-plot (A) and QQ-plot (B) of the genome-wide association results for the OCD meta-analysis leaving out 23andMe.** (A) The x-axis shows the position in the genome (chromosome 1 to 22), the y-axis represents –log10 p-values for the association of variants with OCD from metaanalysis using an inverse-variance weighted fixed effects model. The horizontal red line shows the threshold for genome-wide significance ($5x10^{-08}$). Each dot represents one SNP that was tested in the GWAS, with a green diamond indicating the lead SNP of a genome-wide significant locus, with green dots below belonging to that locus. (B) the expected -log10(p) under the null is plotted against the observed -log10(p). The shading indicates the 95% confidence region under the null. Lambda and Lambda1000 (which is the Lambda if the GWAS contained 1000 cases and 1000 controls) indicate genomic inflation factors. Number of SNPs (N (pvals)) and Number of cases and controls are given in parentheses.

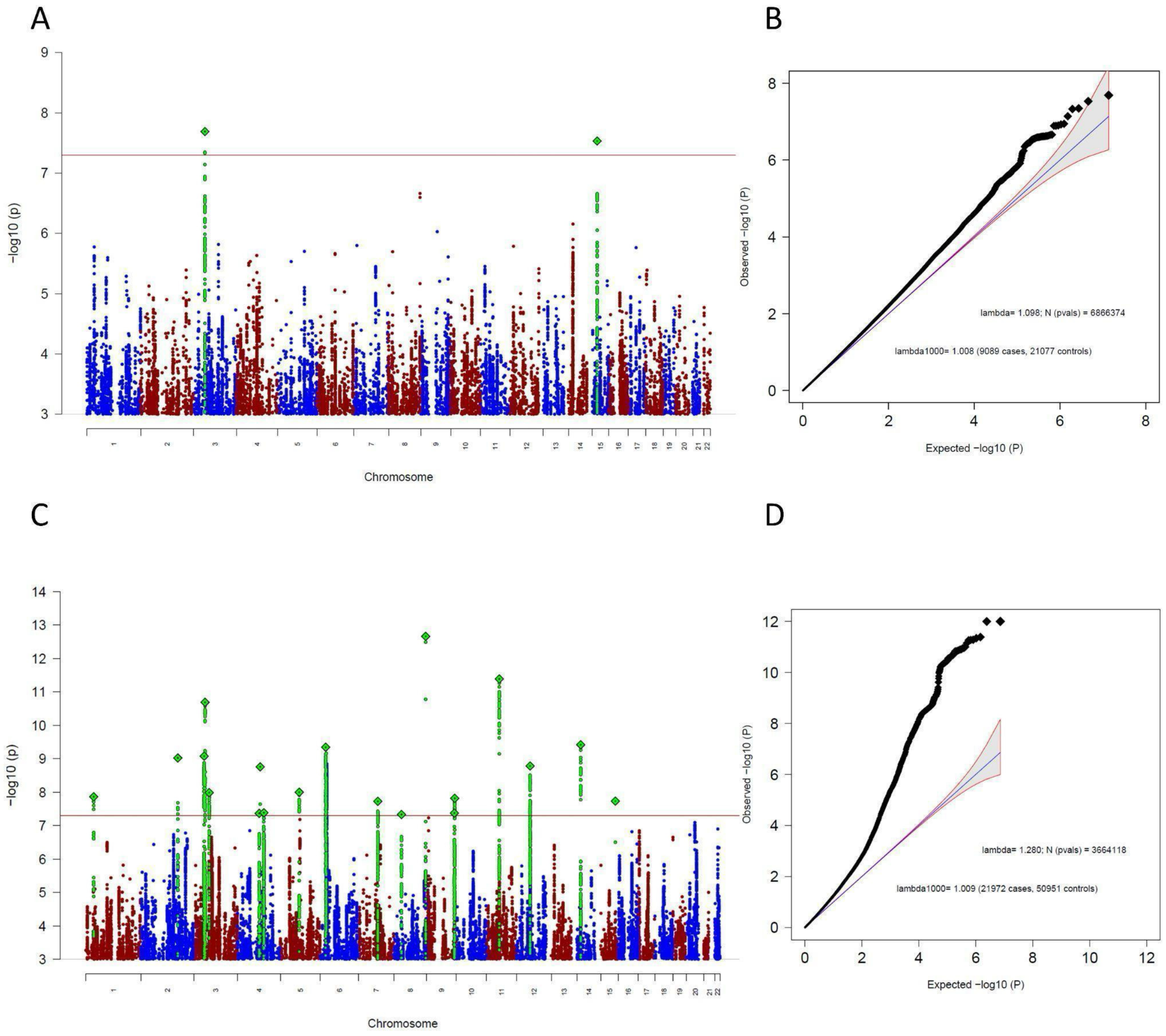

**Supplementary Figure 34: Manhattan-plot (A) and QQ-plot (B) of the sub-group specific GWAS analysis, including only clinical cohorts and Manhattan-plot (C) and QQ-plot (D) of the MTAG analysis specific to clinical cohorts.** Both analyses aimed to find ascertainment-specific signals associated with OCD specifically in clinical cohorts. (A,C) The x-axis shows the position in the genome (chromosome 1 to 22), the y-axis represents –log10 p-values for the association of variants with OCD from metaanalysis using an inverse-variance weighted fixed effects model (A) or MTAG (C). The horizontal red line shows the threshold for genome-wide significance ($5x10^{-08}$). Each dot represents one SNP that was tested in the GWAS, with a green diamond indicating the lead SNP of a genome-wide significant locus, with green dots below belonging to that locus. (B) the expected -log10(p) under the null is plotted against the observed -log10(p). The shading indicates the 95% confidence region under the null. Lambda and Lambda1000 (which is the Lambda if the GWAS contained 1000 cases and 1000 controls) indicate genomic inflation factors. Number of SNPs (N (pvals)) and Number of cases and controls are given in parentheses.

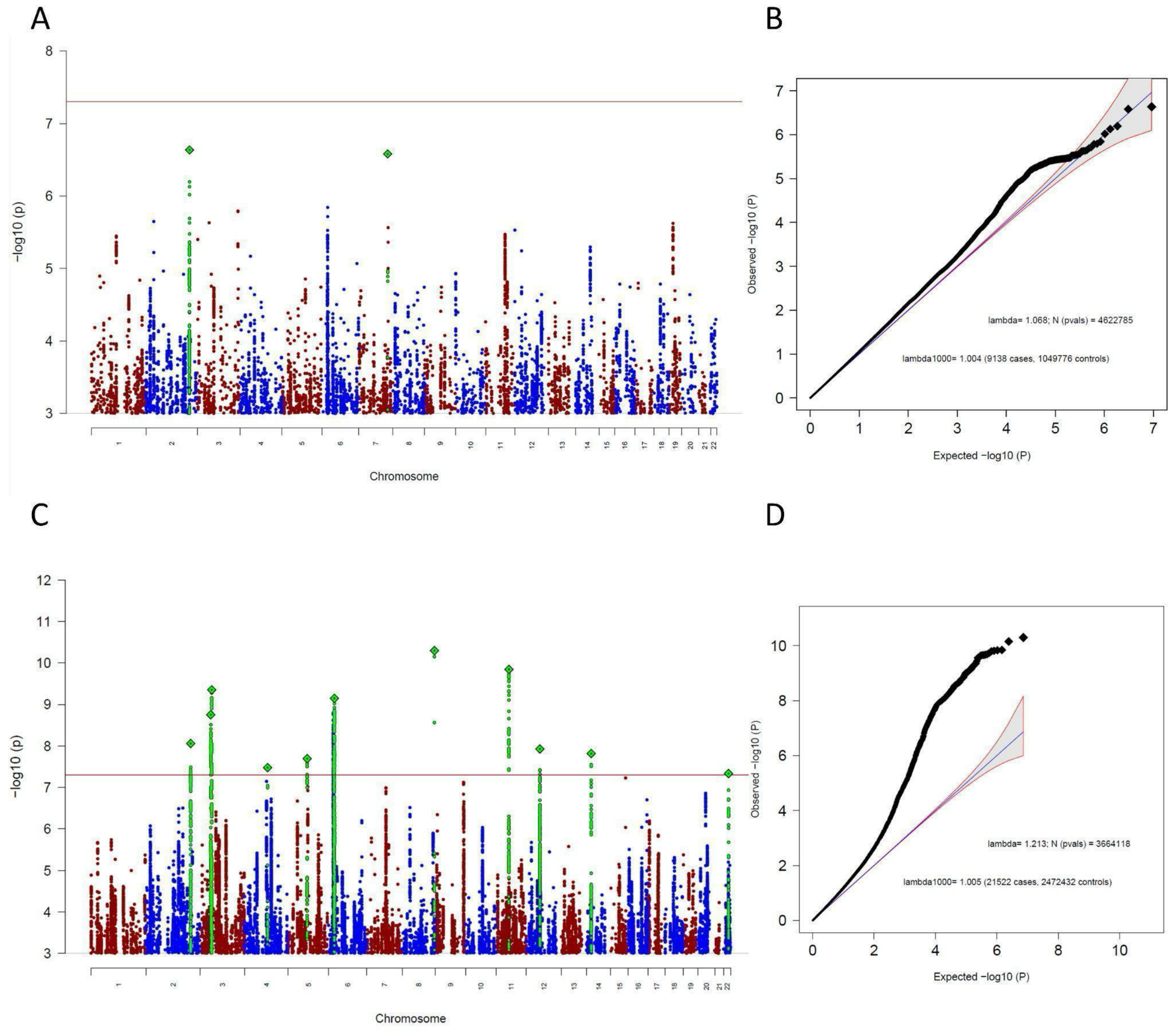

**Supplementary Figure 35: Manhattan-plot (A) and QQ-plot (B) of the sub-group specific GWAS analysis, including only biobank cohorts and Manhattan-plot (C) and QQ-plot (D) of the MTAG analysis specific to biobank cohorts.** Both analyses aimed to find ascertainment-specific signals associated with OCD specifically in biobank cohorts. (A,C) The x-axis shows the position in the genome (chromosome 1 to 22), the y-axis represents –log10 p-values for the association of variants with OCD from metaanalysis using an inverse-variance weighted fixed effects model (A) or MTAG (C). The horizontal red line shows the threshold for genome-wide significance ($5x10^{-08}$). Each dot represents one SNP that was tested in the GWAS, with a green diamond indicating the lead SNP of a genome-wide significant locus, with green dots below belonging to that locus. (B) the expected -log10(p) under the null is plotted against the observed -log10(p). The shading indicates the 95% confidence region under the null. Lambda and Lambda1000 (which is the Lambda if the GWAS contained 1000 cases and 1000 controls) indicate genomic inflation factors. Number of SNPs (N (pvals)) and Number of cases and controls are given in parentheses.

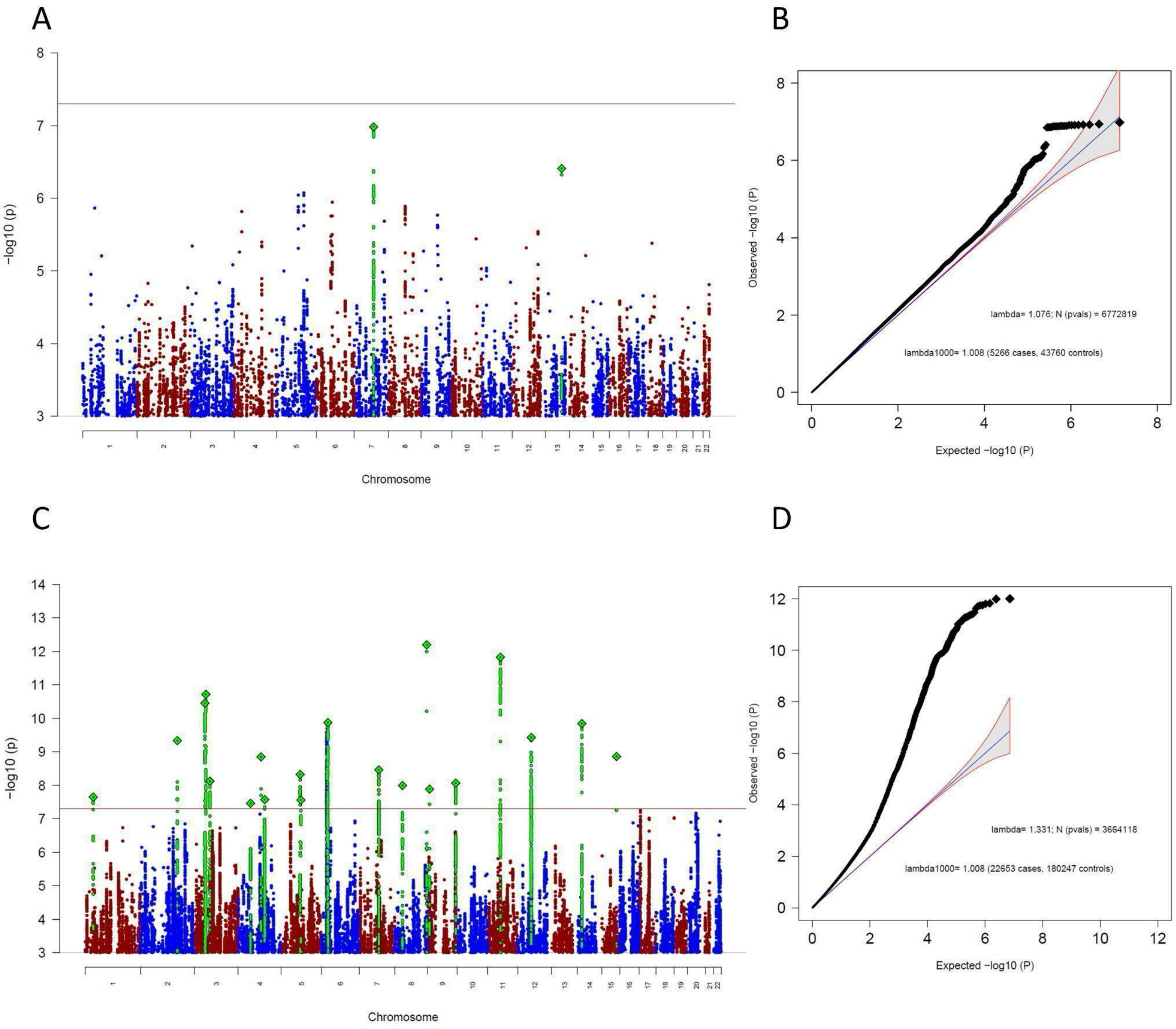

**Supplementary Figure 36: Manhattan-plot (A) and QQ-plot (B) of the sub-group specific GWAS analysis, including only comorbid cohorts and Manhattan-plot (C) and QQ-plot (D) of the MTAG analysis specific to comorbid cohorts.** Both analyses aimed to find ascertainment-specific signals associated with OCD specifically in comorbid cohorts. (A,C) The x-axis shows the position in the genome (chromosome 1 to 22), the y-axis represents –log10 p-values for the association of variants with OCD from metaanalysis using an inverse-variance weighted fixed effects model (A) or MTAG (C). The horizontal red line shows the threshold for genome-wide significance ($5x10^{-08}$). Each dot represents one SNP that was tested in the GWAS, with a green diamond indicating the lead SNP of a genome-wide significant locus, with green dots below belonging to that locus. (B) the expected -log10(p) under the null is plotted against the observed -log10(p). The shading indicates the 95% confidence region under the null. Lambda and Lambda1000 (which is the Lambda if the GWAS contained 1000 cases and 1000 controls) indicate genomic inflation factors. Number of SNPs (N (pvals)) and Number of cases and controls are given in parentheses.

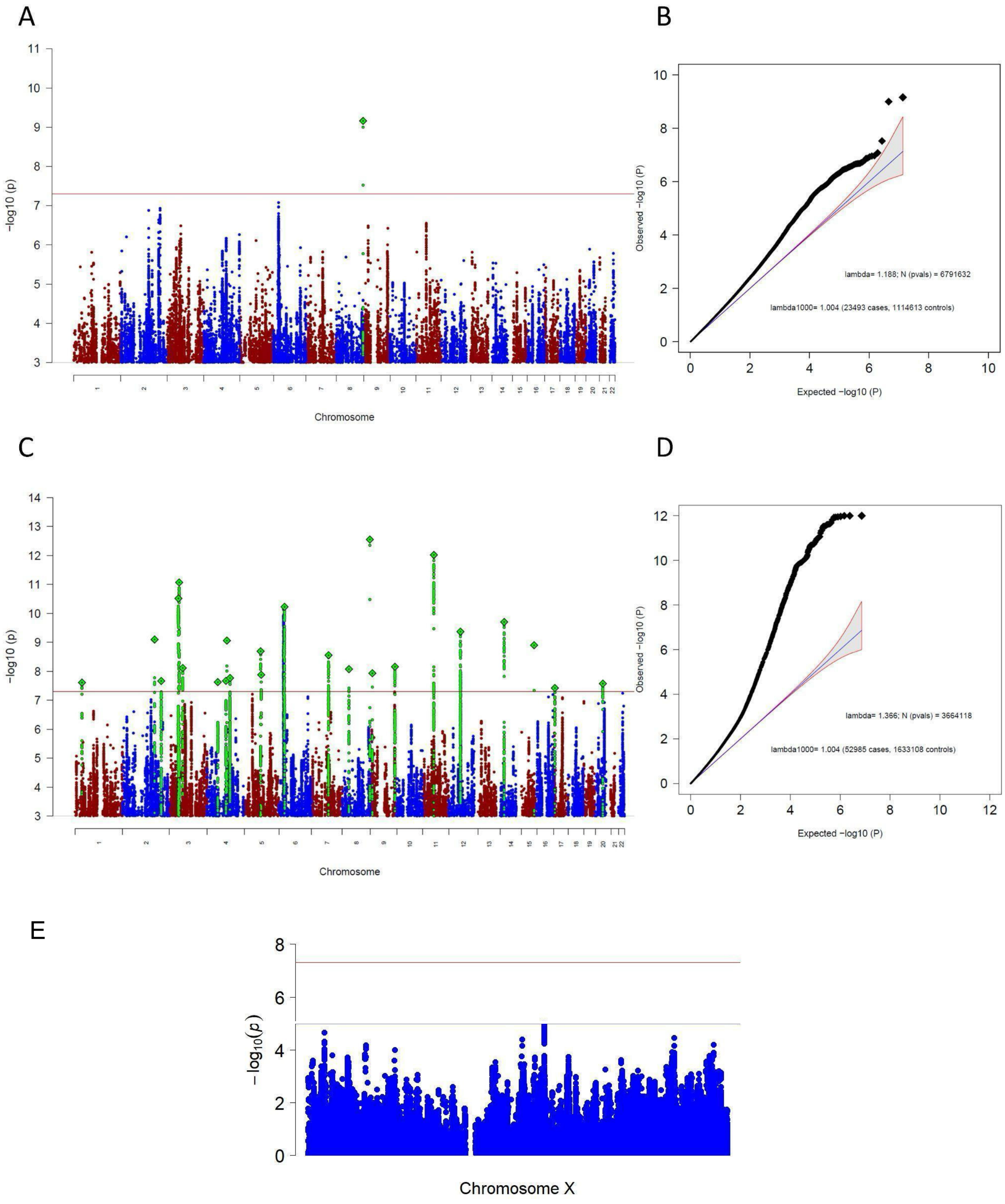

**Supplementary Figure 37: Manhattan-plot (A) and QQ-plot (B) of the sub-group specific GWAS analysis, including only 23andMe, Manhattan-plot (C) and QQ-plot (D) of the MTAG analysis specific to 23andMe, and (E) a plot of the X-chromosome findings specific to 23andMe.** Both analyses (A-D) aimed to find ascertainment-specific signals associated with OCD specifically in 23andMe. (A,C) The x-axis shows the position in the genome (chromosome 1 to 22), the y-axis represents –log10 p-values for the association of variants with OCD from metaanalysis using an inverse-variance weighted fixed effects model (A) or MTAG (C). The horizontal red line shows the threshold for genome-wide significance ($5x10^{-08}$). Each dot represents one SNP that was tested in the GWAS, with a green diamond indicating the lead SNP of a genome-wide significant locus, with green dots below belonging to that locus. (B) the expected -log10(p) under the null is plotted against the observed -log10(p). The shading indicates the 95% confidence region under the null. Lambda and Lambda1000 (which is the Lambda if the GWAS contained 1000 cases and 1000 controls) indicate genomic inflation factors. Number of SNPs (N (pvals)) and Number of cases and controls are given in parentheses.

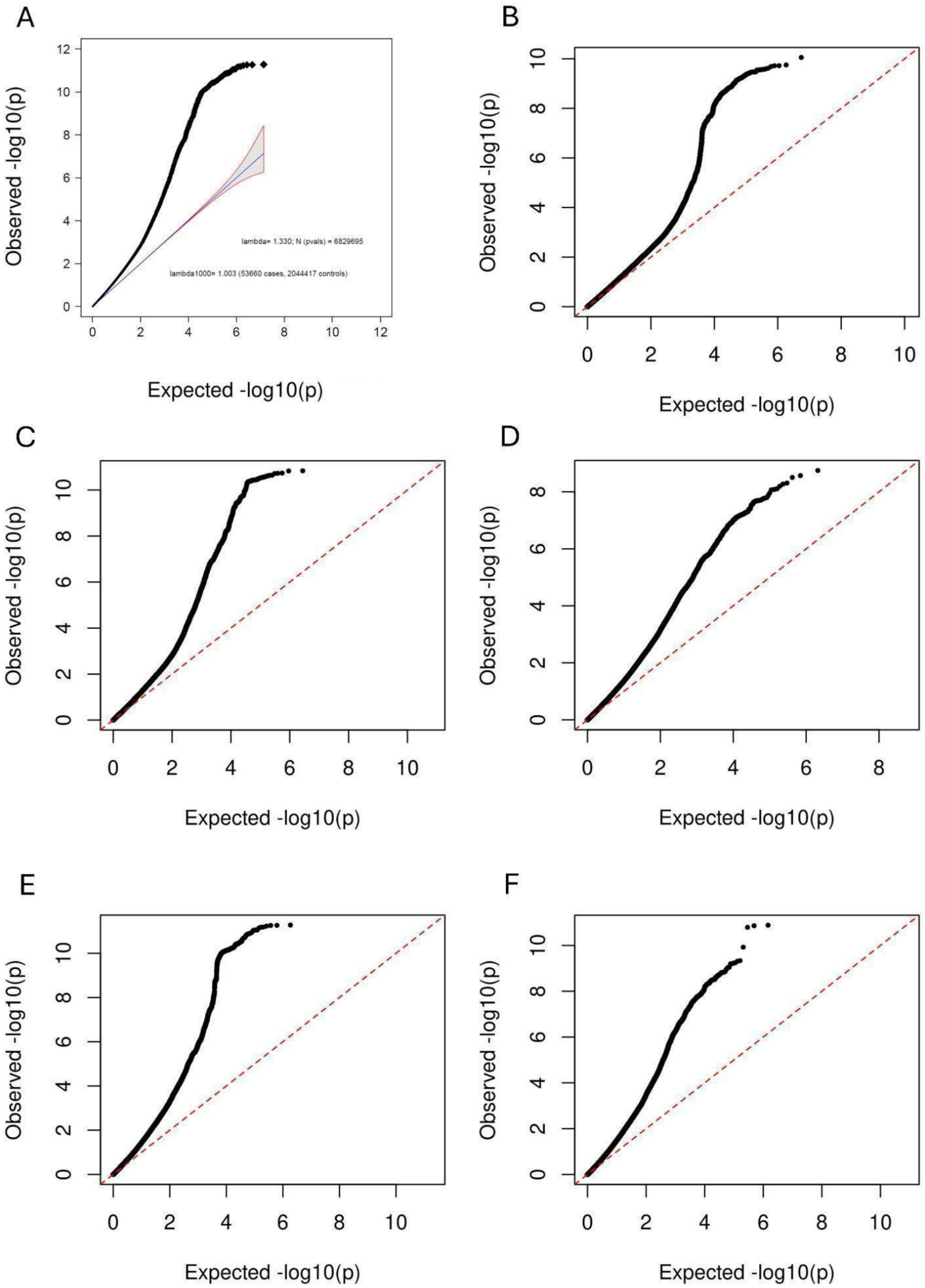

**Supplementary Figure 38: QQ-plot for main GWAS results. (A) across the entire allele-frequency spectrum; (B)-(F) partitioned by MAF bins (B) 0.01-0.1, (C) 0.1-0.2, (D) 0.2-0.3, (E) 0.3-0.4, (F) 0.4-0.5.** For each bin, SNPs were included if their MAF in the overall analysis fell within that range. The expected -log10(p) values under the null hypothesis are plotted against the observed -log10(p) values.

### GenomicSEM one factor model of the four OCD ascertainment sub-groups

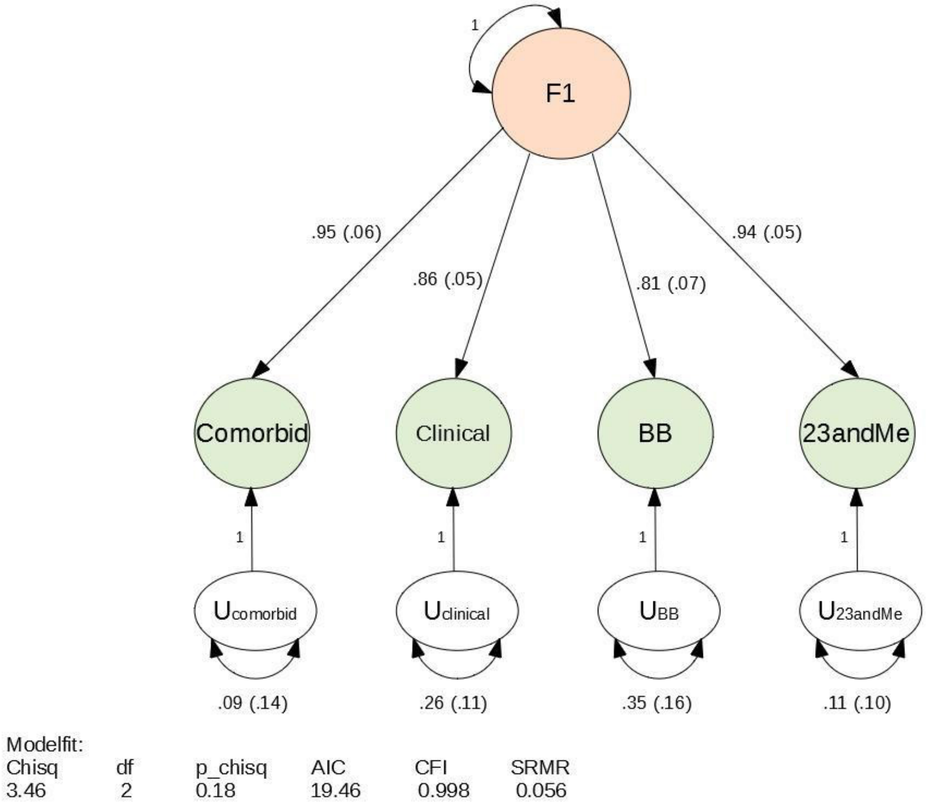

**Supplementary Figure 39: A path diagram of the common-factor GenomicSEM model without SNP effects, specified with unit variance identification, fixing the variance of the common factor F1 to 1.** All estimates are standardized. Model-fit indices below are Chi-square statistic (Chisq); degrees of freedom of the model (df); p-value of the Chi-square (p_chisq); Akaike Information Criterion (AIC), which is a comparative measure of fit with lower values indicating a better fit; comparative fit index (CFI) which assesses the relative improvement in fit compared with the baseline model, ranging between 0 and 1; and standardized root mean square residual (SRMR), which is an absolute measure of fit defined as the standardized difference between observed correlation and the predicted correlation with a value of 0 indicating perfect fit.

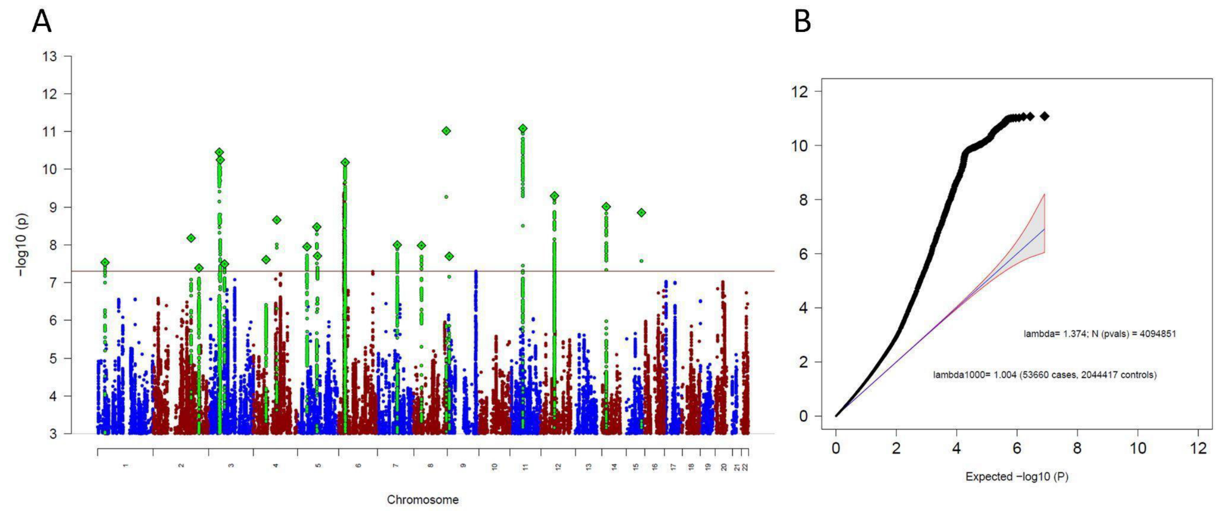

**Supplementary Figure 40: Manhattan-plot (A) and QQ-plot (B) of the common-factor GenomicSEM GWAS as shown in Supplementary Figure 39.** (A) The x-axis shows the position in the genome (chromosome 1 to 22), the y-axis represents –log10 p-values for the association of variants with OCD (A). The horizontal red line shows the threshold for genome-wide significance ($5x10^{-08}$). Each dot represents one SNP that was tested in the GWAS, with a green diamond indicating the lead SNP of a genome-wide significant locus, with green dots below belonging to that locus. (B) the expected -log10(p) under the null is plotted against the observed -log10(p). The shading indicates the 95% confidence region under the null. Lambda and Lambda1000 (which is the Lambda if the GWAS contained 1000 cases and 1000 controls) indicate genomic inflation factors. Number of SNPs (N (pvals)) and Number of cases and controls are given in parentheses.

### Cross-trait genetic correlations of the ascertainment-specific subgroup analyses

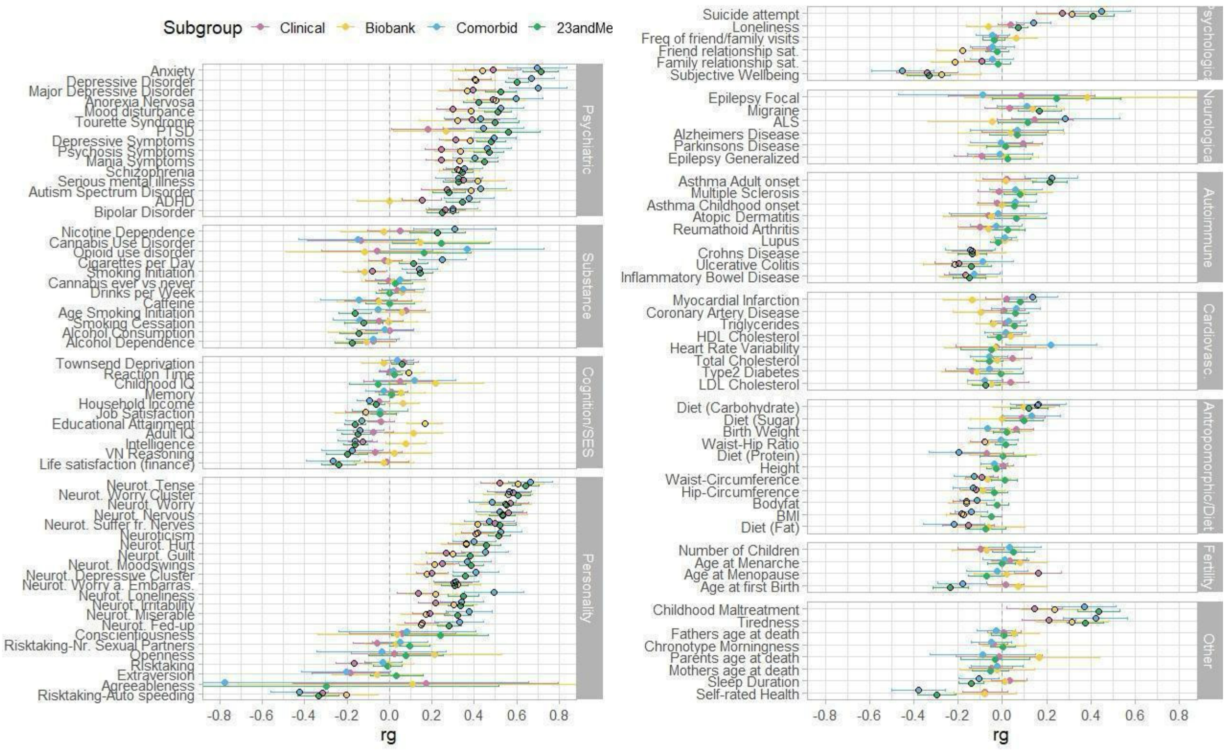

**Supplementary Figure 41: Genetic correlations (rg) between the ascertainment-specific OCD GWASs (23andMe, Biobank, Clinical, comorbid) and 112 psychiatric, substance use, cognition/socioeconomic status (SES), personality, psychological, neurological, autoimmune, cardiovascular, anthropomorphic/diet, fertility, and other phenotypes**. References and sample sizes of the corresponding summary statistics of the GWAS studies can be found in **Supplementary Table 19**. The OCD summary statistics are of the main meta-analysis (Ncases = 53,660 and Ncontrols = 2,044,417). Genetic correlations of OCD specific to 23andMe are in green, specific to biobanks are in yellow, specific to clinical cohorts are in pink, and specific to comorbid cohorts are in blue. Error bars represent 95% confidence intervals for the genetic correlation estimates (rg), black encircled estimates indicate significant associations with a p-value adjusted for multiple testing with the Benjamini-Hochberg procedure to control the FDR (< 0.05), adjusted separately for each OCD ascertainment-specific subgroup (see full list of results in Supplementary Table 18).

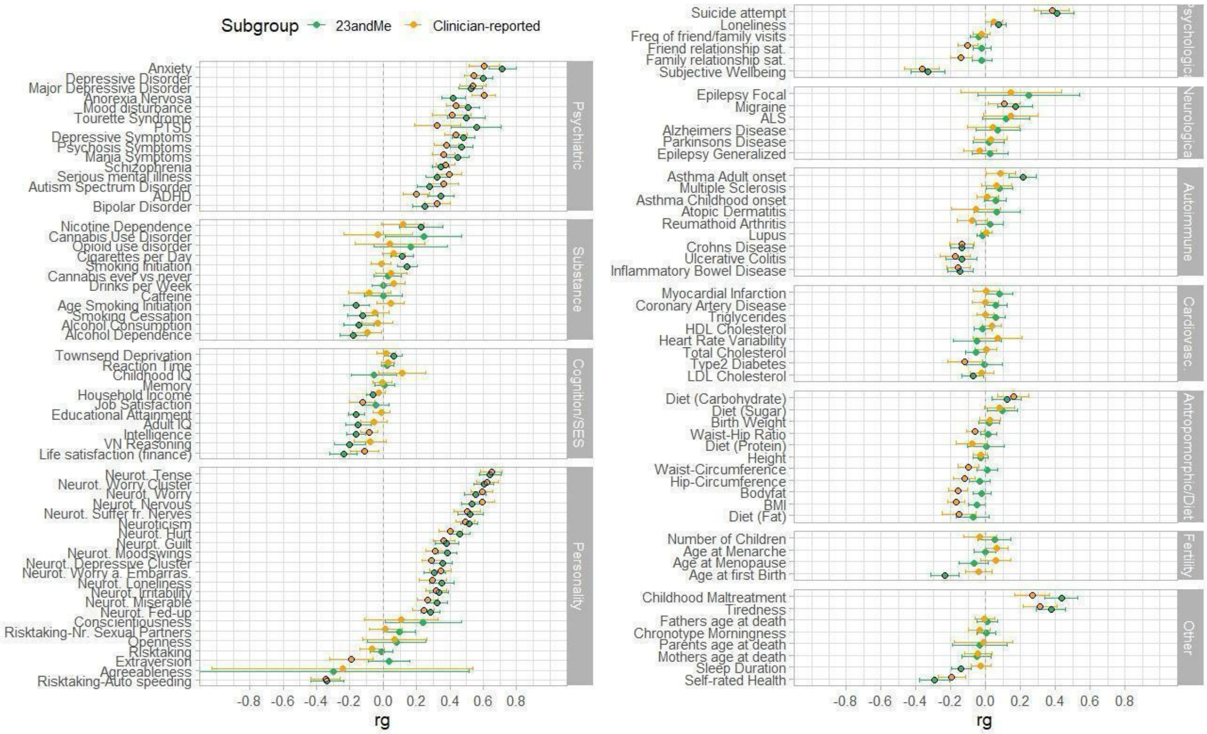

**Supplementary Figure 42: Genetic correlations (rg) of the OCD GWASs including only 23andMe and the OCD GWAS leaving out 23andMe (Clinician-reported) with 112 psychiatric, substance use, cognition/socioeconomic status (SES), personality, psychological, neurological, autoimmune, cardiovascular, anthropomorphic/diet, fertility, and other phenotypes**. References and sample sizes of the corresponding summary statistics of the GWAS studies can be found in **Supplementary Table 19**. The OCD summary statistics are of the main meta-analysis (Ncases = 53,660 and Ncontrols = 2,044,417). Genetic correlations of OCD specific to 23andMe are in green, specific to the GWAS excluding 23andMe (Clinician-reported) in orange. Error bars represent 95% confidence intervals for the genetic correlation estimates (rg), black encircled estimates indicate significant associations with a p-value adjusted for multiple testing with the Benjamini-Hochberg procedure to control the FDR (< 0.05), adjusted separately for each OCD subgroup.

### Consistency of SNP findings across OCD GWAS versions

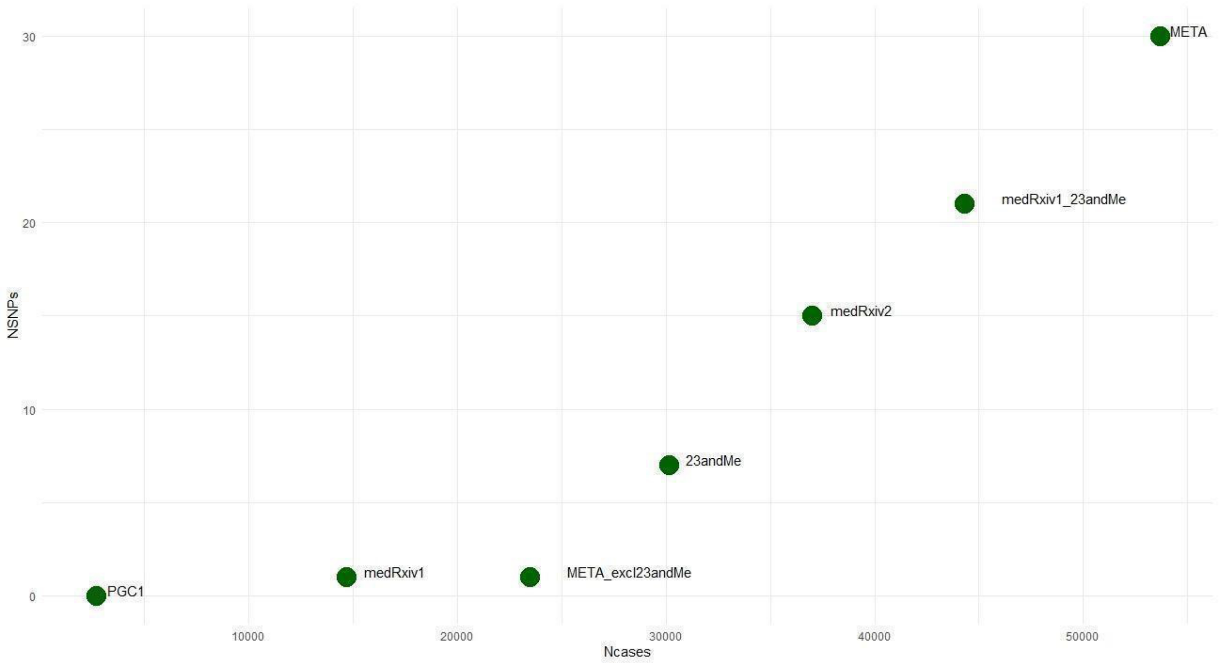

**Supplementary Figure 43: Number of GWAS cases (x-axis) and the respective number of independent significant SNPs they identified (y-axis).** Shown are seven GWAS studies - PGC1 (IOCDF-GC and OCGAS, 2018) ; PGC OCD2 GWAS (medRxiv1; Strom et al. 2021); the meta-analysis of the current paper (META) excluding 23andMe (META_excl23andMe); 23andMe, iPSYCH-PGC-23andMe GWAS (medRxiv2; Strom et al. 2024), PGC OCD2 GWAS and 23andMe (medRxiv1_23andMe), and the current meta-analysis (META). Please also see **Supplementary Table 20** for details. Note that these iterations of OCD GWAS meta-analyses are not independent of each other. Cohorts with fewer cases (to the left of the plot) are included in meta-analyses with more cases (to the right of the plot). An overview of the significant SNPs from the published medRxiv GWASs (medRxiv1 and medRxiv2) and their corresponding p-values in the current OCD GWAS can be found in **Supplementary Table 5**.

### References

Abraham, G., & Inouye, M. (2014). Fast Principal Component Analysis of Large-Scale Genome-Wide Data. *PLOS ONE*, *9*(4), e93766. https://doi.org/10.1371/journal.pone.0093766

Altshuler, D. M., Gibbs, R. A., Peltonen, L., Schaffner, S. F., Yu, F., Dermitzakis, E., Bonnen, P. E., De Bakker, P. I. W., Deloukas, P., Gabriel, S. B., Gwilliam, R., Hunt, S., Inouye, M., Jia, X., Aarno Palotie, Parkin, M., Whittaker, P., Chang, K., Hawes, A., … McEwen, J. E. (2010). Integrating common and rare genetic variation in diverse human populations. *Nature*, *467*(7311), 52–58. https://doi.org/10.1038/nature09298

Arnold, P. D., Askland, K. D., Barlassina, C., Bellodi, L., Bienvenu, O. J., Black, D., Bloch, M., Brentani, H., Burton, C. L., Camarena, B., Cappi, C., Cath, D., Cavallini, M., Conti, D., Cook, E., Coric, V., Cullen, B. A., Cusi, D., Davis, L. K., … Zai, G. (2018). Revealing the complex genetic architecture of obsessive-compulsive disorder using meta-analysis. *Molecular Psychiatry*, *23*(5). https://doi.org/10.1038/mp.2017.154

Bosch, R., Pagerols, M., Rivas, C., Sixto, L., Bricollé, L., Español-Martín, G., Prat, R., Ramos-Quiroga, J. A., & Casas, M. (2021). Neurodevelopmental disorders among Spanish school-age children: Prevalence and sociodemographic correlates. *Psychological Medicine*, 1–11. https://doi.org/10.1017/S0033291720005115

Browning, B. L. (2017). Beagle 4.1. *Https://Faculty.Washington.Edu/Browning/Beagle/Beagle_4.1_21Jan17.Pdf*. https://doi.org/10.1086/521987

Browning, B. L., Zhou, Y., & Browning, S. R. (2018). A One-Penny Imputed Genome from Next-Generation Reference Panels. *American Journal of Human Genetics*, *103*(3), 338–348. https://doi.org/10.1016/j.ajhg.2018.07.015

Browning, S. R., & Browning, B. L. (2007). Rapid and accurate haplotype phasing and missing-data inference for whole-genome association studies by use of localized haplotype clustering. *American Journal of Human Genetics*, *81*(5). https://doi.org/10.1086/521987

Burton, C. L., Lemire, M., Xiao, B., Corfield, E. C., Erdman, L., Bralten, J., Poelmans, G., Yu, D., Shaheen, S. M., Goodale, T., Sinopoli, V. M., Askland, K. D., Barlassina, C., Bienvenu, O. J., Black, D., Bloch, M., Brentani, H., Camarena, B., Cappi, C., … Arnold, P. D. (2021). Genome-wide association study of pediatric obsessive-compulsive traits: Shared genetic risk between traits and disorder. *Translational Psychiatry 2021 11:1*, *11*(1), 1–10. https://doi.org/10.1038/s41398-020-01121-9

Bybjerg-Grauholm, J., Pedersen, C. B., Bækvad-Hansen, M., Pedersen, M. G., Adamsen, D., Hansen, C. S., Agerbo, E., Grove, J., Als, T. D., Schork, A. J., Buil, A., Mors, O., Nordentoft, M., Werge, T., Børglum, A. D., Hougaard, D. M., & Mortensen, P. B. (2020). *The iPSYCH2015 Case-Cohort sample: Updated directions for unravelling genetic and environmental architectures of severe mental disorders* (p. 2020.11.30.20237768). medRxiv. https://doi.org/10.1101/2020.11.30.20237768

Bycroft, C., Freeman, C., Petkova, D., Band, G., Elliott, L. T., Sharp, K., Motyer, A., Vukcevic, D., Delaneau, O., O'Connell, J., Cortes, A., Welsh, S., Young, A., Effingham, M., McVean, G., Leslie, S., Allen, N., Donnelly, P., & Marchini, J. (2018). The UK Biobank resource with deep phenotyping and genomic data. *Nature 2018 562:7726*, *562*(7726), 203–209. https://doi.org/10.1038/s41586-018-0579-z

Calkins, M. E., Merikangas, K. R., Moore, T. M., Burstein, M., Behr, M. A., Satterthwaite, T. D., Ruparel, K., Wolf, D. H., Roalf, D. R., Mentch, F. D., Qiu, H., Chiavacci, R., Connolly, J. J., Sleiman, P. M. A., Gur, R. C., Hakonarson, H., & Gur, R. E. (2015). The Philadelphia Neurodevelopmental Cohort: Constructing a deep phenotyping collaborative. *Journal of Child Psychology and Psychiatry, and Allied Disciplines*, *56*(12), 1356–1369. https://doi.org/10.1111/jcpp.12416

Calkins, M. E., Moore, T. M., Merikangas, K. R., Burstein, M., Satterthwaite, T. D., Bilker, W. B., Ruparel, K., Chiavacci, R., Wolf, D. H., Mentch, F., Qiu, H., Connolly, J. J., Sleiman, P. A., Hakonarson, H., Gur, R. C., & Gur, R. E. (2014). The psychosis spectrum in a young U.S. community sample: Findings from the Philadelphia Neurodevelopmental Cohort. *World*

*Psychiatry: Official Journal of the World Psychiatric Association (WPA)*, *13*(3), 296–305. https://doi.org/10.1002/wps.20152

Chang, C. C., Chow, C. C., Tellier, L. C. A. M., Vattikuti, S., Purcell, S. M., & Lee, J. J. (2015). Second-generation PLINK: Rising to the challenge of larger and richer datasets. *GigaScience*, *4*(1), 7. https://doi.org/10.1186/S13742-015-0047-8/2707533

Cheng, Z., Zhou, H., Sherva, R., Farrer, L. A., Kranzler, H. R., & Gelernter, J. (2018). Genome-wide Association Study Identifies a Regulatory Variant of RGMA Associated With Opioid Dependence in European Americans. *Biological Psychiatry*, *84*(10), 762–770. https://doi.org/10.1016/j.biopsych.2017.12.016

Das, S., Forer, L., Schönherr, S., Sidore, C., Locke, A. E., Kwong, A., Vrieze, S. I., Chew, E. Y., Levy, S., McGue, M., Schlessinger, D., Stambolian, D., Loh, P. R., Iacono, W. G., Swaroop, A., Scott, L. J., Cucca, F., Kronenberg, F., Boehnke, M., … Fuchsberger, C. (2016). Next-generation genotype imputation service and methods. *Nature Genetics*, *48*(10), 1284–1287. https://doi.org/10.1038/NG.3656

de Klein, N., Tsai, E. A., Vochteloo, M., Baird, D., Huang, Y., Chen, C.-Y., van Dam, S., Oelen, R., Deelen, P., Bakker, O. B., El Garwany, O., Ouyang, Z., Marshall, E. E., Zavodszky, M. I., van Rheenen, W., Bakker, M. K., Veldink, J., Gaunt, T. R., Runz, H., … Westra, H.-J. (2023). Brain expression quantitative trait locus and network analyses reveal downstream effects and putative drivers for brain-related diseases. *Nature Genetics*, *55*(3), Article 3. https://doi.org/10.1038/s41588-023-01300-6

Delaneau, O., Marchini, J., & Zagury, J. F. (2011). A linear complexity phasing method for thousands of genomes. *Nature Methods 2011 9:2*, *9*(2), 179–181. https://doi.org/10.1038/nmeth.1785

Delaneau, O., Zagury, J.-F., & Marchini, J. (2013). Improved whole-chromosome phasing for disease and population genetic studies. *Nature Methods*, *10*(1), Article 1. https://doi.org/10.1038/nmeth.2307

Dunham, I., Kundaje, A., Aldred, S. F., Collins, P. J., Davis, C. A., Doyle, F., Epstein, C. B., Frietze, S., Harrow, J., Kaul, R., Khatun, J., Lajoie, B. R., Landt, S. G., Lee, B. K., Pauli, F.,

Rosenbloom, K. R., Sabo, P., Safi, A., Sanyal, A., … Lochovsky, L. (2012). An integrated encyclopedia of DNA elements in the human genome. *Nature*, *489*(7414), 57–74. https://doi.org/10.1038/nature11247

Durbin, R. M., Altshuler, D., Durbin, R. M., Abecasis, G. R., Bentley, D. R., Chakravarti, A., Clark, A. G., Collins, F. S., De La Vega, F. M., Donnelly, P., Egholm, M., Flicek, P., Gabriel, S. B., Gibbs, R. A., Knoppers, B. M., Lander, E. S., Lehrach, H., Mardis, E. R., McVean, G. A., … The Translational Genomics Research Institute. (2010). A map of human genome variation from population-scale sequencing. *Nature*, *467*(7319), Article 7319. https://doi.org/10.1038/nature09534

Evans, L. M., Tahmasbi, R., Vrieze, S. I., Abecasis, G. R., Das, S., Gazal, S., Bjelland, D. W., de Candia, T. R., Haplotype Reference Consortium, Goddard, M. E., Neale, B. M., Yang, J., Visscher, P. M., & Keller, M. C. (2018). Comparison of methods that use whole genome data to estimate the heritability and genetic architecture of complex traits. Nature genetics, 50(5), 737–745. https://doi.org/10.1038/s41588-018-0108-x

Gaziano, J. M., Concato, J., Brophy, M., Fiore, L., Pyarajan, S., Breeling, J., Whitbourne, S., Deen, J., Shannon, C., Humphries, D., Guarino, P., Aslan, M., Anderson, D., LaFleur, R., Hammond, T., Schaa, K., Moser, J., Huang, G., Muralidhar, S., … O'Leary, T. J. (2016). Million Veteran Program: A mega-biobank to study genetic influences on health and disease. *Journal of Clinical Epidemiology*, *70*, 214–223. https://doi.org/10.1016/j.jclinepi.2015.09.016

Gazzellone, M. J., Zarrei, M., Burton, C. L., Walker, S., Uddin, M., Shaheen, S. M., Coste, J., Rajendram, R., Schachter, R. J., Colasanto, M., Hanna, G. L., Rosenberg, D. R., Soreni, N., Fitzgerald, K. D., Marshall, C. R., Buchanan, J. A., Merico, D., Arnold, P. D., & Scherer, S. W. (2016). Uncovering obsessive-compulsive disorder risk genes in a pediatric cohort by high-resolution analysis of copy number variation. *Journal of Neurodevelopmental Disorders*, *8*(1), 1–10. https://doi.org/10.1186/s11689-016-9170-9

Goldstein, J. I., Crenshaw, A., Carey, J., Grant, G. B., Maguire, J., Fromer, M., O'Dushlaine, C., Moran, J. L., Chambert, K., Stevens, C., Swedish Schizophrenia Consortium, ARRA

Autism Sequencing Consortium, Sklar, P., Hultman, C. M., Purcell, S., McCarroll, S. A., Sullivan, P. F., Daly, M. J., & Neale, B. M. (2012). zCall: A rare variant caller for array-based genotyping: genetics and population analysis. *Bioinformatics (Oxford, England)*, *28*(19), 2543–2545. https://doi.org/10.1093/bioinformatics/bts479

Grotzinger, A.D., Rhemtulla, M., de Vlaming, R. et al. Genomic structural equation modelling provides insights into the multivariate genetic architecture of complex traits. Nat Hum Behav 3, 513–525 (2019). https://doi.org/10.1038/s41562-019-0566-x

Guo, Y., He, J., Zhao, S., Wu, H., Zhong, X., Sheng, Q., Samuels, D. C., Shyr, Y., & Long, J. (2014). Illumina human exome genotyping array clustering and quality control. *Nature Protocols*, *9*(11), 2643–2662. https://doi.org/10.1038/nprot.2014.174

Gur, R. C., Richard, J., Calkins, M. E., Chiavacci, R., Hansen, J. A., Bilker, W. B., Loughead, J., Connolly, J. J., Qiu, H., Mentch, F. D., Abou-Sleiman, P. M., Hakonarson, H., & Gur, R. E. (2012). Age group and sex differences in performance on a computerized neurocognitive battery in children age 8-21. *Neuropsychology*, *26*(2), 251–265. https://doi.org/10.1037/a0026712

Halvorsen M, Samuels J, Wang Y, Greenberg BD, Fyer AJ, McCracken JT, *et al.* (2021): Exome sequencing in obsessive-compulsive disorder reveals a burden of rare damaging coding variants. *Nat Neurosci* 24: 1071–1076.

Harrington, K. M., Nguyen, X.-M. T., Song, R. J., Hannagan, K., Quaden, R., Gagnon, D. R., Cho, K., Deen, J. E., Muralidhar, S., O'Leary, T. J., Gaziano, J. M., Whitbourne, S. B., & VA Million Veteran Program. (2019). Gender Differences in Demographic and Health Characteristics of the Million Veteran Program Cohort. *Women's Health Issues: Official Publication of the Jacobs Institute of Women's Health*, *29 Suppl 1*, S56–S66. https://doi.org/10.1016/j.whi.2019.04.012

Helgeland, Ø., Vaudel, M., Juliusson, P. B., Lingaas Holmen, O., Juodakis, J., Bacelis, J., Jacobsson, B., Lindekleiv, H., Hveem, K., Lie, R. T., Knudsen, G. P., Stoltenberg, C., Magnus, P., Sagen, J. V., Molven, A., Johansson, S., & Njølstad, P. R. (2019). Genome-wide association study reveals dynamic role of genetic variation in infant and early childhood growth. *Nature Communications*, *10*(1), 1–10. https://doi.org/10.1038/s41467-019-12308-0

Howie, B. N., Donnelly, P., & Marchini, J. (2009). A Flexible and Accurate Genotype Imputation Method for the Next Generation of Genome-Wide Association Studies. *PLOS Genetics*, *5*(6), e1000529. https://doi.org/10.1371/journal.pgen.1000529

Hunter-Zinck, H., Shi, Y., Li, M., Gorman, B. R., Ji, S.-G., Sun, N., Webster, T., Liem, A., Hsieh, P., Devineni, P., Karnam, P., Gong, X., Radhakrishnan, L., Schmidt, J., Assimes, T. L., Huang, J., Pan, C., Humphries, D., Brophy, M., … Pyarajan, S. (2020). Genotyping Array Design and Data Quality Control in the Million Veteran Program. *American Journal of Human Genetics*, *106*(4), 535–548. https://doi.org/10.1016/j.ajhg.2020.03.004

Hyde, C., Nagle, M., Tian, C., Chen, X., Paciga, S., Wendland, J., Tung, J., Hinds, D., Perlis, R., & Winslow, A. (2016). Identification of 15 genetic loci associated with risk of major depression in individuals of European descent. *Nature Genetics*, *48*(9), 1031–1036. https://doi.org/10.1038/NG.3623

International Obsessive Compulsive Disorder Foundation Genetics Collaborative (IOCDF-GC) and OCD Collaborative Genetics Association Studies (OCGAS) (2018). Revealing the complex genetic architecture of obsessive-compulsive disorder using meta-analysis. Molecular psychiatry, 23(5), 1181–1188. https://doi.org/10.1038/mp.2017.154

Jun, G., Flickinger, M., Hetrick, K. N., Romm, J. M., Doheny, K. F., Abecasis, G. R., Boehnke, M., & Kang, H. M. (2012). Detecting and estimating contamination of human DNA samples in sequencing and array-based genotype data. *American Journal of Human Genetics*, *91*(5), 839–848. https://doi.org/10.1016/j.ajhg.2012.09.004

Kaufman, J., Birmaher, B., Brent, D. A., Ryan, N. D., & Rao, U. (2000). K-SADS-PL. *Journal of the American Academy of Child & Adolescent Psychiatry*, *39*(10), 1208–1208. https://doi.org/10.1097/00004583-200010000-00002

Korn, J. M., Kuruvilla, F. G., McCarroll, S. A., Wysoker, A., Nemesh, J., Cawley, S., Hubbell, E., Veitch, J., Collins, P. J., Darvishi, K., Lee, C., Nizzari, M. M., Gabriel, S. B., Purcell, S., Daly, M. J., & Altshuler, D. (2008). Integrated genotype calling and association analysis of

SNPs, common copy number polymorphisms and rare CNVs. *Nature Genetics*, *40*(10), Article 10. https://doi.org/10.1038/ng.237

Krokstad, S., Langhammer, A., Hveem, K., Holmen, T. L., Midthjell, K., Stene, T. R., Bratberg, G., Heggland, J., & Holmen, J. (2013). Cohort profile: The HUNT study, Norway. *International Journal of Epidemiology*. https://doi.org/10.1093/ije/dys095

Lai, D., Wetherill, L., Bertelsen, S., Carey, C. E., Kamarajan, C., Kapoor, M., Meyers, J. L., Anokhin, A. P., Bennett, D. A., Bucholz, K. K., Chang, K. K., De Jager, P. L., Dick, D. M., Hesselbrock, V., Kramer, J., Kuperman, S., Nurnberger Jr, J. I., Raj, T., Schuckit, M., … Foroud, T. (2019). Genome-wide association studies of alcohol dependence, DSM-IV criterion count and individual criteria. *Genes, Brain and Behavior*, *18*(6), e12579. https://doi.org/10.1111/gbb.12579

Lai, D., Wetherill, L., Kapoor, M., Johnson, E. C., Schwandt, M., Ramchandani, V. A., Goldman, D., Joslyn, G., Rao, X., Liu, Y., Farris, S., Mayfield, R. D., Dick, D., Hesselbrock, V., Kramer, J., McCutcheon, V. V., Nurnberger, J., Tischfield, J., Goate, A., … Schuckit, M. (2020). Genome-wide association studies of the self-rating of effects of ethanol (SRE). *Addiction Biology*, *25*(2), e12800. https://doi.org/10.1111/adb.12800

Lam, M., Awasthi, S., Watson, H. J., Goldstein, J., Panagiotaropoulou, G., Trubetskoy, V., Karlsson, R., Frei, O., Fan, C. C., De Witte, W., Mota, N. R., Mullins, N., Brügger, K., Hong Lee, S., Wray, N. R., Skarabis, N., Huang, H., Neale, B., Daly, M. J., … Ripke, S. (2020). RICOPILI: Rapid Imputation for COnsortias PIpeLIne. *Bioinformatics*, *36*(3), 930–933. https://doi.org/10.1093/bioinformatics/btz633

Leckman, J. F., Riddle, M. A., Hardin, M. T., Ort, S. I., Swartz, K. L., Stevenson, J., & Cohen, D. J. (1989). The Yale Global Tic Severity Scale: Initial Testing of a Clinician-Rated Scale of Tic Severity. *Journal of the American Academy of Child and Adolescent Psychiatry*, *28*(4), 566–573. https://doi.org/10.1097/00004583-198907000-00015

Lee, J. H., Cheng, R., Graff-Radford, N., Foroud, T., & Mayeux, R. (2008). Analyses of the national institute on aging late-onset Alzheimer's disease family study: Implication of additional loci. *Archives of Neurology*, *65*(11), 1518–1526. https://doi.org/10.1001/archneur.65.11.1518

Leitsalu, L., Alavere, H., Tammesoo, M. L., Leego, E., & Metspalu, A. (2015). Linking a population biobank with national health registries—The Estonian experience. *Journal of Personalized Medicine*, *5*(2). https://doi.org/10.3390/jpm5020096

Lek M, Karczewski KJ, Minikel EV, Samocha KE, Banks E, Fennell T, *et al.* (2016): Analysis of protein-coding genetic variation in 60,706 humans. *Nature* 536: 285–291.

Li, J. Z., Absher, D. M., Tang, H., Southwick, A. M., Casto, A. M., Ramachandran, S., Cann, H. M., Barsh, G. S., Feldman, M., Cavalli-Sforza, L. L., & Myers, R. M. (2008). Worldwide human relationships inferred from genome-wide patterns of variation. *Science*, *319*(5866), 1100–1104. https://doi.org/10.1126/science.1153717

Loh, P. R., Danecek, P., Palamara, P. F., Fuchsberger, C., Reshef, Y. A., Finucane, H. K., Schoenherr, S., Forer, L., McCarthy, S., Abecasis, G. R., Durbin, R., & Price, A. L. (2016). Reference-based phasing using the Haplotype Reference Consortium panel. *Nature Genetics 2016 48:11*, *48*(11), 1443–1448. https://doi.org/10.1038/ng.3679

Loh, P. R., Palamara, P. F., & Price, A. L. (2016). Fast and accurate long-range phasing in a UK Biobank cohort. *Nature Genetics 2016 48:7*, *48*(7), 811–816. https://doi.org/10.1038/ng.3571

Mahjani, B., Dellenvall, K., Grahnat, A. C. S., Karlsson, G., Tuuliainen, A., Reichert, J., Mahjani, C. G., Klei, L., De Rubeis, S., Reichenberg, A., Devlin, B., Hultman, C. M., Buxbaum, J. D., Sandin, S., & Grice, D. E. (2020). Cohort profile: Epidemiology and Genetics of Obsessive–compulsive disorder and chronic tic disorders in Sweden (EGOS). *Social Psychiatry and Psychiatric Epidemiology*, *55*(10), 1383–1393. https://doi.org/10.1007/s00127-019-01822-7

Manichaikul, A., Mychaleckyj, J. C., Rich, S. S., Daly, K., Sale, M., & Chen, W. M. (2010). Robust relationship inference in genome-wide association studies. *Bioinformatics*, *26*(22), 2867–2873. https://doi.org/10.1093/BIOINFORMATICS/BTQ559

Marchini, J., Howie, B., Myers, S., McVean, G., & Donnelly, P. (2007). A new multipoint method for genome-wide association studies by imputation of genotypes. *Nature Genetics*, *39*(7), Article 7. https://doi.org/10.1038/ng2088

Margraf, J., Cwik, J. C., Pflug, V., & Schneider, S. (2017). Strukturierte klinische Interviews zur Erfassung psychischer Störungen über die Lebensspanne. *Zeitschrift Für Klinische Psychologie Und Psychotherapie*, *46*(3), 176–186. https://doi.org/10.1026/1616-3443/a000430

Mars, N., Koskela, J. T., Ripatti, P., Kiiskinen, T. T. J., Havulinna, A. S., Lindbohm, J. V., Ahola-Olli, A., Kurki, M., Karjalainen, J., Palta, P., Neale, B. M., Daly, M., Salomaa, V., Palotie, A., Widén, E., & Ripatti, S. (2020). Polygenic and clinical risk scores and their impact on age at onset and prediction of cardiometabolic diseases and common cancers. *Nature Medicine*, *26*(4), 549–557. https://doi.org/10.1038/s41591-020-0800-0

Mataix-Cols, D., Hansen, B., Mattheisen, M., Karlsson, E. K., Addington, A. M., Boberg, J., Djurfeldt, D. R., Halvorsen, M., Lichtenstein, P., Solem, S., Lindblad-Toh, K., Nordic OCD and Related Disorders Consortium (NORDiC), Haavik, J., Kvale, G., Rück, C., & Crowley, J. J. (2020). Nordic OCD & Related Disorders Consortium: Rationale, design, and methods. *American Journal of Medical Genetics. Part B, Neuropsychiatric Genetics: The Official Publication of the International Society of Psychiatric Genetics*, *183*(1), 38–50. https://doi.org/10.1002/ajmg.b.32756

Mattheisen, M., Samuels, J. F., Wang, Y., Greenberg, B. D., Fyer, A. J., Mccracken, J. T., Geller, D. A., Murphy, D. L., Knowles, J. A., Grados, M. A., Riddle, M. A., Rasmussen, S. A., Mclaughlin, N. C., Nurmi, E. L., Askland, K. D., Qin, H. D., Cullen, B. A., Piacentini, J., Pauls, D. L., … Nestadt, G. (2015). Genome-wide association study in obsessive-compulsive disorder: Results from the OCGAS. *Molecular Psychiatry*, *20*(3), 337–344. https://doi.org/10.1038/mp.2014.43

Mbatchou, J., Barnard, L., Backman, J., Marcketta, A., Kosmicki, J. A., Ziyatdinov, A., Benner, C., O'Dushlaine, C., Barber, M., Boutkov, B., Habegger, L., Ferreira, M., Baras, A., Reid, J., Abecasis, G., Maxwell, E., & Marchini, J. (2021). Computationally efficient whole-genome regression for quantitative and binary traits. *Nature Genetics*, *53*(7), Article 7. https://doi.org/10.1038/s41588-021-00870-7

McCarthy, S., Das, S., Kretzschmar, W., Delaneau, O., Wood, A. R., Teumer, A., Kang, H. M., Fuchsberger, C., Danecek, P., Sharp, K., Luo, Y., Sidore, C., Kwong, A., Timpson, N., Koskinen, S., Vrieze, S., Scott, L. J., Zhang, H., Mahajan, A., … Marchini, J. (2016). A reference panel of 64,976 haplotypes for genotype imputation. *Nature Genetics 2016 48:10*, *48*(10), 1279–1283. https://doi.org/10.1038/ng.3643

Mitt, M., Kals, M., Pärn, K., Gabriel, S. B., Lander, E. S., Palotie, A., Ripatti, S., Morris, A. P., Metspalu, A., Esko, T., Mägi, R., & Palta, P. (2017). Improved imputation accuracy of rare and low-frequency variants using population-specific high-coverage WGS-based imputation reference panel. *European Journal of Human Genetics*, *25*(7). https://doi.org/10.1038/ejhg.2017.51

O'Connell, J. R., & Weeks, D. E. (1998). PedCheck: A program for identification of genotype incompatibilities in linkage analysis. *American Journal of Human Genetics*, *63*(1), 259–266. https://doi.org/10.1086/301904

Pato, M. T., Sobell, J. L., Medeiros, H., Abbott, C., Sklar, B. M., Buckley, P. F., Bromet, E. J., Escamilla, M. A., Fanous, A. H., Lehrer, D. S., Macciardi, F., Malaspina, D., McCarroll, S. A., Marder, S. R., Moran, J., Morley, C. P., Nicolini, H., Perkins, D. O., Purcell, S. M., … Pato, C. N. (2013). The genomic psychiatry cohort: Partners in discovery. *American Journal of Medical Genetics. Part B, Neuropsychiatric Genetics: The Official Publication of the International Society of Psychiatric Genetics*, *162B*(4), 306–312. https://doi.org/10.1002/ajmg.b.32160

Pedersen, B. S., & Quinlan, A. R. (2017). Who's Who? Detecting and Resolving Sample Anomalies in Human DNA Sequencing Studies with Peddy. *The American Journal of Human Genetics*, *100*(3), 406–413. https://doi.org/10.1016/j.ajhg.2017.01.017

Pedersen, C. B., Bybjerg-Grauholm, J., Pedersen, M. G., Grove, J., Agerbo, E., Bækvad-Hansen, M., Poulsen, J. B., Hansen, C. S., McGrath, J. J., Als, T. D., Goldstein, J. I., Neale, B. M., Daly, M. J., Hougaard, D. M., Mors, O., Nordentoft, M., Børglum, A. D., Werge, T., &

Mortensen, P. B. (2018). The iPSYCH2012 case-cohort sample: New directions for unravelling genetic and environmental architectures of severe mental disorders. *Mol Psychiatry*, *23*(1), 6–14. https://doi.org/10.1038/mp.2017.196

Price, A. L., Patterson, N. J., Plenge, R. M., Weinblatt, M. E., Shadick, N. A., & Reich, D. (2006). Principal components analysis corrects for stratification in genome-wide association studies. *Nature Genetics*, *38*(8), 904–909. https://doi.org/10.1038/NG1847

Purcell, S., Neale, B., Todd-Brown, K., Thomas, L., Ferreira, M. A. R., Bender, D., Maller, J., Sklar, P., De Bakker, P. I. W., Daly, M. J., & Sham, P. C. (2007). PLINK: a tool set for whole-genome association and population-based linkage analyses. *American Journal of Human Genetics*, *81*(3), 559–575. https://doi.org/10.1086/519795

Rovira, P., Demontis, D., Sánchez-Mora, C., Zayats, T., Klein, M., Mota, N. R., Weber, H., Garcia-Martínez, I., Pagerols, M., Vilar-Ribó, L., Arribas, L., Richarte, V., Corrales, M., Fadeuilhe, C., Bosch, R., Martin, G. E., Almos, P., Doyle, A. E., Grevet, E. H., … Ribasés, M. (2020). Shared genetic background between children and adults with attention deficit/hyperactivity disorder. *Neuropsychopharmacology*, *45*(10), Article 10. https://doi.org/10.1038/s41386-020-0664-5

Satterthwaite, T. D., Connolly, J. J., Ruparel, K., Calkins, M. E., Jackson, C., Elliott, M. A., Roalf, D. R., Hopson, R., Prabhakaran, K., Behr, M., Qiu, H., Mentch, F. D., Chiavacci, R., Sleiman, P. M. A., Gur, R. C., Hakonarson, H., & Gur, R. E. (2016). The Philadelphia Neurodevelopmental Cohort: A publicly available resource for the study of normal and abnormal brain development in youth. *NeuroImage*, *124*(Pt B), 1115–1119. https://doi.org/10.1016/j.neuroimage.2015.03.056

Scahill, L., Riddle, M. A., McSwiggin-Hardin, M., Ort, S. I., King, R. A., Goodman, W. K., Cicchetti, D., & Leckman, J. F. (1997). Children’s Yale-Brown Obsessive Compulsive Scale: Reliability and validity. *Journal of the American Academy of Child and Adolescent Psychiatry*, *36*(6), 844–852. https://doi.org/10.1097/00004583-199706000-00023

Schmidt, M. H., Döpfner, M., Berner, W., Flechtner, H., Lehmkuhl, G., & Steinhausen, H.-C. E. (2000). *Psychopathologisches Befund-System für Kinder und Jugendliche (CASCAP-D)*. 28(1):64-64. https://doi.org/10.1024//1422-4917.28.1.64

Schumacher, F. R., Berndt, S. I., Siddiq, A., Jacobs, K. B., Wang, Z., Lindstrom, S., Stevens, V. L., Chen, C., Mondul, A. M., Travis, R. C., Stram, D. O., Eeles, R. A., Easton, D. F., Giles, G., Hopper, J. L., Neal, D. E., Hamdy, F. C., Donovan, J. L., Muir, K., … Kraft, P. (2011). Genome-wide association study identifies new prostate cancer susceptibility loci. *Human Molecular Genetics*, *20*(19), 3867–3875. https://doi.org/10.1093/hmg/ddr295

Sherva, R., Wang, Q., Kranzler, H., Zhao, H., Koesterer, R., Herman, A., Farrer, L. A., & Gelernter, J. (2016). Genome-wide Association Study of Cannabis Dependence Severity, Novel Risk Variants, and Shared Genetic Risks. *JAMA Psychiatry*, *73*(5), 472–480. https://doi.org/10.1001/jamapsychiatry.2016.0036

Solovieff, N., Hartley, S. W., Baldwin, C. T., Perls, T. T., Steinberg, M. H., & Sebastiani, P. (2010). Clustering by genetic ancestry using genome-wide SNP data. *BMC Genetics*, *11*, 108. https://doi.org/10.1186/1471-2156-11-108

Stewart, S. E., Mayerfeld, C., Arnold, P. D., Crane, J. R., O'Dushlaine, C., Fagerness, J. A., Yu, D., Scharf, J. M., Chan, E., Kassam, F., Moya, P. R., Wendland, J. R., Delorme, R., Richter, M. A., Kennedy, J. L., Veenstra-Vanderweele, J., Samuels, J., Greenberg, B. D., Mccracken, J. T., … Mathews, C. A. (2013). Meta-analysis of association between obsessive-compulsive disorder and the 3′ region of neuronal glutamate transporter gene SLC1A1. *American Journal of Medical Genetics, Part B: Neuropsychiatric Genetics*, *162*(4), 367–379. https://doi.org/10.1002/ajmg.b.32137

Strom, N. I., Verhulst, B., Bacanu, S.-A., Cheesman, R., Purves, K. L., Gedik, H., … Hettema, J. (2024) Genome-wide association study of major anxiety disorders in 122,341 European-ancestry cases identifies 58 loci and highlights GABAergic signaling. medRxiv 2024.07.03.24309466; doi: https://doi.org/10.1101/2024.07.03.24309466

The 1000 Genomes Project Consortium. (2015). A global reference for human genetic variation. *Nature*, *526*(7571), 68–74. https://doi.org/10.1038/nature15393

Võsa, U., Claringbould, A., Westra, H.-J., Bonder, M. J., Deelen, P., Zeng, B., Kirsten, H., Saha, A., Kreuzhuber, R., Yazar, S., Brugge, H., Oelen, R., de Vries, D. H., van der Wijst, M. G. P., Kasela, S., Pervjakova, N., Alves, I., Favé, M.-J., Agbessi, M., … Franke, L. (2021). Large-scale cis- and trans-eQTL analyses identify thousands of genetic loci and polygenic scores that regulate blood gene expression. *Nature Genetics*, *53*(9), 1300–1310. https://doi.org/10.1038/s41588-021-00913-z

Wainberg, M., Merico, D., Keller, M. C., Fauman, E. B., & Tripathy, S. J. (2022). Predicting causal genes from psychiatric genome-wide association studies using high-level etiological knowledge. *Molecular Psychiatry*, *27*(7), Article 7. https://doi.org/10.1038/s41380-022-01542-6

Wang, C., Zhan, X., Bragg-Gresham, J., Kang, H. M., Stambolian, D., Chew, E. Y., Branham, K. E., Heckenlively, J., Fulton, R., Wilson, R. K., Mardis, E. R., Lin, X., Swaroop, A., Zöllner, S., & Abecasis, G. R. (2014). Ancestry estimation and control of population stratification for sequence-based association studies. *Nature Genetics*, *46*(4), 409–415. https://doi.org/10.1038/ng.2924

Wang S, Mandell JD, Kumar Y, Sun N, Morris MT, Arbelaez J, *et al.* (2018): De Novo Sequence and Copy Number Variants Are Strongly Associated with Tourette Disorder and Implicate Cell Polarity in Pathogenesis. *Cell Rep* 25: 3544.

Warren, H. R., Evangelou, E., Cabrera, C. P., Gao, H., Ren, M., Mifsud, B., Ntalla, I., Surendran, P., Liu, C., Cook, J. P., Kraja, A. T., Drenos, F., Loh, M., Verweij, N., Marten, J., Karaman, I., Segura Lepe, M. P., O'Reilly, P. F., Knight, J., … Morris, A. P. (2017). Genome-wide association analysis identifies novel blood pressure loci and offers biological insights into cardiovascular risk. *Nature Genetics*, *49*(3), 403–415. https://doi.org/10.1038/ng.3768

Wray, N.R., Ripke, S., Mattheisen, M. et al. Genome-wide association analyses identify 44 risk variants and refine the genetic architecture of major depression. Nat Genet 50, 668–681 (2018). https://doi.org/10.1038/s41588-018-0090-3

Yang, J., Lee, S. H., Goddard, M. E., & Visscher, P. M. (2011). GCTA: a tool for genome-wide complex trait analysis. American journal of human genetics, 88(1), 76–82. https://doi.org/10.1016/j.ajhg.2010.11.011

Zhou, W., Nielsen, J. B., Fritsche, L. G., Dey, R., Gabrielsen, M. E., Wolford, B. N., LeFaive, J., VandeHaar, P., Gagliano, S. A., Gifford, A., Bastarache, L. A., Wei, W. Q., Denny, J. C., Lin, M., Hveem, K., Kang, H. M., Abecasis, G. R., Willer, C. J., & Lee, S. (2018). Efficiently controlling for case-control imbalance and sample relatedness in large-scale genetic association studies. *Nature Genetics*, *50*(9), 1335–1341. https://doi.org/10.1038/s41588-018-0184-y
